# Causes of Outcome Learning: A causal inference-inspired machine learning approach to disentangling common combinations of potential causes of a health outcome

**DOI:** 10.1101/2020.12.10.20225243

**Authors:** A Rieckmann, P Dworzynski, L Arras, S Lapuschkin, W Samek, OA Arah, NH Rod, CT Ekstrøm

## Abstract

Nearly all diseases can be caused by different combinations of exposures. Yet, most epidemiological studies focus on the causal effect of a single exposure on an outcome. We present the Causes of Outcome Learning (CoOL) approach, which seeks to identify combinations of exposures (which can be interpreted causally if all causal assumptions are met) that could be responsible for an increased risk of a health outcome in population sub-groups. The approach allows for exposures acting alone and in synergy with others. It involves (a) a pre-computational phase that proposes a causal model; (b) a computational phase with three steps, namely (i) analytically fitting a non-negative additive model, (ii) decomposing risk contributions, and (iii) clustering individuals based on the risk contributions into sub-groups based on the predefined causal model; and (c) a post-computational phase on hypothesis development and validation by triangulation on new data before eventually updating the causal model. The computational phase uses a tailored neural network for the non-negative additive model and Layer-wise Relevance Propagation for the risk decomposition through this model. We demonstrate the approach on simulated and real-life data using the R package ‘CoOL’. The presentation is focused on binary exposures and outcomes but can be extended to other measurement types. This approach encourages and enables epidemiologists to identify combinations of pre-outcome exposures as potential causes of the health outcome of interest. Expanding our ability to discover complex causes could eventually result in more effective, targeted, and informed interventions prioritized for their public health impact.

## Introduction

Many diseases may be caused by several different combinations of exposures. As putative causes, such exposures may act together and lead to a combined effect that exceeds the sum of the individual effects, also called synergism. A common example of synergism is how the combined effect of smoking and asbestos on lung cancer exceeds the sum of their individual effects.[1] The most established theoretical framework for studying synergistic effects of multiple causes in epidemiology is the *sufficient cause model*,[2]. Assessment of synergism between causes may provide etiological insight into how to prevent and treat disease. It may also help to identify and quantify the disease burden in high-risk sub-groups. Thus, understanding the spectra of exposures rather than single exposures for effective preventive strategies has been highlighted as essential for decades. Rose for example says “*… risk assessment must consider all relevant factors together rather than confine attention to a single test, for nearly all diseases are multifactorial*” when discussing effective policy decisions.[3]

Despite the policy relevance, few epidemiological studies have analytically tried to identify combination of causes for specific outcomes. We suspect that the apparent lack of epidemiological studies for questions about causes of outcomes is due to frequently taught frameworks for epidemiologists that warn against type 1 errors from multiple testing (false positive findings),[4] various confounding structures for each exposure,[5] the overwhelming number of combinations between exposures that can be created,[6] and lack of established theoretically founded approaches for applied data analysis,[6,7] though some do exist.[8–10]

We introduce a machine learning- and causal inference-based approach called the Causes of Outcome Learning (CoOL) approach that attempts to generate insights into questions like *“Given a particular health outcome, what are the most common combinations of exposures, which might have been its causes?”*. We present the approach assisted by a simple simulated example. A step-by-step tutorial is included in Supplementary simulations 1-6, robustness checks in Supplementary simulations 7-9, and a real-life application in Supplementary real life data analysis. Terms that may need additional explanation are marked with * and explained in the Supplementary Table 1.

### Motivating example

We use a simple simulation setting to motivate the CoOL approach. We generate a study population of 10,000 individuals, of whom 50% are men and 50% are women, 20% are exposed to drug A, and 20% are exposed to drug B. Sex, drug A, and drug B are independent. All individuals have a baseline risk of disease Y of 5% throughout follow up, men who are exposed to drug A have a 15% increased risk of disease Y, and so do women who are exposed to drug B. If we were using a real-life dataset instead of a simulated one, we would need methods that could help us to identify the different risks associated with the measured exposures and population characteristics in order to eventually prevent disease Y. We will show how the CoOL approach can help direct us towards sets of exposures which might have caused our health outcome of interest in this example and in more complex simulations (see Supplementary material).

### The Causes of Outcome Learning approach

The CoOL approach is enabled by three major new developments in the fields of machine learning* and causal inference*. Firstly, advances in computing and machine learning allow for identification of complex structures in large datasets. Secondly, there has been a recent breakthrough in understanding why machine learning models produce the results they do (explainable AI such as Layer-Wise Relevance Propagation [LRP] [11–13]). Lastly, by assuming a causal structure of data, models may be interpreted as structural causal models, which allows for causal interpretation.[14] The CoOL approach follows 3 phases (a-c) to ensure the embeddedness in an inductive-deductive scientific process, of which our contribution is specifically related to the computational phase (Figure 1).

a. Pre-computational phase: Propose a causal model using a Directed Acyclic Graph (DAG) of the exposures and the outcome based on prior domain expertise of selected actionable exposures and contextual factors.
b. Computational phase (We provide the R package ‘CoOL’, see Supplementary information 1 for installation):

- On a training dataset:

i) Fit a *non-negative additive model*\* based on the features from the assumed causal model
ii) Decompose the risk contributions.
iii) Cluster individuals based on the risk contributions.
- On an internal validation dataset:

iv. Ensure the robustness of the findings in an internal validation dataset.
c. Post-computational phase: Based on the new learnings from the computational phase and existing knowledge, develop hypotheses to be assessed in further (intervention) studies on new data. Our approach focuses on common sub-groups with high risks, and thus the presentation of the results can direct the researchers towards insight with potential large public health impact. The scientific process will continue in an inductive-deductive continuum towards inference to the best explanation.

**Figure 1.**
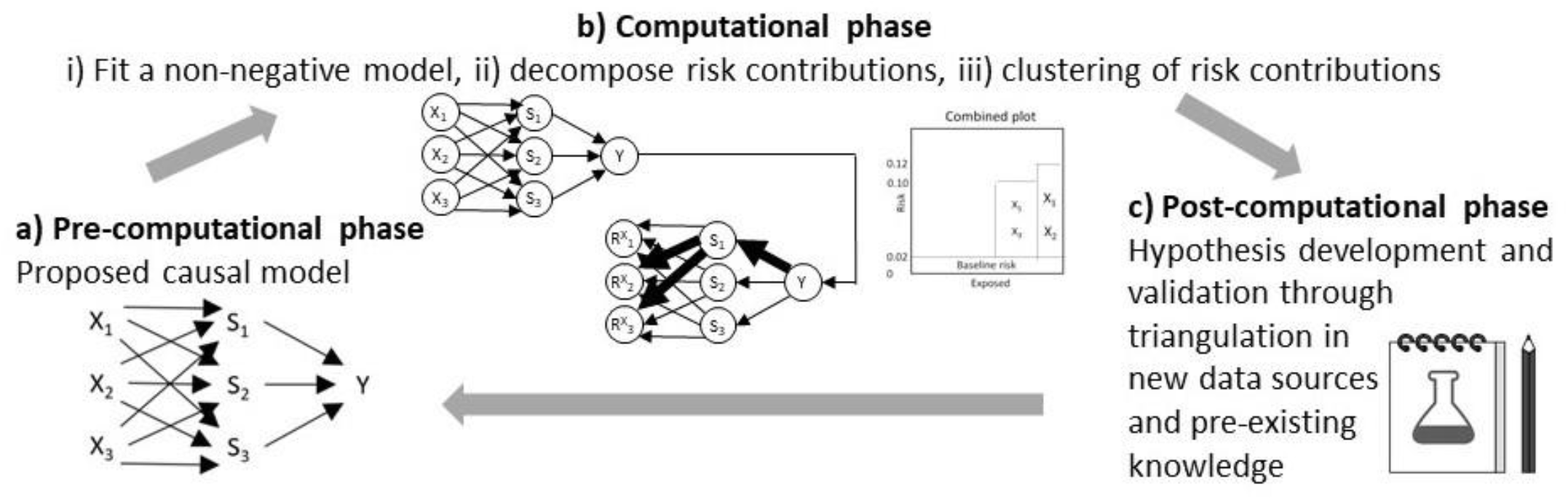
The phases of the CoOL approach towards inference to the best explanation. A) Pre-computational phase: Scoping the research question and causal structure assumptions. Computational phase: I) A non-negative model as close to the assumed causal model is fitted, II) risk contributions are decomposed and III) individuals are clustered into sub-groups. IV) Manual validation of the results is suggested in an internal validation dataset to assess the stability of the results. C) Post-computational phase: the results are held against existing evidence in order to develop new hypotheses that can be tested in new studies. new understandings will update our initial assumed causal model.

### Pre-computational phase on proposing a causal model

Causal structures are commonly depicted with DAGs,[15] which allow for a causal interpretation of associations given a set of *causal assumptions*\*: exchangeability, positivity, consistency, no measurement error, and no model misspecification.[6]

The intuition behind the assumed causal model is to link exposures to unknown sufficient causes.[16] Figure 2A ideally shows unknown combinations of exposures that cause the health outcome. Figure 2C shows the theoretical DAG, where *X*_*i*_ denotes exposures, *SC*_*j*_ denotes j unknown sets of sufficient causes for the outcome (inspired by the notation by VanderWeele and Robins [16]), and *Y* denotes the outcome. The U_SCi_ and U denotes different types of unmeasured (including unmeasurable and unknown) causes; U_SCi_ denotes the unmeasured component causes of SC_j_, while U denotes unmeasured causes of Y of which we assume all individuals are exposed to. This theoretical DAG avoids making assumptions of the lack of causal effects between exposures and sufficient causes, and the computational steps will aim at reducing these causal effects towards the minimal sets of component causes. The assumed causal model assists in the selection of exposures to include in the model: there are actionable exposures, i.e. those we can intervene on such as drug intake, and contextual factors, which help describe sub-groups in risk. It also helps to decide whether proximal non-actionable exposures should be left out of the model, as they may mediate effects of actionable exposures. Further, the assumed causal model is used for the interpretation of the results because only direct and joint effects are returned.[5]

**Figure 2.**
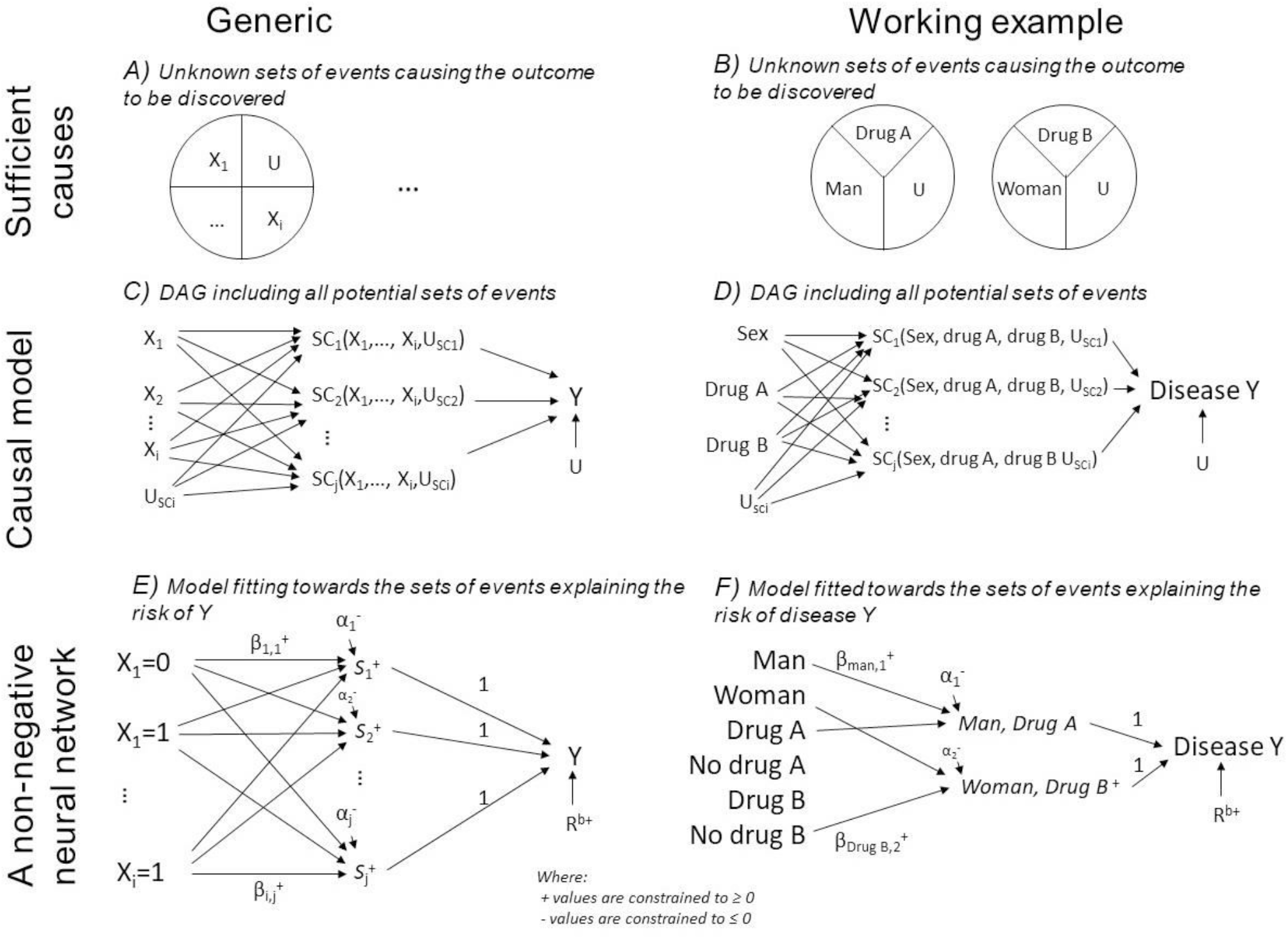
Sufficient causes, causal model and non-negative neural network. The pictogram shows the relation between epidemiological theory, structural models and a non-negative neural network. The left column is a generic presentation, and the right column shows the simulated example. A-B) An illustration of sufficient causes. The example to the right shows that a certain disease occurs if men are exposed to Drug A and some unknown factors and if women are exposed to drug B and some unknown factors. C-D) An assumed causal model illustrated using a directed acyclic graph, where *X*_*i*_ denotes the exposures, U_SCi_ denotes the unmeasured causes of the sufficient causes, U denotes the unmeasured causes of Y assumed to affect all individuals, *SC*_*j*_ denotes *hidden* sufficient causes, and *Y* denotes the outcome. E-F) A non-negative neural network resembling the assumed causal model. *X*_*i*_ denotes exposures, 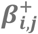 denotes non-negative parameters, 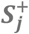 denotes hidden synergy-functions, 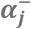 denotes non-positive intercepts, acting as activation thresholds for synergy-functions, and 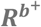 denotes the baseline risk.

The model choice affects our causal interpretations,[17] and models for estimating synergistic effects assume positive monotonicity, i.e. exposures either have no effect or always act in the same direction on the outcome.[18,19] The proposed non-negative model (next section) relaxes the monotonicity assumption by letting us explore all directions of exposures on the outcome simultaneously for which effects act independently or synergistically with others (e.g. if there exists exposures that are especially harmful for men and other exposures that are especially harmful for women).

To identify an elevated risk, we need to define a reference *baseline risk, R*^*b*+^, of all unmeasured causes assumed to affect all individuals. Given the causal structure is correct, the average effect for being exposed to combination *z* of the exposures compared with the baseline risk is given by 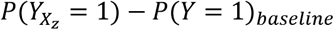, which can be estimated as *P*(*Y* = 1|*X* = *x*_*z*_) − *R*^*b*+^, and the average effect of removing one exposure, by setting X_i_ to 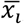, can be estimated as 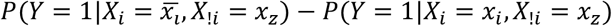.

As in all studies aiming at causal inference, appropriate adjustment of exposures causing confounding is of great concern.[6] In the CoOL approach, *confounders*\* are of interest as they could be causes of the outcome and be additionally associated with the exposures or they could be causes of the exposures in addition to being associated with the outcome.[20] By including the factors responsible for confounding, we block spurious associations between exposures and the outcome, which they would otherwise introduce (*the backdoor path criterion*\*) [14]. Researchers should also consider situations where, e.g. differences in disease definition or surveillance and changes in registrations of exposures are known to have happened over time.[21] Issues with selection or collider bias and measurement bias should equally well be considered.

For our motivating example, we assume that sex, drug A, and drug B do not share a common cause. Ideally, we want to identify the sufficient causes shown in Figure 2B, and the DAG showing our scientific interest can be drawn as in Figure 2D.

### Computational phase

Since the number of potential combinations of exposures is large, there is a risk of type 1 errors. To prevent the model from *overfitting*\* to noise, data is split into a training dataset and a dataset for internal validation. We suggest training until the model converges based on the error function for the training dataset. The findings from the training dataset can be manually confirmed in the yet unseen internal validation dataset before developing hypotheses.

### 1. Fitting a non-negative model

Various non-negative models may be used - we suggest a *non-negative*\* *additive*\* *single-hidden layer*\* *neural network*\* (Figure 2E) designed to mimic our assumed causal model (Figure 2C). This model resembles a linear regression model estimating risk differences but with two main modifications. First, the model includes a series of unobserved mediators that can combine the effects of various exposures. We call these unobserved mediators the *synergy-functions*\*, *S*^+^(), represented in the hidden layer between the exposures and the outcome. Second, we restrict all connection parameters to have non-negative values,[22] so that exposures can only increase the occurrence of the outcome and thereby meet a relaxed version of the monotonicity assumption.[18] In Figure 2E, *X*_*i*_ denotes *i* exposures (each category of the variable is *binary (/one-hot) encoded*\* into one new variable each with 0 if not present and 1 if present) and *Y* denotes the disease outcome (coded 0 and 1); 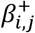 denotes connection parameters from the exposures to the synergy-functions (parameters can only take non-negative values), *S*^+^(), which return the non-negative sum of its input value or zero; 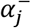 is an intercept (can only take non-positive values) that acts as an activation threshold which only allows combinations of exposures with large 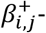 weighted sum to pass *S*^+^(); *R*^*b*+^ represents the baseline risk (can only take non-negative values); and the parameter of connections between the synergy functions and the outcome has a fixed value of 1. The model estimates the risk on an additive scale so that synergism is defined as combined effects that are larger than the sum of individual effects.[2] Clearly, probabilities larger than 1 indicate model misspecification.

This model can be denoted as below, where *S*^+^(*x*) = *max*(0, *x*), and likewise ^+^ denotes restrictions to non-negative values, and ^−^ denotes restrictions to non-positive values. This model satisfies the assumption that the added risk is independent of the baseline risk or phrased as an “independent of background” model by Beyea and Greenland.[17]

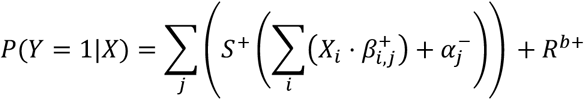

Fitting the model is done using *stochastic gradient decent*\* on the training dataset: In a step-wise procedure run on one individual at a time, the model estimates the individual’s risk of the disease outcome, *P*(*Y*|*X*), calculates the squared prediction error (*Y* − *P*(*Y*|*X*))^2^ and adjusts the model parameters to minimize this error.[23] By iterating through all individuals for multiple *epochs*\*, we obtain model parameters, which minimizes the sum of prediction errors across the entire population. The *initial values*\*, *derivatives*\*, *learning rates*\*, and *regularizations*\* are described in Supplementary information 2.

While the prediction performance measured by the area under the receiver operating characteristic curve (AUC) provides a useful metric for evaluating model discriminatory performance across the entire population, it is important to consider that in case the outcome is caused by multiple distinct sets of causes, a model with low AUC can still capture sets of causes for a particular sub-group.[24]

Figure 2F shows the model for our motivating example. We binary encode new variables for each possible category of each exposure, such that sex (coded 0 if man, 1 if women) becomes two factors; man (coded 1 if man, 0 if not man) and woman (coded 1 if woman, 0 if not woman) and so forth for drug A and drug B. This data is used to fit the proposed non-negative model with 10 synergy-functions. Figure 4A-C shows how the error decreases by each epoch, it visualizes the neural network connections, and shows the accuracy using the prediction performance measure, AUC.

### 2. Decomposing risk contributions

Machine learning models are commonly referred to as black boxes due to the limited interpretability of their parameters and how they interact with the input-variables.[25] Instead of attempting to interpret the model directly, we use LRP [11–13] to decompose the risk of the outcome to *risk contributions*\* for each individual (in particular, we use the LRP_alpha,beta_-rule, with alpha=1 and beta=0). LRP was introduced by Bach et al. in 2015[11] as a decomposition technique for pre-trained neural networks, and was later justified via Deep Taylor Decomposition.[26] As opposed to other explanation techniques for neural networks, LRP is aimed at conserving the information for predicting the outcome when assigning relevance to the inputs that were driving the prediction. In the CoOL approach, the predicted risk of the outcome, *P*(*Y* = 1|*X*) is decomposed into a baseline risk, *R*^*b*+^, and the risk contributions by each exposure, 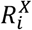 (where *P*(*Y* = 1|*X*) can take values between 0 and 1):

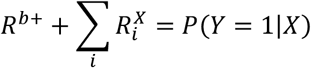

These risk contributions may be interpreted as the exposures’ positive contribution to the risk given the model and the individual’s set of exposures. No risk contributions are decomposed to the intercepts, 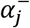 The below procedure is conducted for all individuals in a one-by-one fashion. The baseline risk, *R*^*b*+^, is represented by its own parameter as illustrated in Figure 2E, and is therefore estimated as part of fitting the non-negative neural network. The decomposition of the risk contributions for exposures, 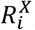,takes 3 steps:

Step 1 - Subtract the baseline risk, 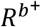:

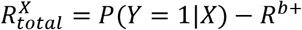

Step 2 - Decompose risk contributions to the synergy-functions, where *S*_*j*_ is the value returned by each of the *j* synergy-functions given the exposure distribution *X*_*i*_, parameters, 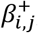, and intercepts, 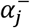:

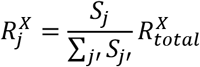

Step 3 - Decompose risk contributions from the synergy-functions to the exposures:

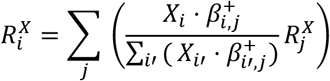

As a result of the risk decomposition, each individual is assigned a set of risk contributions, 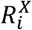, one for each exposure plus a baseline risk, *R*^*b*+^. The decomposition of risk contributions can be illustrated in Figure 3E-F using the motivating example and explanation in the figure legend.

**Figure 3.**
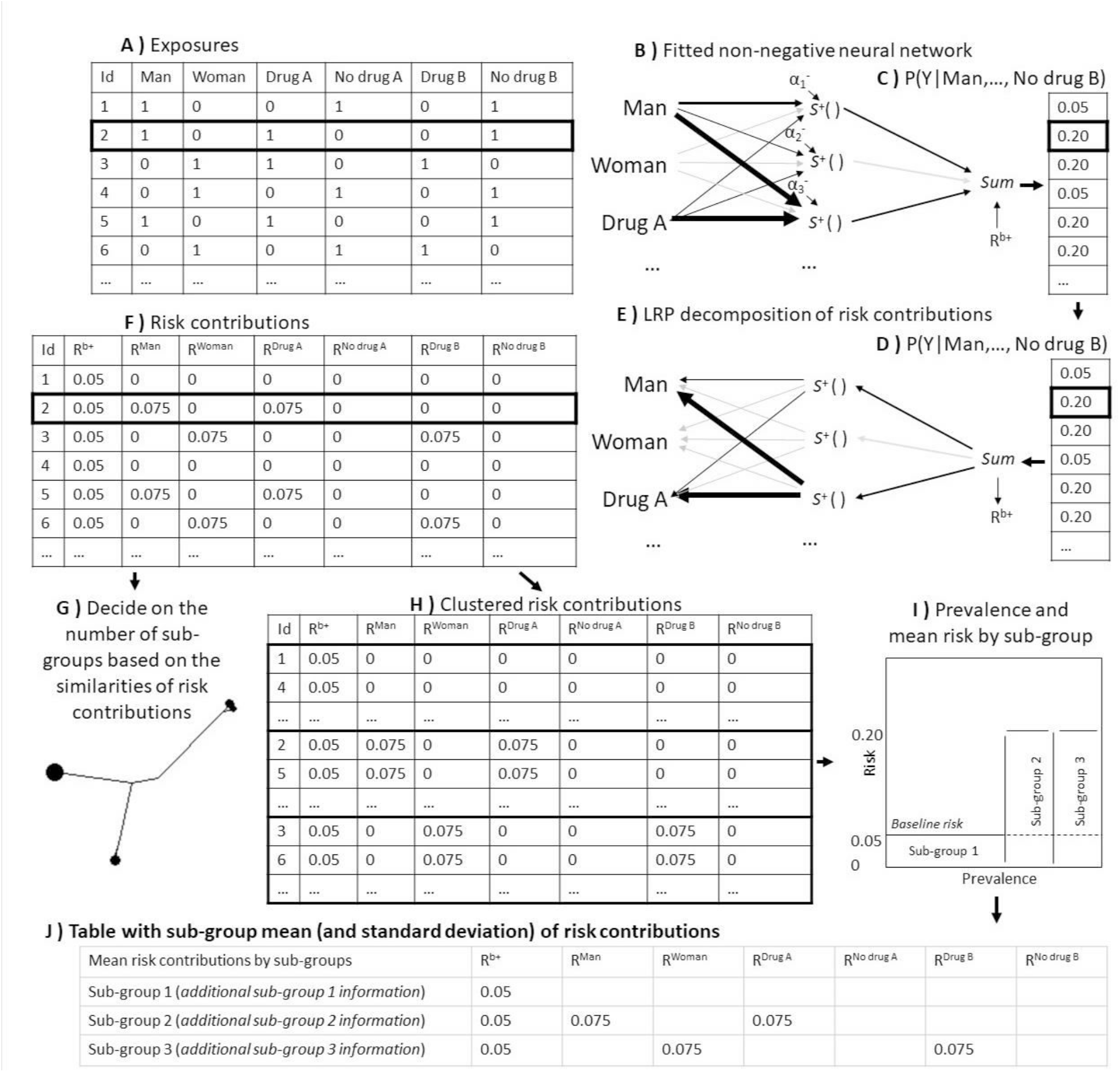
Workflow of the computational phase of CoOL. The flowchart of how sub-groups are identified as part of the computational phase of Causes of Outcome Learning. A) The expanded dataset of sex (one variable for man, one for woman), drug A (one variable for drug A, one for no drug A), and drug B (one variable for drug B, one for no drug B). B) The fitted non-negative model is illustrated. wide edges indicate large connection parameters. C-D) The predicted risk, *P*(*Y*|*X*). E) The predicted risk is decomposed using LRP to risk contributions of the baseline, *R*^*b*+^, and exposures, *R*^*X*^. F) The risk contribution matrix. G) A dendrogram to help decide on the number of sub-groups. H) Clustered risk contribution matrix into sub-groups. I) Prevalence and mean risk by sub-group plot. This plot indicate areas for greater public health impact. J) A table with sub-group mean of risk contributions. It can hold more information which can be useful when developing hypotheses, such as quantifications of the excess proportion of all cases found is this sub-groups when considering the prevalence of the subgroup, the risk in the sub-group and the baseline risk.

### 3. Clustering of risk contributions

We suggest to sub-group the individuals based on risk contributions using *Manhattan distances*\* and *the Ward method*\*.[27,28] One helpful technique that could inform deciding on number of subgroups is a dendrogram[29] of the distance matrix with node sizes representing the prevalence of similar risk contributions to be used to decide the number of sub-groups (Figure 3G). Additionally, we can plot the prevalence and mean risk of each sub-groups (inspired by excess probability plots[30]) to help identify the sub-groups with a high impact by identifying the area within a sub-group above the baseline risk (Figure 3I). Further, we can make a table of mean risk contributions and standard deviations (SD) by sub-groups to illuminate which exposures elevate the risk in each sub-group (Figure 3J). An indication of synergism is when the combined risk contribution of a set of exposures is higher than the sum of stand-alone risk contributions of each of the exposures, which can also be added to the table (Supplementary information 3, but it should be interpreted with caution as deviation may occur in noisy datasets). Formal investigation of synergism should be done in the yet unseen internal validation dataset before developing hypotheses for phase 3 of the Causes of Outcome Learning approach (see formulas for formal interaction analyses by VanderWeele[31]).

If exposures identified as high risk contributors are associated with the outcome due to uncontrolled confounding, the risk contributors only help identify the high-risk group and removing the exposure will not necessarily affect the risk. If two component causes act solely in synergy on the outcome, then removing just one of them is sufficient by itself, and thus the estimated risk contributions underestimate the causal effects in a counterfactual framework. The area within a sub-group above the baseline risk (Figure 3I) indicate the excess fraction of all cases due to the combination of exposures in the sub-group and thus indicate groups where a large public health impact may be made, but the interpretation should depend on how well the causal mechanisms are understood. The term also relates to the concept of grouped partial attributable risks[32] or termed formally as the attributable proportion in the population[19] and can be defined for a subgroup Z as:

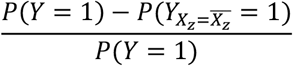

where 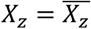 denotes eliminating risk contributors in subgroup Z. Given the combined risk contributions causally affect the outcome and meet the assumption of positive monotonicity, the excess fraction can be calculated as (Supplementary information 4):

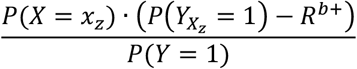

Analyzing our motivating example, we can apply the fitted non-negative model, decompose the risk contributions using LRP and show a dendrogram of how similar the population are to each other in Figure 4D, which indicated 3 groups. Figure 4E shows the risk and prevalence of the 3 sub-groups, where one sub-group which has a risk of 5%, a second sub-group that has a risk of approximately 20% with a prevalence of 10%, and a third sub-group that has a risk of approximately 20% with a prevalence of 10%. Figure 4F shows us that we correctly identified that men (sex_0) who are exposed to drug A (drug_a_1) have a 5% baseline risk, which reaches a near 20% risk through the contributions from being a man and drug A. Similar are the findings for women (sex_1) and drug B (drug_b_1). Though not observed in this analysis, we may expect that the predicted risks are slightly underestimated since we apply regularization to reduce noise signals in data.

**Figure 4.**
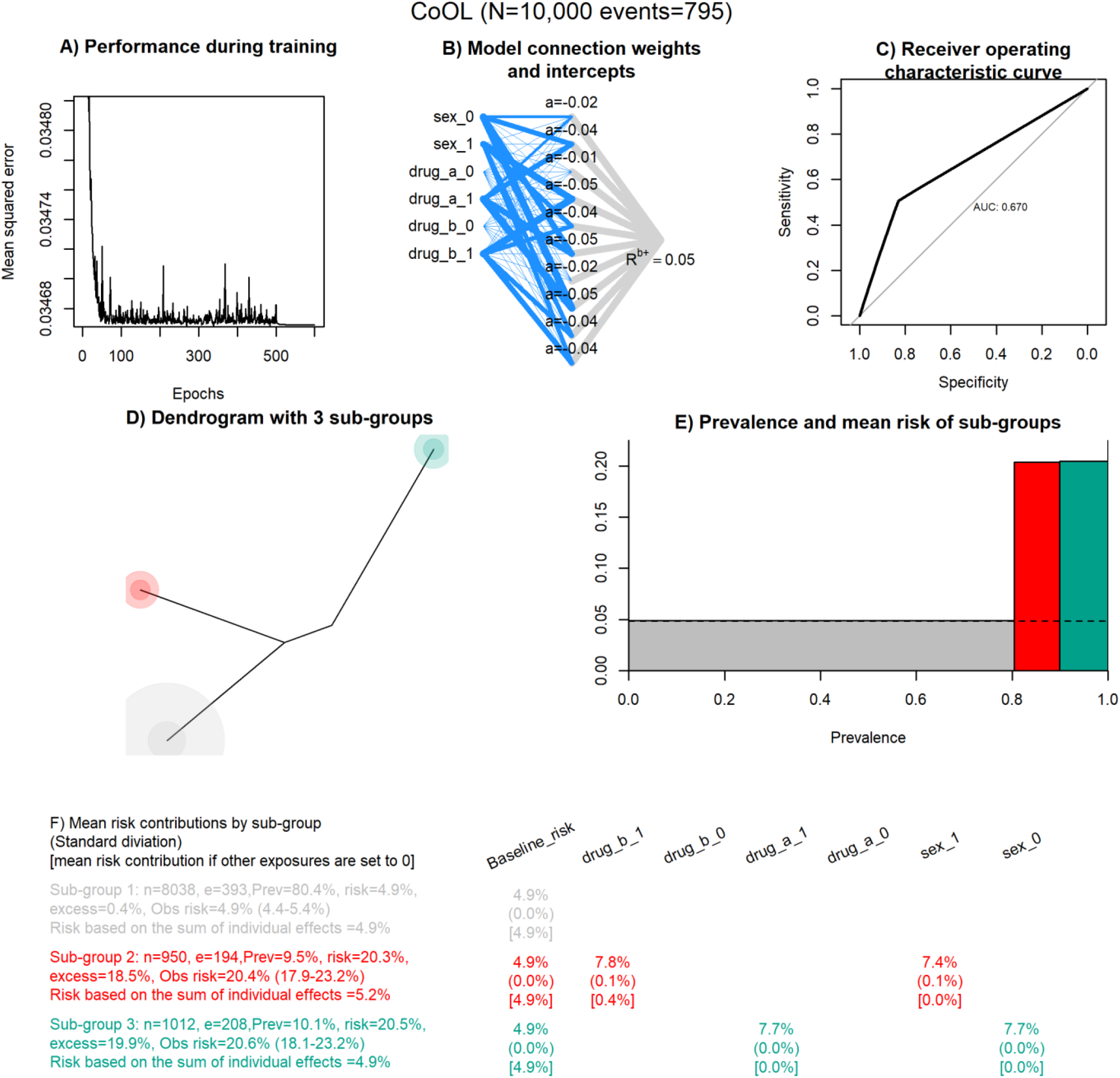
Results of the computational phase of CoOL. The main results are combined in one plot. A) Performance measured by the mean squared error by epoch. B) A visualisation of the fitted non-negative neural network. The width of the line indicate the strength of each connection. C) A plot on prediction performance as measured by a ROC curve. D) A dendrogram colored by 3 groups. E) the mean risk and prevalence by sub-groups. F) the table with the main results for the working example. “n” is the total number of individuals in the sub-group, “e” is the number of events / individuals with the outcome in the subgroup, “prev” is the prevalence of the sub-group, “risk” is the mean risk in the sub-group based on the model, “excess” is the excess fraction being the proportion out of all cases which are more than expected (more than the baseline risk) in this sub-group, “obs risk” is the observed risk in this sub-group (95% confidence interval is calculated using the Wald method in [74]), “risk based on the sum of individual effects” is the risk summed up where all other exposures are set to zero. For the 3 estimates presented at each variable by each sub-group, the first estimate is the mean risk contribution, the estimate in parentheses is the standard deviation, and the estimate in brackets is the risk contribution had all other exposures been set to zero. The baseline risk is by definition the same for all groups.

### Post-computational phase on hypothesis development and validation

The results of the computational step may provide learnings about different sets of exposures, which may have led to the outcome in different sub-populations. This evidence should be interpreted in the light of the assumed causal model that was specified in phase 1, and thus formulated into new hypotheses about multifactorial etiology, which may be denoted in a DAG as done by VanderWeele.[16] The empirical evidence from the computational phase highlights the outcome prevalence and risk distribution across population sub-groups and directs attention towards groups with a potentially large public health impact.

The domain experts will need to assess whether the causal assumptions are met across the identified sub-groups. Hypothetically, it may be that unmeasured confounding influenced our results based on our prior causal assumptions, which suggests that further work needs to be conducted to validate the findings and to understand how risk may be mitigated for the sub-groups. The major gain of using the CoOL approach compared to many other machine learning approaches is that the sub-groups can be defined by specific combinations of exposures that are easily communicated with words rather than by a black box algorithm. Hence, our learnings may be formulated as a hypothetical intervention and explored using established methodological frameworks for causal inference modelling.[6] Here, we can use the frameworks developed for synergistic effects of causes in a causal inference framework to draw our causal assumptions.[16]

Phase 3 for triangulating the hypotheses is conducted in external populations (either temporal validation or more desirably, external validation). If replicable, the researchers should provide sufficient evidence that the replicated finding is causal (and not due to similar bias structures) for example using various triangulation approaches with orthogonal bias structures (i.e. designs with biases in different directions) including studies outside the epidemiological field.[33] Eventually, if possible, the hypotheses needs to be tested using a randomized set-up.

In our example, we now have some learnings to inform two hypotheses: Men taking drug A seem to be at a higher-than-normal risk, women taking drug B seem to be at a higher-than-normal risk. We may supplement with observational data from other settings before we eventually may intervene (stop exposure to drug A for men, and drug B for women) possible in a randomized way if justified by *equipoise*\*.

## Discussion

We have introduced the CoOL approach, which investigates common combinations of exposures, which may have led to a specific health outcome. We have used a simple simulated example in the presentation, however, the approach applies to more complex scenarios (see additional simulations and a real life data analysis in the Supplementary material). New learnings can be formulated as hypotheses in words rather than a black box algorithm, and these hypotheses can subsequently be challenged and tested using for example the framework for hypothetical interventions and by triangulation.

So far, the sufficient cause model and the way of thinking about causes of an outcome have, *de facto*, mostly been a theoretical framework and not a practical approach for applied data analysis in epidemiology.[6] Though, there exists justifications of individual (n=1) explanation rather than group-based explanations by going from studying effects of causes to causes of an outcome,[34] our intention with the CoOL approach is to identify commonly shared sets of events, which are associated with higher risks of the outcome in specific sub-populations for public health interventions. Fully explaining an outcome seems far-fetched in epidemiology,[35] since these sets of events will interplay with multiple unknown or unmeasurable causes, but an approach like ours takes the first steps towards suggesting etiology[36] or – at least – to identify vulnerable subgroups.

The proposed approach may be of relevance to a number of theoretical frameworks: It links to the classical sufficient causes model[2], it may help disentangle structures in the *syndemics* (synergistic epidemics) literature,[37] and add a tool for holistic approaches to “precision” public health.[38] We stress that the CoOL approach is an inductive-deductive approach and that researchers in each of the phases need to carefully consider the most appropriate set-up that eventually may lead to fair public health actions.

### Limitations and extensions

#### Inference

Co-occurring associations between the exposures and the outcome can be due to various causal structures such as interactions,[31] clustered causes (exposures sharing a common cause), mediation,[31] uncontrolled confounding,[6] and conditioning on a common effect (collider-stratification or selection bias)[39] (Figure 5). *Interactions* (Figure 5A) entail a combined structural effect that is beyond the sum of the individual effects of the putative causes, and thus some inference about the underlying structures may be suggested by the CoOL approach and confirmed by formal interaction analysis. When studying non-randomized epidemiological data, complex combinations of all of these structures can be expected. Researchers will need to assess various hypotheses through triangulation to support their explanatory contribution.

**Figure 5.**
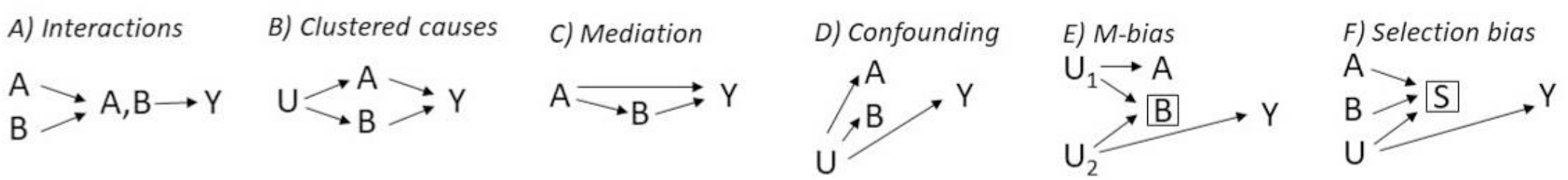
Six causal structures causing co-occurring associations. A and B denotes measured exposures of interest, U denotes an unmeasured cause of A and B, Y denotes the outcome, S denotes a selection mechanism. All six causal structures result in an increased co-occurrence of A and B in the Causes of Outcome Learning approach. It only applies for interactions that the combined effect is larger than the sum of the individual effects. A) Interaction -A and B jointly affect Y, and thus occur often together when assessing risk contributions (see also [31]). B) Clustered causes -A and B occur more often together due to U. C) Mediation -Since B is caused by A, A and B often occur together (see also [31]). D) Confounding -If U is a cause of A, B and Y, all variables occur often together (See also [6]). E) M-bias -Selection on B can cause a non-causal association between A and B, and A and Y (See also [39]). F) Selection bias -Conditioning on S creates a non-causal association between A, B and Y (See also [39]).

Rose described chains of causes by separating causes into distal and proximal causes.[3] Proximal causes, e.g. infectious agents, dietary deficiencies, smoking, toxic exposures and allergens, are close to the outcome in the causal chain, and distal causes, e.g. social and economic positions, as the causes of causes and thus are distal to the outcome in the causal chain. Such frameworks have been further expanded in the exposome literature.[40] The CoOL approach focuses on the most proximal causes of the causal chain, and thus included exposures should be carefully selected according to appropriate actionable exposures and contextual factors. An individualized focus on proximal causes may misdirect our attention away from structural public health interventions and could in the worst case scenario stigmatize parts of the population without offering preventive interventions.[41] Future work is needed to explore the degree of which bias is introduced due to collider bias by using a neural network.[16]

#### Model

The version of CoOL we have presented deals with binary exposures and outcomes, similar to the sufficient cause model.[7] However, the approach can be extended to continuous outcomes, where the value 0 has a meaningful interpretation (as e.g. loss of disease-free years, and in contrast to e.g. body mass index). Further, it may be that the CoOL approach reveals complementary information when studying positive outcomes (high quality of life) in comparison to negative outcomes (diseases and death).[42] Multiple other extensions of the CoOL approach may be possible, e.g. ways to incorporate time, such as time-varying variables, complex confounding scenarios, and censoring, would be of high relevance for epidemiological, and we encourage others to explore these. It could be of interest to explore a variation of outcome-wide approaches,[43] since co-morbidity may be a sign of shared underlying sufficient component causes (e.g. atopic diseases as asthma, dermatitis and nasopharyngitis), and thus analyzing several health outcomes may let us pick up suggestions of common sufficient causes otherwise missed (*multi-task learning*\*). The best presentation of the results will depend on the aim and extensions of the CoOL approach.

#### Robustness checks

If data are sparse for investigating the causal structure, the results may not be reliable both with the potential of type 1 (false positive findings) and type 2 errors (false negative findings). Robustness checks should be conducted to challenge the stability of the approach.[44] It can give insight to try the following to ensure that the baseline risk estimation and the identified groups are robust: change the number of synergy-functions (Supplementary simulation 7) re-run the analysis with sub-samples of the study population (Supplementary simulation 8), change the regularization values (Supplementary simulation 9). A baseline risk above its initial value, 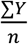, is a sign of model misspecification.

### Theoretical comparison with other approaches

Though not commonly applied in epidemiology, frameworks for identifying component causes exist by selection on either cases[9]or exposure[10]. In the social sciences, Configurational Comparative Methods deal with sufficient causes (referencing earlier work[45]), of which the most famous is the Qualitative Comparative Analysis,[46] which has also been applied in the public health domain.[47] Qualitative comparative analysis works by analyzing all combinations of exposures and uses a top-down search of exposure combinations which fulfill some chosen criteria, such as a risk threshold.[8] Using pre-defined risk thresholds may both have advantages such as transparent protocols and disadvantages such as being sensitive to the chosen threshold level with the risk of not identifying relevant risk groups with moderate increased risks but with large public health impact.

The CoOL approach has similarities to decomposition approaches of mediated and interactive effects in epidemiology,[48,49] however, work is needed to assess the similarities to the LRP properties in the CoOL approach. A recent approach, Algorithm for Learning Pathway Structures (ALPS), which uses a Monte Carlo scheme to update a pathway structure has shown promise for identifying complex interactions in large epidemiological datasets.[50] However, since ALPS focuses on parameter interpretation, its results differs from those of the CoOL approach, which identifies sizeable sub-groups who share exposures, which may have led to their increased risk of the outcome.

Approaches such as the exposome[40,51,52] and exposure- or environment-wide association studies (EWAS)[53,54] assess multiple exposures simultaneously, but few applied studies include interactions.[53–56] The few that do consider interactions tend to investigate interaction of pre-selected factors only.[57] Such studies have been discussed in relation to their potential, especially in light of successes of genome-wide association studies,[58] and limitations such as a challenging causal interpretation.[5]

LRP has previously been successfully demonstrated in image, text and biological data classification,[59–61] as well as for health records to explain clinical decisions on therapy assignment.[62] In this latter case, neither a baseline risk was estimated nor was there interest in identifying sub-groups. The computational phase of the CoOL approach has similarities to existing work on explaining and correcting computer vision,[63,64] but takes its depart from a causal question. The CoOL approach may be viewed as a supervised clustering approach based on an additive feature attribution method guided by a causally inspired model. It should be investigated to which degree other additive feature attribution methods approximate similar results,[65] since they are usually not related to any causal frameworks nor any underlying epidemiological theories. These methods seem to depart from a specified reference group in contrast to the CoOL approach, which estimates a baseline risk. Different model dependent methods for decomposing risks can naturally yield different estimates.[66]

Alternative methods to LRP for decomposing neural network predictions were proposed recently, such as DeepLIFT[67] or Integrated Gradients.[68] However, only LRP and its Deep Taylor Decomposition theoretical framework[26] fit our assumption of a non-negative neural network with negative biases, and allow for a seamless interpretation of relevance as risk contributions in our causal inference setup. Using non-negative models for sets of explanations within certain aims was proposed decades ago[69] but not in relation to causal questions. We did not want to consider sensitivity-, perturbation-, or surrogate-based explanation techniques, since our question of interest relates to the causes of an outcome posed as “*Given a particular health outcome, what are the most common sets of exposures, which might have been its causes?*” rather than effects of causes posed as “*What would have occurred if a particular factor were intervened upon and thus set to a different level than it in fact was?*” These distinctions have previously been discussed both in the causal inference literature[7] and in the literature on LRP.[12] Furthermore, perturbation-based methods produce localized explanations which may not generalize to global causal pathways.[59,70,71]

## Conclusion

We have introduced the Causes of Outcome Learning approach with the aim of disentangling common combinations of pre-outcome exposures that could have caused a specific health outcome. The approach is based on prior knowledge of the causal structure, the flexibility of a non-negative neural network, the LRP explanation technique for decomposing risk contributions and clustering, and, finally, hypothesis development and testing. These are steps towards building better transparency and causal reasoning into hypothesized causal findings from machine learning methods in the health sciences.[72,73] This CoOL approach should encourage and enable epidemiologists to examine common combinations of exposures as causes of the outcome of interest. This could eventually inform the development of more effective, targeted and impactful public health interventions.

## Supporting information

Supplementary results 1 - Repeated

Supplementary results 2 - Noise variables

Supplementary results 3 - Complex example

Supplementary results 4 - Common causes

Supplementary results 5 - Mediation

Supplementary results 6 - Confounding

Supplementary results 7 - Synergy functions

Supplementary results 8 - Data size

Supplementary results 9 - Regularization

## Data Availability

Simulated data can be generated using the provided R package "CoOL". Access to the real life data is explained in the Supplementary Material page 21.

## Acknowledgment

The authors would like to thank the colleagues at Section of Epidemiology, Department of Public Health, University of Copenhagen for valuable comments and suggestions on the idea throughout the development. The authors are grateful to Rasmus Wibæk Christensen, Stine Byberg and Douglas Ezra Morrison for comments on the manuscript, to Tue Kjærgaard Nielsen for assistance with the implementation of dendrograms, and to Thorkild IA Sørensen for guidance on literature regarding body mass index, blood pressure and mortality.

