## Supplementary material for "Causes of Outcome Learning: A causal inference-inspired machine learning approach to disentangling common combinations of potential causes of a health outcome"

### Supplementary materials
### Causes of Outcome Learning

#### A causal inference-inspired machine learning approach to disentangling common combinations of potential causes of a health outcome

A Rieckmann[1], P Dworzynski[2], L Arras[3], S Lapuschkin[3], W Samek[3], OA Arah[4], NH Rod[1], CT Ekstrøm[5]

[1] Section of Epidemiology, Department of Public Health, University of Copenhagen

[2] Novo Nordisk Foundation Center for Basic Metabolic Research, University of Copenhagen

[3] Machine Learning Group, Department of Video Coding & Analytics, Fraunhofer Heinrich Hertz Institute

[4] Department of Epidemiology, Fielding School of Public Health, University of California, Los Angeles and Department of Statistics, UCLA College of Letters and Science, Los Angeles

[5] Section of Biostatistics, Department of Public Health, University of Copenhagen

Pre-print (not peer reviewed nor accepted by a journal)

Supplementary table 1. Glossary

| Term used | Description |
|---|---|
| Additive | Effects are modeled in an additive space. A frequent additive model used is a linear regression. In contrast, a logistic regression is not additive. In an additive model, a synergistic interaction can be defined as a combined effect which is larger than the sum of the independent effects. |
| The backdoor path criterion | A spurious association between two variables due to a common cause, can be adjusted for by including any variable on the non-causal pathway through the common cause. |
| Baseline risk | The assumed risk for everyone given the unknown and not measured causes of the outcome shared by the full study population. |
| Binary/one-hot encoding | Called binary encoding or one-hot encoding. All categorical variables are split into several binary variables, which reference group is interpreted as the category is not present, thus introducing dependency between these binary variables. |
| Causal assumptions | Assumptions to be met such that associations can be interpreted as average causal effects. |
| Causal inference | Inference about causal relations based on the observed data and assumptions about the causal structure of data. Various closely linked frameworks exist such as the counter factual framework and the potential outcome framework. |
| Confounders | We used the word confounders pragmatically, though only the term confounding is strictly relevant in causal inference. |
| Derivatives | Training is done using derivatives of the error function for each given data point. |
| Epoch | One complete training iteration through all individuals is called an epoch. |
| Equipoise | We are uncertain whether intervening is better than not intervening. |
| Initial values | The initial neural network will be given random initial parameters (connection weights and intercepts) before any training on data had been done. |
| Learning rates | The pace of which training is done. The learning rate is reduced during training for the parameters to converge. |
| Machine learning | Algorithms that learn patterns in data. Models with high flexibility / degree of freedom. The term may cover a large variety of models fitted to data – usually with the ability to incorporate nonlinearites. It covers models such as logistic regressions, random forest, deep neural networks etc. |
| Manhattan distances | The sum of all absolute differences between risk contributions between all combinations of individuals observed. Risk contribution differences will be summed, thus if two variable express the same risk, their risk contribution differences will be similar to only having included one of the variables. |
| Monotonicity assumption | For an exposure, x, assumes that it has either no effect or its effect always takes the same direction on Y. |
| Multi-task learning | The improvement of task specific models by training one model to simultaneously solve more than one task. |
| Neural networks | Artificial neural network-models are inspired by brain synapses, and allows information from inputs to the outcome to go through various paths, which are found by fitting the model to data. A neural network can be viewed as a parallel and sequential expansion of well-known models in epidemiology. The paralleled and sequential expansion makes it difficult to interpret the model parameters directly. This is why an explanation method for neural networks (such as LRP) is needed. |
| Non-negative model | A model with only positive parameters of each input. Such models may facilitate monotonicity assumptions. |
| Overfitting | A model is fitted to chance findings in the observed data such that it is not generalizable to other data sets. The concept relates to chance findings, type 1 errors, false positive and negative findings, lack of external validity, and lack of transportability. |
| Regularization | Forced reduction in the parameters – typically to prevent extreme parameters or to aid in feature selection. Regularization is also used in e.g. lasso functions. |
| Risk contributions | The original term for risk contributions in the LRP literature is relevance measures, but given our causal model assumptions, we can now interpret relevance measures as risk contributions. |
| Signal-to-noise ratio | There exists a balance between the signal in data and the amount of input variables included. If too many noisy input variables are included the noise may mask a true effect of an exposure. |
| Single-hidden layer | In neural networks, this defined the number of hidden layers between the exposures and the outcome Can be viewed as latent constructs |
| Stochastic gradient decent | During an iterative learning process, the error of the model is reduced for each individual at a time, to develop eventually a model with a lower average error. |
| Synergy-function | A “Synergy-function” is not a machine learning term. We use the synergy functions to allow that some exposures work together and cause higher risks than expected by their individual effects. The operation $f(x) = \max(0,x)$ is also sometimes called Rectified Linear Units (ReLU) in the neural network literature. The synergy functions can return linear effects if the negative intercepts are close to zero. |
| Test data | In this paper, we define test data as data to manually confirm the observations on before developing hypotheses. Normally a test data set can be used for stopping the training of the model, when the model is indicated to overfit to the training data. |
| Training data | The data that the model is fitted to. |
| Unobserved layer in neural networks | Normally called a hidden layer. |
| Validation data | In this paper, a validation data set is the data set saved for testing the developed hypotheses. It may be a part of the original data set or data from very different sources but where the hypotheses can be triangulated on. |
| Ward method | One of many approaches to cluster distance measures. The Ward method balances the size of the sub-groups too when assigning sub-groups. It centers around centroids and penalizes large sub-groups, which also results in larger sizes of the smallest subgroups, thus all sub-groups will have a certain potential public health impact. |

Preprint (not peer reviewed nor accepted by a journal)

#### Supplementary information 1. Installation of the R package ‘CoOL’

We provide the R package ‘CoOL’ for the computational phase, which can be installed in R using the commands:

```
if(!require(“devtools”)) install.packages(“devtools”)
devtools::install_github(‘ekstroem/CoOL’)
```

Pre-print (not peer reviewed nor accepted by a journal)

#### Supplementary information 2. Initial values, derivatives, learning rates, and regularization

$n$ is the sample size, the predicted risk, $P(Y|X^+)$ is denoted $O$, the true outcome is denoted $Y$, the error is estimated as $\frac{(O-Y)^2}{2}$ and denoted $E$, the value taken by each of the hidden synergy-functions $S_j^+()$ activation functions are denoted $S_j$, the input value to $S_j^+()$ activation function values are denoted $s_j$, the connection parameters from the exposures to the synergy are denoted $\beta_{i,j}^+$, the intercepts for the hidden $S_j^+()$ activation functions are denoted $\alpha_j^-$, the baseline risk is denoted $R^{b+}$, and the exposures are denoted $X_i^+$. $\lambda$ denotes to which degree the input parameters are regularized with at each iteration.

| | Initial values | Derivative | Learning rate | Constrains | Regularization | Reasoning |
|---|---|---|---|---|---|---|
| $\beta_{i,j}^+$ | Gamma distribution with a rate parameter of 1 and a shape parameter of 100. | $\frac{\delta E}{\delta \beta_{i,j}^+} = \frac{\delta E}{\delta O} \cdot \frac{\delta O}{\delta S_j} \cdot \frac{\delta S_j}{\delta s_j} \cdot \frac{\delta s_j}{\delta \beta_{ij}}$ | learning rate | $\geq 0$ | - learning rate * $\lambda$, where the default $\lambda$ is $10^{-4}$, but should be adjusted balancing noise signals and false negative findings | We initiate the values close to zero (but positive), to assume we have no known knowledge of the directions for which the effects will take. |
| $\alpha_j^-$ | Gamma distribution with a rate parameter of 1 and a shape parameter of 100. | $\frac{\delta E}{\delta \alpha_j^-} = \frac{\delta E}{\delta O} \cdot \frac{\delta O}{\delta S_j} \cdot \frac{\delta S_j}{\delta s_j} \cdot 1$ | learning rate | $\leq 0$ | None | We initiate the values close zero (but negative) as the intercepts will function as gate keepers in the synergy-functions. |
| $R^{b+}$ | $\frac{\sum Y}{n}$ | $\frac{\delta E}{\delta R^{b+}} = \frac{\delta E}{\delta O} \cdot 1$ | learning rate / 10 | $\geq 0$ | None | We initiate the baseline risk as the mean risk of the outcome, as if we had no information from the exposures. |

Where: $\frac{\delta E}{\delta O} = O - Y, \frac{\delta O}{\delta S_j} = 1, \frac{\delta S_j}{\delta s_j} = 1$ if $S_j > 0$ otherwise 0, $\frac{\delta s_j}{\delta \beta_{i,j}} = X_i^+$

Pre-print (not peer reviewed nor accepted by a journal)

#### Supplementary information 3. Estimating the risk based on the sum of individual effects

If we denote the combined risk as $P(Y = 1|X)$, the risk based on the sum of the individual effects can be estimated as:

$$R^{b+} + \sum_i (P(Y = 1|\mathrm{X}_i = x_i, X_{!i}^{+} = 0) - R^{b+})$$

Where $R^{b+}$ is the baseline risk, $x_i$ is the actual value taken by $X_i$, and $!\,i$ denotes all but $i$.

Pre-print (not peer reviewed nor accepted by a journal)

#### Supplementary information 4. Excess fraction

The excess fraction or formally the attributable proportion in the population[1] for subgroup Z can be defined as $\frac{P(Y=1)-P(Y_{X_z=\overline{X_z}}=1)}{P(Y=1)}$ and can be calculated as follows:

$$\frac{\mathrm{P(X = x_z)} \cdot \left(\mathrm{P(Y = 1|X = x_z) - R^{b+}}\right)}{\mathrm{P(Y = 1)}}$$

Argumentation: Given the combined risk contributions causally affect the outcome and meet the assumption of positive monotonicity, we are interested in estimating how much the area marked in the bold dashed rectangle (the numerator) constitutes out of all the shaded areas (the denominator)  in below figure showing the prevalence and mean risk by sub-groups. The area of the bold dashed rectangle (the numerator) can be estimated by the product of each orthogonal line, which is equal to the proportion exposed, $\mathrm{P(X = x_z)}$, times the causal effect, $\mathrm{P(Y = 1|X = x_z) - R^{b+}}$. All of the shaded area (the denominator) is equal to $\mathrm{P(Y = 1)}$.

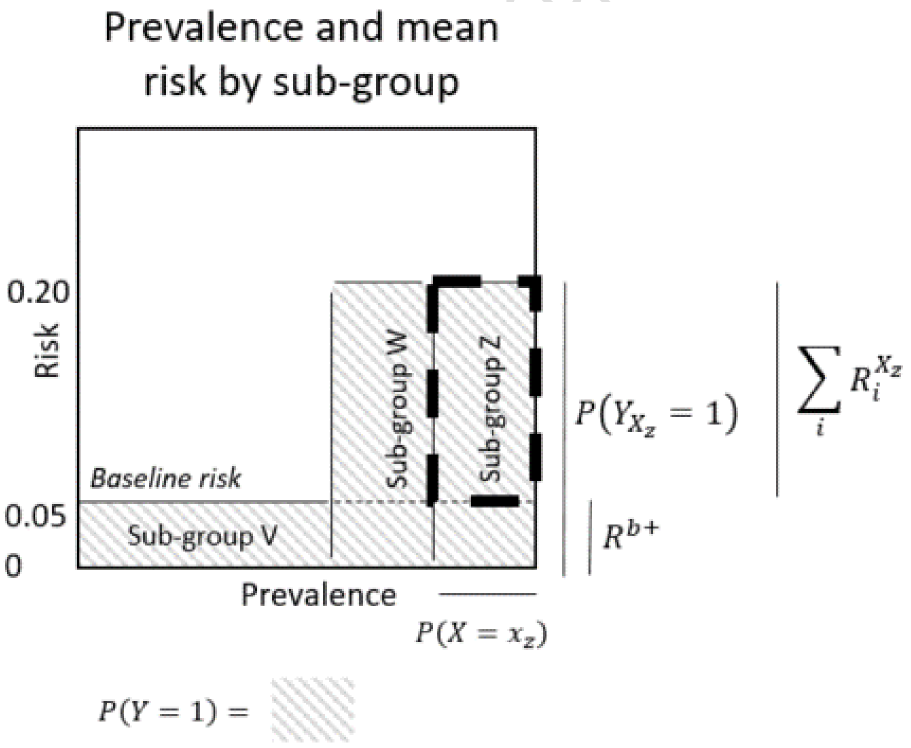

---

[1] Suzuki, Etsuji, Eiji Yamamoto, and Toshihide Tsuda. "On the relations between excess fraction, attributable fraction, and etiologic fraction." *American journal of epidemiology* 175.6 (2012): 567-575.

#### Supplementary simulation 1. Working example with tutorial

With below bit, the presented motivating example can be replicated. Use View(function_name) to explore the underlying code, as modifications may be needed to better present the specific research question. In the subsequently "Step-by-step tutorial" we explain each line. In Supplementary results 1, we present the results of 10 simulations which all give consistent estimates.

```
library(CoOL)
data <- CoOL_0_working_example(n=10000)
outcome_data <- data[,1]
exposure_data <- CoOL_0_binary_encode_exposure_data(data[,-1])
model <- CoOL_1_initiate_neural_network(inputs=ncol(exposure_data), output = outcome_data)
model <- CoOL_2_train_neural_network(X_train=exposure_data, Y_train=outcome_data, model=model)
plot(model$train_performance,type='l',yaxs='i',ylab="Mean squared error", xlab="Epochs",
main="Performance - training data")
CoOL_3_plot_neural_network(model,names(exposure_data))
CoOL_4_AUC(outcome_data,exposure_data,model)
risk_contributions <- CoOL_5_layerwise_relevance_propagation(exposure_data,model)
CoOL_6_dendrogram(risk_contributions,number_of_subgroups = 3)
sub_groups <- CoOL_6_sub_groups(risk_contributions,number_of_subgroups = 3)
CoOL_7_prevalence_and_mean_risk_plot(risk_contributions,sub_groups)
CoOL_8_mean_risk_contributions_by_sub_group(risk_contributions, sub_groups)
```

**Step-by-step tutorial**

We read the R package (see Supplementary information 1 for installation).

```
library(CoOL)
```

We then generate data for the motivating example. The larger the data sample, the more consistent the estimations.

```
data <- CoOL_0_working_example(n=10000)
```

We identify the outcome, and exposures. The exposures are binary encoded, such that we generate a new variable for each category value each exposure may take.

```
outcome_data <- data[,1]
exposure_data <- CoOL_0_binary_encode_exposure_data(data[,-1])
```

We initiate the model and train it.

```
model <- CoOL_1_initiate_neural_network(inputs=ncol(exposure_data), output = outcome_data)
model <- CoOL_2_train_neural_network(X_train=exposure_data, Y_train=outcome_data, model=model)
```

During training the following screens for monitoring the process are showed, which shows the mean squared error by epoch, the log of the mean squared weight difference between each epoch, and the estimated baseline risk by epoch. The mean squared error in the first plot should decrease, the mean squared weight differences should converge and naturally drops when the learning rate is decreased, and the baseline risk should converge.

Pre-print (not peer reviewed nor accepted by a journal)

If the baseline risk drops towards zero, this is an indication of too large noise to signal ratio, and more regularization may be needed or data may be too sparse for the number of included features.

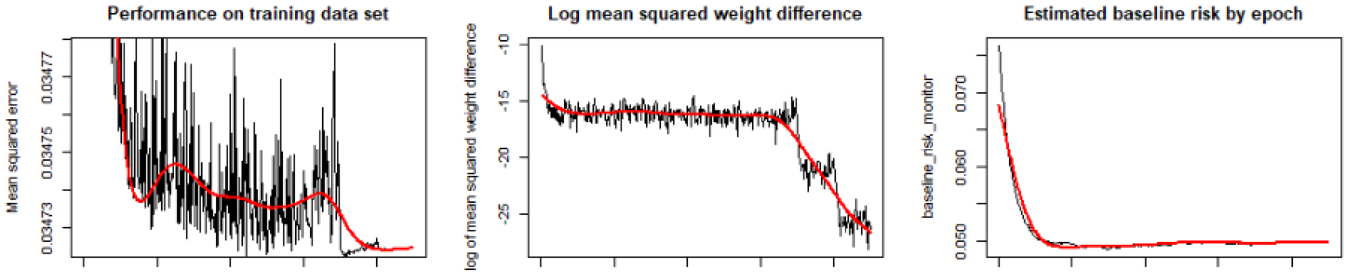

The following plots gives the increase in performance, a visualization of the model's connection weights and intercepts, and the ROC plot.

```
plot(model$train_performance,type='l',yaxs='i',ylab="Mean squared error", xlab="Epochs",
main="Performance - training data")
CoOL_3_plot_neural_network(model,names(exposure_data))
CoOL_4_AUC(outcome_data,exposure_data,model)
```

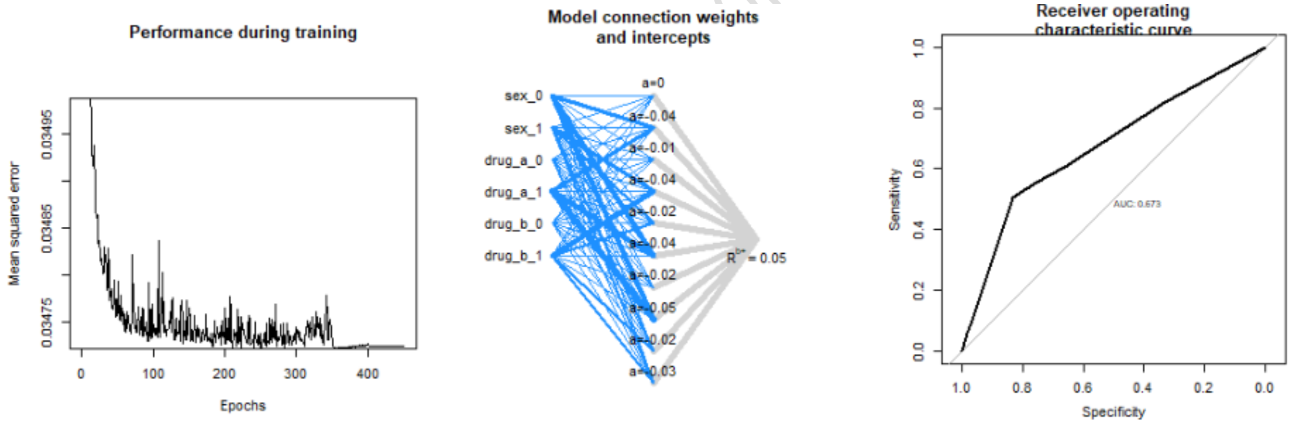

Pre-print (nc
or accepted by a jou

We then calculate the risk contributions and plot dendrograms. Each of the below plots are colored by different numbers of sub-groups, while the dendrogram stays static as it is independent of the number of chosen sub-groups.

```
risk_contributions <- CoOL_5_layerwise_relevance_propagation(exposure_data,model)
CoOL_6_dendrogram(risk_contributions,number_of_subgroups = 2)
CoOL_6_dendrogram(risk_contributions,number_of_subgroups = 3)
CoOL_6_dendrogram(risk_contributions,number_of_subgroups = 4)
```

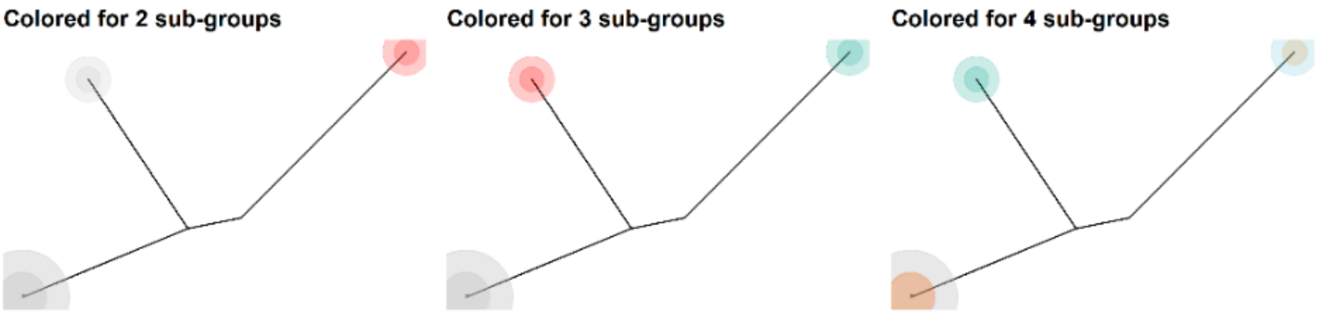

Based on the dendrogram, we decide on 3 sub-groups:

```
sub_groups <- CoOL_6_sub_groups(risk_contributions,number_of_subgroups = 3)
```

We then plot the prevalence and mean risk plot

```
CoOL_7_prevalence_and_mean_risk_plot(risk_contributions,sub_groups)
```

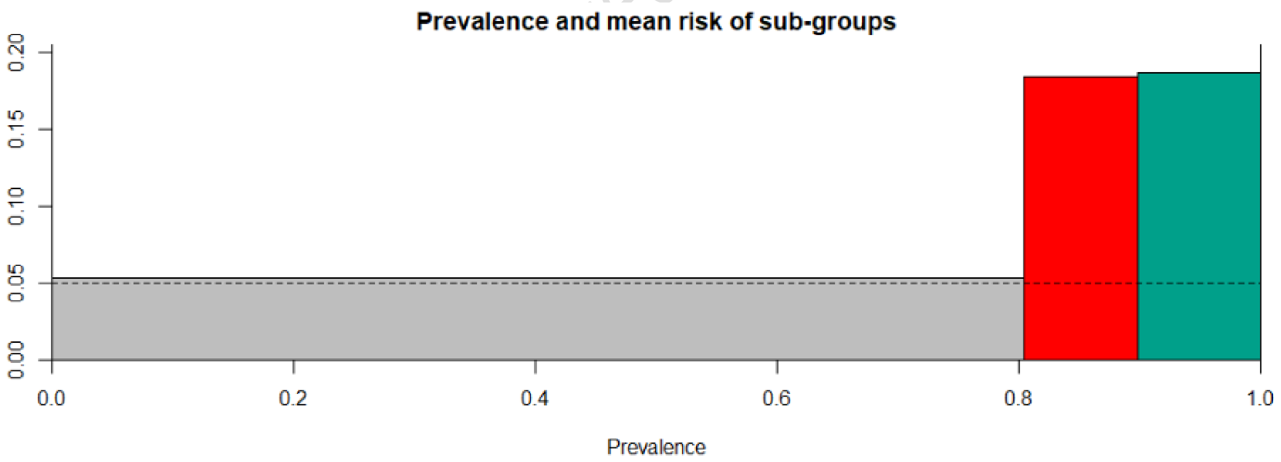

We can see that the grey group has a similar risk as the baseline risk, and that the red and green group has a risk close to 20% percentage.

We use the following function to show the mean risk contributions by sub-groups indicating that men who take drug A have a 15% increased risk of the outcome, and so does women who take drug B. We also see that this is indicated to be more than the sum of the individual effects, and that approximately equal risk contributions come from sex and the drugs.

```
CoOL_8_mean_risk_contributions_by_sub_group(risk_contributions, sub_groups)
```

| F) Mean risk contributions by sub-group<br>(Standard diviation)<br>[mean risk contribution if other exposures are set to 0] | Baseline_risk | drug_b_1 | drug_b_0 | drug_a_1 | drug_a_0 | sex_1 | sex_0 |
|---|---|---|---|---|---|---|---|
| Sub-group 1: n=8038, e=393,Prev=80.4%, risk=5.3%, excess=3.4%, Obs risk=4.9% (4.4-5.4%)<br>Risk based on the sum of individual effects =5.3% | 5%<br>(0.0%)<br>[5.0%] | | | | | | |
| Sub-group 2: n=950, e=194,Prev=9.5%, risk=18.4%, excess=16.1%, Obs risk=20.4% (17.9-23.2%)<br>Risk based on the sum of individual effects =6.8% | 5%<br>(0.0%)<br>[5.0%] | 7.2%<br>(0.0%)<br>[1.6%] | | | | 6%<br>(0.0%)<br>[0.0%] | |
| Sub-group 3: n=1012, e=208,Prev=10.1%, risk=18.7%, excess=17.5%, Obs risk=20.6% (18.1-23.2%)<br>Risk based on the sum of individual effects =6.3% | 5%<br>(0.0%)<br>[5.0%] | | | 6.8%<br>(0.0%)<br>[0.8%] | | | 6.5%<br>(0.0%)<br>[0.2%] |

Pre-print (not peer reviewed nor acce

#### Supplementary simulation 2. Working example with increasing number of irrelevant exposures

We ran simulations with increasing number of randomly flipped binary variables ∈ $[0, 5, 10, \ldots 50]$ added to the exposure variables in our motivating example (Supplementary results 2). Due to memory issues, we reduced the number of individuals to 10,000 and used 10 synergy functions. We see that the CoOL approach is able to identify the sub-groups even with 50 noise variables included in the analysis as seen below. Too many noise variables is suspected at some point to decrease the signal-to-noise too much.

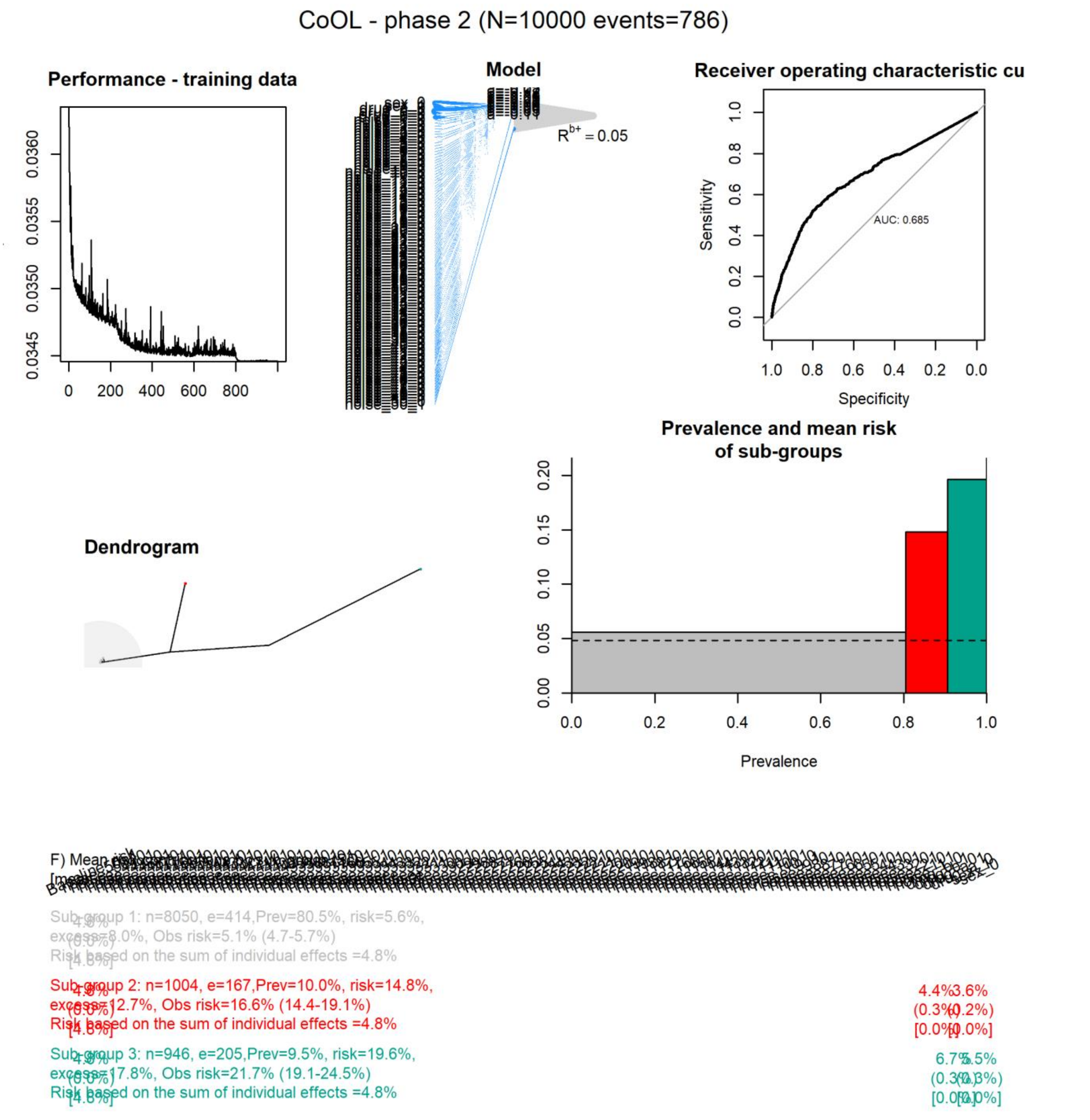

#### Supplementary simulation 3. Complex scenario

Let us consider a fictive population of 50,000 individuals. The data is generated according to Figure A. Simulating a real-life setting, we aim at identifying the different exposures which may have led to the outcome. Given the way data is generated, we should ideally be able to identify the following exposures which together caused the outcome: **Cause combination 1** with: Not *physically active* and *low-density lipoprotein (LDL)* and *night shift*, and **Cause combination 2** with: *Mutation X* and *air pollution*. As a result, the example data includes synergism; no cause acts alone, but depends on other measured and unmeasured exposures to become sufficient to cause disease Y. Low socio-economic status (SES), genes and living area act through mediators. U denotes an unmeasured exposure.

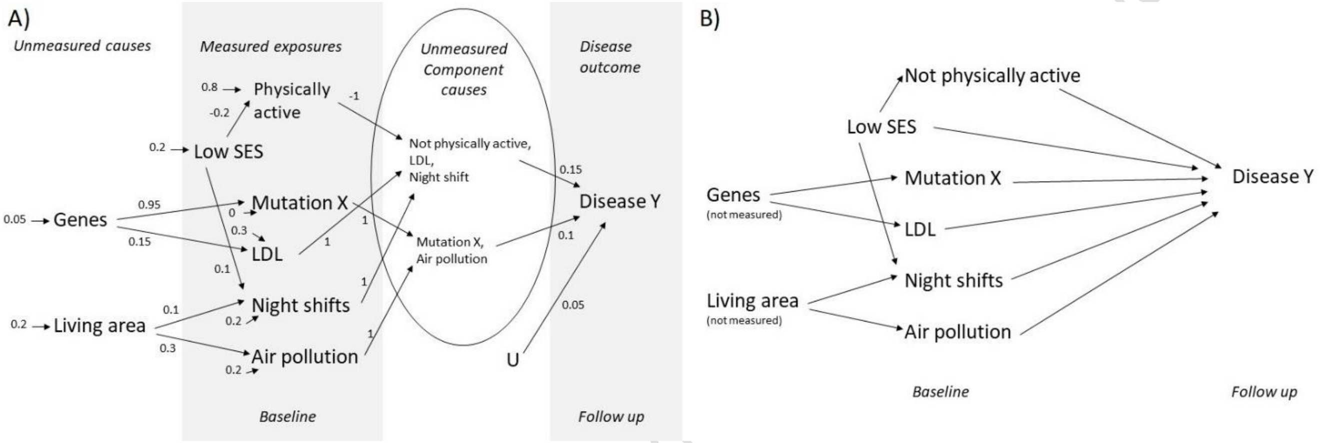

**A) Data generating process. Each arrow between factors shows the causal direction and the risk difference the exposure affects the outcome with. E.g. the background probabligy of being physical active is 80%, which is reduced by 20% if one has a low socio economic position. B) Directed Acyclic Graph based on prior knowledge for this example.**

The theoretical expectations to this simulation is:

$$P(Y|U) = 0.05$$

$$P(Y|MutationX, Airpollution) = 0.1$$

$$P(MutationX, Airpollution) = (0.05 \cdot 0.95) \cdot (0.2 \cdot 0.3 + 0.2) = 0.01235$$

$$P(Y|NotPhysicallyActive, LDL, NightShift) = 0.15, \;\; P(Y|NotPhysicallyActive, LDL, NightShift) = 0.15$$

$$P(NotPhysicallyActive, LDL, NightShift) = \left(1 - (0.8 - 0.2 \cdot 0.2)\right) \cdot (0.3 + 0.05 \cdot 0.15) \cdot (0.2 + 0.2 \cdot 0.1 + 0.2 \cdot 0.1) = 0.017712$$

Using complementary rules, we can calculate Y as 1 minus the probability of not having the outcome:

$$P(Y) = 1 - \left(1 - P(Y|U)\right) \cdot \left(1 - P(Y|MutationX, Airpollution) \cdot P(MutationX, Airpollution)\right) \cdot \left(1 - P(Y|NotPhysicallyActive, LDL, NightShift) \cdot P(NotPhysicallyActive, LDL, NightShift)\right) = 0.0536941$$

Pre-print (not peer reviewed journal)

$$Excess\ fraction\ (Mutation\ X, Airpolltion) = \frac{P(Y|MutationX, Airpollution) \cdot P(MutationX, Airpollution)}{P(Y)} = 0.0230007$$

$$Excess\ fraction\ (Not\ physically\ active, LDL, night\ shift) = \frac{P(Y|NotPhysicallyActive, LDL, NightShift) \cdot P(NotPhysicallyActive, LDL, NightShift)}{P(Y)} = 0.0494803$$

Our data simulation included relatively rare outcomes with a high degree of uncertainty. As a result, the baseline risk is considerable and risks are far from certain if individuals have any of the cause combinations. Due to the data generating mechanisms, our expectations to data are that 1.8% of the study population are exposed to the cause combination 1: Not *Physically active* and *LDL* and *Night shift*, which increases the risk by 15%. We expect that this combination of exposures contribute to 4.9% excess cases. The cause combination 2: *Mutation X* and *Air pollution* has a prevalence of 1.2% in the study population and increases the risk of the disease by 10%. This combination of exposures contribute to 2.3% excess cases.

In this example, we use a regularization parameter of $10^{-5}$ for the CoOL approach. The dendrogram suggests 3 groups. The results of clustering the risk contribution matrix to 3 sub-groups are shown in the plot visualizing sub-groups' prevalences and risks as well as in the table with the mean risk contributions by sub-group. First, we find that that the baseline risk is estimated to the expected 0.05. We also find that the correct sets of combination of exposures are identified (Sub-group 2 with: Not *Physically active* and *LDL* and *Night shift*. Sub-group 3 with: *Mutation X* and *Air pollution*). The model-based predicted risk is close to the expected risk from the data generating process, but also the measured risk in each cluster, and the "obs risk" being the risk in the assigned sub-group. Not only are the component causes identified, but also the proportion of cases attributed to these causes, as seen under "excess". We also calculate, in brackets, what would be the risk contribution had all other values been zero.

In conclusion, the CoOL approach identifies the sub-groups in this simulated example.

Pre-print (not peer reviewed nor accepted by a journal)

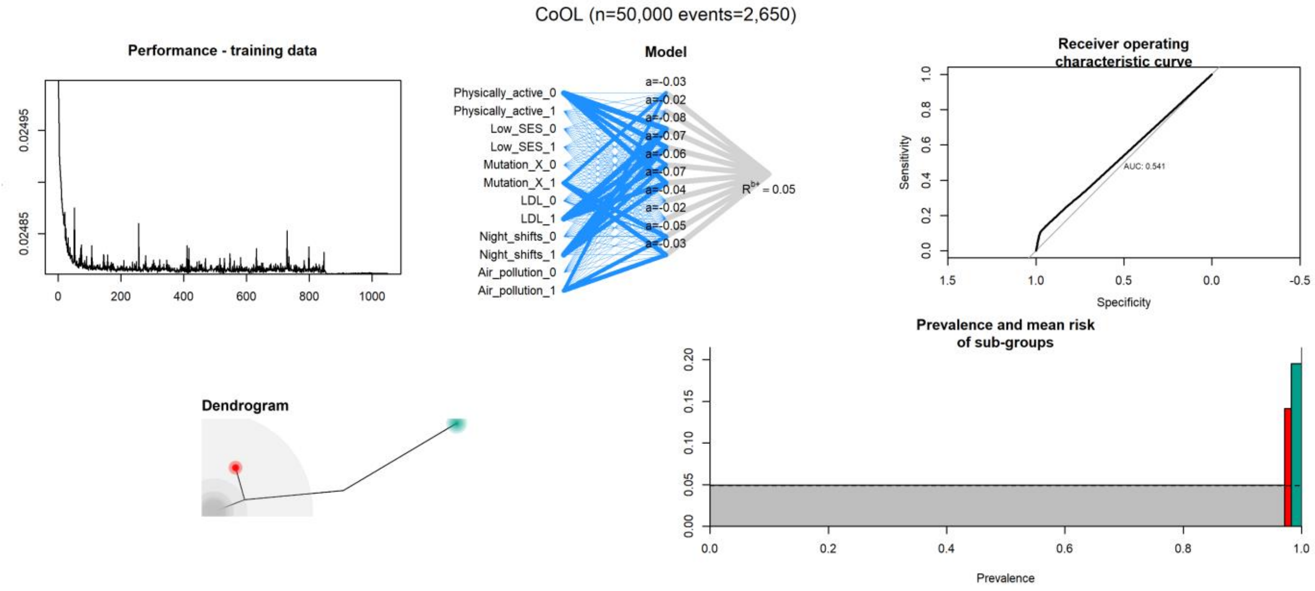

F) Mean risk contributions by sub-group
(Standard diviation)
[mean risk contribution if other exposures are set to 0]

| | Baseline_risk | Air_pollution_1 | Air_pollution_0 | Night_shifts_1 | Night_shifts_0 | LDL_1 | LDL_0 | Mutation_X_1 | Mutation_X_0 | Low_SES_1 | Low_SES_0 | Physically_active_1 | Physically_active_0 |
|---|---|---|---|---|---|---|---|---|---|---|---|---|---|
| Sub-group 1: n=48564, e=2378,Prev=97.1%, risk=4.9%, excess=1.2%, Obs risk=4.9% (4.7-5.1%) Risk based on the sum of individual effects =4.9% | 4.9% (0.0%) [4.9%] | | | | | | | | | | | | |
| Sub-group 2: n=560, e=89,Prev=1.1%, risk=14.1%, excess=2.0%, Obs risk=15.9% (13.0-19.2%) Risk based on the sum of individual effects =4.9% | 4.9% (0.0%) [4.9%] | 4.1% (0.1%) [0.0%] | | | | | | 4.4% (0.2%) [0.0%] | | | | | |
| Sub-group 3: n=876, e=183,Prev=1.8%, risk=19.5%, excess=4.8%, Obs risk=20.9% (18.3-23.8%) Risk based on the sum of individual effects =4.9% | 4.9% (0.0%) [4.9%] | | | 4.8% (0.0%) [0.0%] | | 4.8% (0.0%) [0.0%] | | | | | | | 4.8% (0.0%) [0.0%] |

**A) The performance plot shows the decline in mean squared error by each epoch of training. B) The model plot visualises the parameters of the trained monotonistic neural network. C) The accuracy plot shows the area under the receiver operating characteristic. D) The dendrogram shows the similarity of risk contributions by individuals. E) The prevalence and mean risk of sub-groups plot gives an indication of which sub-group carries the greatest disease burden. F) The table of mean risk contributions by sub-group indicates which exposures elevate the risk for the specific groups. In the first column in the table; n is the number of individuals in the sub-group, e is the number of events in the sub-group, 'prev' is the prevalence of the sub-group, 'risk' is the estimated risk for the sub-group, 'excess' is the estimated proportion of cases above baseline in this sub-group, and 'obs risk' gives the 95% confidence interval for the risk of being a case in the subgroup estimated from the actual proportion of cases in the sub-group.**

Pre-print (not peer reviewed)

#### Supplementary simulation 4. Clustered causes

We simulate below causal structure. Each value at each arrow indicates the probability difference in a linear system. All variables are binary and initiated with a probability of being 1 with 30%, while Y is initiated with a probability of being 1 with a probability of 1%. In Supplementary results 4 are the results of 10 simulations (n=40000). A typical result looks as below. It is shown that the risk contribution when B and E are combined are similar to the expected risk contribution had all other exposures been set to zero as shown in the brackets.

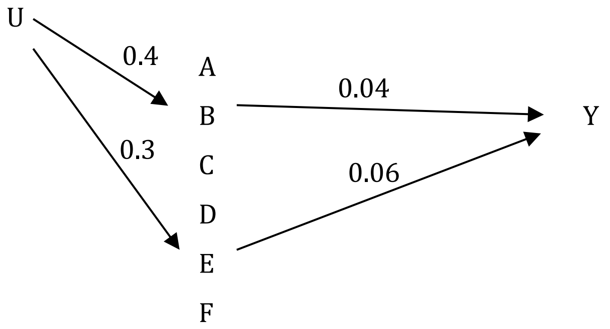

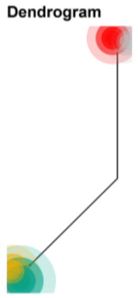

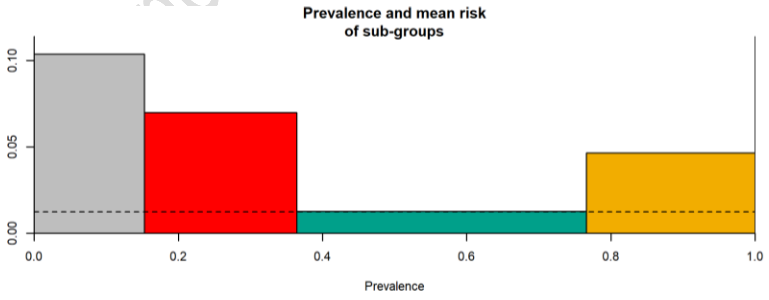

F) Mean risk contributions by sub-group (SD)

| | Baseline_risk | F_1 | F_0 | E_1 | E_0 | D_1 | D_0 | C_1 | C_0 | B_1 | B_0 | A_1 | A_0 |
|---|---|---|---|---|---|---|---|---|---|---|---|---|---|
| Sub-group 1: n=6110, e=653,Prev=15.3%, risk=10.4%, excess=29.8%, Obs risk=10.7% ( 9.9-11.5%)<br>Risk based on the sum of individual effects =10.3% | 1.3%<br>(0.0%)<br>[1.3%] | | | 5.7%<br>(0.0%)<br>[5.7%] | | | | | | 3.4%<br>(0.0%)<br>[3.4%] | | | |
| Sub-group 2: n=8465, e=612,Prev=21.2%, risk=7.0%, excess=26.0%, Obs risk=7.2% (6.7-7.8%)<br>Risk based on the sum of individual effects =7.0% | 1.3%<br>(0.0%)<br>[1.3%] | | | 5.7%<br>(0.0%)<br>[5.7%] | | | | | | | | | |
| Sub-group 3: n=16054, e=173,Prev=40.1%, risk=1.3%, excess=0.2%, Obs risk=1.1% (0.9-1.3%)<br>Risk based on the sum of individual effects =1.3% | 1.3%<br>(0.0%)<br>[1.3%] | | | | | | | | | | | | |
| Sub-group 4: n=9371, e=456,Prev=23.4%, risk=4.7%, excess=17.0%, Obs risk=4.9% (4.4-5.3%)<br>Risk based on the sum of individual effects =4.6% | 1.3%<br>(0.0%)<br>[1.3%] | | | | | | | | | 3.4%<br>(0.0%)<br>[3.4%] | | | |

Pre ... or accepted by a journal)

#### Supplementary simulation 5. Mediated causes

We simulate below causal structure. Each value at each arrow indicates the probability difference in a linear system. All variables are binary and initiated with a probability of being 1 with 30%, while Y is initiated with a probability of being 1 with a probability of 1%. In Supplementary results 5 are the results of 10 simulations (n=40000). A typical result looks as below. It is shown that the risk contribution when B and E are combined are similar to the expected risk contribution had all other exposures been set to zero as shown in the brackets. Only the direct effects of B and E are shown, thus the total effect of B is not identified.

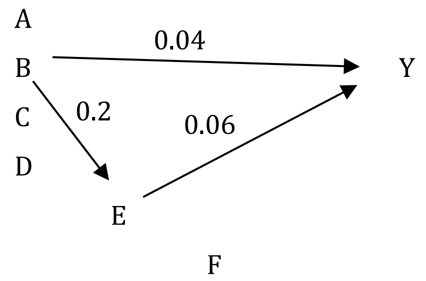

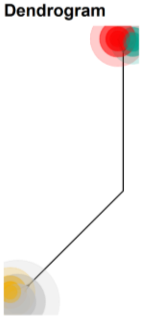

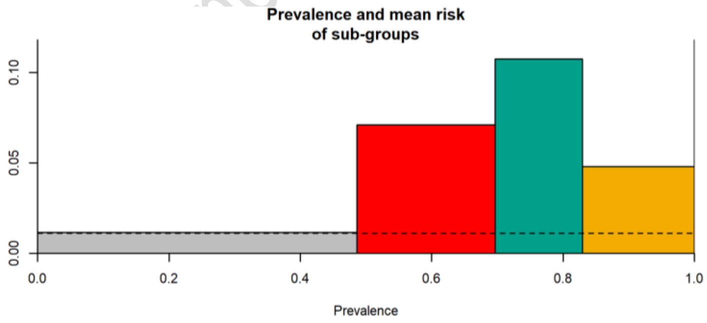

F) Mean risk contributions by sub-group (SD)

| | Baseline_risk | F_1 | F_0 | E_1 | E_0 | D_1 | D_0 | C_1 | C_0 | B_1 | B_0 | A_1 | A_0 |
|---|---|---|---|---|---|---|---|---|---|---|---|---|---|
| Sub-group 1: n=19447, e=206,Prev=48.6%, risk=1.2%, excess=0.7%, Obs risk=1.1% (0.9-1.2%) Risk based on the sum of individual effects =1.2% | 1.1% (0.0%) [1.1%] | | | | | | | | | | | | |
| Sub-group 2: n=8421, e=596,Prev=21.1%, risk=7.1%, excess=29.3%, Obs risk=7.1% (6.5-7.7%) Risk based on the sum of individual effects =7.1% | 1.1% (0.0%) [1.1%] | | | 5.9% (0.0%) [5.9%] | | | | | | | | | |
| Sub-group 3: n=5300, e=613,Prev=13.2%, risk=10.7%, excess=29.6%, Obs risk=11.6% (10.7-12.5%) Risk based on the sum of individual effects =10.7% | 1.1% (0.0%) [1.1%] | | | 5.9% (0.0%) [5.9%] | | | | | | 3.6% (0.0%) [3.6%] | | | |
| Sub-group 4: n=6832, e=326,Prev=17.1%, risk=4.8%, excess=14.7%, Obs risk=4.8% (4.3-5.3%) Risk based on the sum of individual effects =4.8% | 1.1% (0.0%) [1.1%] | | | | | | | | | 3.6% (0.0%) [3.6%] | | | |

or accepted by a journal)

### Supplementary simulation 6. Confounding

We simulate below causal structure. Each value at each arrow indicates the probability difference in a linear system. All variables are binary and initiated with a probability of being 1 with 30%, while Y is initiated with a probability of being 1 with a probability of 1%. In Supplementary results 6 are the results of 10 simulations (n=40000) where data is analysed with and without C. A typical result looks as below for each of the analytical approaches. When C is not included in the model, B and F are given risk contributions, however when C is included then only C contains risk contributions.

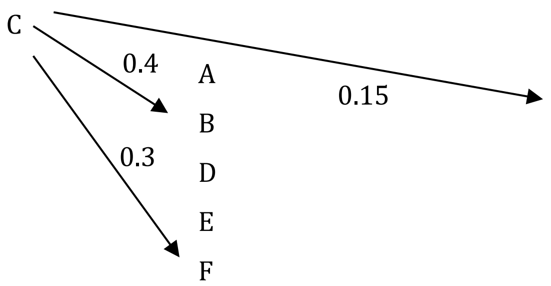

Without C

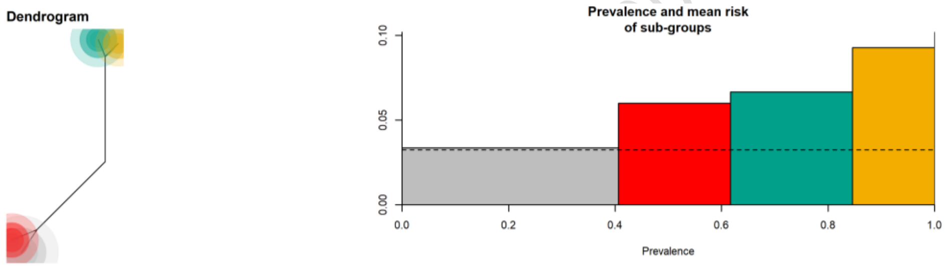

F) Mean risk contributions by sub-group (SD)
[mean risk contribution if other exposures are set to 0]

| | Baseline_risk | F_1 | F_0 | E_1 | E_0 | D_1 | D_0 | B_1 | B_0 | A_1 | A_0 |
|---|---|---|---|---|---|---|---|---|---|---|---|
| Sub-group 1: n=16245, e=525,Prev=40.6%, risk=3.4%, excess=0.9%, Obs risk=3.2% (3.0-3.5%) Risk based on the sum of individual effects =3.4% | 3.2% (0.0%) [3.2%] | | | | | | | | | | |
| Sub-group 2: n=8428, e=496,Prev=21.1%, risk=6.0%, excess=10.3%, Obs risk=5.9% (5.4-6.4%) Risk based on the sum of individual effects =6.0% | 3.2% (0.0%) [3.2%] | 2.6% (0.0%) [2.6%] | | | | | | | | | |
| Sub-group 3: n=9153, e=600,Prev=22.9%, risk=6.7%, excess=13.9%, Obs risk=6.6% (6.1-7.1%) Risk based on the sum of individual effects =6.6% | 3.2% (0.0%) [3.2%] | | | | | | | 3.3% (0.0%) [3.3%] | | | |
| Sub-group 4: n=6174, e=621,Prev=15.4%, risk=9.3%, excess=16.6%, Obs risk=10.1% ( 9.3-10.8%) Risk based on the sum of individual effects =9.3% | 3.2% (0.0%) [3.2%] | 2.6% (0.0%) [2.6%] | | | | | | 3.3% (0.0%) [3.3%] | | | |

With C

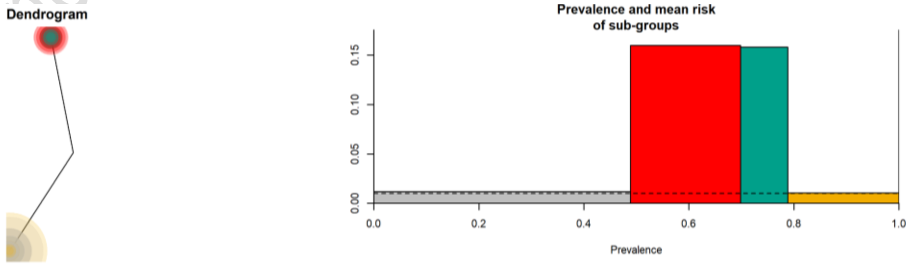

F) Mean risk contributions by sub-group (SD)
[mean risk contribution if other exposures are set to 0]

| | Baseline_risk | F_1 | F_0 | E_1 | E_0 | D_1 | D_0 | C_1 | C_0 | B_1 | B_0 | A_1 | A_0 |
|---|---|---|---|---|---|---|---|---|---|---|---|---|---|
| Sub-group 1: n=19547, e=212,Prev=48.9%, risk=1.2%, excess=1.4%, Obs risk=1.1% (0.9-1.2%) Risk based on the sum of individual effects =1.2% | 1% (0.0%) [1.0%] | | | | | | | | | | | | |
| Sub-group 2: n=8382, e=1404,Prev=21.0%, risk=16.0%, excess=56.1%, Obs risk=16.8% (16.0-17.6%) Risk based on the sum of individual effects =16.0% | 1% (0.0%) [1.0%] | | | | | | | 14.8% (0.0%) [14.8%] | | | | | |
| Sub-group 3: n=3609, e=548,Prev=9.0%, risk=15.8%, excess=23.9%, Obs risk=15.2% (14.0-16.4%) Risk based on the sum of individual effects =15.8% | 1% (0.0%) [1.0%] | | | | | | | 14.8% (0.0%) [14.8%] | | | | | |
| Sub-group 4: n=8462, e=78,Prev=21.2%, risk=1.0%, excess=0.0%, Obs risk=0.9% (0.7-1.2%) Risk based on the sum of individual effects =1.0% | 1% (0.0%) [1.0%] | | | | | | | | | | | | |

#### Supplementary simulation 7. Robustness check – synergy functions

We ran the motivating example-simulation with increasing number of synergy functions ∈ [1,2,3,4,5,10, … , 50] (Supplementary results 7). As expected, we see that the CoOL approach fails to identify the sub-groups when there are too few synergy functions such as 1 as seen below. The results are stable also with 20 synergy functions.

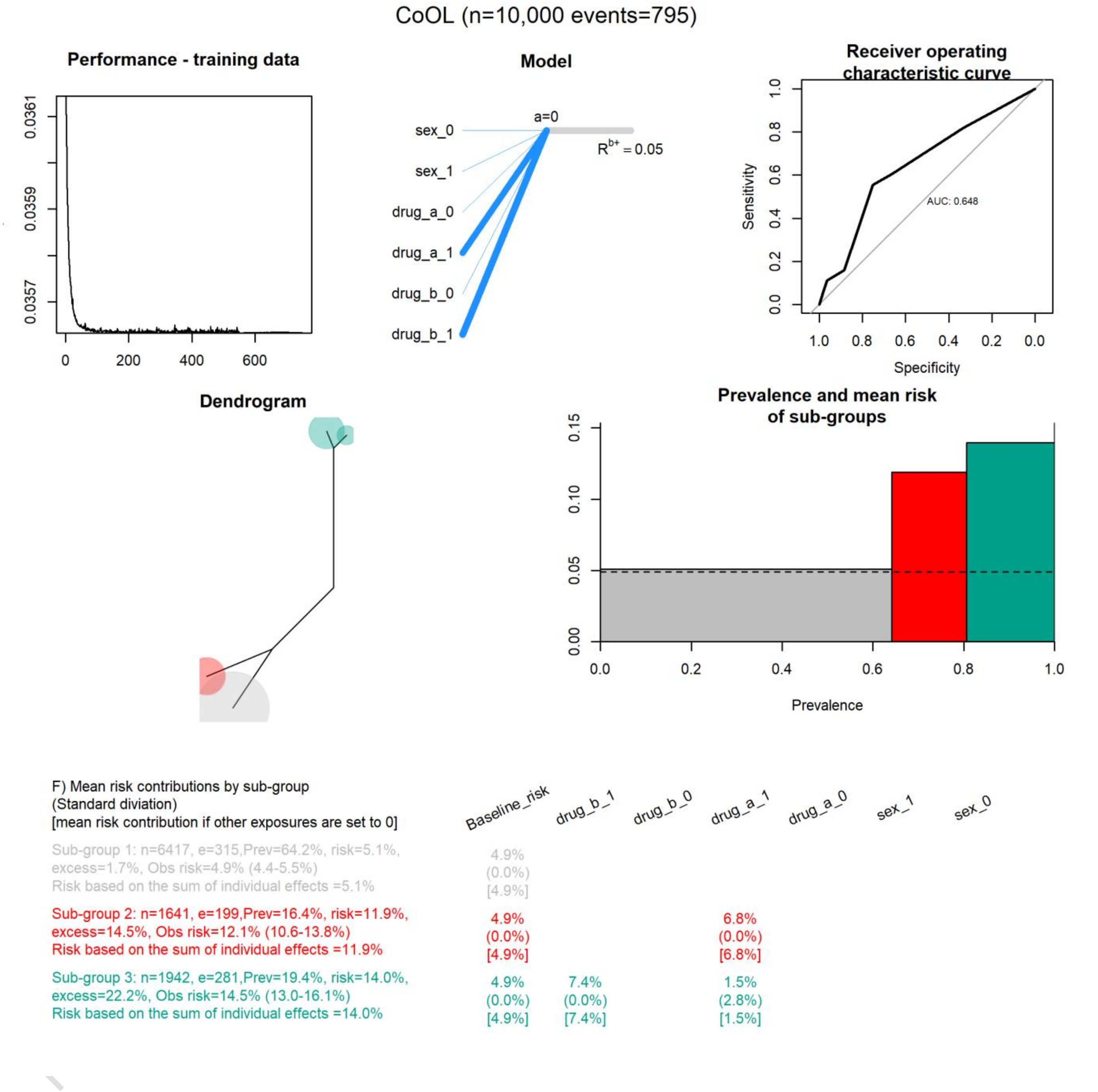

#### Supplementary simulation 8. Robustness check – data size

We ran the motivating example-simulation with different sample sizes ∈[100, 250, 500, 750, 1000, ..., 5000, ..., 40000] (Supplementary results 8). We see that the CoOL approach does not identify the sub-groups when the sample size was 100 individuals and only 11 events but with 1000 individuals and 89 events a pattern emerge. With 2000 individuals (162 events) and more individual the results are stable. The baseline risk decreases towards zero with sparse data, and stabilizes when enough data is available.

**Results based on 100 individuals (11 events)**

| F) Mean risk contributions by sub-group<br>(Standard diviation)<br>[mean risk contribution if other exposures are set to 0] | Baseline_risk | drug_b_1 | drug_b_0 | drug_a_1 | drug_a_0 | sex_1 | sex_0 |
|---|---|---|---|---|---|---|---|
| Sub-group 1: n=83, e=4,Prev=83.0%, risk=5.7%,<br>excess=42.7%, Obs risk=4.8% ( 1.6-12.5%)<br>Risk based on the sum of individual effects =1.5% | | 1.1%<br>(3.8%)<br>[0.9%] | 1.9%<br>(2.1%)<br>[0.0%] | | | | 2.2%<br>(2.1%)<br>[0.0%] |
| Sub-group 2: n=11, e=5,Prev=11.0%, risk=42.6%,<br>excess=42.2%, Obs risk=45.5% (18.1-75.4%)<br>Risk based on the sum of individual effects =13.3% | | 27.5%<br>(0.7%)<br>[13.0%] | | | | 14.4%<br>(0.0%)<br>[0.0%] | |
| Sub-group 3: n=6, e=2,Prev=6.0%, risk=28.0%,<br>excess=15.1%, Obs risk=33.3% ( 6.0-75.9%)<br>Risk based on the sum of individual effects =4.2% | | | 7.1%<br>(0.0%)<br>[0.0%] | 10.2%<br>(0.0%)<br>[4.2%] | | | 10.7%<br>(0.0%)<br>[0.0%] |

**Results based on 1000 individuals (89 events)**

| F) Mean risk contributions by sub-group<br>(Standard diviation)<br>[mean risk contribution if other exposures are set to 0] | Baseline_risk | drug_b_1 | drug_b_0 | drug_a_1 | drug_a_0 | sex_1 | sex_0 |
|---|---|---|---|---|---|---|---|
| Sub-group 1: n=836, e=43,Prev=83.6%, risk=5.5%,<br>excess=10.2%, Obs risk=5.1% (3.8-6.9%)<br>Risk based on the sum of individual effects =5.1% | 4.5%<br>(0.0%)<br>[4.5%] | | | | | | |
| Sub-group 2: n=78, e=23,Prev=7.8%, risk=27.1%,<br>excess=19.9%, Obs risk=29.5% (20.0-41.0%)<br>Risk based on the sum of individual effects =8.9% | 4.5%<br>(0.0%)<br>[4.5%] | 12.8%<br>(0.0%)<br>[3.7%] | | | | 9.1%<br>(0.0%)<br>[0.0%] | |
| Sub-group 3: n=86, e=23,Prev=8.6%, risk=24.5%,<br>excess=19.5%, Obs risk=26.7% (18.0-37.6%)<br>Risk based on the sum of individual effects =8.8% | 4.5%<br>(0.0%)<br>[4.5%] | 1.1%<br>(1.7%)<br>[1.1%] | | 10.9%<br>(0.1%)<br>[3.3%] | | | 7.7%<br>(0.1%)<br>[0.0%] |

**Results based on 2000 individuals (162 events)**

| F) Mean risk contributions by sub-group<br>(Standard diviation)<br>[mean risk contribution if other exposures are set to 0] | Baseline_risk | drug_b_1 | drug_b_0 | drug_a_1 | drug_a_0 | sex_1 | sex_0 |
|---|---|---|---|---|---|---|---|
| Sub-group 1: n=1609, e=82,Prev=80.4%, risk=5.6%,<br>excess=7.9%, Obs risk=5.1% (4.1-6.3%)<br>Risk based on the sum of individual effects =4.9% | 4.8%<br>(0.0%)<br>[4.8%] | | | | | | |
| Sub-group 2: n=181, e=35,Prev=9.0%, risk=17.2%,<br>excess=14.0%, Obs risk=19.3% (14.0-26.0%)<br>Risk based on the sum of individual effects =5.9% | 4.8%<br>(0.0%)<br>[4.8%] | 6.3%<br>(0.1%)<br>[1.1%] | | | | 5.3%<br>(0.1%)<br>[0.0%] | |
| Sub-group 3: n=210, e=45,Prev=10.5%, risk=19.6%,<br>excess=19.3%, Obs risk=21.4% (16.2-27.7%)<br>Risk based on the sum of individual effects =5.0% | 4.8%<br>(0.0%)<br>[4.8%] | | | 7.3%<br>(0.0%)<br>[0.0%] | | | 7.2%<br>(0.0%)<br>[0.0%] |

#### Supplementary simulation 9. Robustness check - regularization

We ran the motivating example-simulation with increasing regularizations of the input parameters ∈[$10^{-10}$, $10^{-5}$, $10^{-4}$, $10^{-3}$, $10^{-2}$] (Supplementary results 9). We see that with too much regularization such as $10^{-2}$, we cannot identify the sub-groups, as shown below.

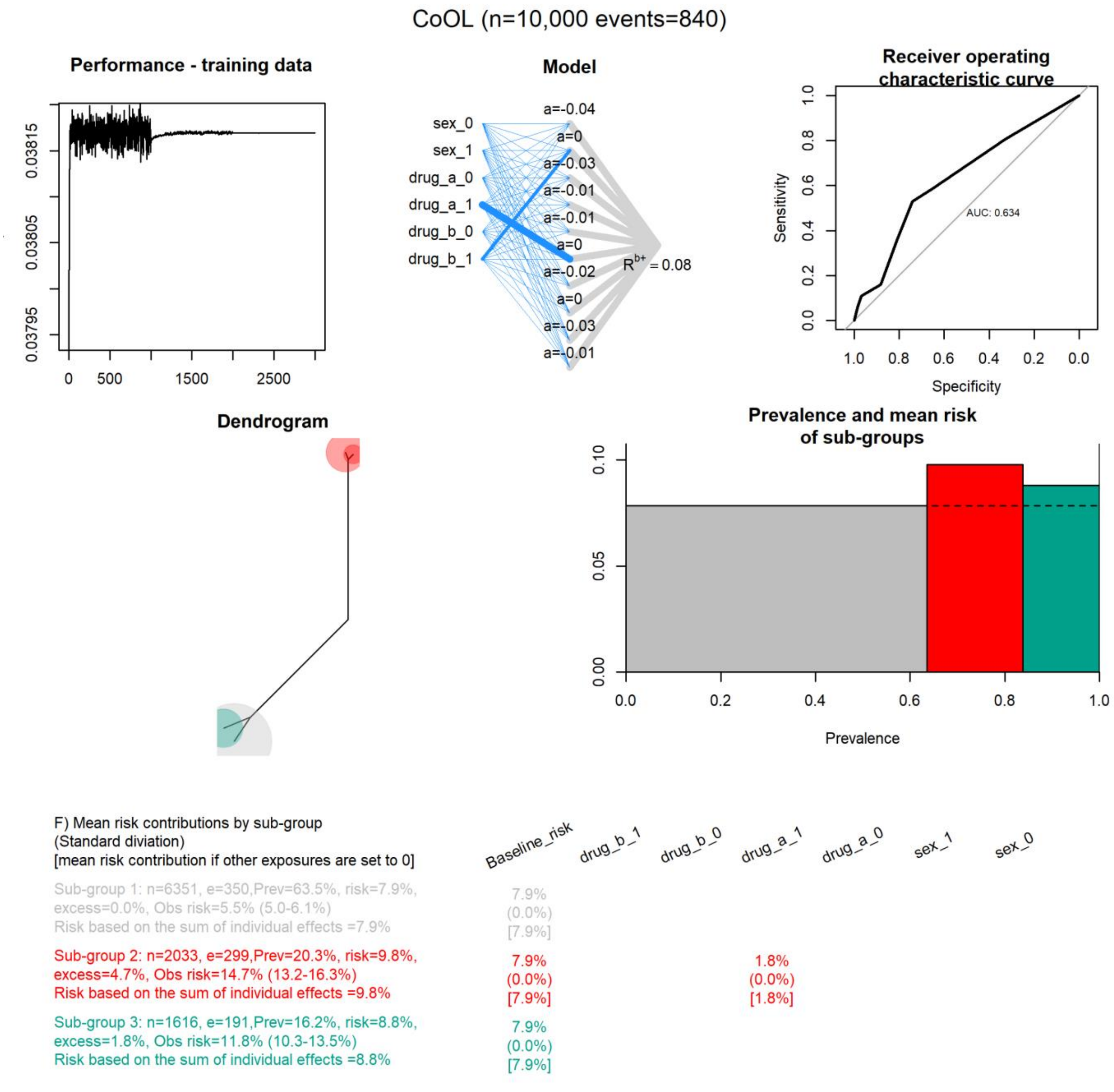

#### Supplementary real life data analysis. Example of the CoOL approach on a real life data set by the U.S. Centers for Disease Control about causes of premature mortality

We demonstrate the CoOL approach on a real life data set, which is publicly available. A full application of the causes of outcome learning approach is a study by itself – a greater endeavor than for an appendix – and thus this text primarily serves to demonstrate the computational phase of the CoOL approach on real health data rather than from simulations. Also notice that the risk of mortality in this data set is high, and that synergy as the combined effects is more than the sum of individual effects becomes violated when applied to common outcomes. We expect that the approach will be even more powerful in larger data sets with rarer outcomes.

Data is from the National Health and Nutrition Examination Survey (NHANES I) conducted by the U.S. Centers for Disease Control (CDC), as well as the NHANES I Epidemiologic Follow-up Study (NHEFS). In 1971-75, NHANES I invited 20729 American adults 25-74 years, of whom 14407 completed a medical examination (70%) A follow up study was conducted in 1992 to identify the vital status of the participants, with of whom 4785 died during the 20 years of follow up (34%) and 1116 were lost to follow up (8%).[2] Original raw tape-format data can be obtained from CDC's website[3]. Lundberg et al. (2019)[4] prepared a python script for preparing this dataset available from their Github website.[5] Using their python script, the dataset contains 14264 individuals and 4711 deaths (33%). This data has no indicator of whether an individual was lost to follow up, and we will make a very strong assumption that they were alive to the end of follow up. We restrict our analysis to adults below 50 years old at baseline, which results in 7539 individuals, of which 739 individuals die during the follow up period (9.8%), to ask the research question; "*What are the different common sets of circumstances, which might have caused young Americans to die prematurely?*" (1971 US life expectancy was 71 years[6]). We included the following information shown in Table 1.

Pre-print (not peer reviewed nor accepted by a journal)

**Table 1. Variable operationalization and prevalence.**

| Variable | Category | Total (n = 7539) | Data for training (n=3770) | Data for internal validating (n=3769) |
|---|---|---|---|---|
| Vital status | Alive | 6800 (90.2%) | 3410 (90.5%) | 3390 (89.9%) |
| | Died | 739 (9.8%) | 360 (9.5%) | 379 (10.1%) |
| Sex | Men | 2571 (34.1%) | 1296 (34.4%) | 1275 (33.8%) |
| | Women | 4968 (65.9%) | 2474 (65.6%) | 2494 (66.2%) |
| Age | 25-29 years | 1926 (25.5%) | 976 (25.9%) | 950 (25.2%) |
| | 30-39 years | 3015 (40.0%) | 1510 (40.1%) | 1505 (39.9%) |
| | 40-49 years | 2598 (34.5%) | 1284 (34.1%) | 1314 (34.9%) |
| BMI | Low (< 18.5) | 297 (3.9%) | 154 (4.1%) | 143 (3.8%) |
| | Normal (18.5 to 25) | 3969 (52.6%) | 1999 (53.0%) | 1970 (52.3%) |
| | High (> 25) | 3273 (43.4%) | 1617 (42.9%) | 1656 (43.9%) |
| Systolic blood pressure | Normal (<= 120 mm Hg) | 3908 (51.8%) | 1956 (51.9%) | 1952 (51.8%) |
| | High (> 120 mm Hg) | 3631 (48.2%) | 1814 (48.1%) | 1817 (48.2%) |

[2] Cox, Christine S. Plan and operation of the NHANES I Epidemiologic Followup Study, 1987. No. 27. US Department of Health and Human Services, Public Health Service, Centers for Disease Control, National Center for Health Statistics, 1992.

[3] HTTPS://WWWN.CDC.GOV/NCHS/NHANES/

[4] Lundberg, Scott M., et al. "Explainable AI for trees: From local explanations to global understanding." *arXiv preprint arXiv:1905.04610* (2019).

[5] HTTPS://GITHUB.COM/SUINLEELAB/TREEEXPLAINER-STUDY/TREE/MASTER/NOTEBOOKS/MORTALITY

[6] HTTPS://WWW.GOOGLE.COM/PUBLICDATA/EXPLORE?DS=D5BNCPPJOF8F9_&MET_Y=SP_DYN_LE00_IN&IDIM=COUNTRY:USA:GBR:CAN&HL=DA&DL=DA

From the marginal associations from Table 2, we see – based on all data (n=7539) - that men compared with women are associated with a higher risk of dying; age 40-49 compared to both 25-29 and 30-39 are associated with a higher risk of dying; high and low BMI compared with normal BMI is associated with a higher risk of dying; and high compared with normal systolic blood pressure is associated with a higher risk of dying.

**Table 2. Risk differences from a mutual adjusted model on all data (n=7539)**

| Variable | Risk difference (95% confidence interval) | | |
|---|---|---|---|
| | Unadjusted | Causally adjusted* | Mutually adjusted** (direct effects and potential m-bias) |
| Women (compared with men) | -0.05 (-0.06 to -0.03) | -0.05 (-0.06 to -0.03) | -0.04 (-0.05 to -0.02) |
| Age 30-39 (compared with age 25-29 years) | 0.01 (-0.01 to 0.02) | 0.01 (-0.01 to 0.02) | 0.01 (-0.01 to 0.02) |
| Age 40-49 (compared with age 25-29 years) | 0.09 (0.07 to 0.1) | 0.09 (0.07 to 0.1) | 0.08 (0.06 to 0.1) |
| Normal BMI (compared with low BMI) | -0.05 (-0.08 to -0.01) | -0.05 (-0.09 to -0.02) | -0.06 (-0.09 to -0.02) |
| High BMI (compared with low BMI) | -0.02 (-0.06 to 0.01) | -0.04 (-0.07 to 0) | -0.06 (-0.09 to -0.02) |
| High systolic blood pressure (compared with normal) | 0.05 (0.04 to 0.07) | 0.04 (0.03 to 0.05) | 0.03 (0.02 to 0.05) |

* Adjusted for common measured causes according to the DAG

** Adjusted for all other co-variables

##### Pre-computational phase

Body mass index (BMI) has a U shaped association with mortality, and it has been speculated that individuals suffering underlying illness may have a higher propensity of being both under- and overweight.[7] An IV study suggest that the effect of low BMI on mortality may be overestimated due to reverse causation.[8] High blood pressure is also an established risk factor of mortality,[9] and interventions lowering blood pressure shows preventive effects.[10] Higher BMI is also associated with higher blood pressure.[11] Both BMI and systolic blood pressure may be affected by underlying illness and health related behavior, which are not measured in the data we have available, and needs to be considered when interpreting the results. Our assumed potential causal structure is sketched in Figure 1.

**Figure 1. Potential causal structure**

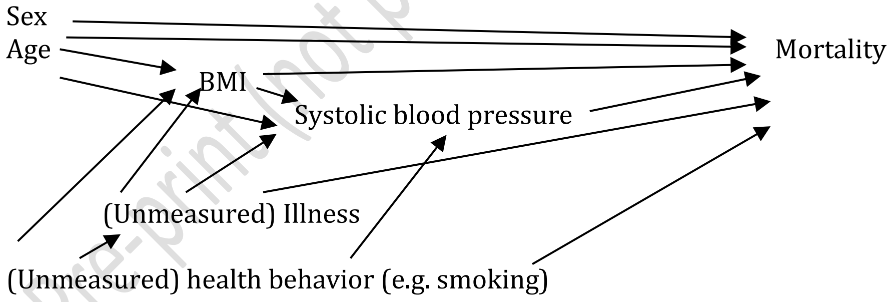

Pre-print (not peer reviewed nor accepted by a journal)

#### Computational phase

We randomly split data into 50% to a training data set (n=3770) and 50% to an internal validation data set for manually confirming the findings from the training set (n=3769) to ensure the robustness of the findings. Based on the prevalence in Table 1, the two split datasets looks balanced.

When we analyse the training data using the Causes of Outcome Learning approach, we see that there are six clusters of different sizes (Figure 2). The red sub-group is the majority of 51% and have the baseline risk of approximately 6.2%. High systolic blood pressure is relevant in all the remaining identified sub-groups. The orange group of 18% of the population have a risk of 10% due to being in their 30s and have high systolic blood pressure, and the grey group of 12% of the population has a risk of 13% due to being in their 40s and have high systolic blood pressure. The remaining three sub-groups are men in their 40s with high systolic blood pressure and different BMI values. Among these, those with a high BMI (the yellow group) are 6% of the population with a risk of 21%, those with a normal BMI (the blue group) are 3% of the population with a risk of 29%, and those with a low BMI (the green group) are 0.5% of the population with a risk of 37%. The potential largest public health impact is among the yellow group with excess deaths equal to 9% of all deaths, the grey group with excess deaths equal to 8% of all deaths, the orange group with excess deaths equal to 7% of all deaths, the blue group with excess deaths equal to 7% of all deaths, the green group with excess deaths equal to 1% of all deaths. **Finding 1 on public health impact:** From a public health point of view, we need to understand whether systolic blood pressure by itself cause deaths or it is a proxy of other causes. When understood, population wide approaches seem to be an approach which may affect many of the sub-groups. **Finding 2 on etiology:** Our findings suggest that among men in their 40s with high systolic blood pressure, a dose-response relationship exists with the lower BMI is, the higher mortality is. To test whether these findings are robust and not chance findings (type 1 errors / false positive), we manually assess these in our yet unseen internal validation dataset.

eer reviewed nor accepted by a journal)

**Figure 2. Results from the computational phase of the CoOL approach (continues)**

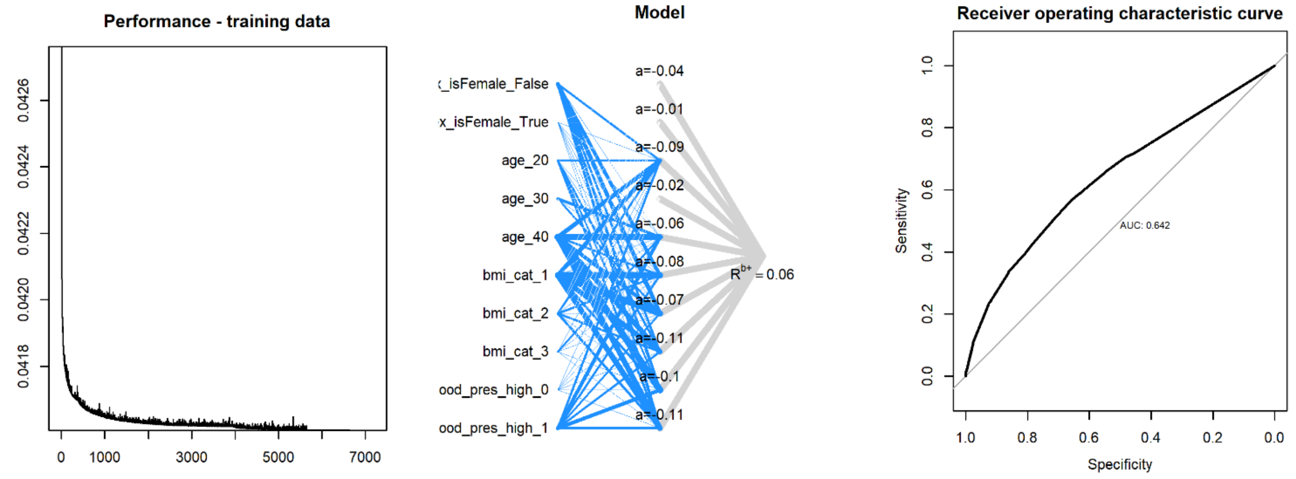

**Figure 2. Results from the computational phase of the CoOL approach (continued)**

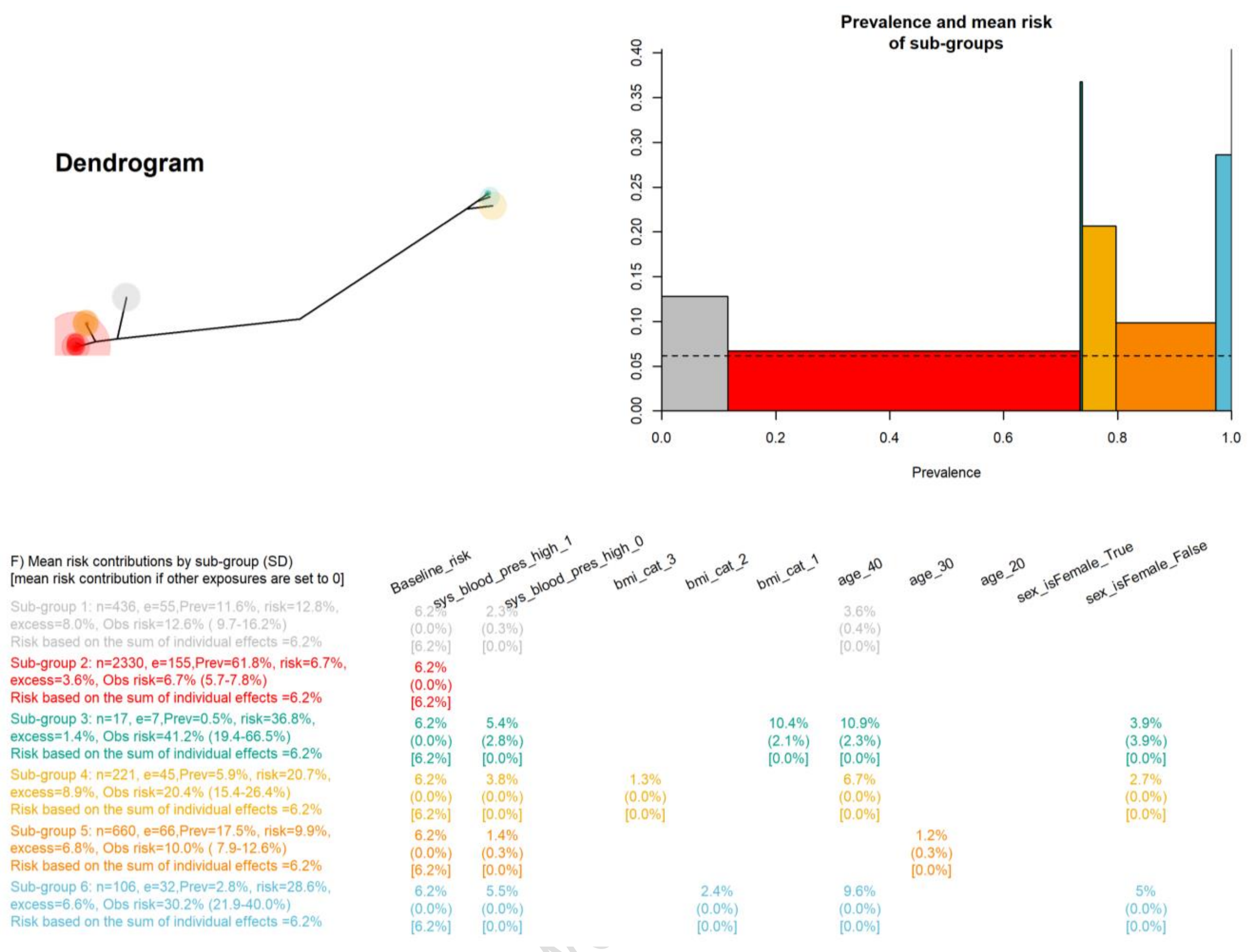

We first check in the internal validation data that high systolic blood pressure is associated with a higher risk of mortality among all individuals in their 30s and 40s compared with low systolic blood pressure and individuals in their 20s (Table 3). This is the case and supports that we need an understanding of whether systolic blood pressure is by itself cause death or whether it is a proxy of other distal causes, which may guide population wide interventions.

**Table 3. Validation data set to manually check the robustness of finding 1**

| Risk of death within 20 years. All of the validation dataset (n=3769) | 25-29 (95% CI) | 30-39 (95% CI) | 40-49 (95% CI) |
|---|---|---|---|
| Normal systolic blood pressure | 7% (5-9%) | 6% (4-8%) | 12% (9-14%) |
| High systolic blood pressure | 6 (3-10%) | 8% (5-10%) | 20% (17-22%) |

We also observed that low BMI was associated with higher mortality among men in their 40s with a high systolic blood pressure. We now check whether these findings can be found in our internal validation data set (Table 4).

Preprint (not peer reviewed)

**Table 4. Internal validation data set to manually check the robustness of finding 2**

| Risk of death within 20 years. Men in their 40s with high systolic blood pressure (n=1275) | High BMI (95% CI) | Normal BMI (95% CI) | Low BMI (95% CI) |
|---|---|---|---|
| | 23% (16-27%) | 28% (20-35%) | 67% (32-101%) |

We can further formally test for a linear dose-response (results from the training dataset indicated a linear relationship) in the validation data set using a linear regression which gives a parameter of 9.6% (95%CI 1-18%) increase in risk when moving one BMI class down. We can also test if the combined risk due to being a man in the 40s with high blood pressure and a low BMI is more than the sum of the individual effects of being a man vs a woman, in the 40s vs below 40, with high blood pressure vs normal blood pressure, and low BMI vs normal or high BMI among all individuals in the internal validation dataset. The additional risk difference is 43% (95% CI 19%-67%) higher, and if there were no synergy, we would have expected this value to be 0. It should be noted that this analysis relies on few cases in the strata with all combined exposures, but these finding were present in both splits of original data, which suggest that the findings are internally robust. We discuss whether the findings are causal in phase 3.

**Post-computational phase**

The study population was American individuals followed from 1970s. We have confirmed that the findings are unlikely chance findings, however, various biases may be present, which challenge a causal interpretation. We can sketch our new working hypotheses in a DAG, where the bold arrows indicate the combination of circumstances associated with a higher than normal risk of mortality in the studied cohort, and the dotted lines indicate reluctance for drawing causal conclusions (Figure 3).

**Figure 3. Updated potential causal structure**

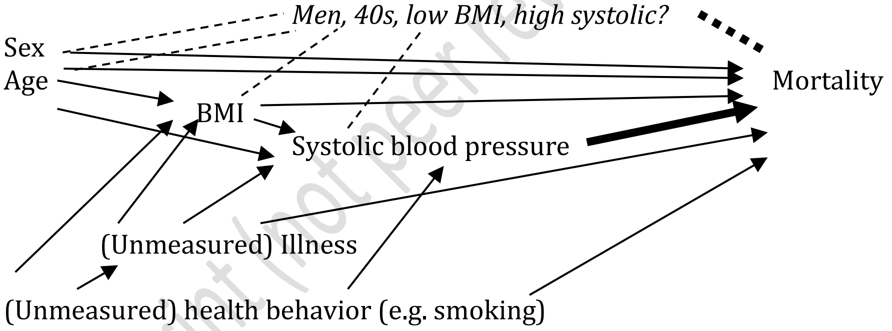

We explored the literature for above association pattern regarding men in the 40s with low BMI and high systolic blood pressure, which is previously reported, and explained as "*In individuals without cardiovascular disease, elevated body mass index (BMI) is associated with an increased risk of death. However, in patients with certain chronic diseases, including heart failure, low BMI has been associated with increased mortality.*", where 540 patients of whom 84% men with a mean age of 59 years, were studied and the authors conclude "*In patients with*

Pre-print (not peer reviewed nor accepted by a journal)

*systolic heart failure, lower BMI was an independent predictor of increased all cause death rate, cardiac death rate and HF death rate*.".[12]

As discussed by Sudharsanan et al (2020),[13] "*Estimating the relationship between BMI and mortality presents a distinct methodologic challenge because individuals tend to lose weight when sick and especially before death. This introduces a form of unobserved confounding that often results in empirical estimates implying that higher BMI is actually protective of mortality. One way to address this source of confounding is to use BMI measurements taken earlier in an individual's life*". Since we study relatively young individuals with a follow up period of 20 years, our estimates are likely less affected by acute illness affecting both BMI and mortality, however, a chronic underlying illness could confound our association with low BMI and mortality.

Our results may have identified some high risk groups – known elsewhere in the literature - but a causal interpretation of the finding is challenging. E.g. symptoms of chronic illness may materialize with low BMI and high blood pressure, and thus are simply indicators. By extracting such information from a complex model, screening indicators can be explained with words and do not rely on a black box algorithm. This example of applying the Causes of Outcome Learning approach on real life data indicate it to be a promising first step for causal discovery by first describing combination of circumstances for whom are in an elevated risk of the outcome, and secondly the iterative process of disentangling cause and effect of these phenomena. It also highlight the importance of a well-considered research question, domain expertise, and high quality data, and thus the necessity of an iterative process through all 3 phases of the causes of outcome learning approach. Again, a full application of the causes of outcome learning approach is a study by itself – a greater endeavor than for an appendix.

Pre-print (not peer reviewed nor accepted by a journal)

[12] Hua, Y. H., et al. "Body mass index and prognosis in patients with systolic heart failure." *Zhonghua xin xue guan bing za zhi* 37.10 (2009): 870-874.

[13] Sudharsanan, Nikkil, and Jessica Y. Ho. "Rural–Urban Differences in Adult Life Expectancy in Indonesia: A Parametric g-formula–based Decomposition Approach." *Epidemiology (Cambridge, Mass.)* 31.3 (2020): 393.
