## Supplementary figures and images for "Causes of Outcome Learning: A causal inference-inspired machine learning approach to disentangling common combinations of potential causes of a health outcome"

### Add_noise_0.png

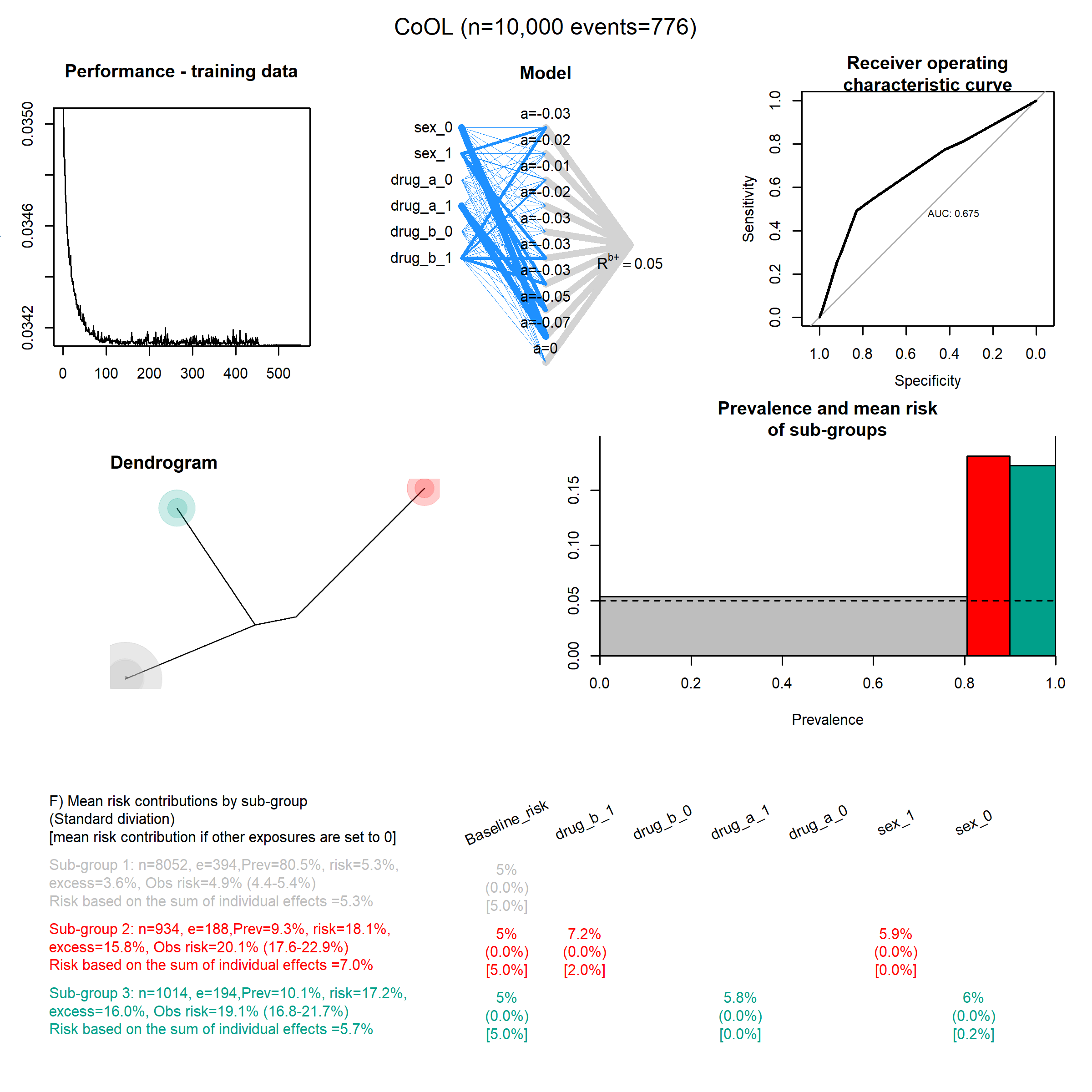

### Add_noise_5.png

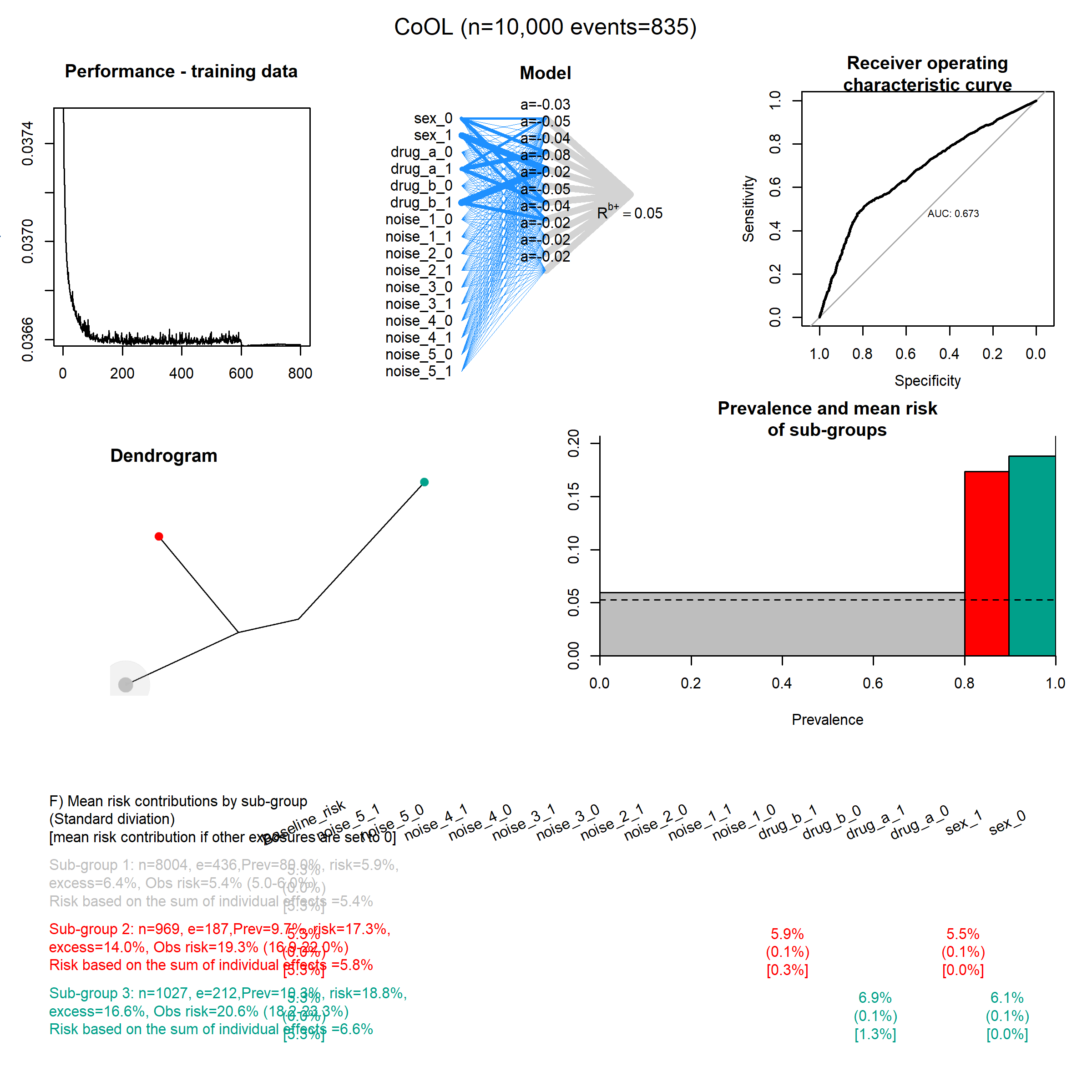

### Add_noise_10.png

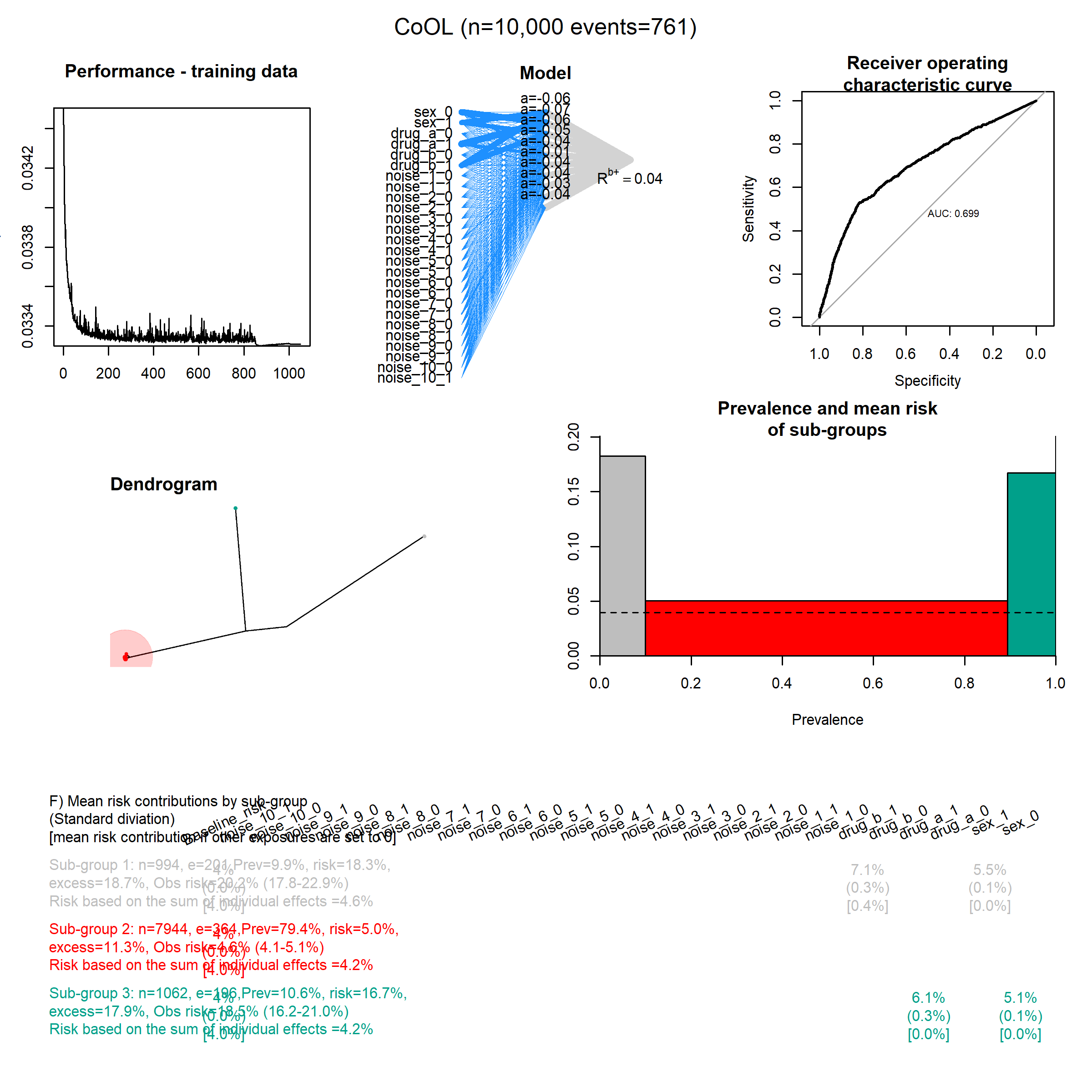

### Add_noise_15.png

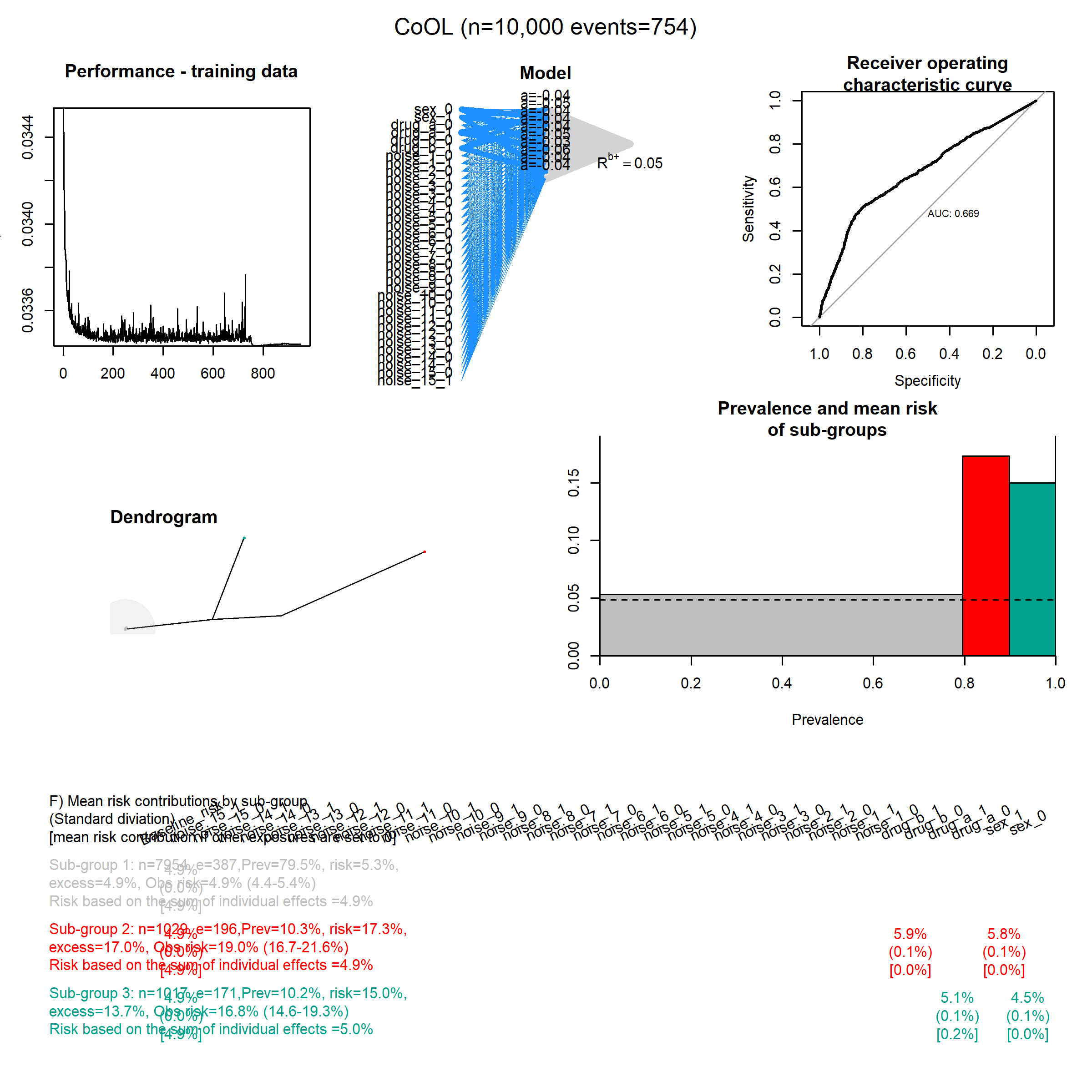

### Add_noise_20.png

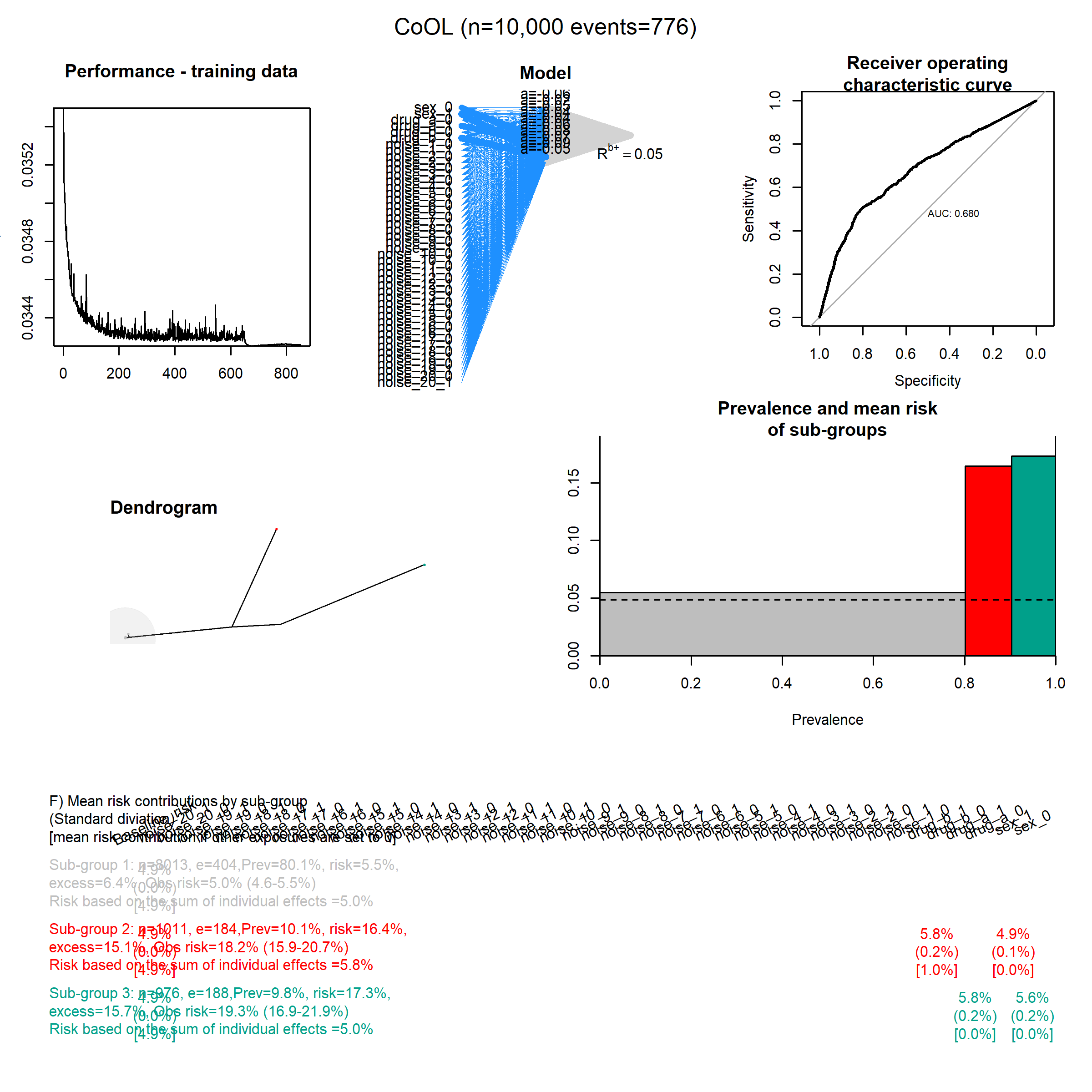

### Add_noise_25.png

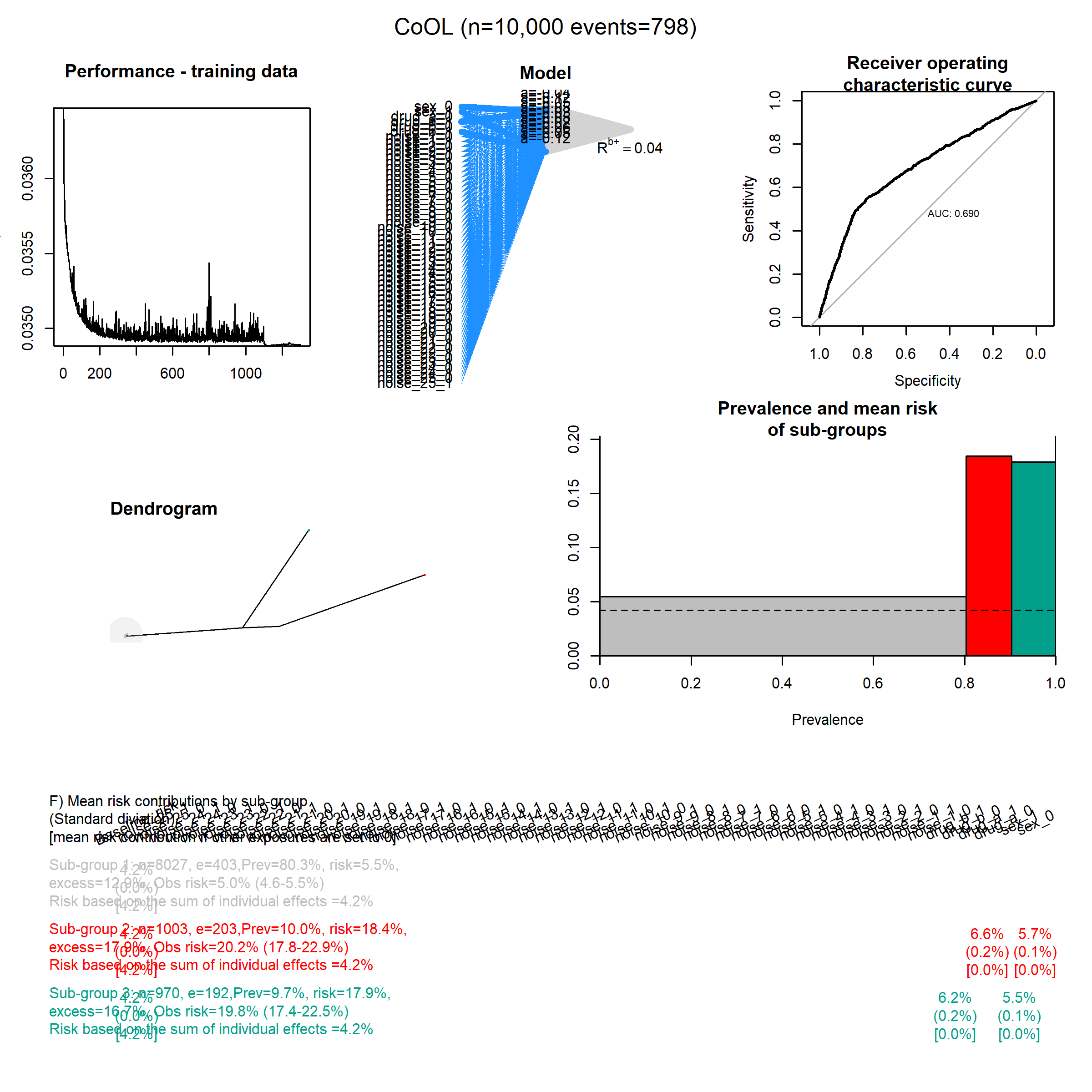

### Add_noise_30.png

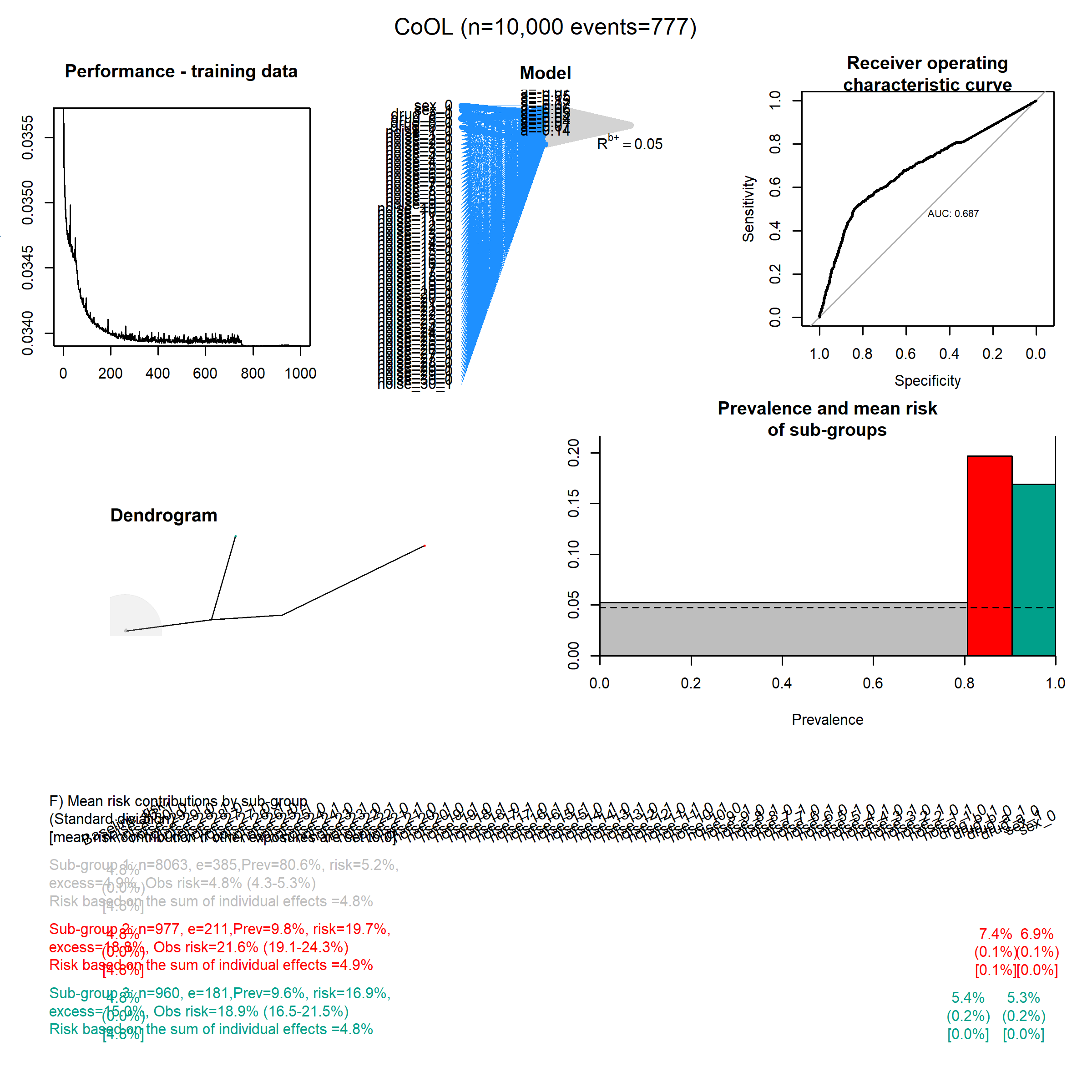

### Add_noise_35.png

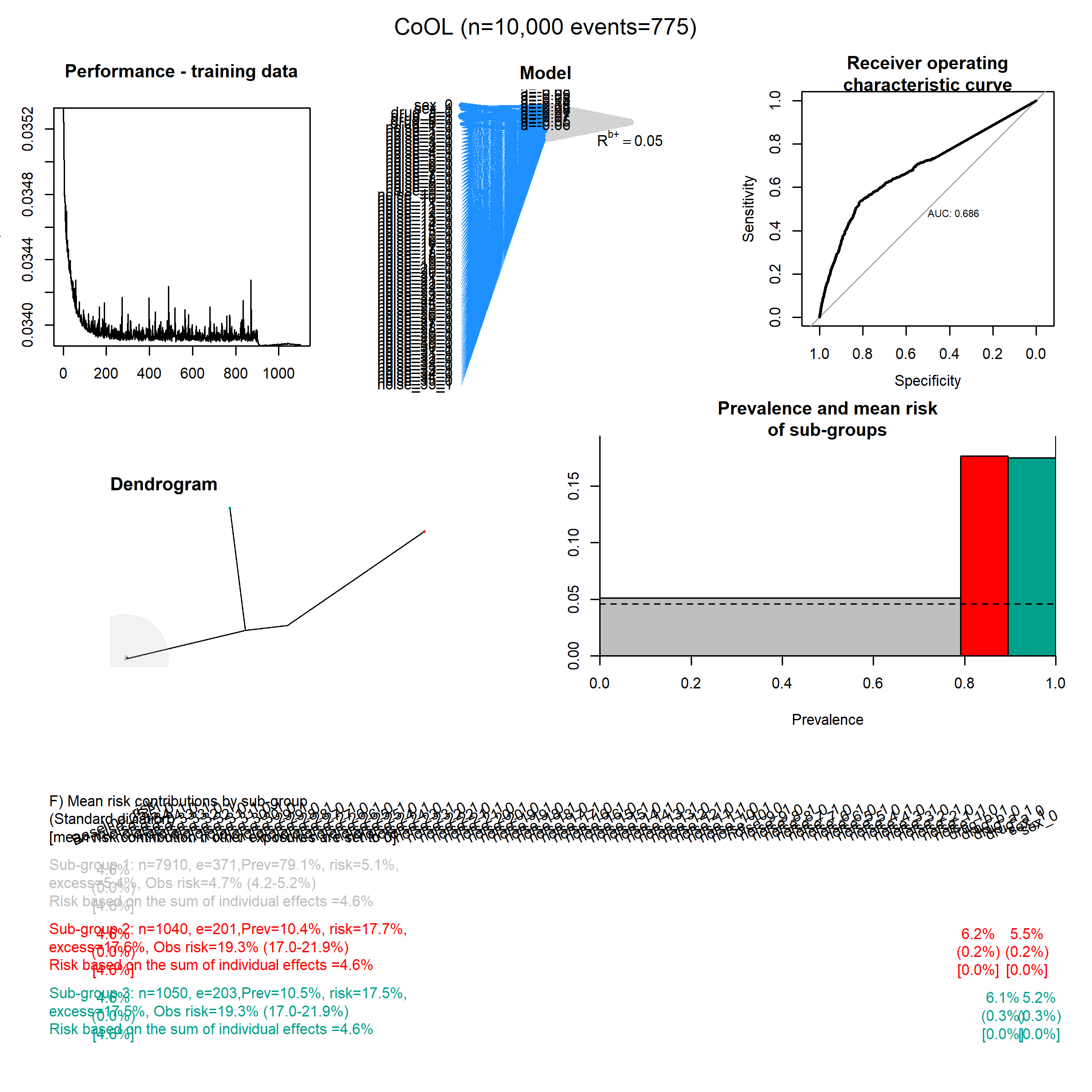

### Add_noise_40.png

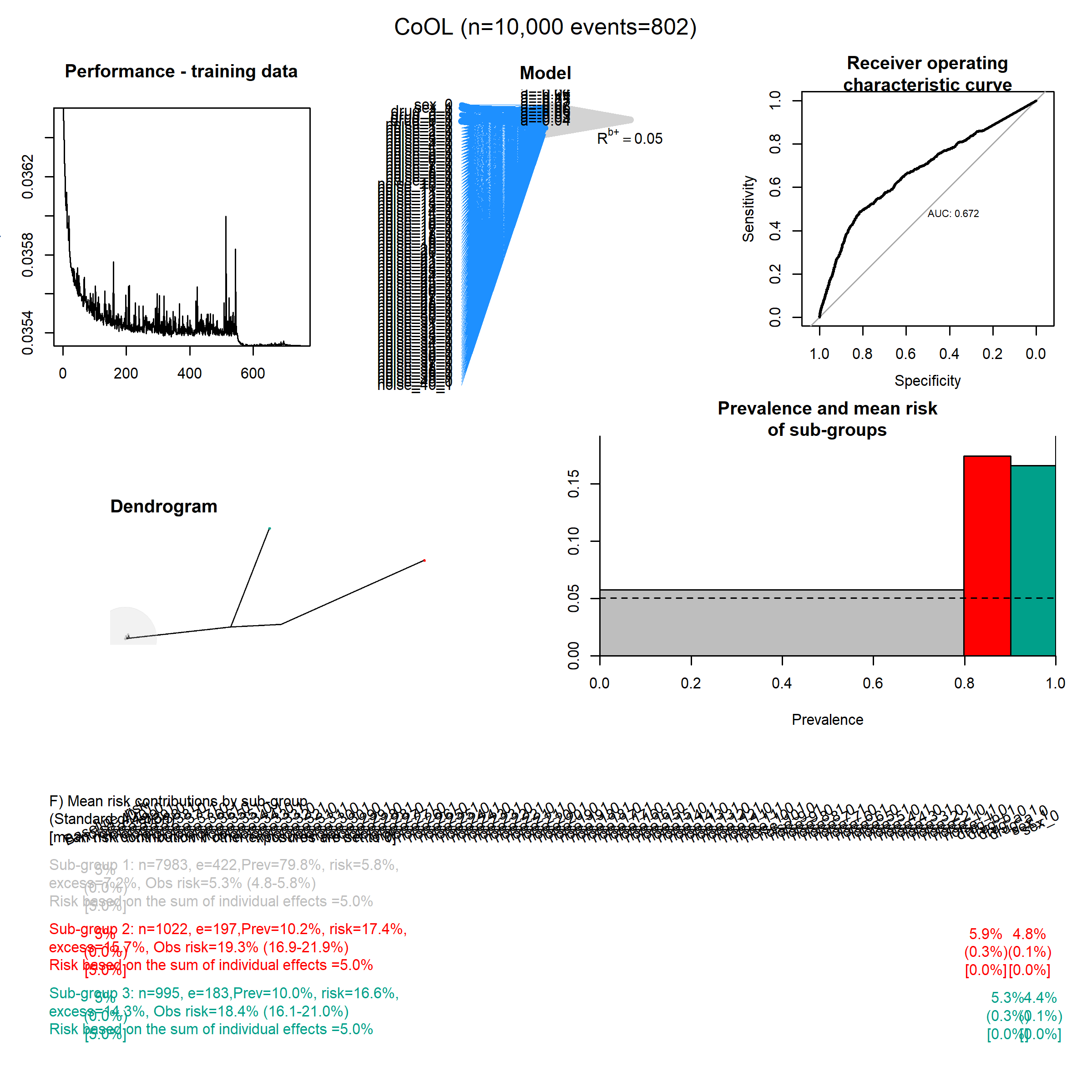

### Add_noise_45.png

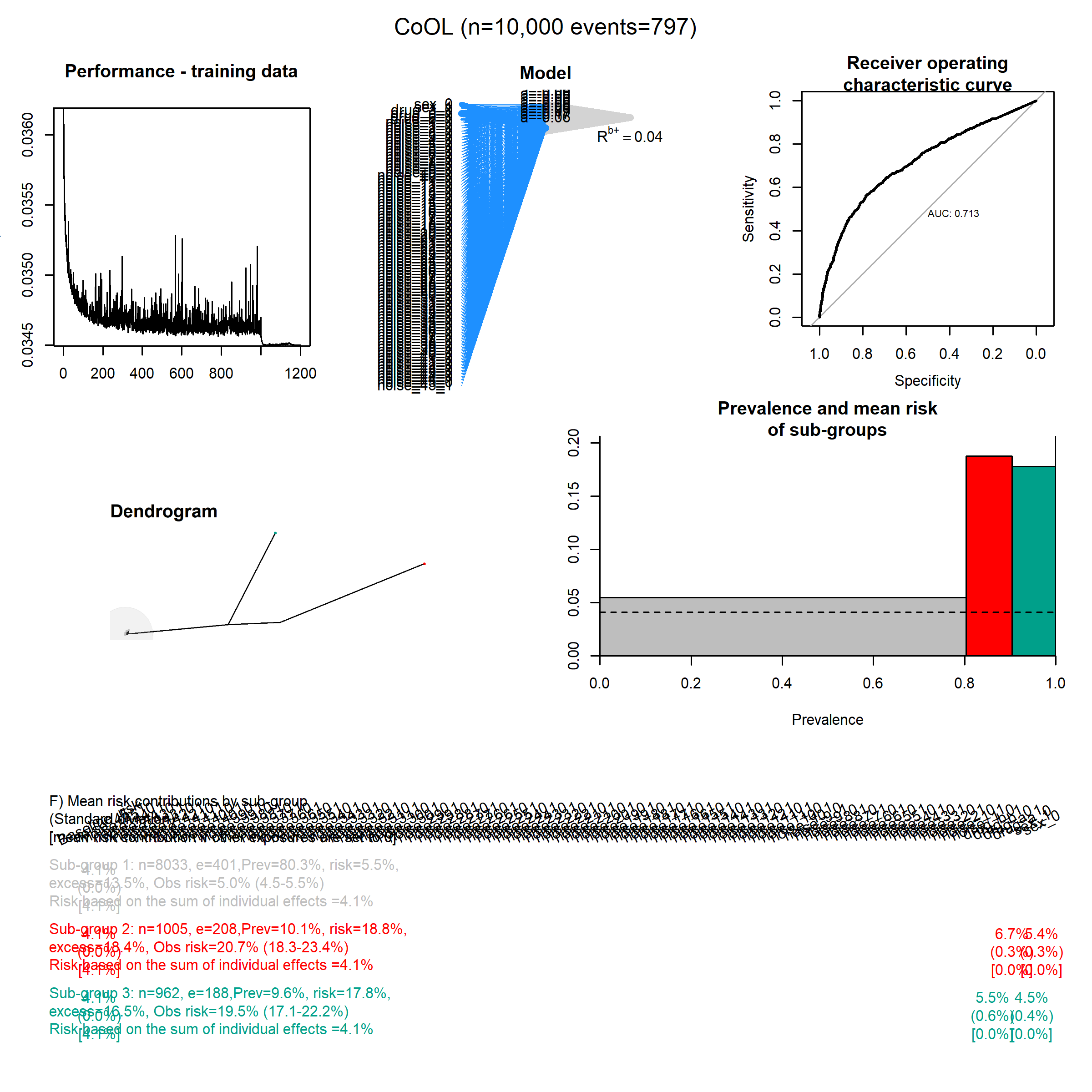

### Add_noise_50.png

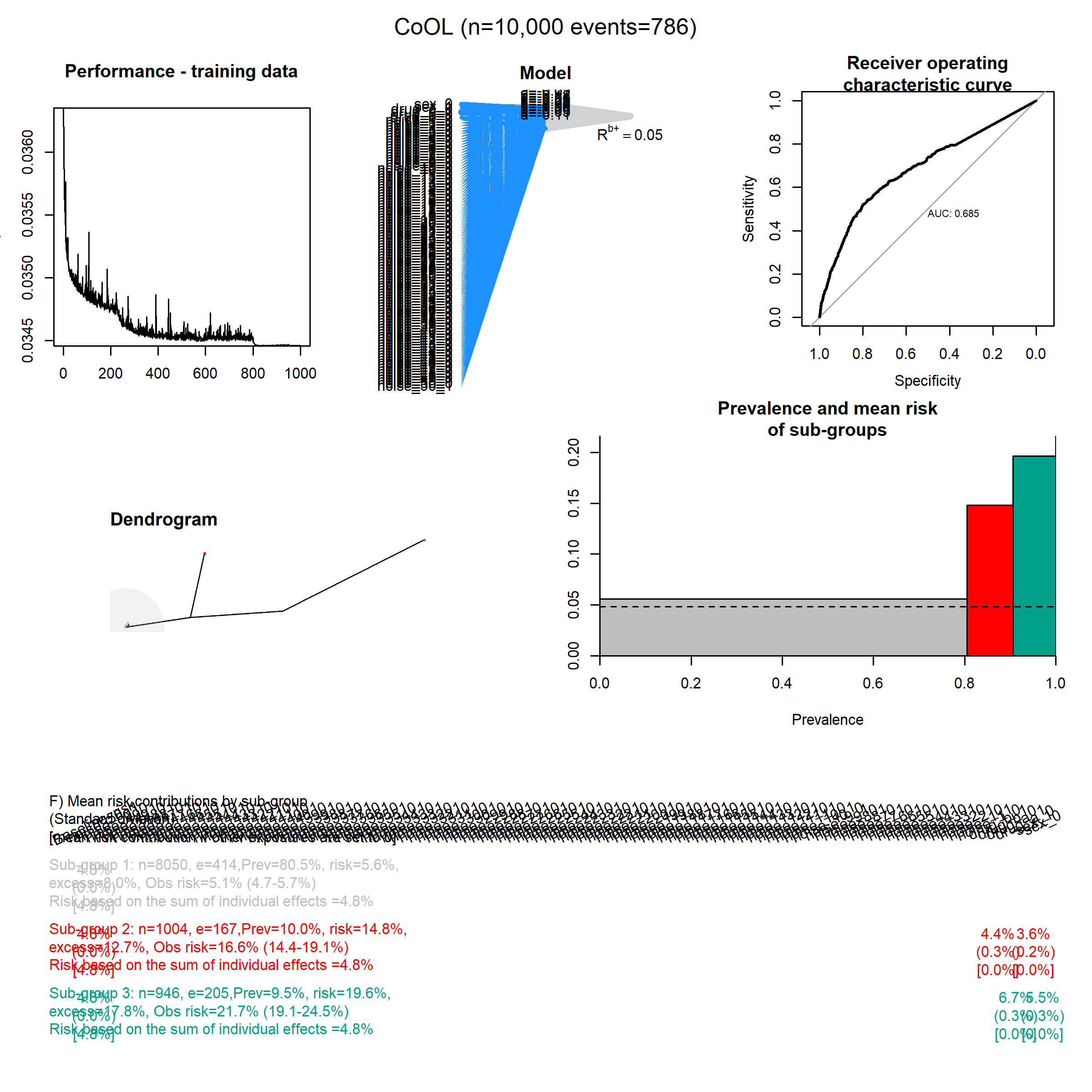

### Common_1.png

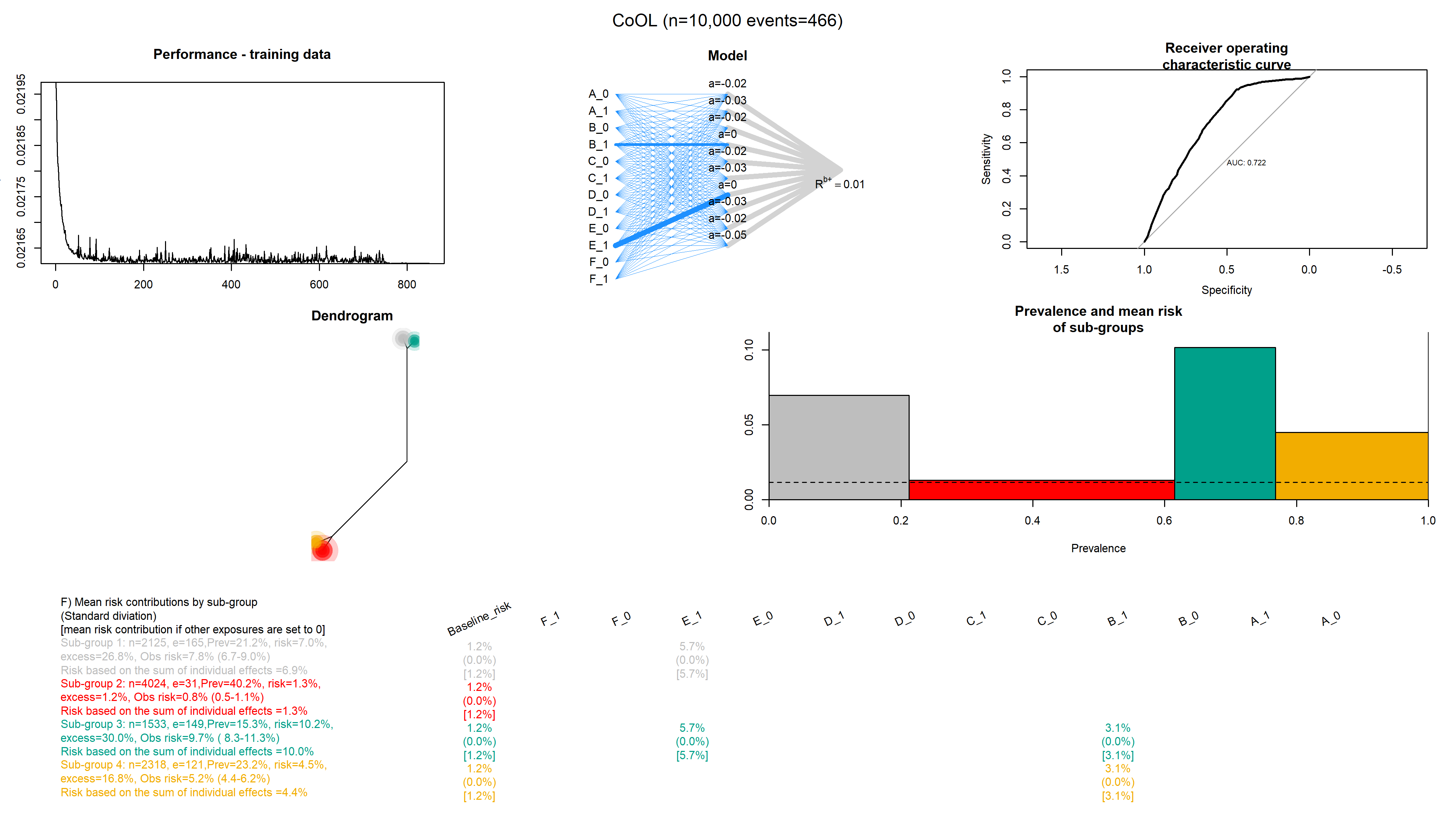

### Common_2.png

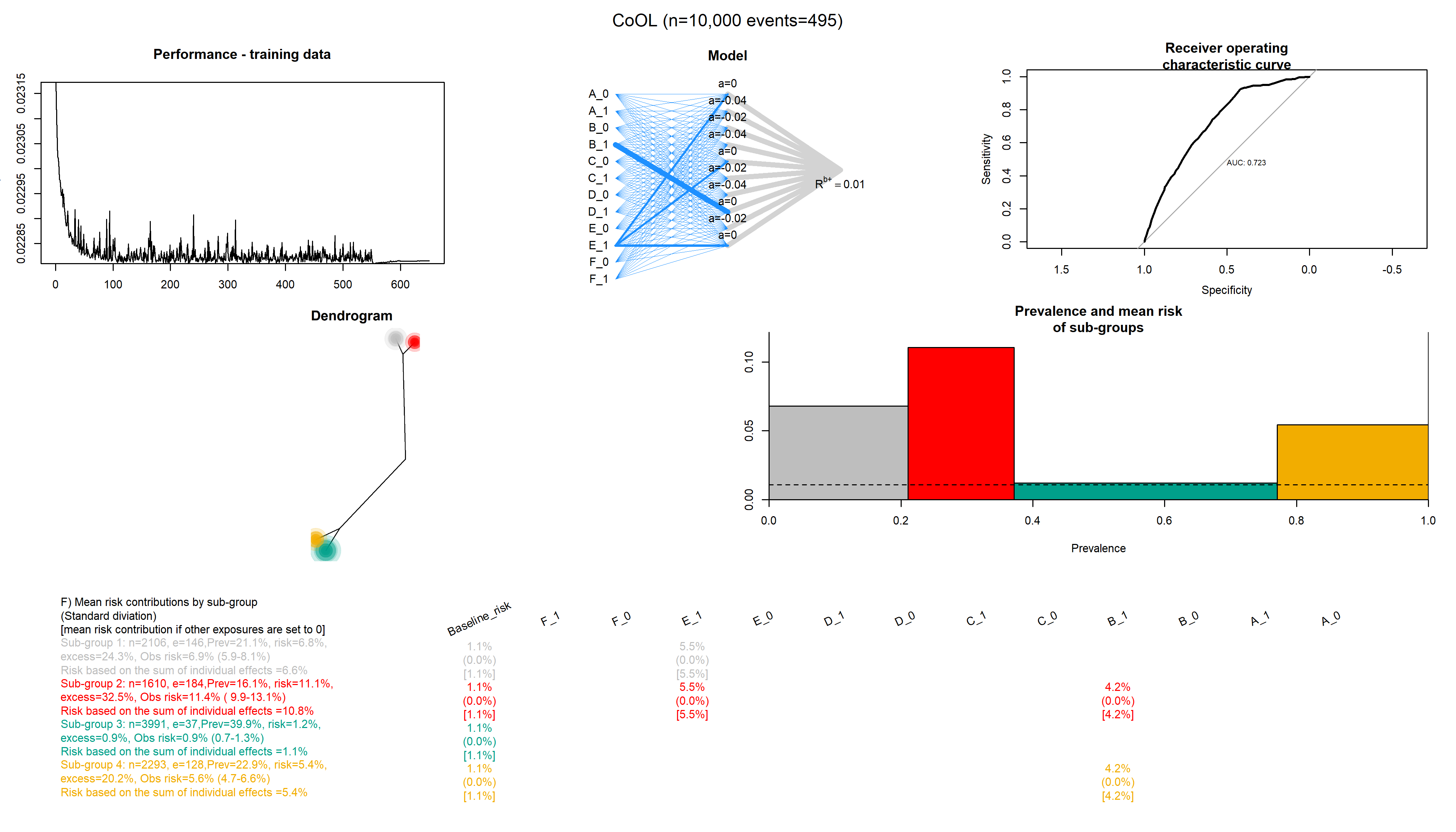

### Common_3.png

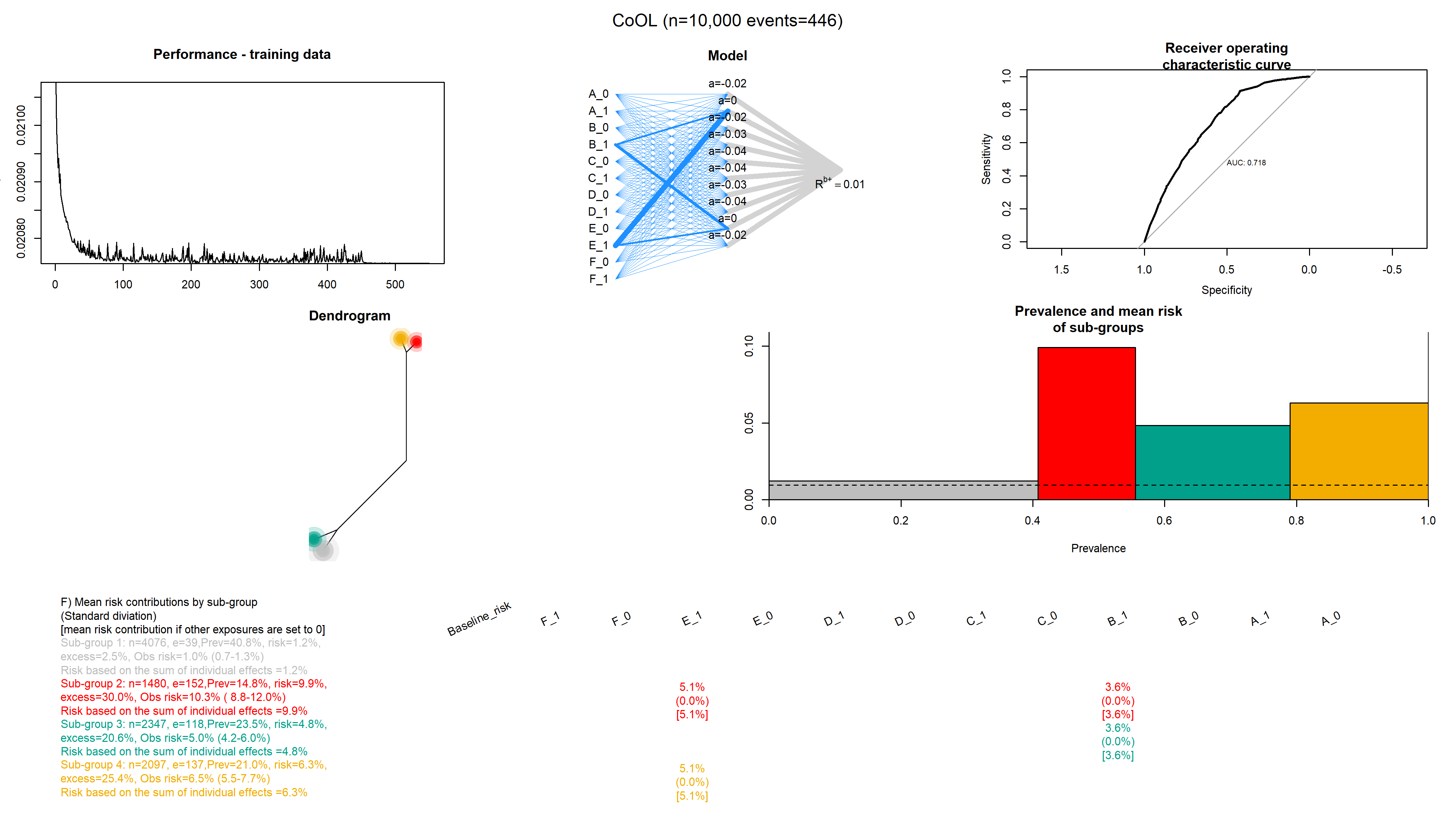

### Common_4.png

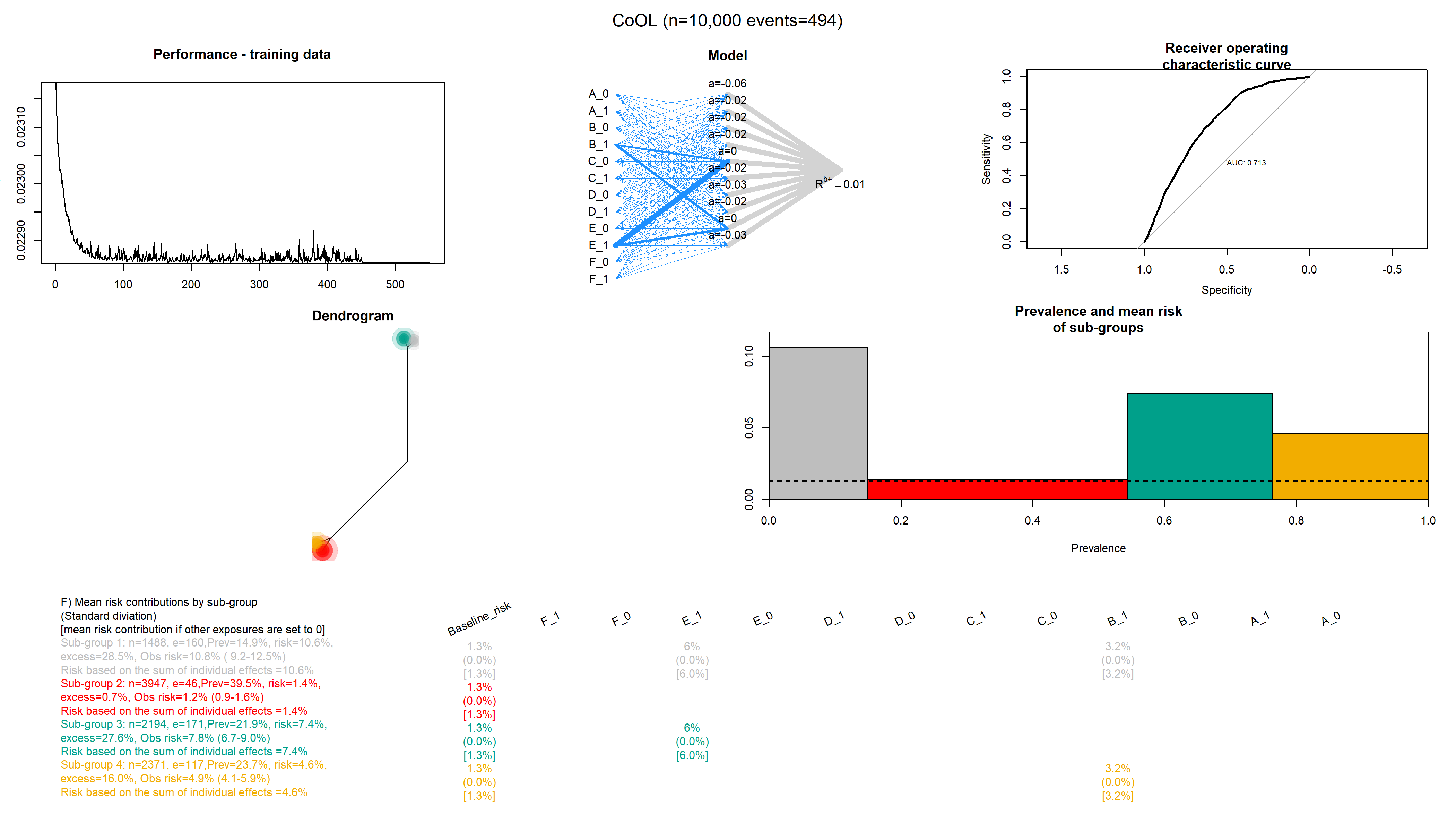

### Common_5.png

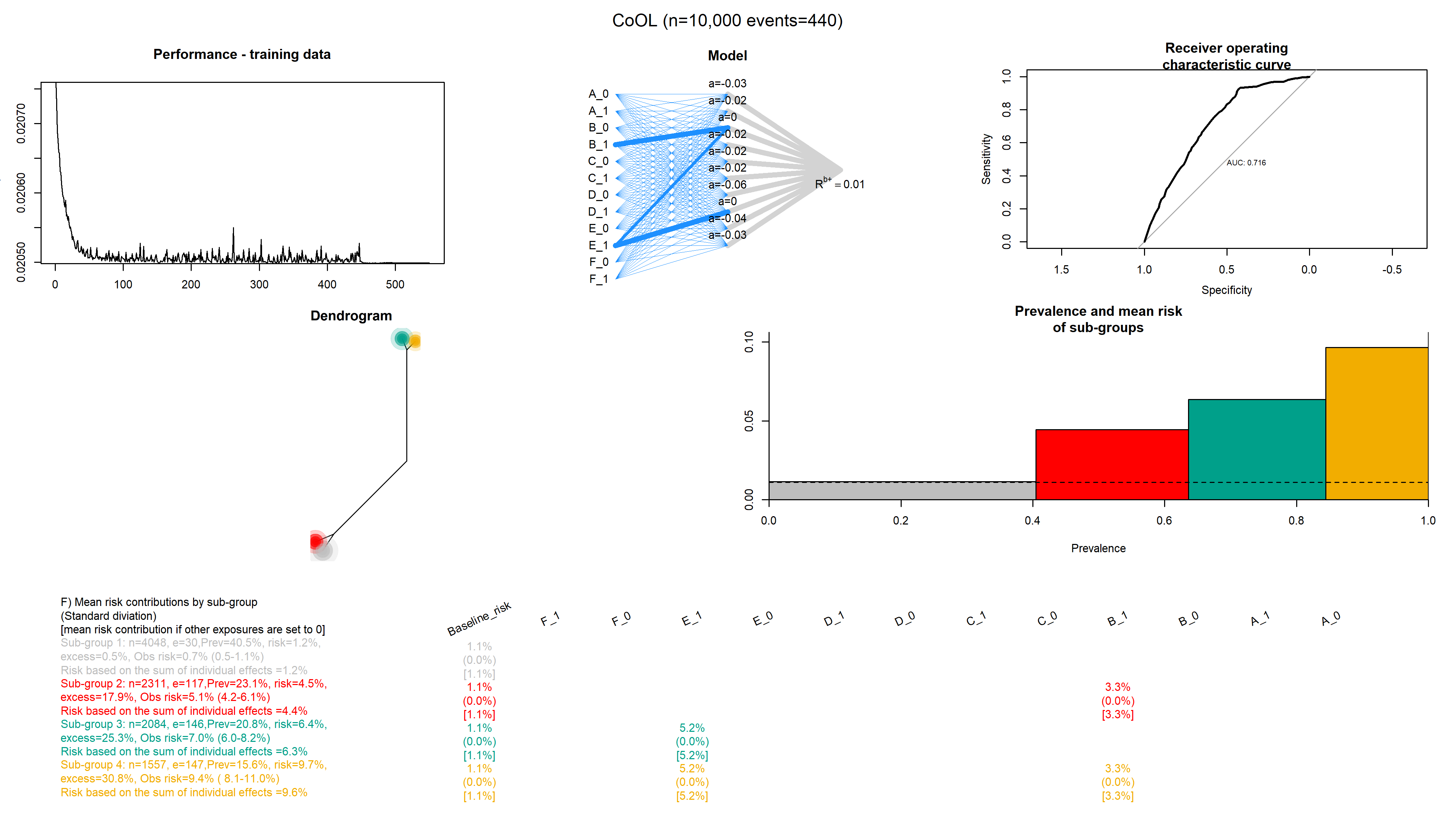

### Common_6.png

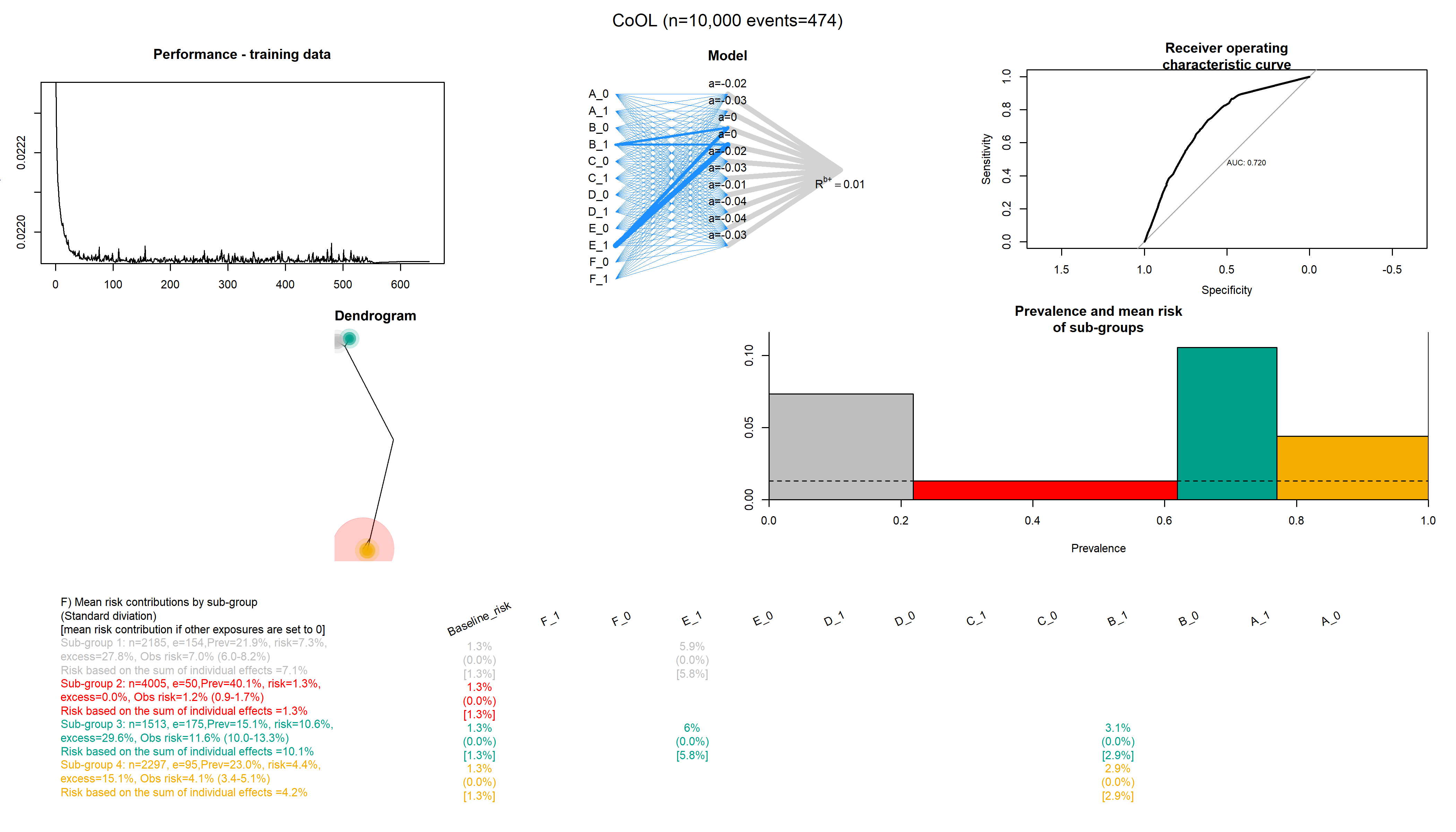

### Common_7.png

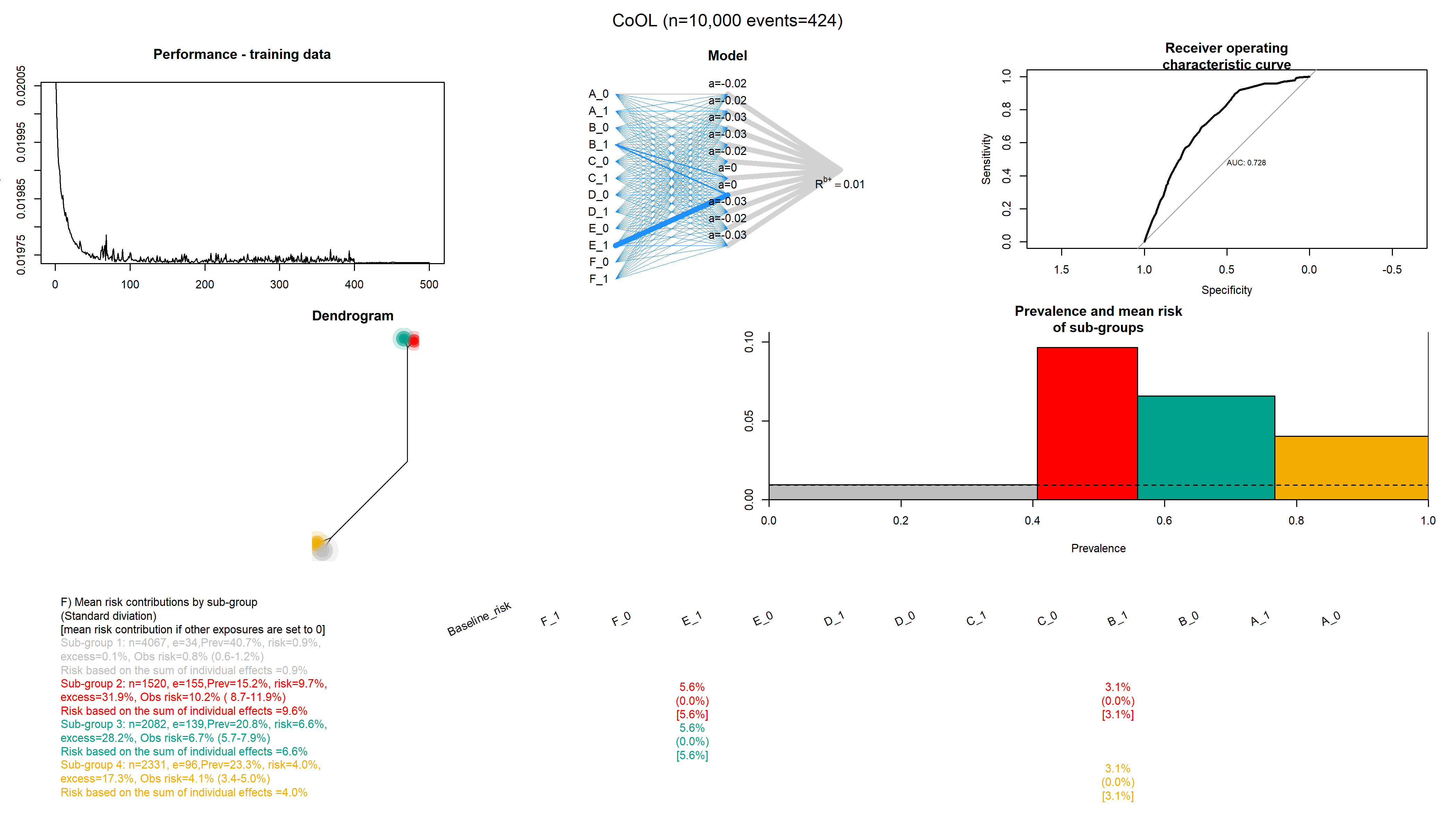

### Common_8.png

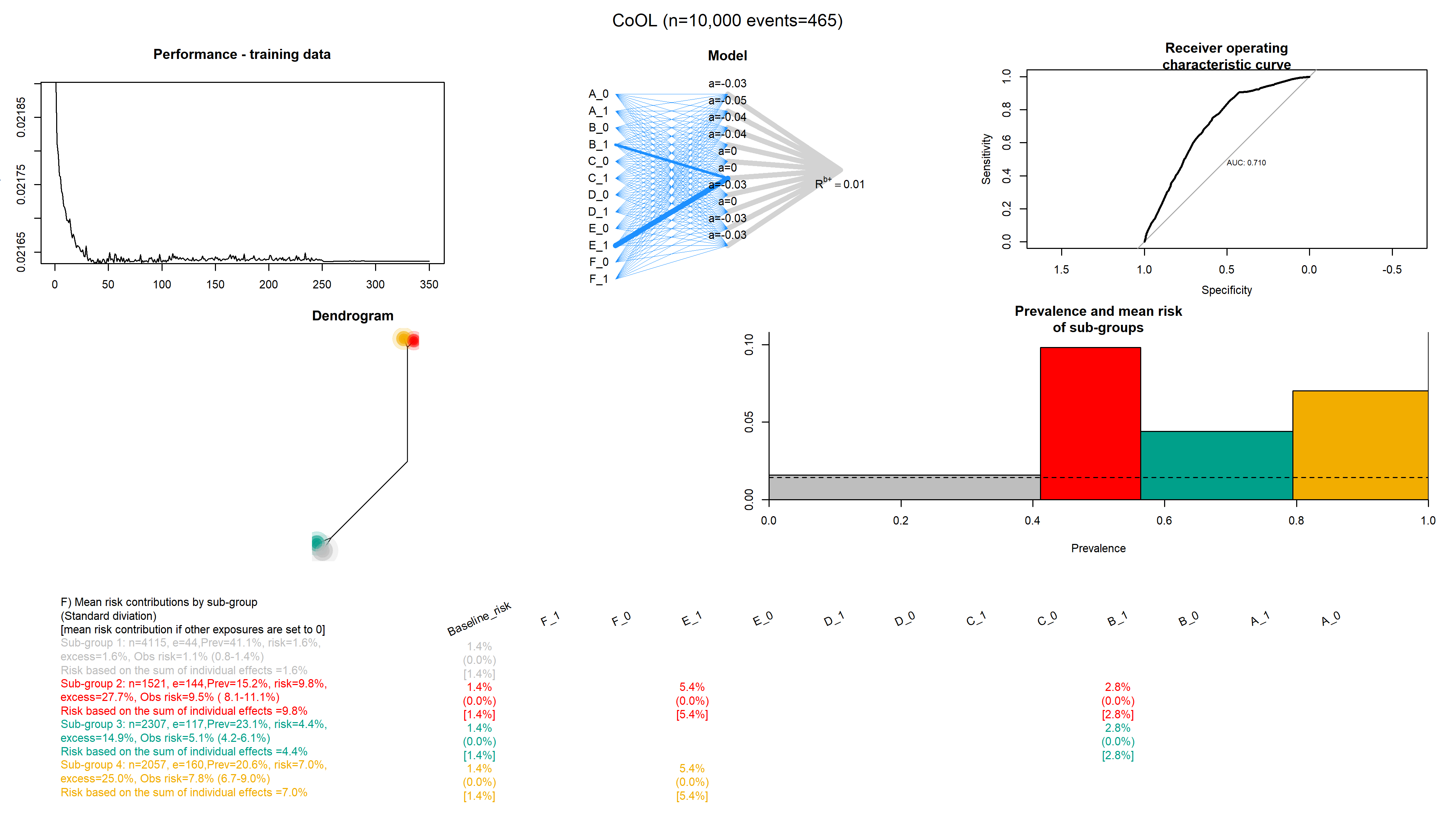

### Common_9.png

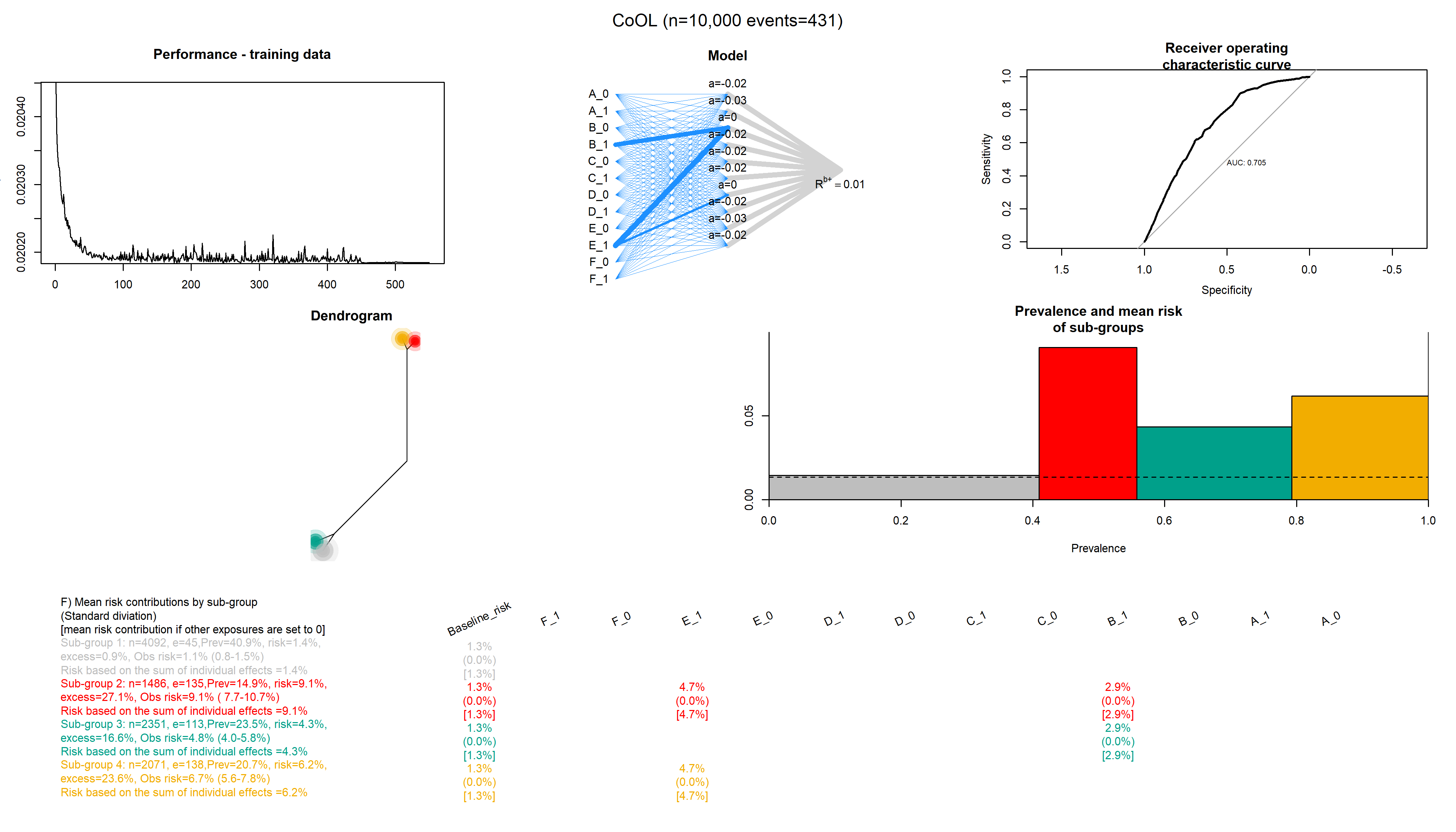

### Common_10.png

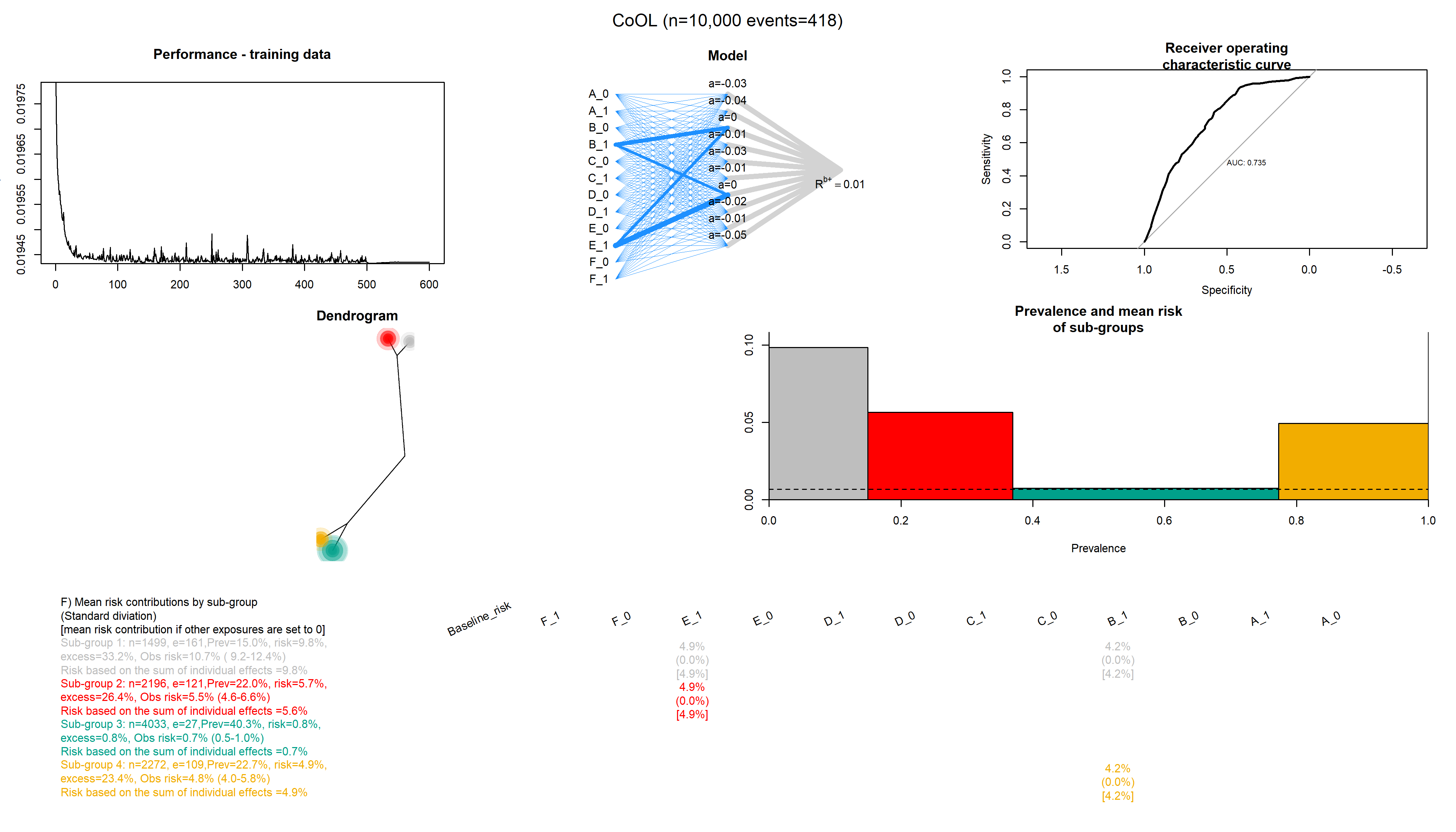

### Complex_1.png

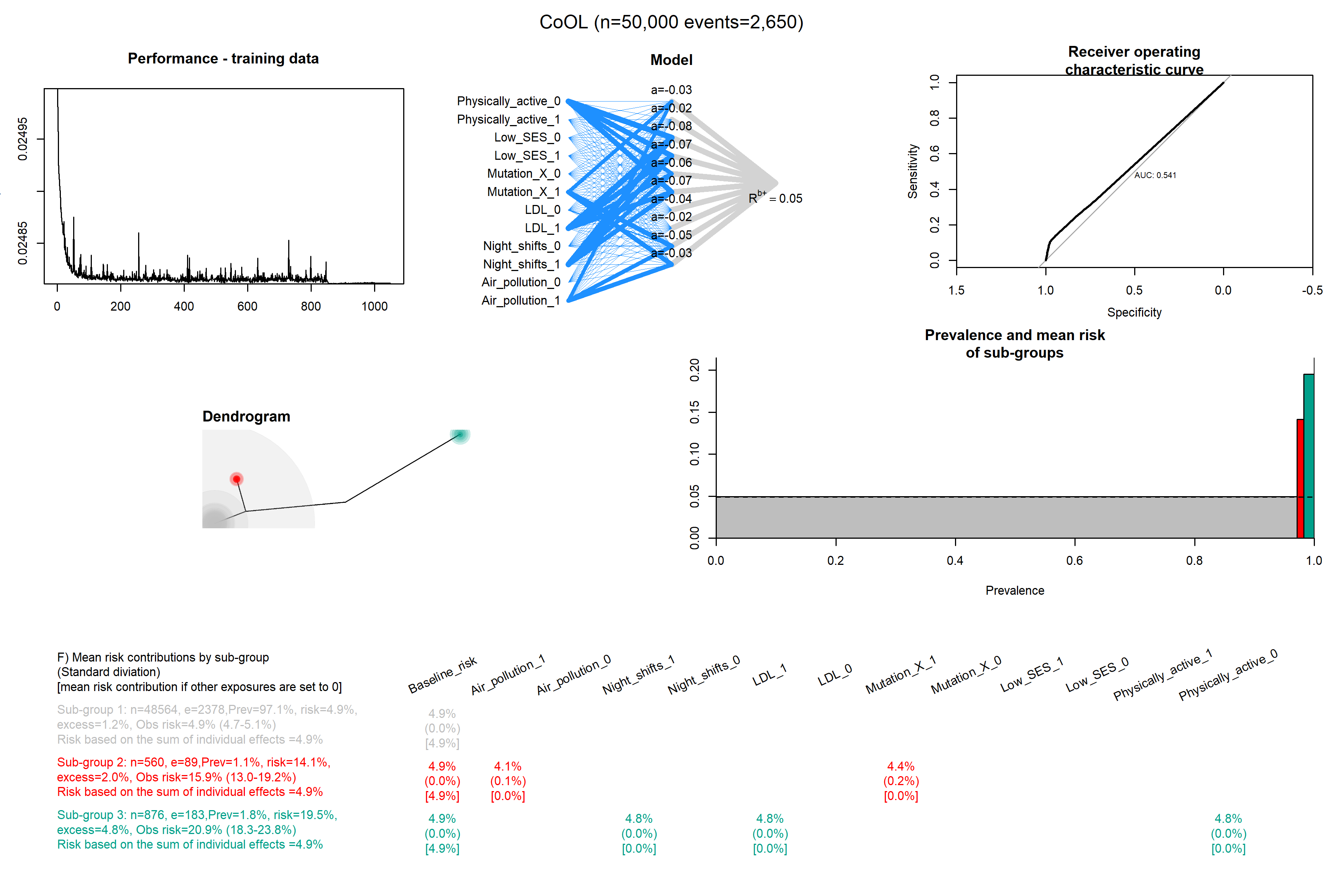

### Complex_2.png

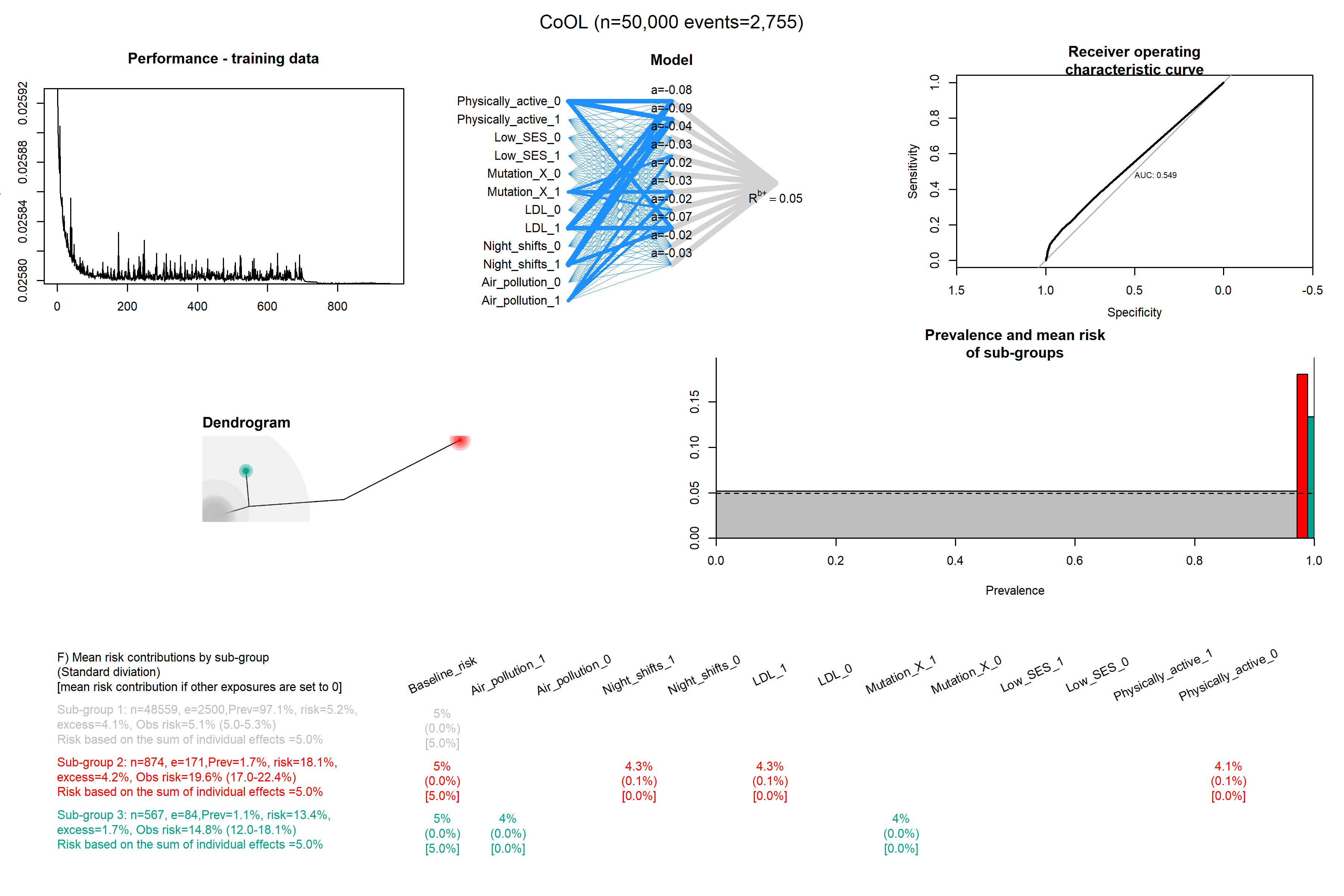

### Complex_3.png

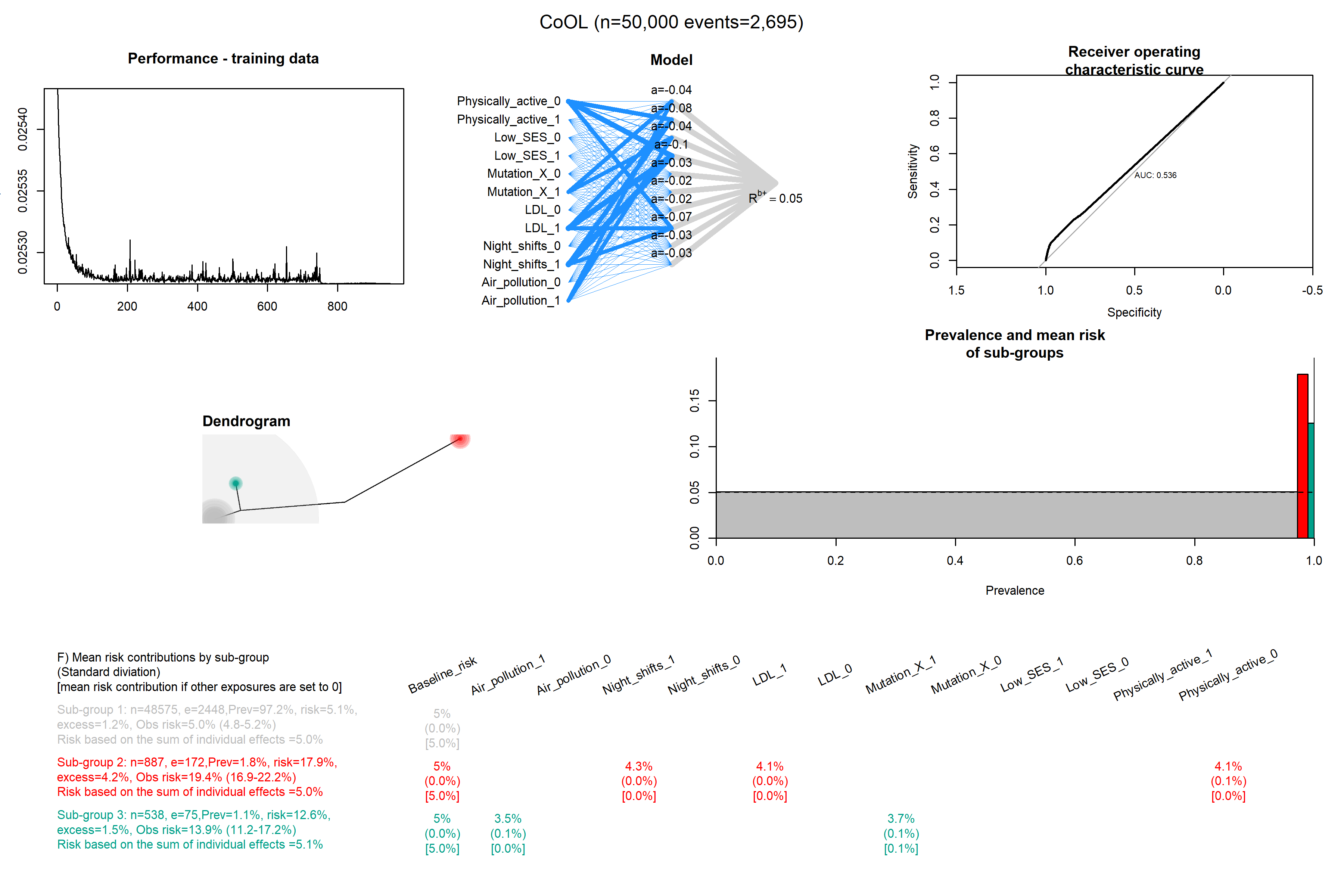

### Complex_4.png

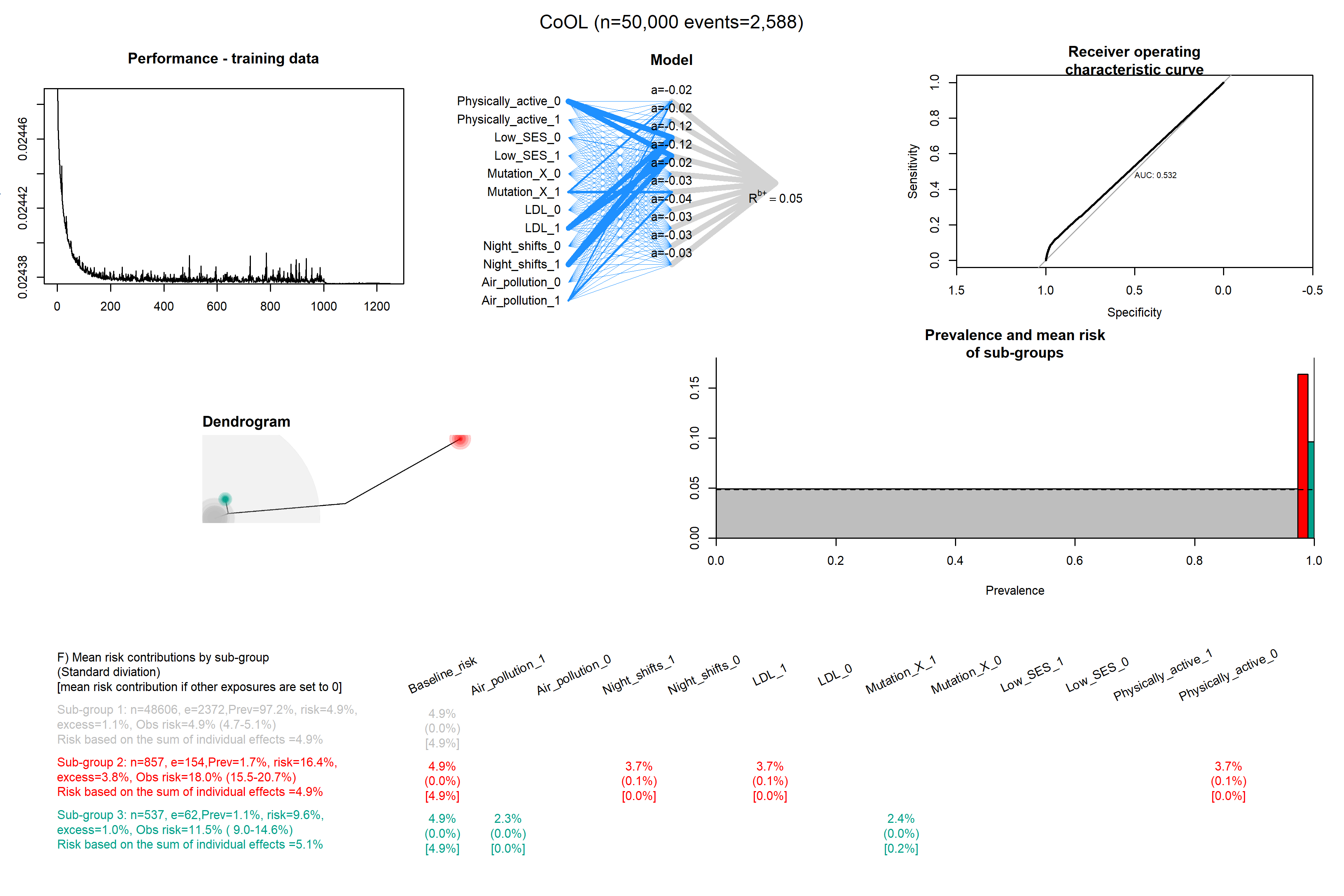

### Complex_5.png

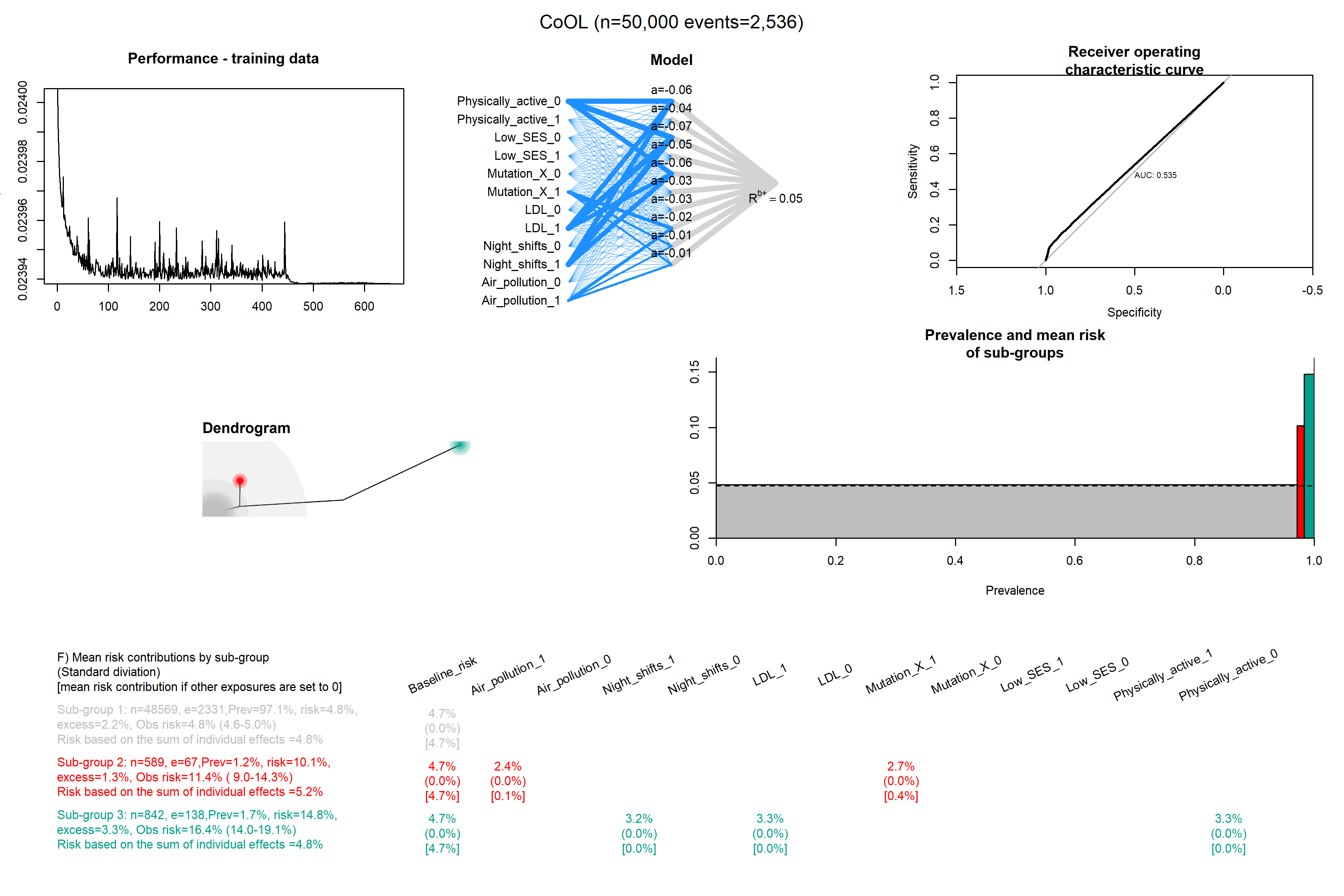

### Complex_6.png

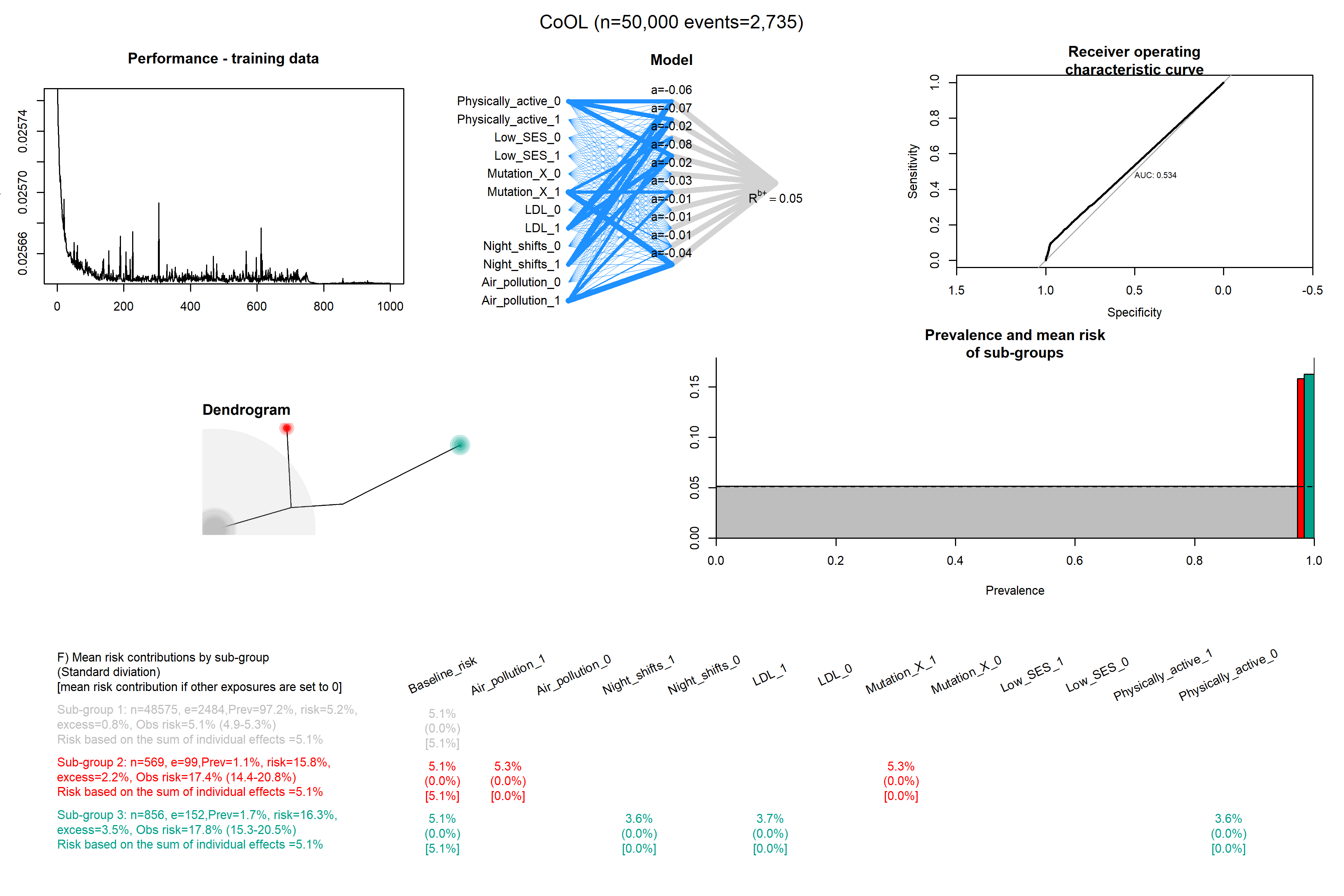

### Complex_7.png

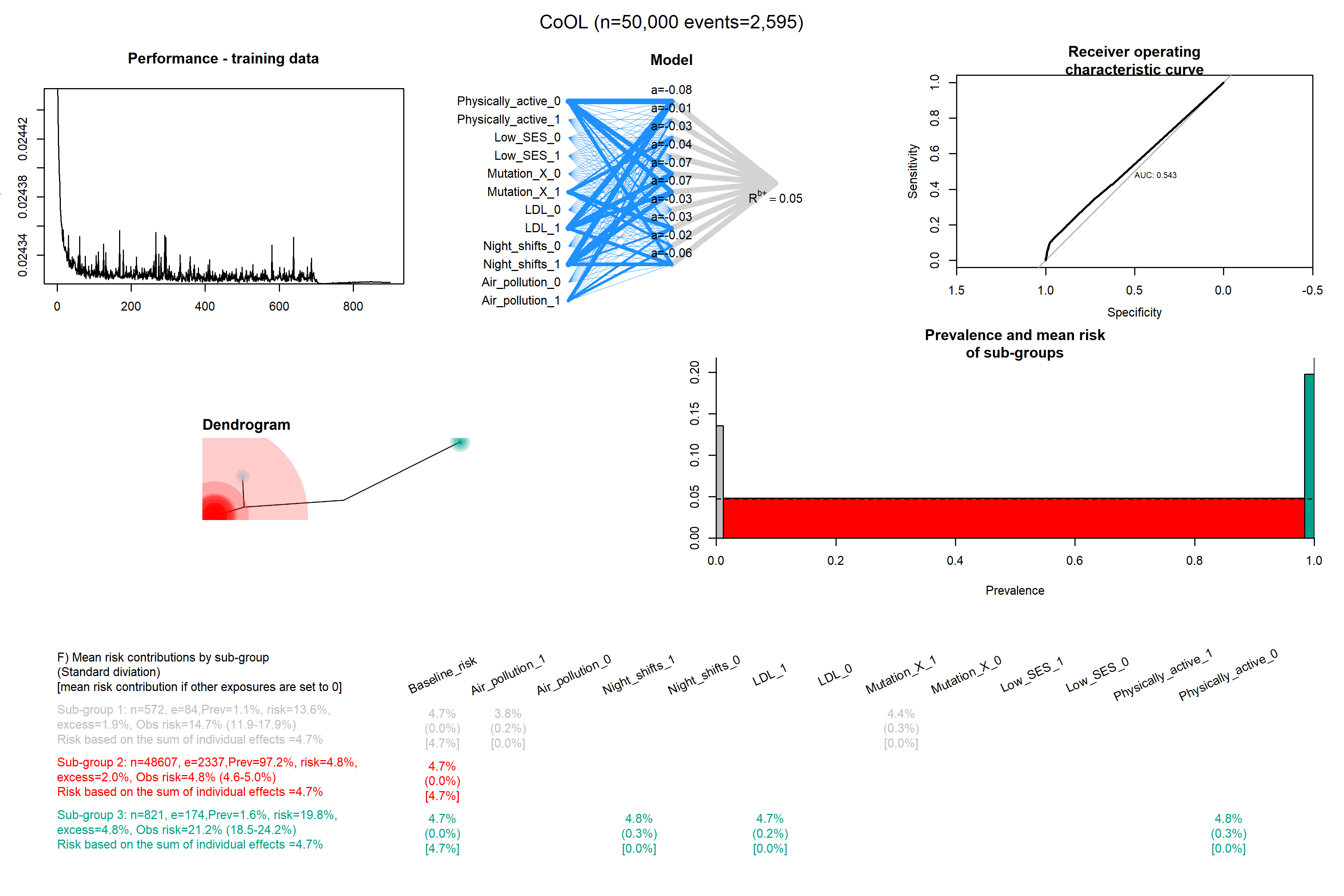

### Complex_8.png

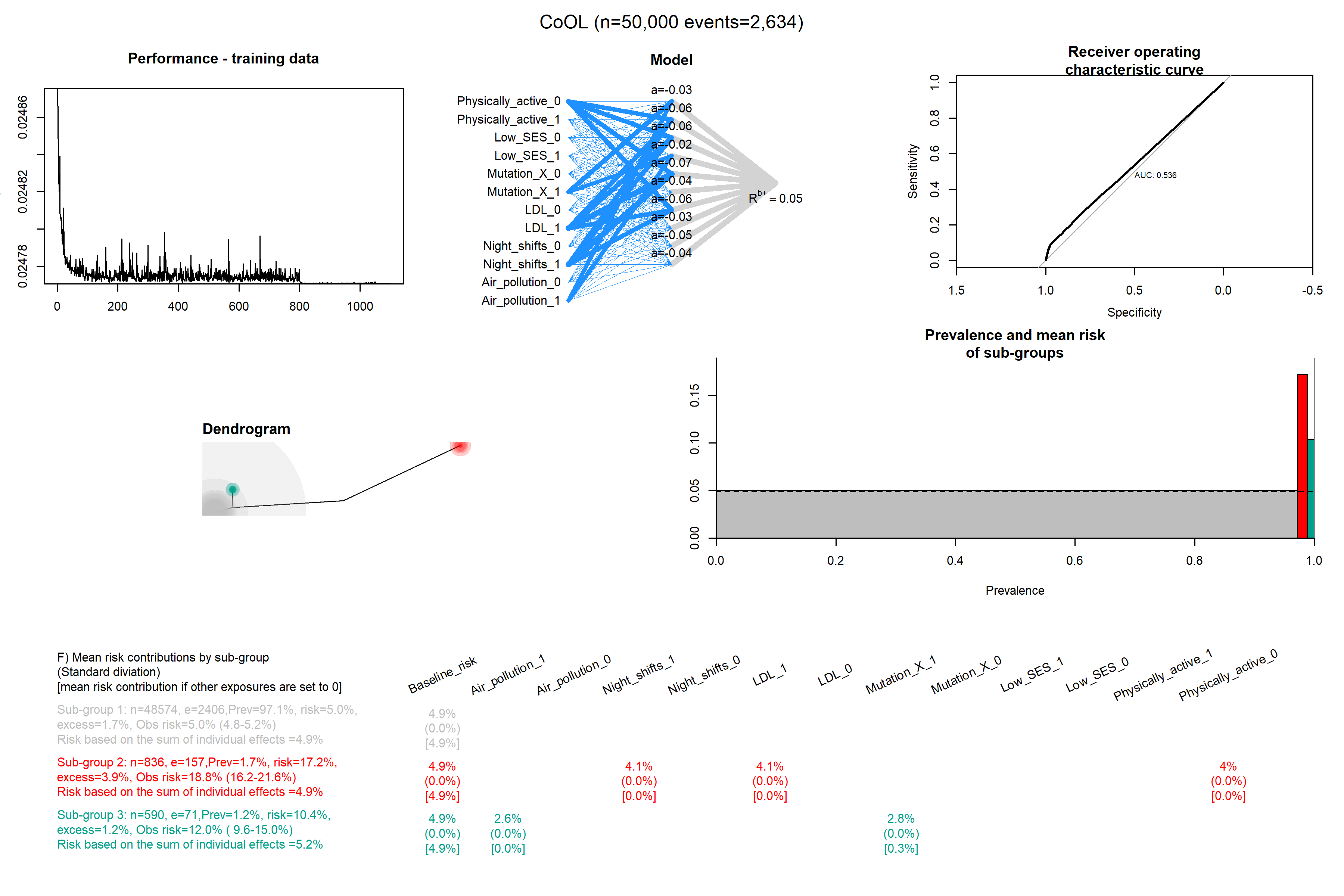

### Complex_9.png

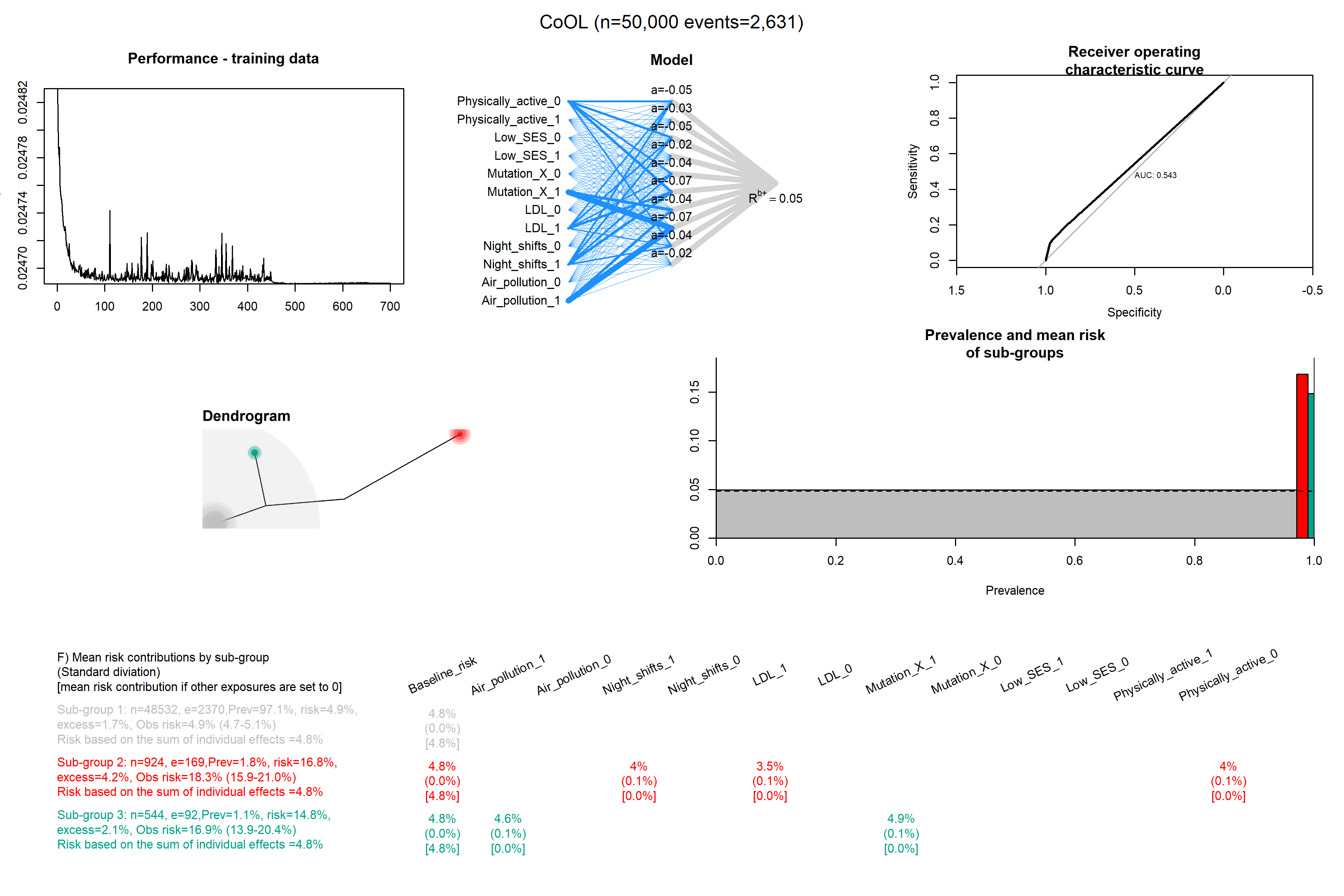

### Complex_10.png

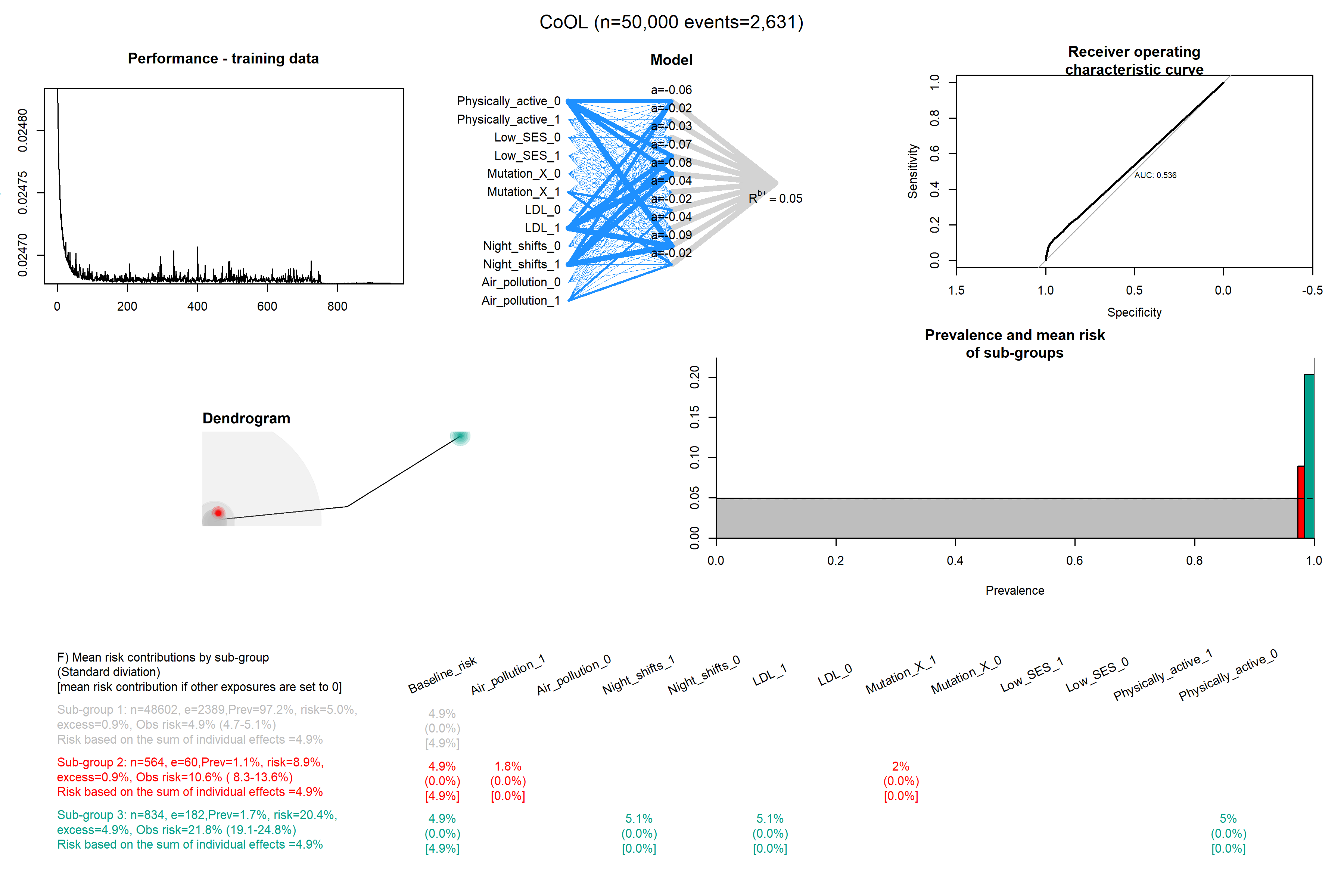

### Confounding_with_c_1.png

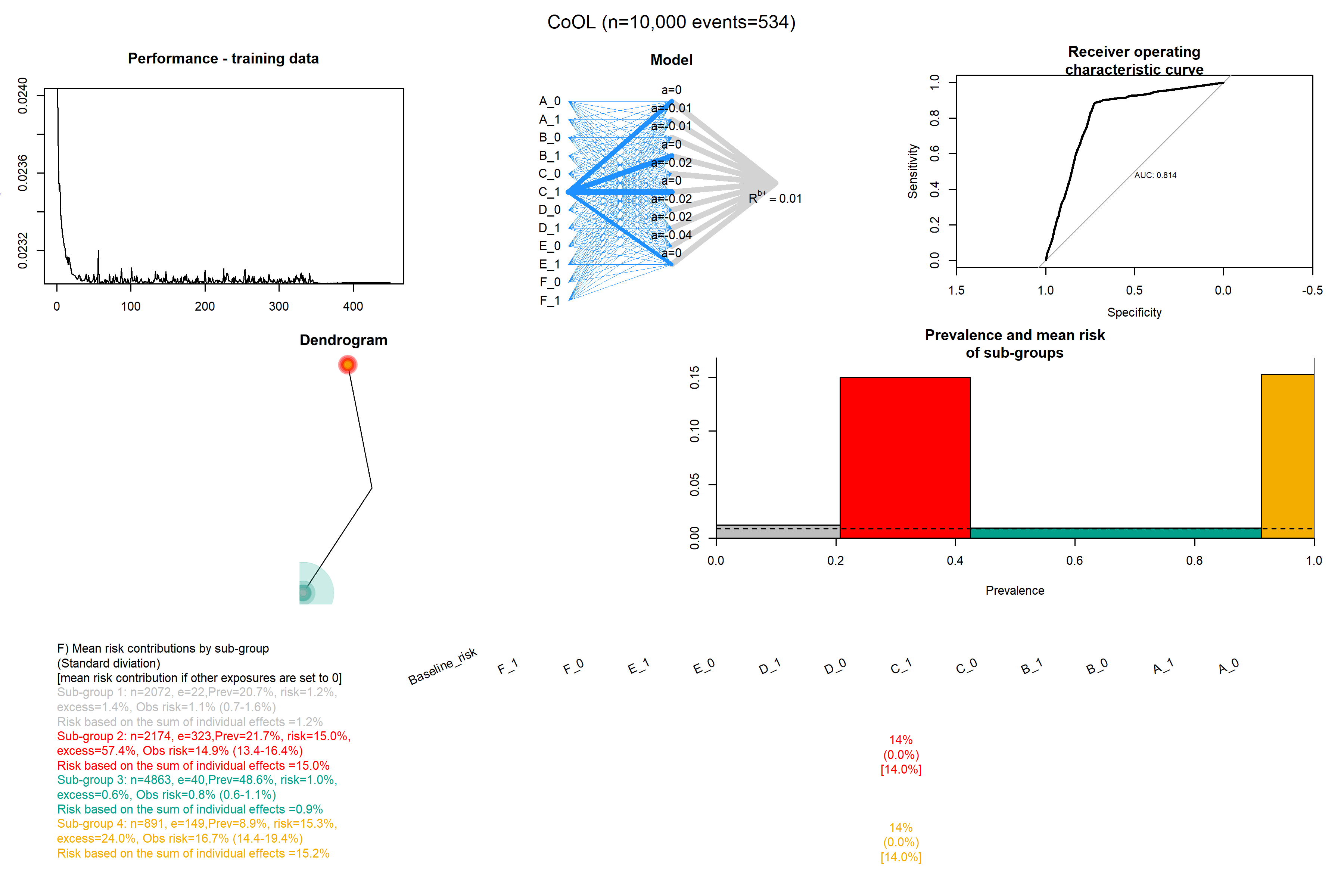

### Confounding_with_c_2.png

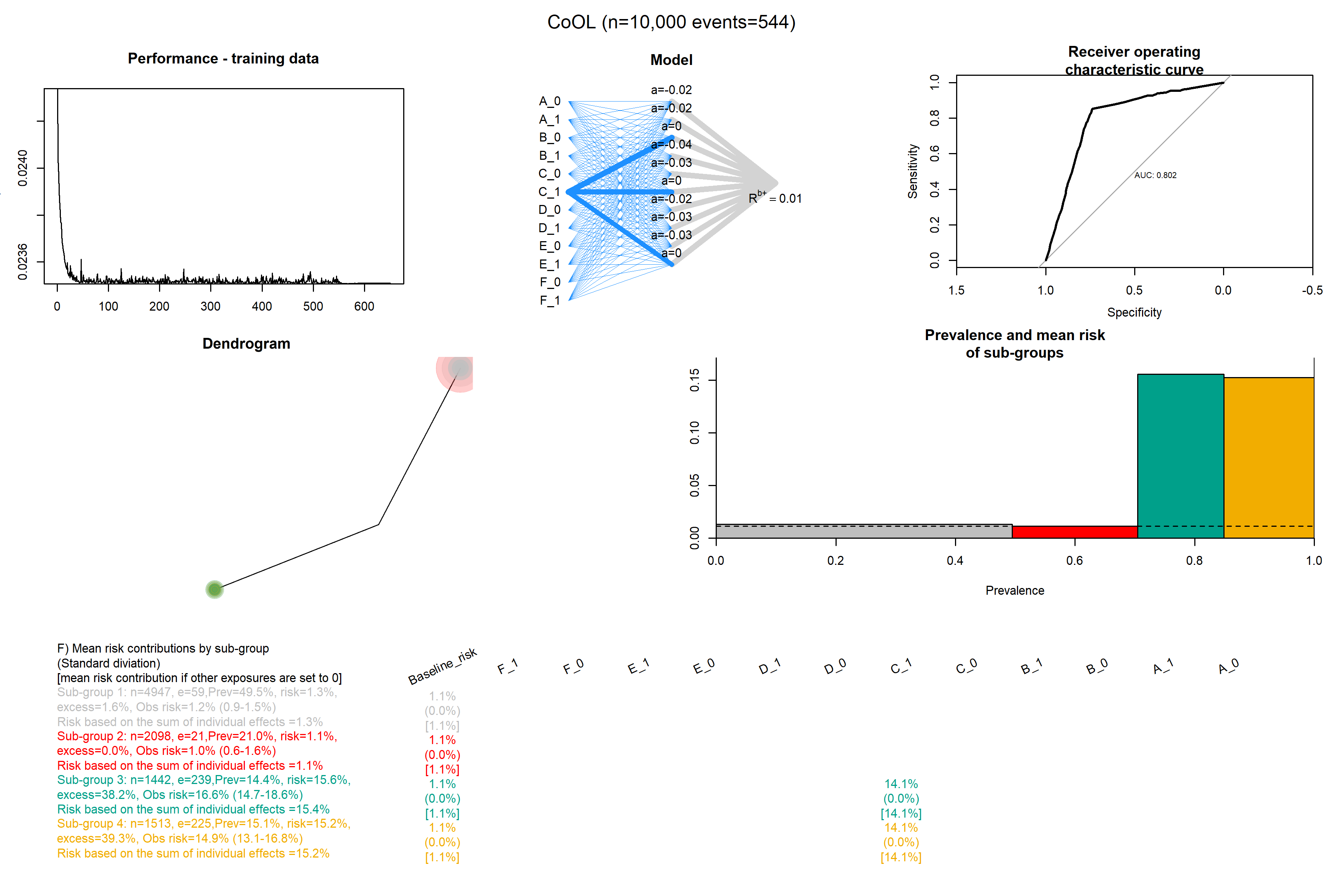

### Confounding_with_c_3.png

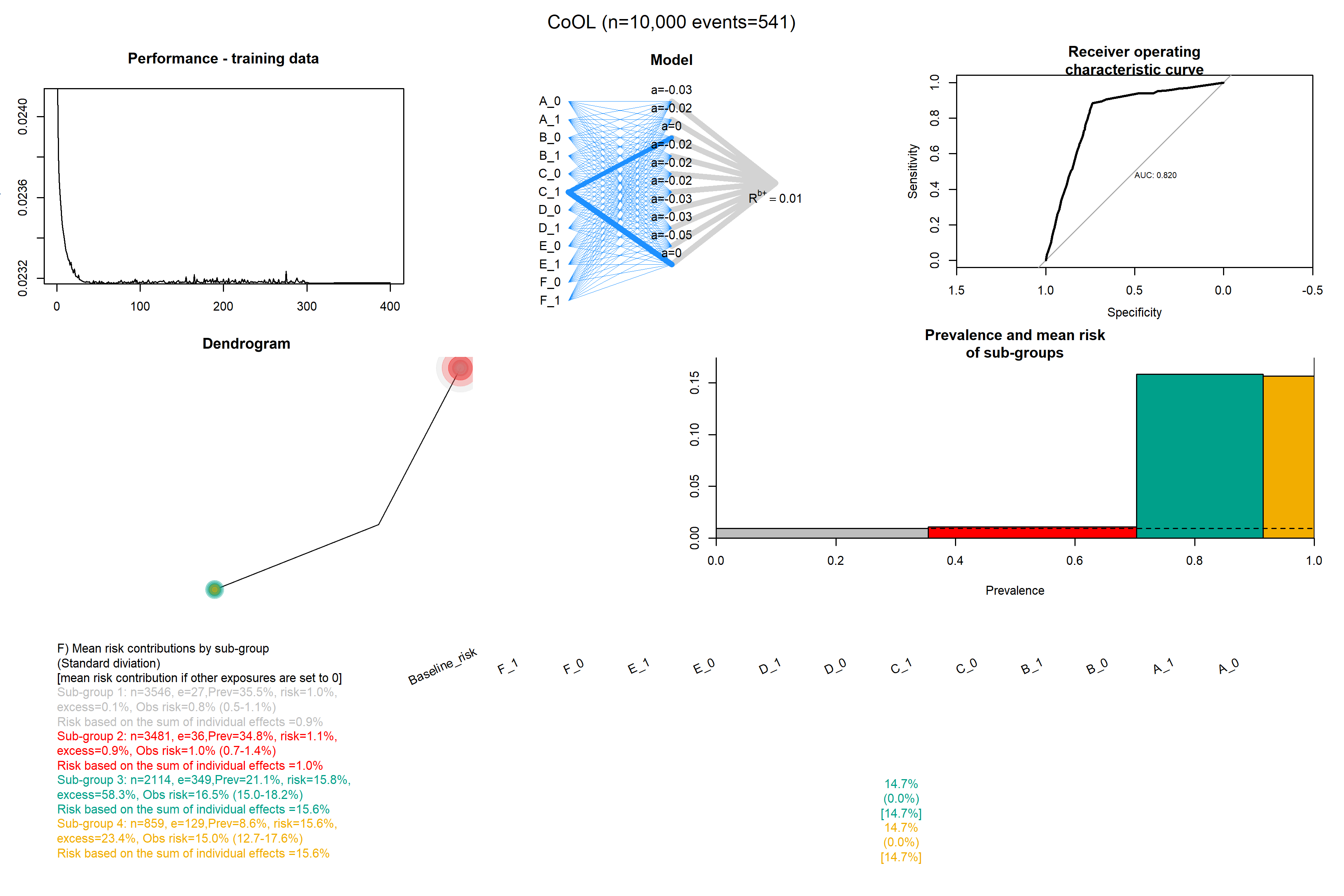

### Confounding_with_c_4.png

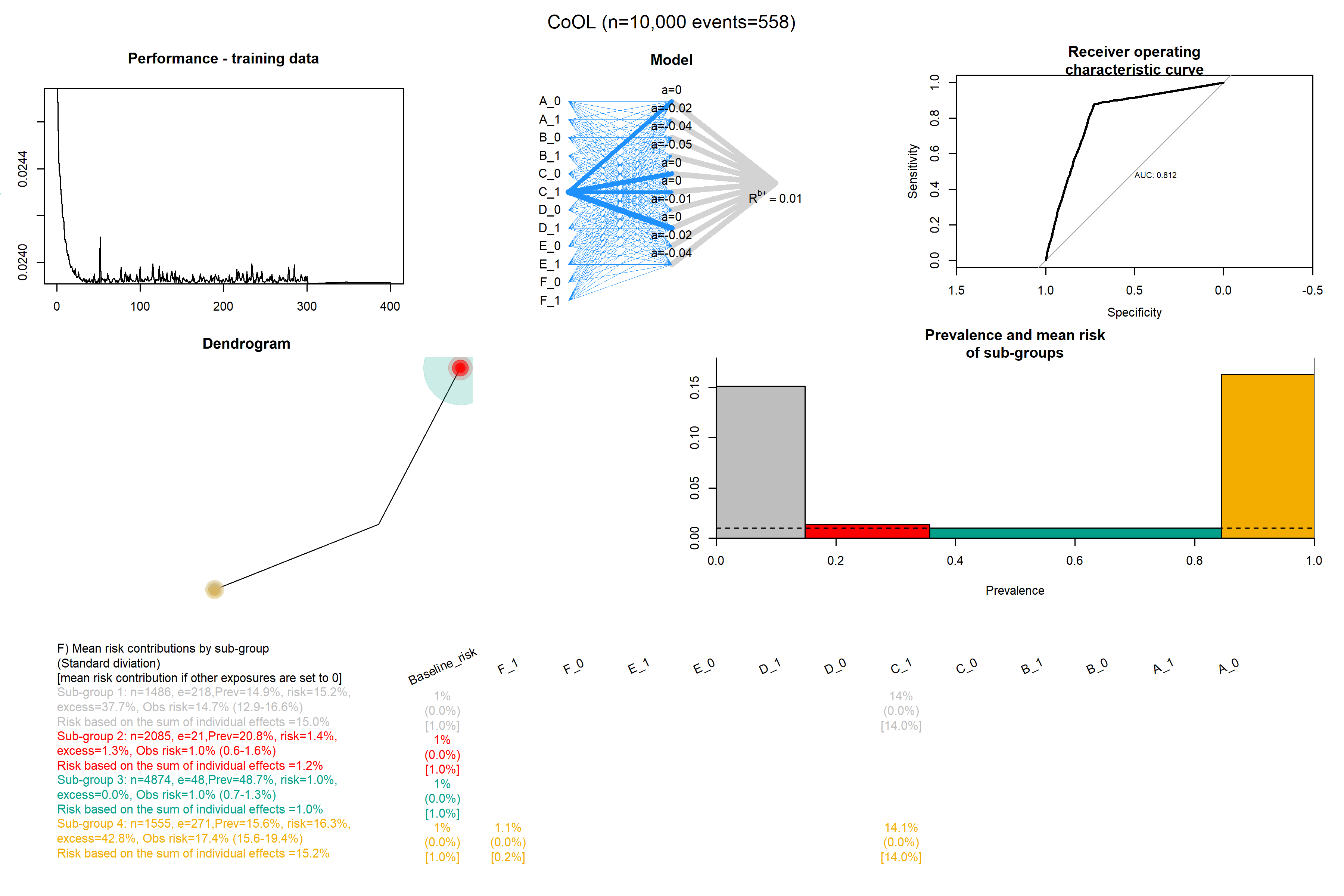

### Confounding_with_c_5.png

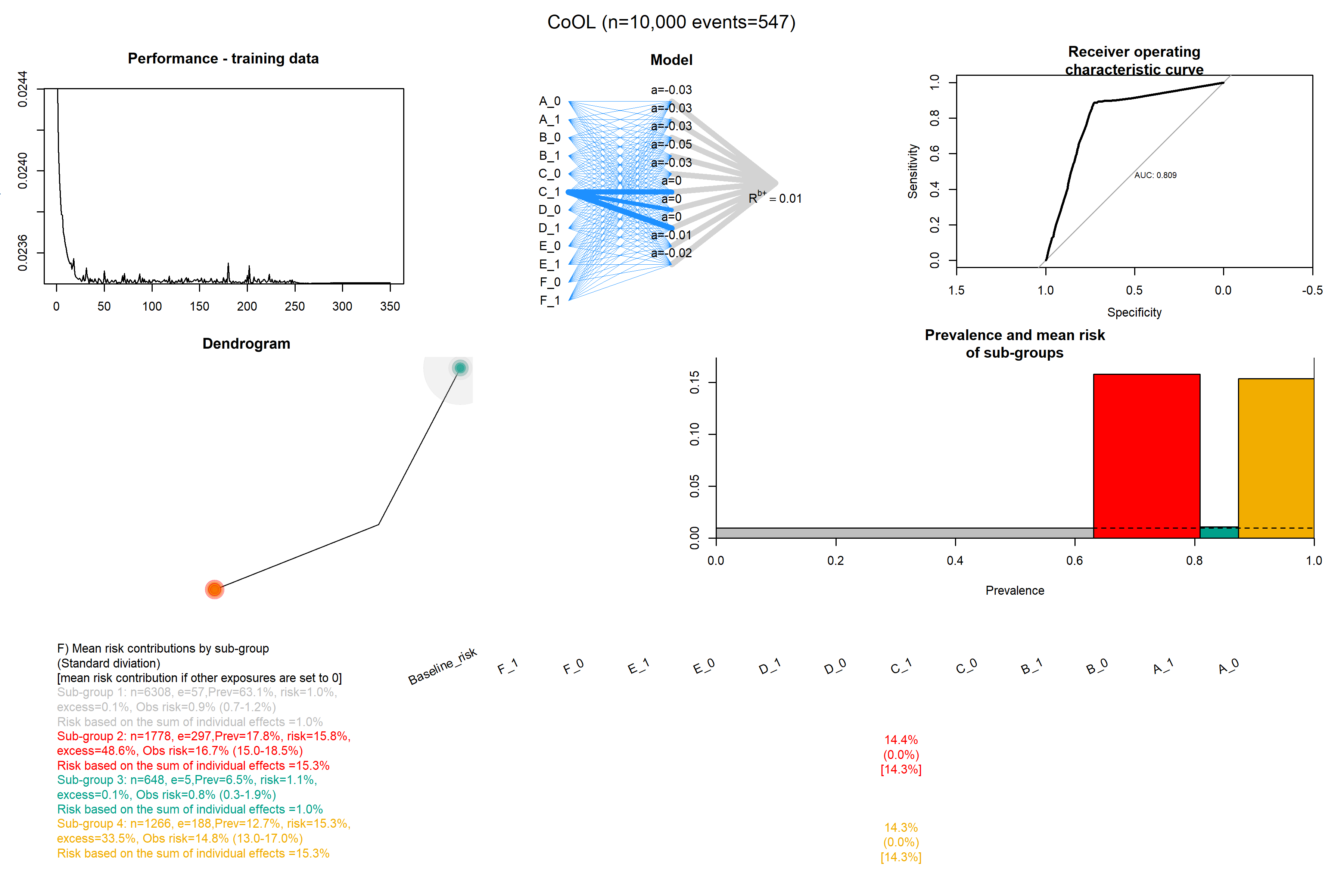

### Confounding_with_c_6.png

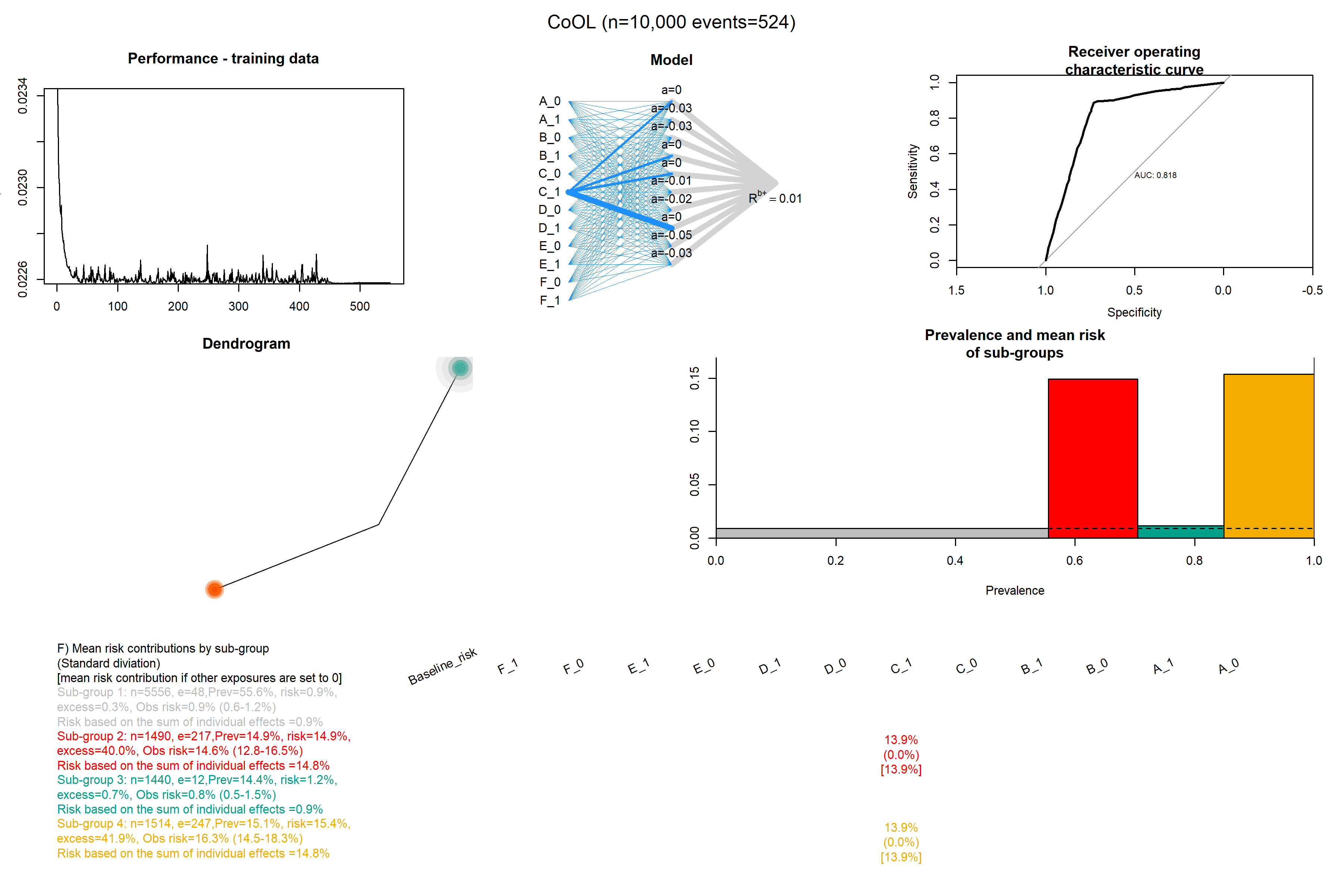

### Confounding_with_c_7.png

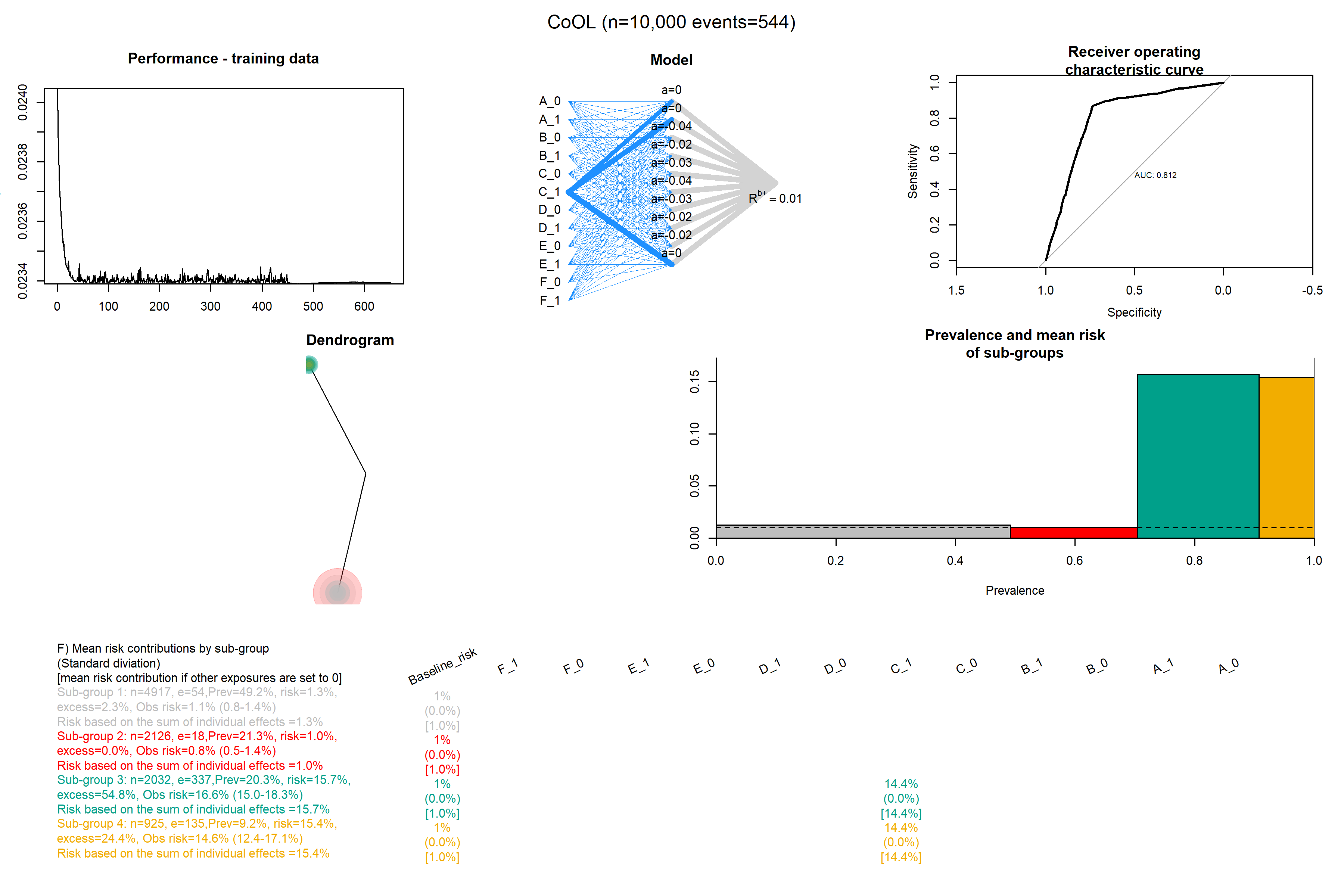

### Confounding_with_c_8.png

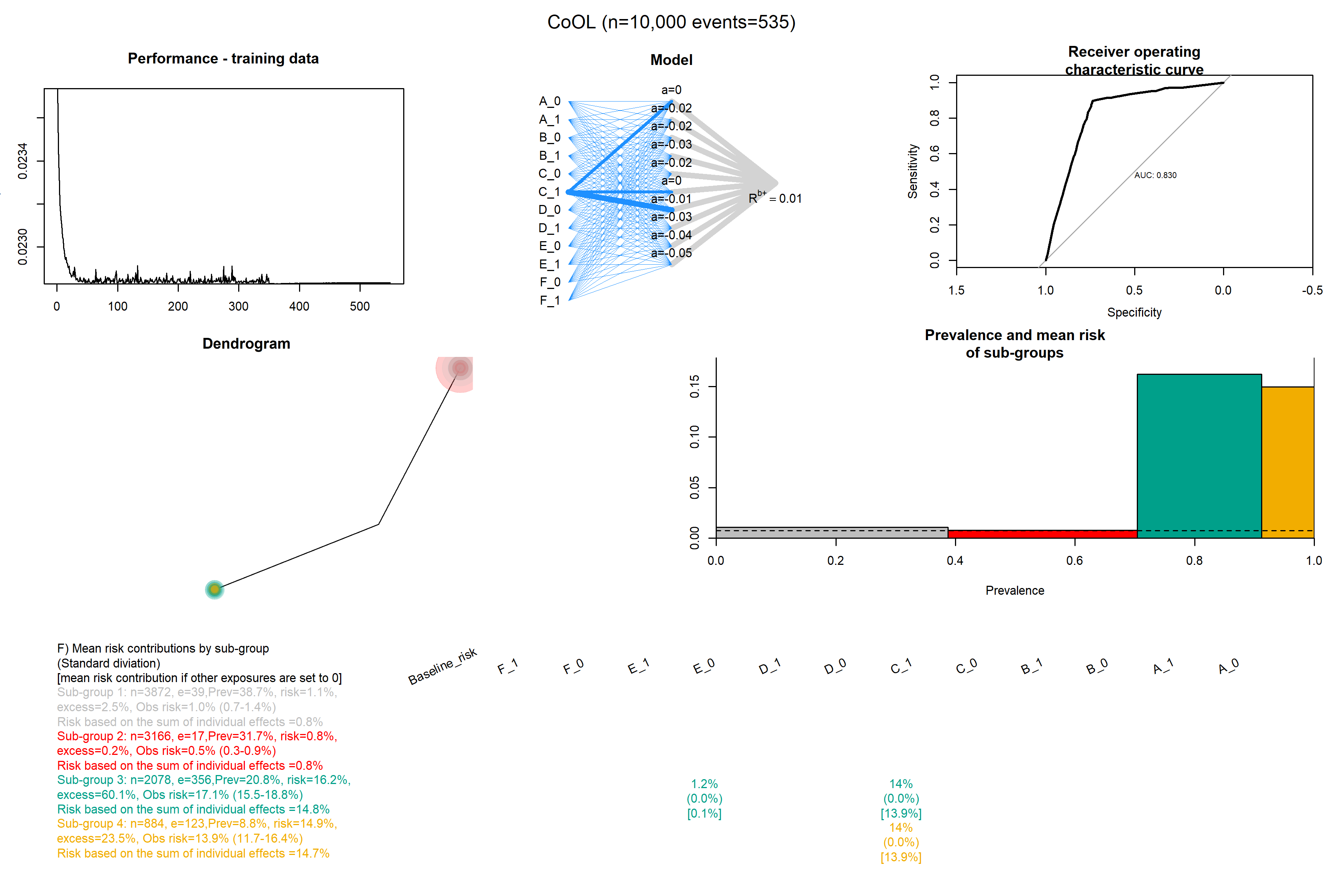

### Confounding_with_c_9.png

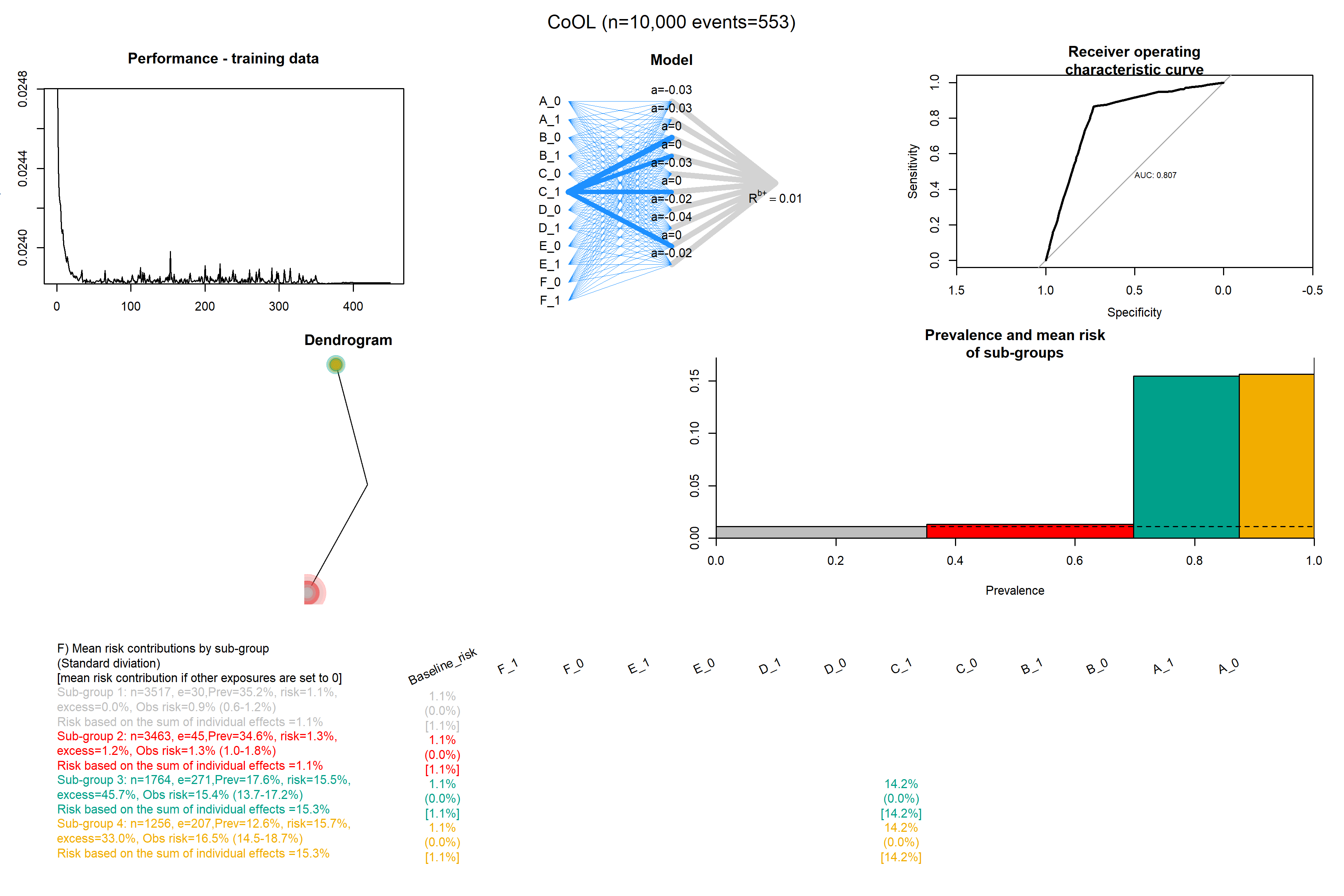

### Confounding_with_c_10.png

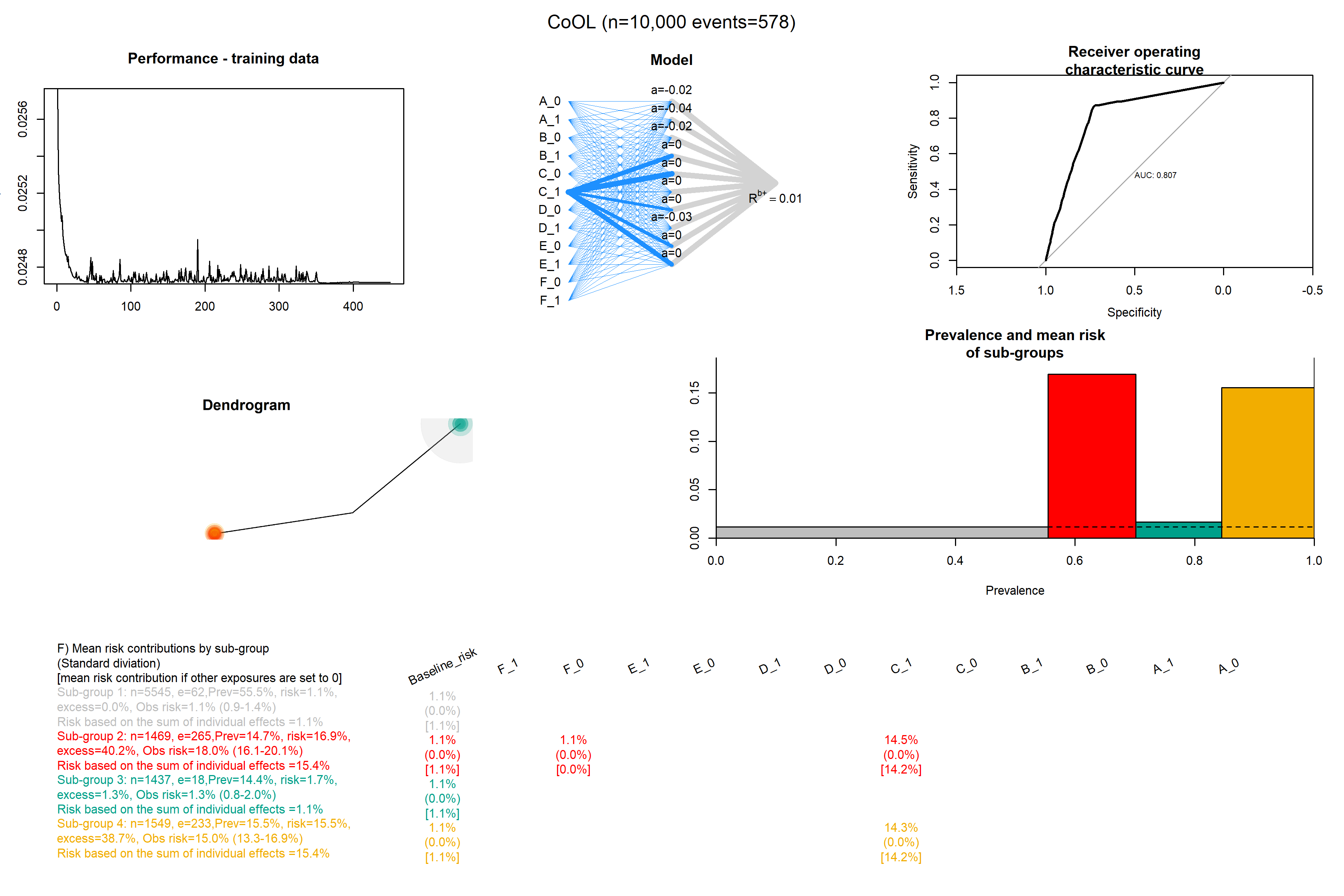

### Confounding_without_c_1.png

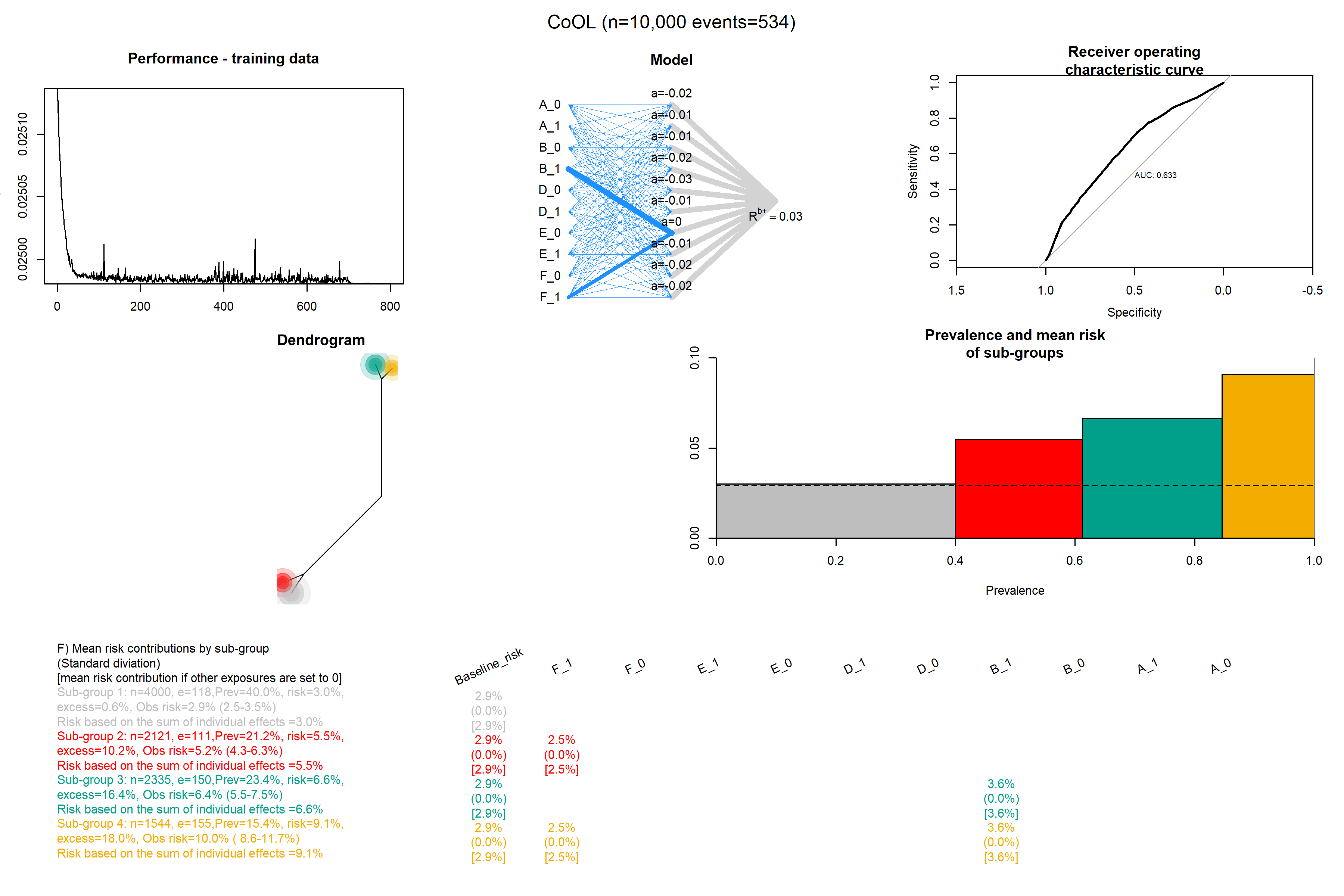

### Confounding_without_c_2.png

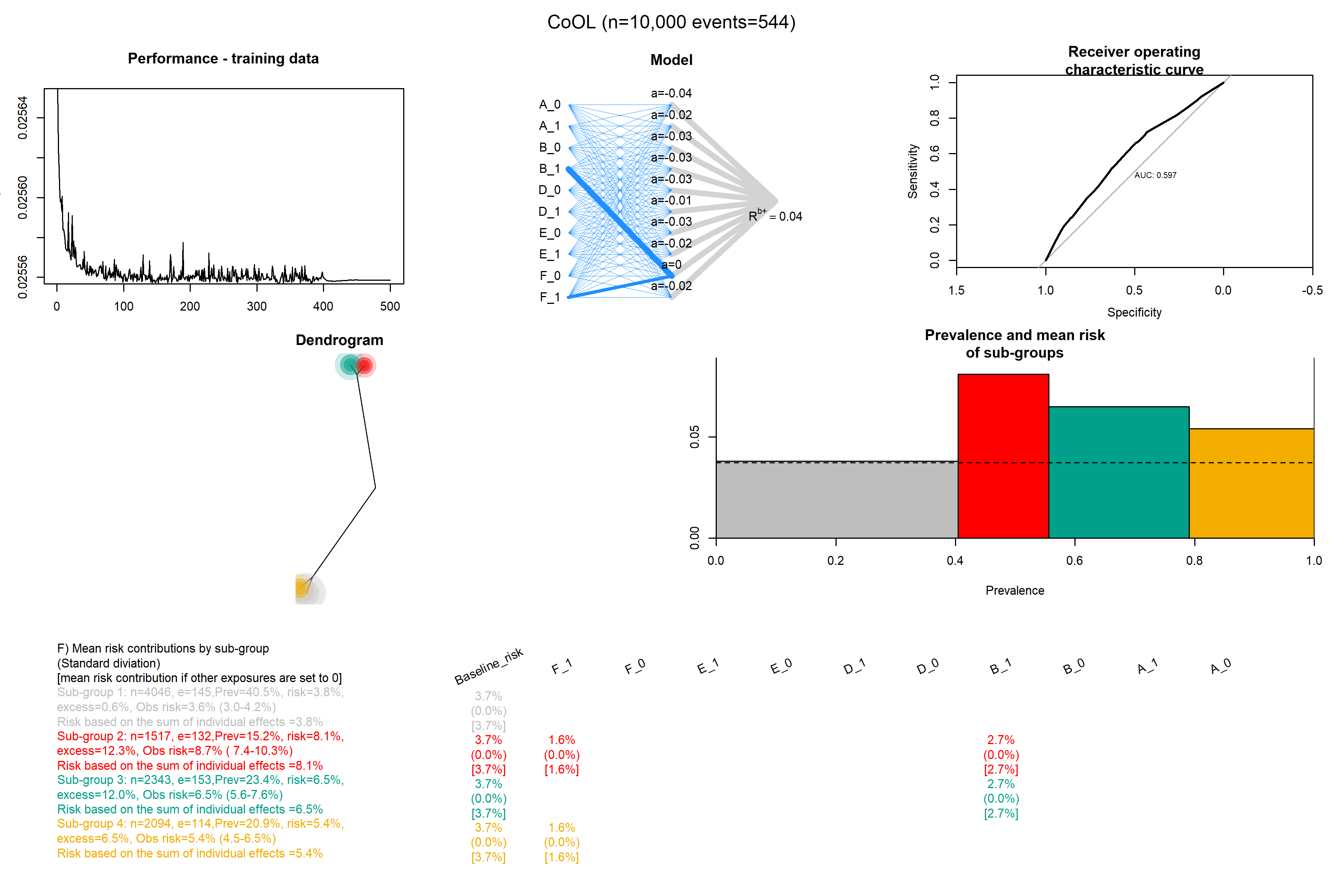

### Confounding_without_c_3.png

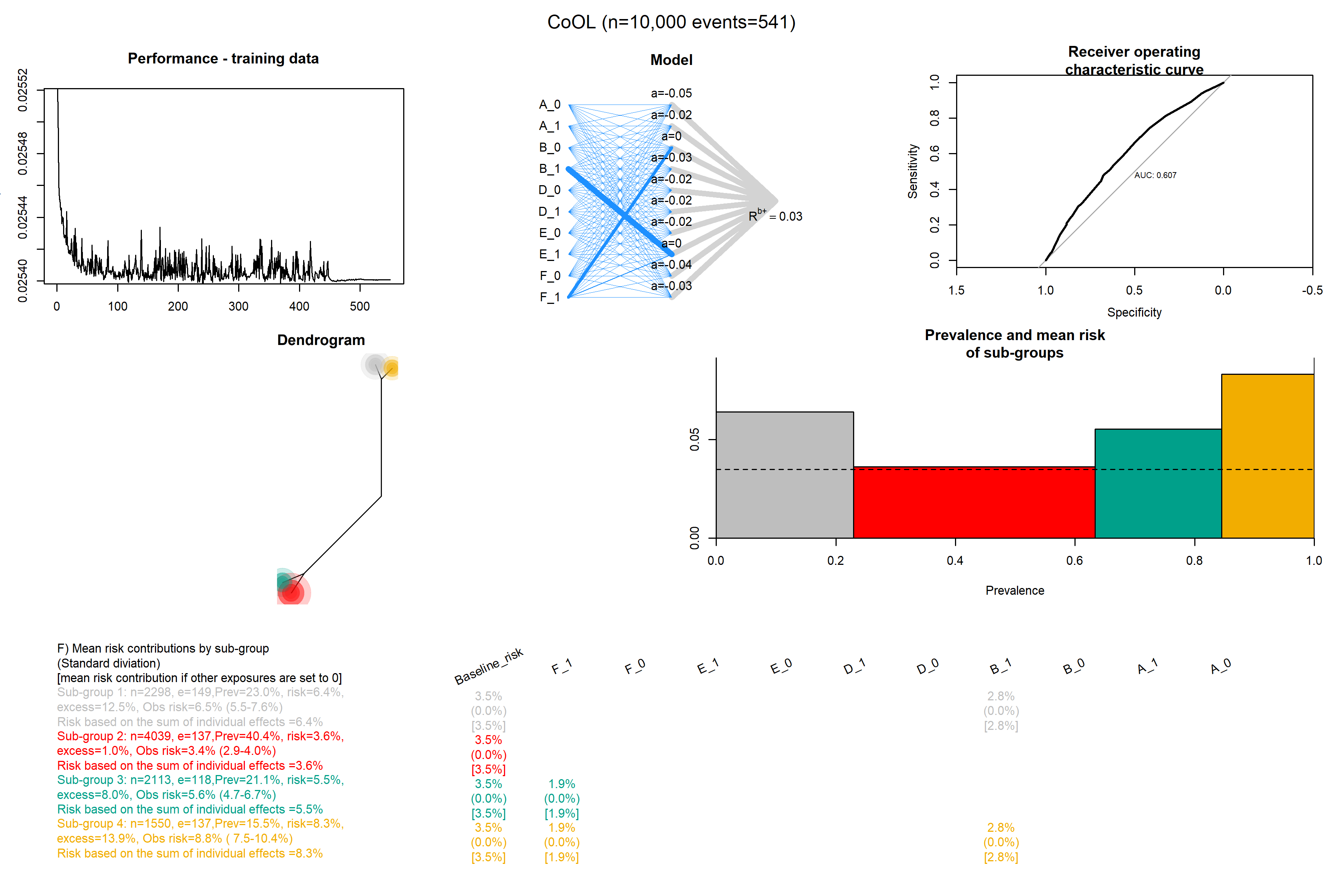

### Confounding_without_c_4.png

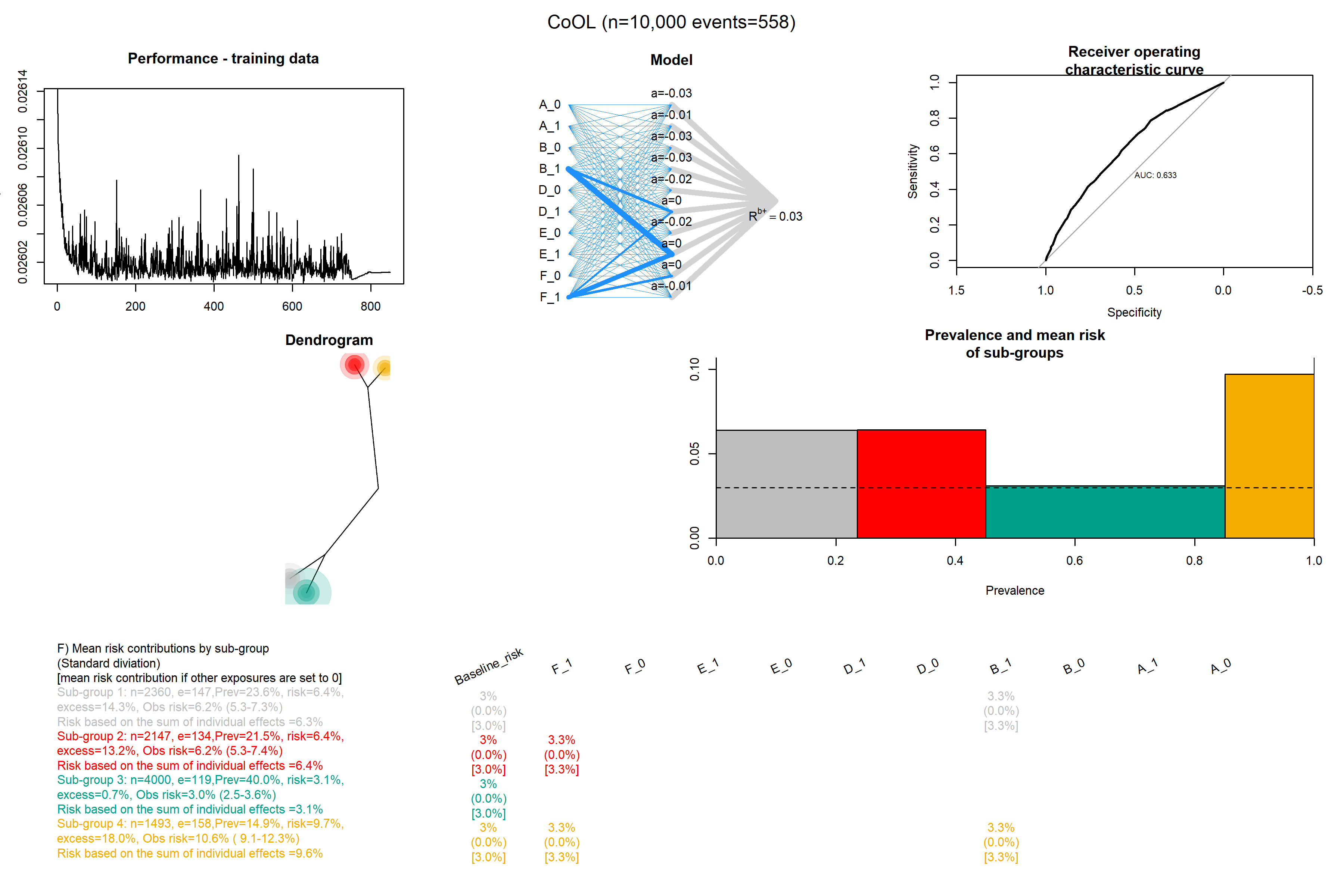

### Confounding_without_c_5.png

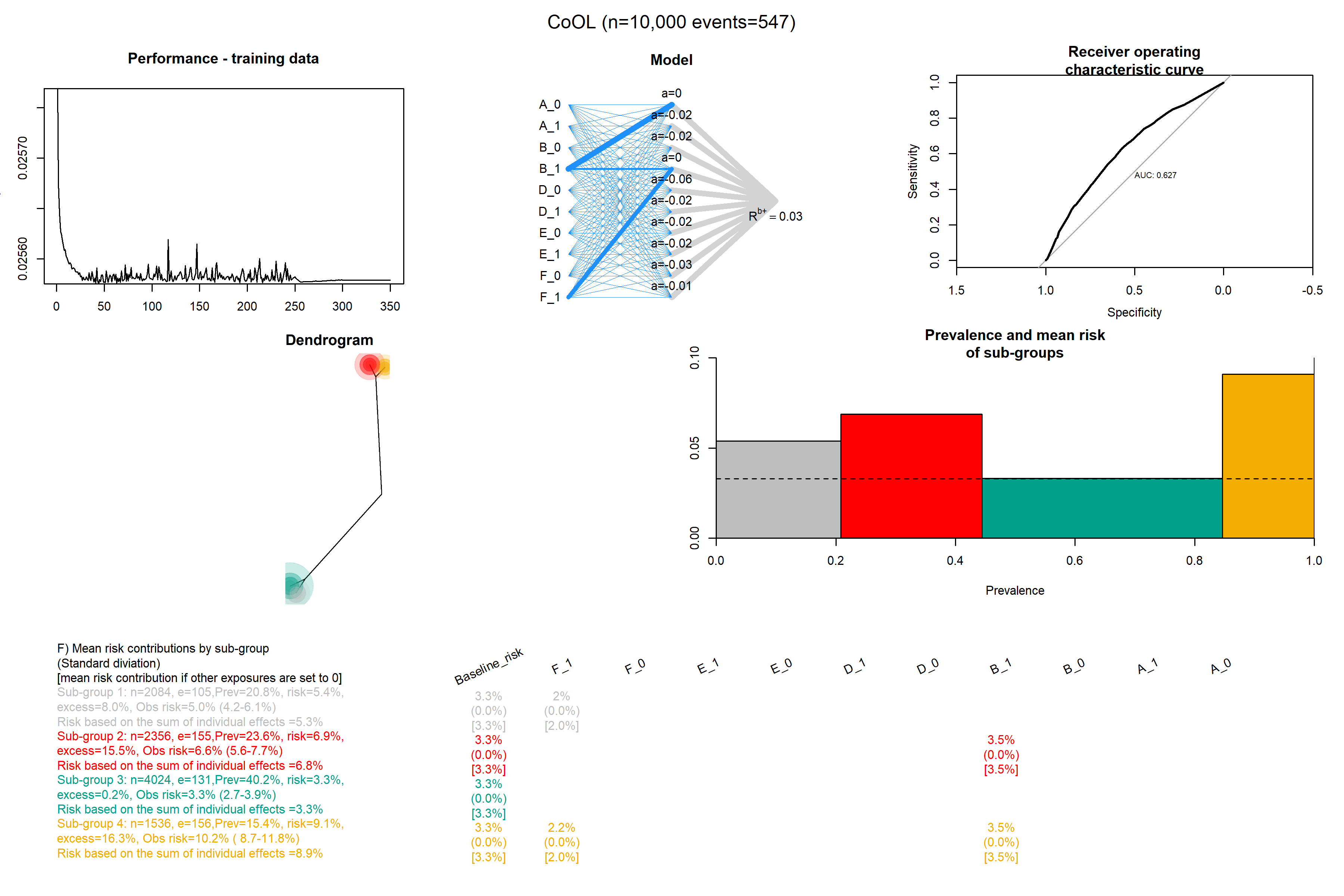

### Confounding_without_c_6.png

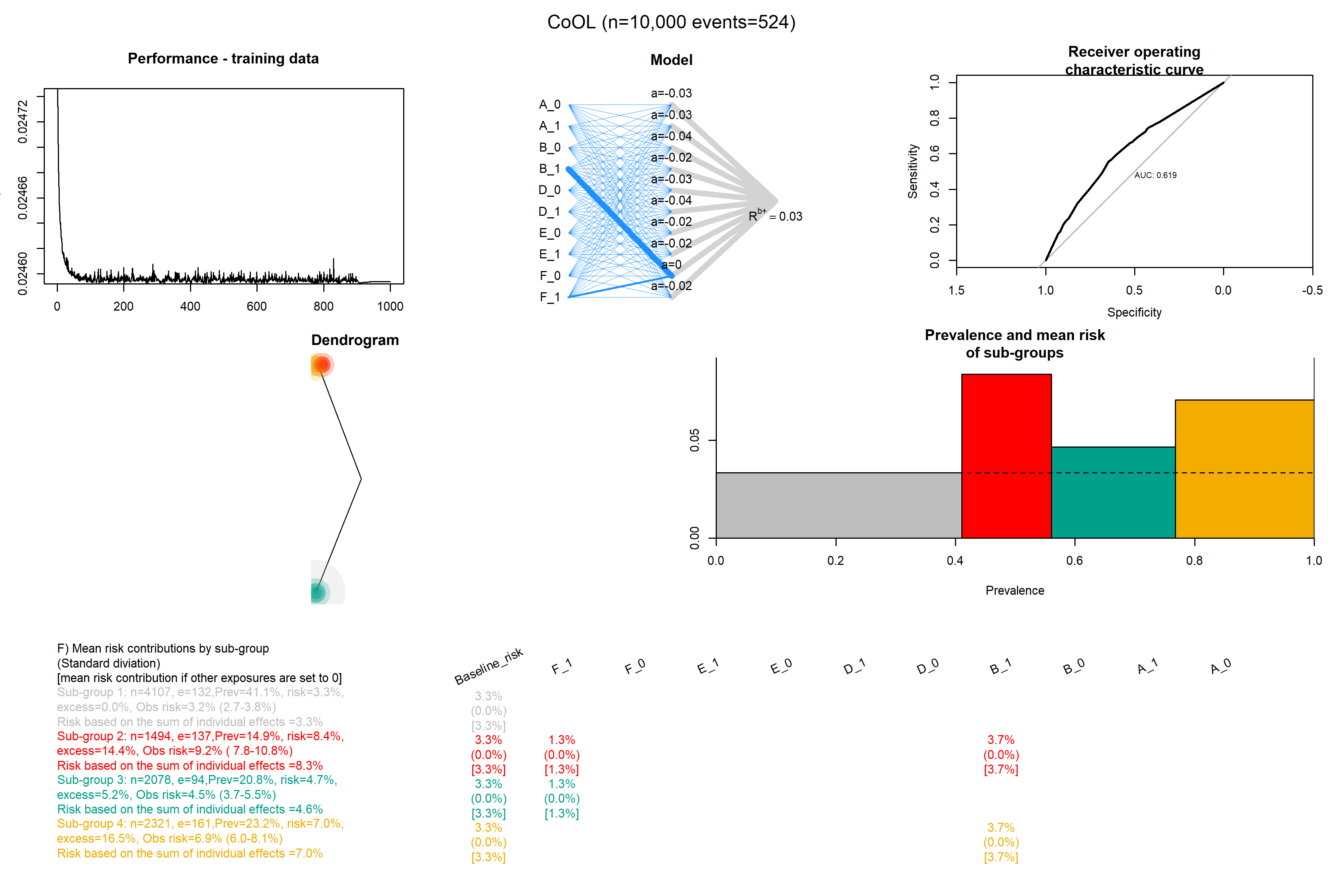

### Confounding_without_c_7.png

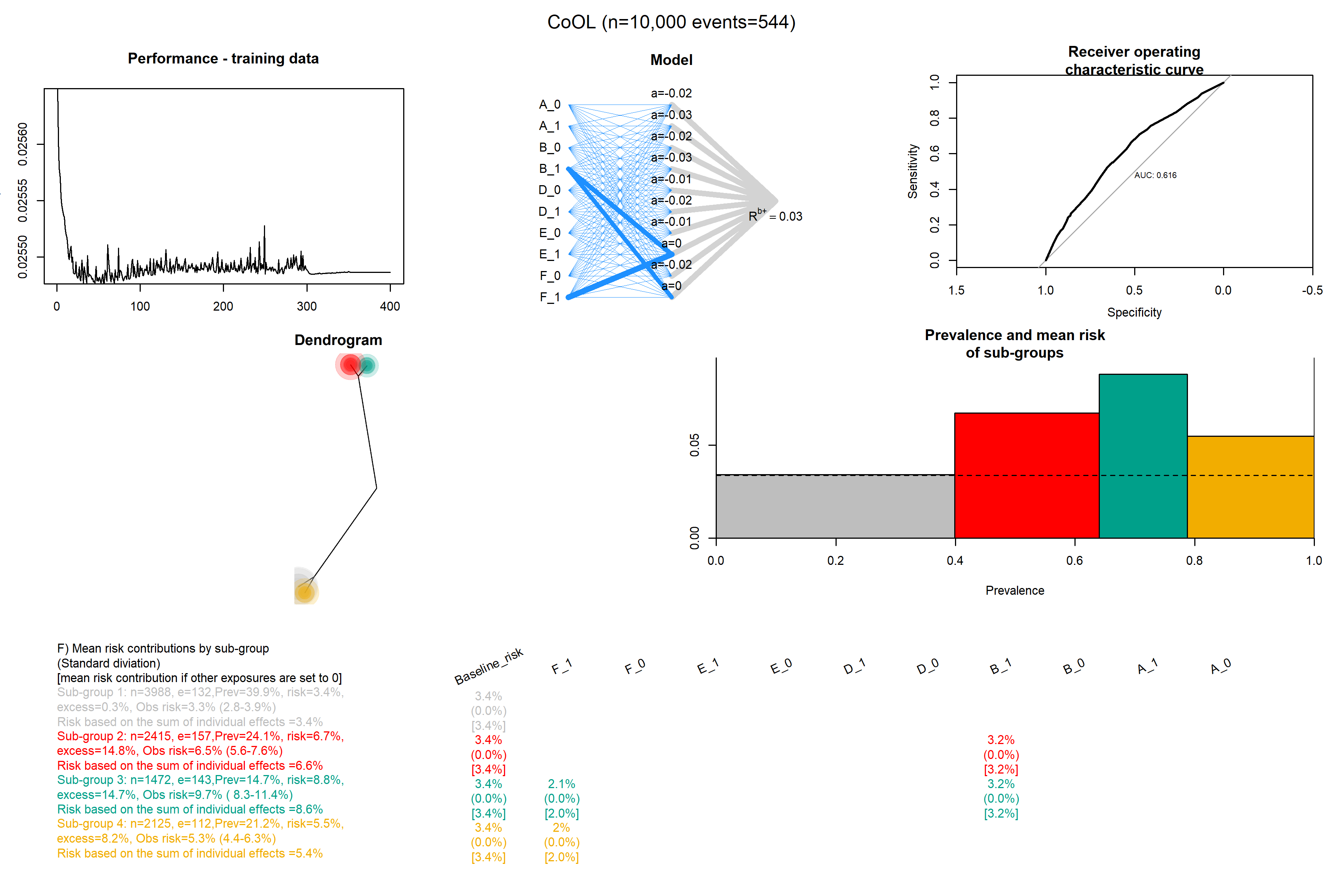

### Confounding_without_c_8.png

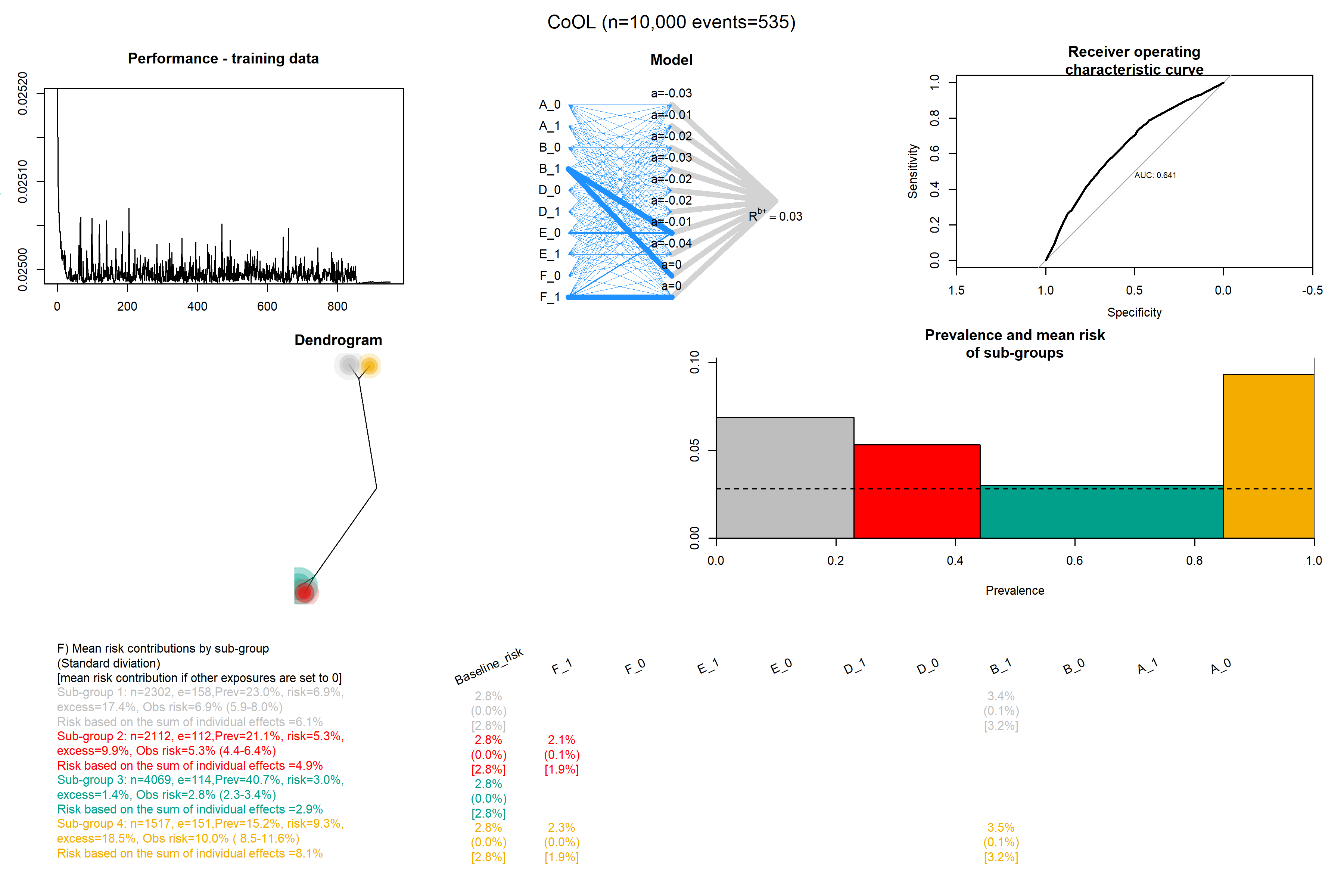

### Confounding_without_c_9.png

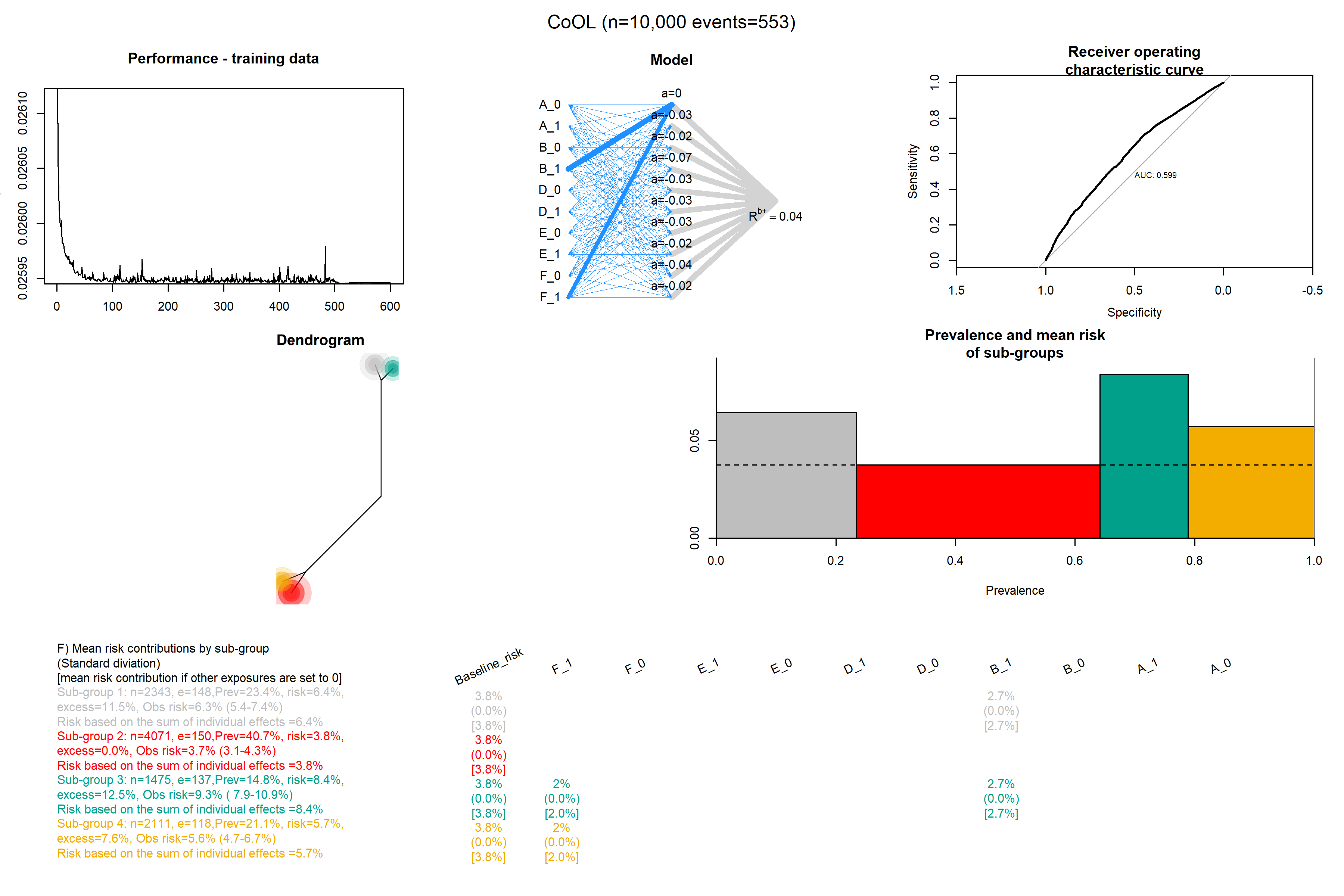

### Confounding_without_c_10.png

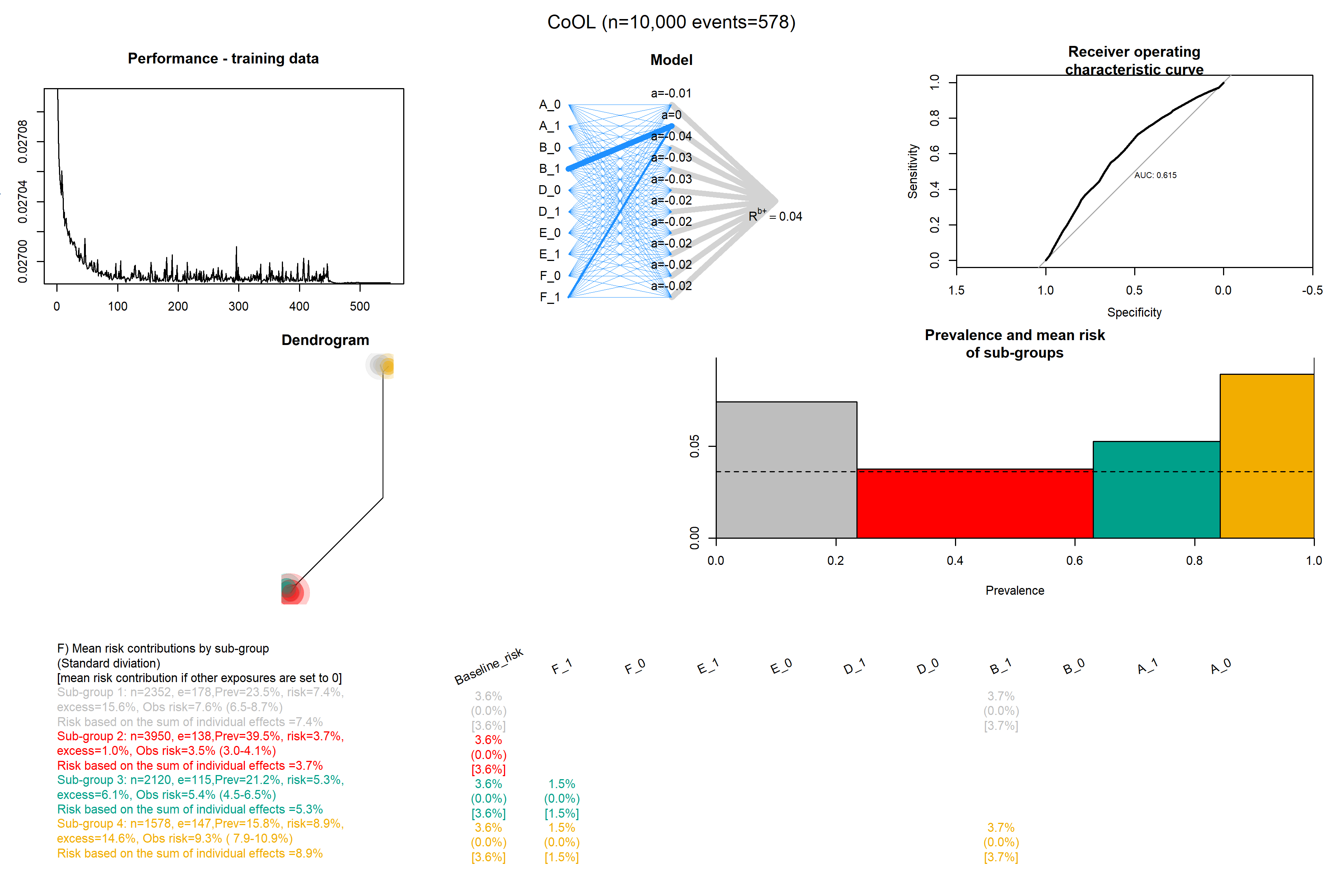

### Data_size_100.png

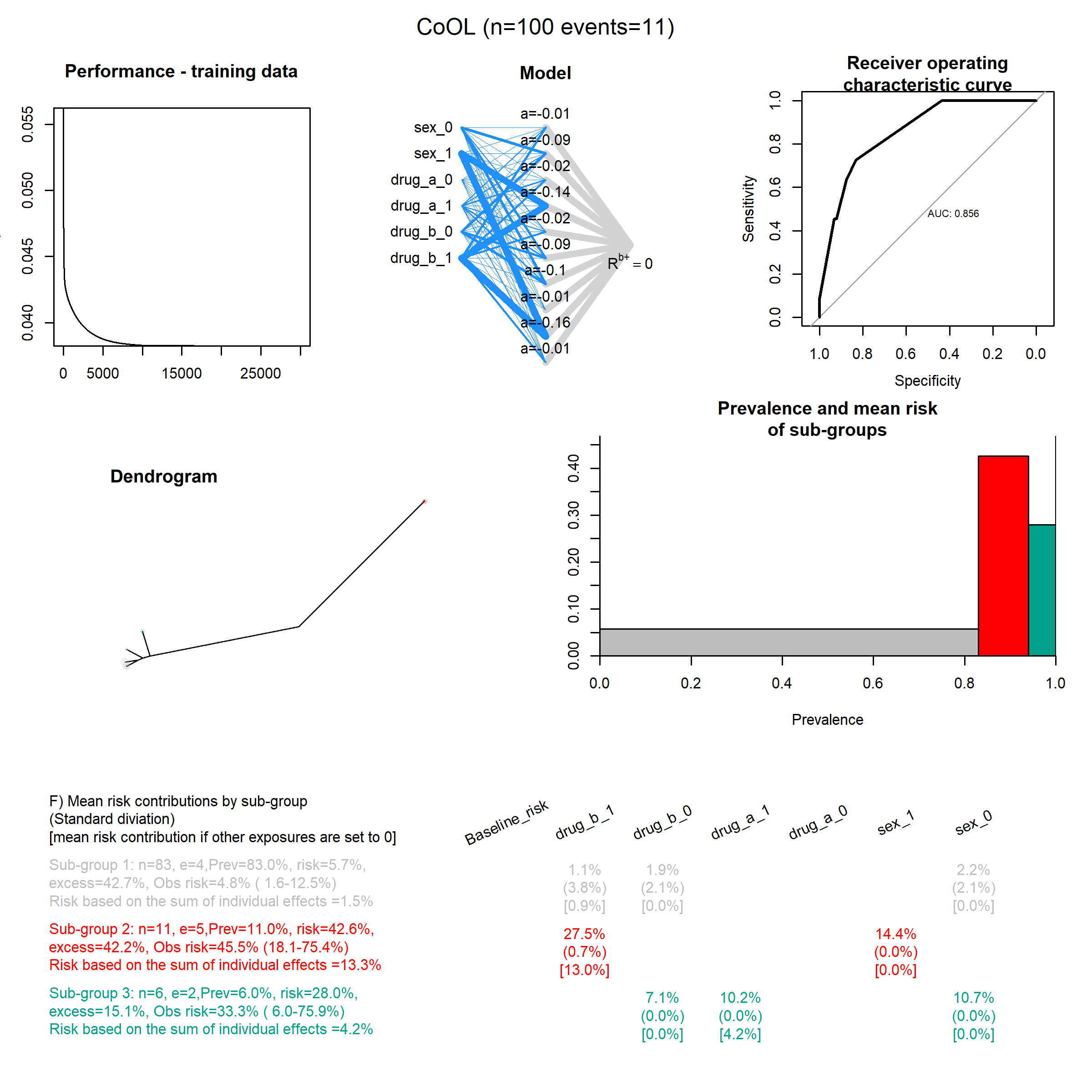

### Data_size_250.png

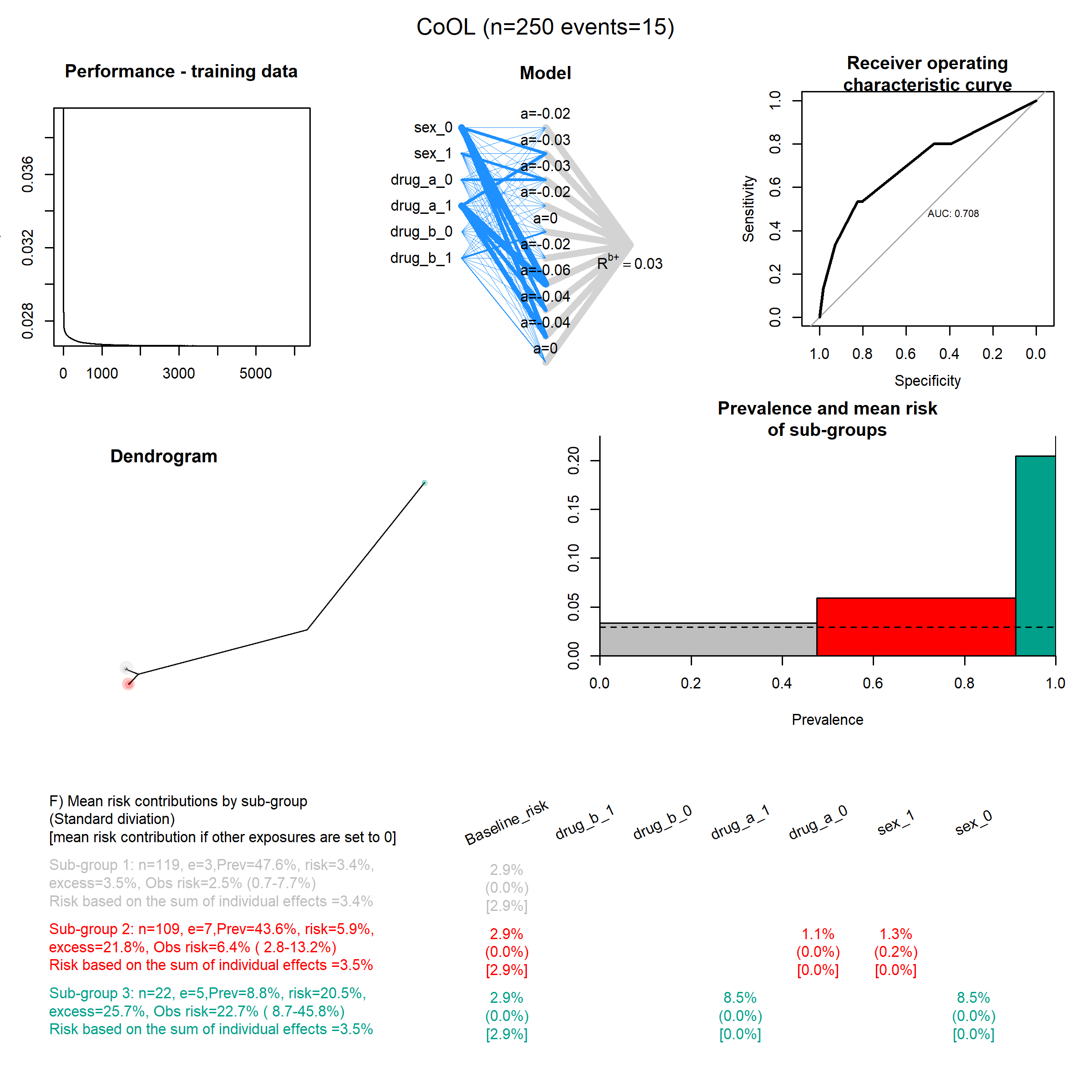

### Data_size_500.png

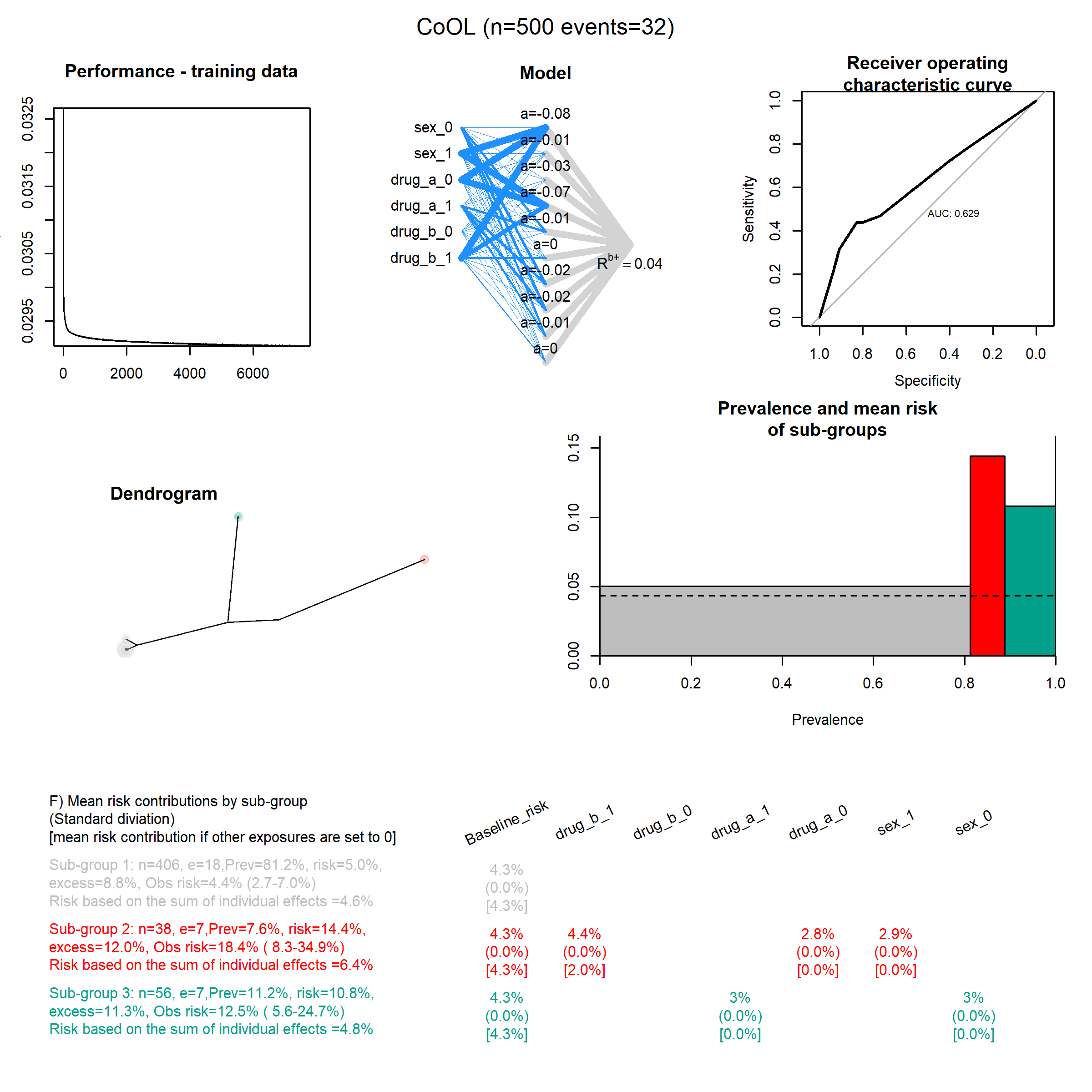

### Data_size_750.png

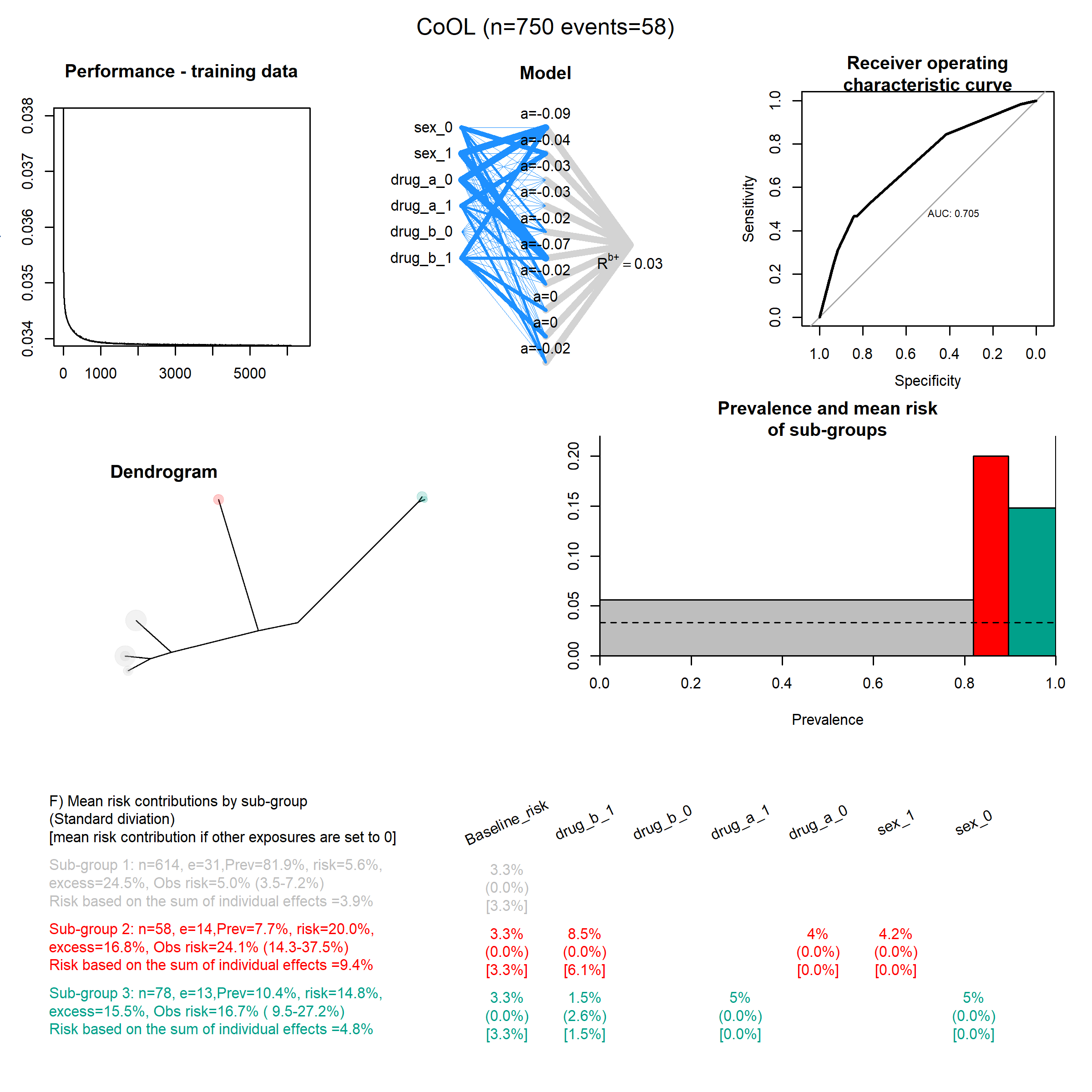

### Data_size_1000.png

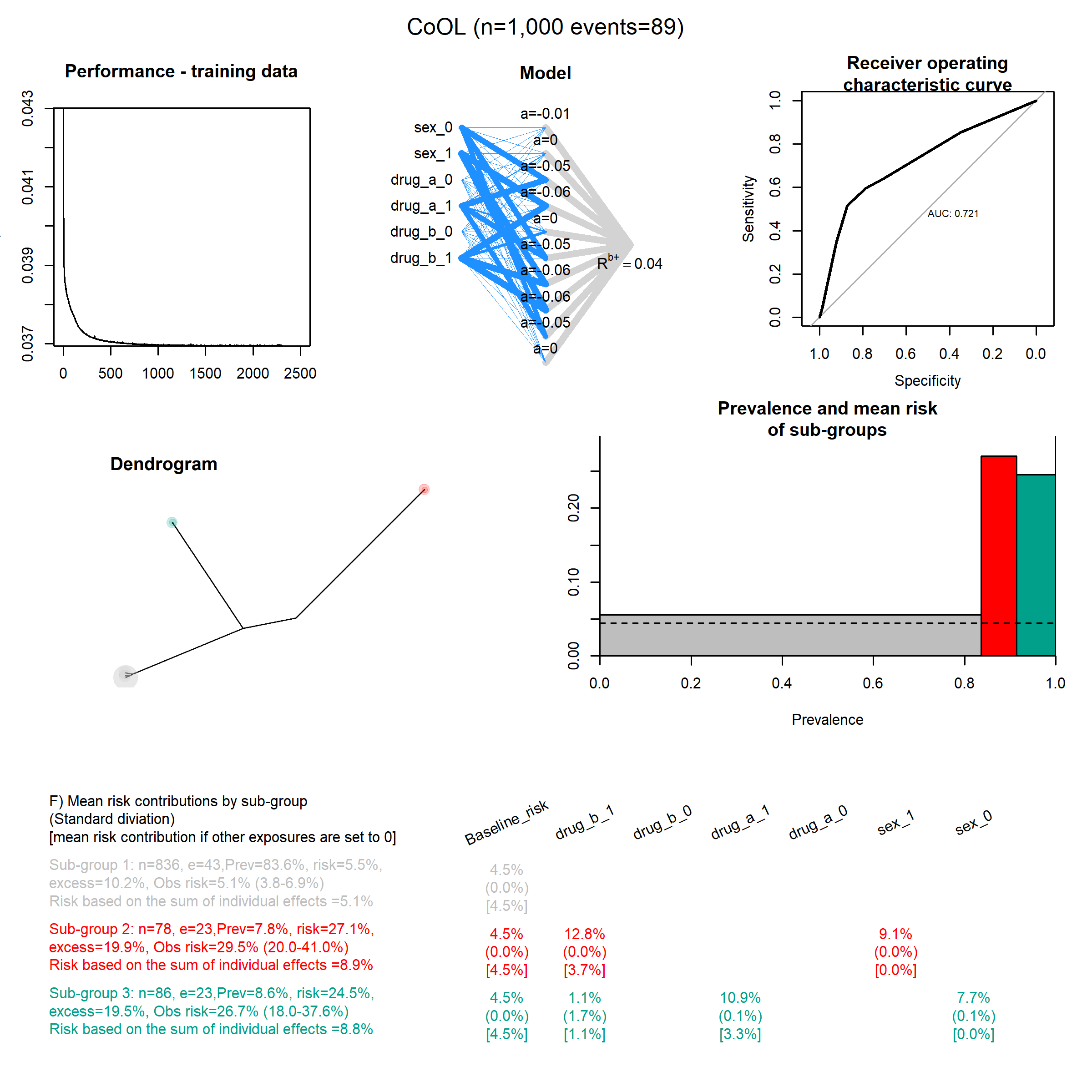

### Data_size_2000.png

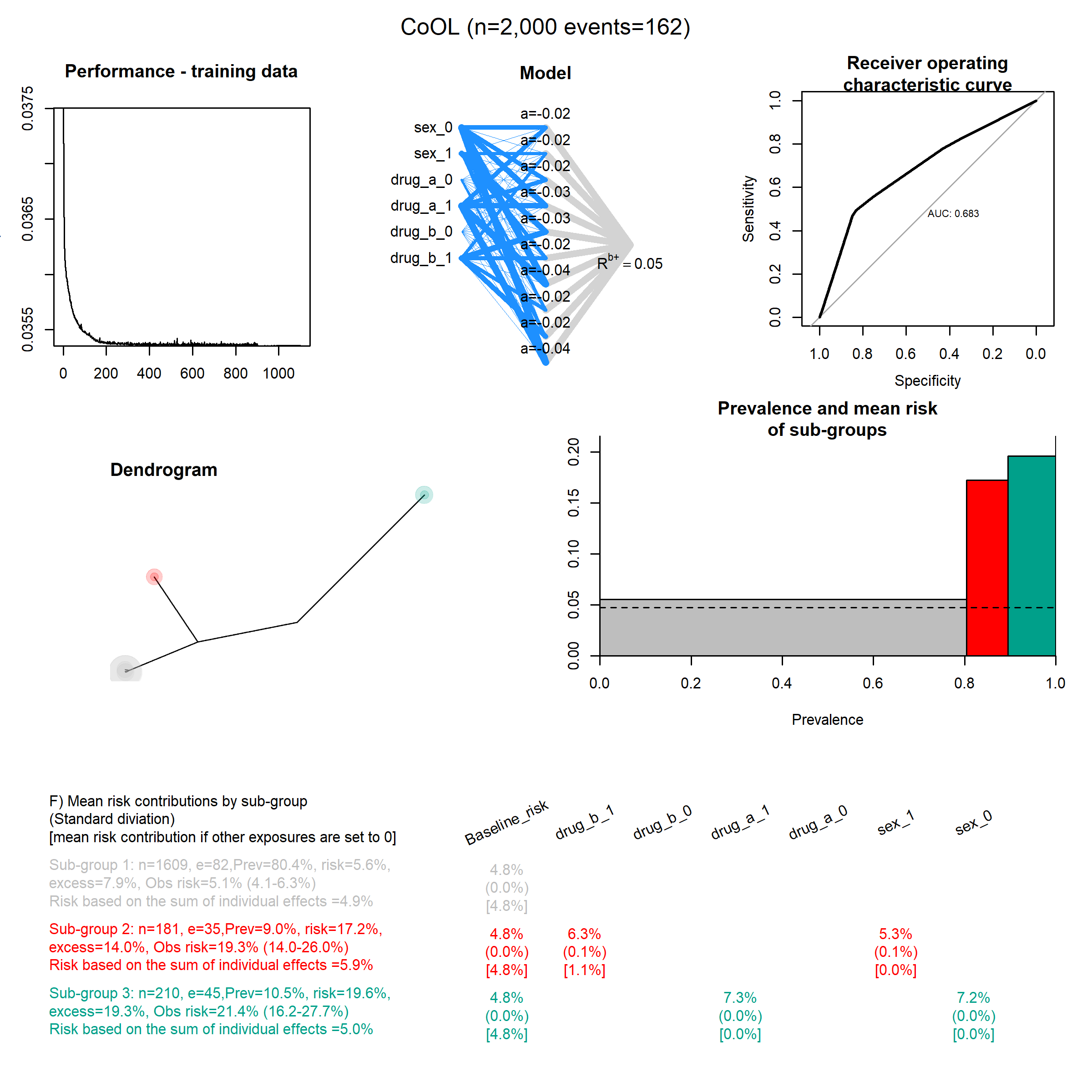

### Data_size_3000.png

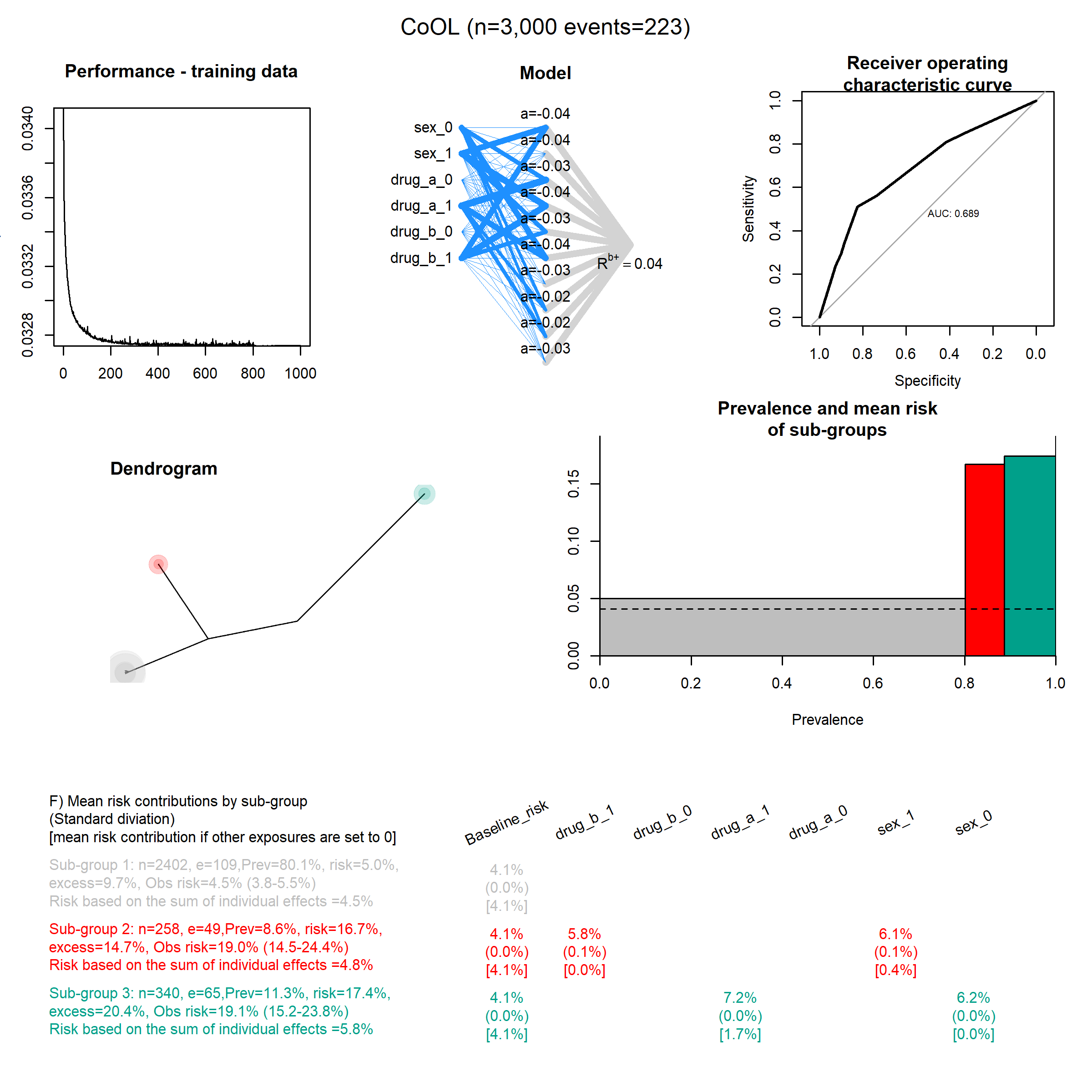

### Data_size_4000.png

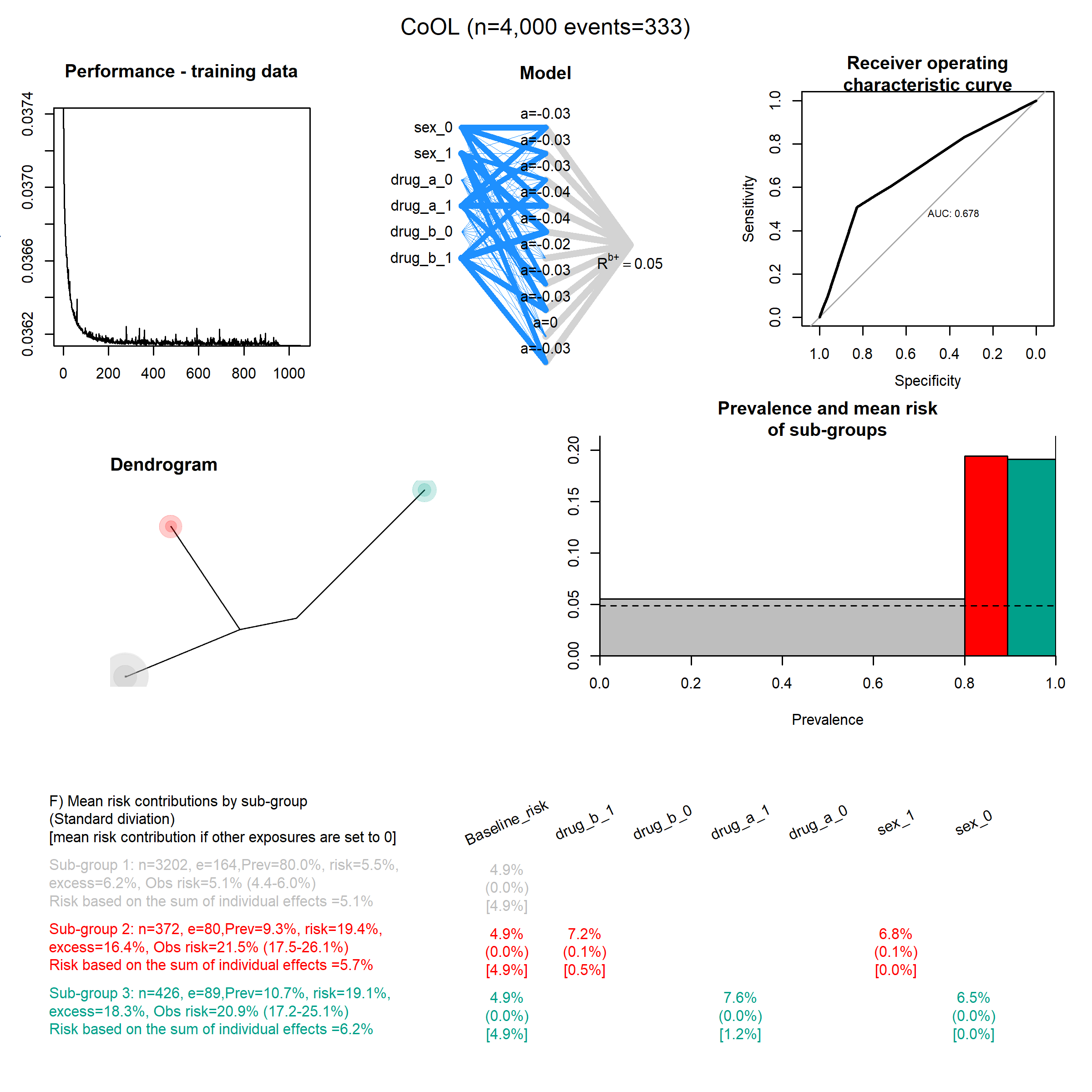

### Data_size_5000.png

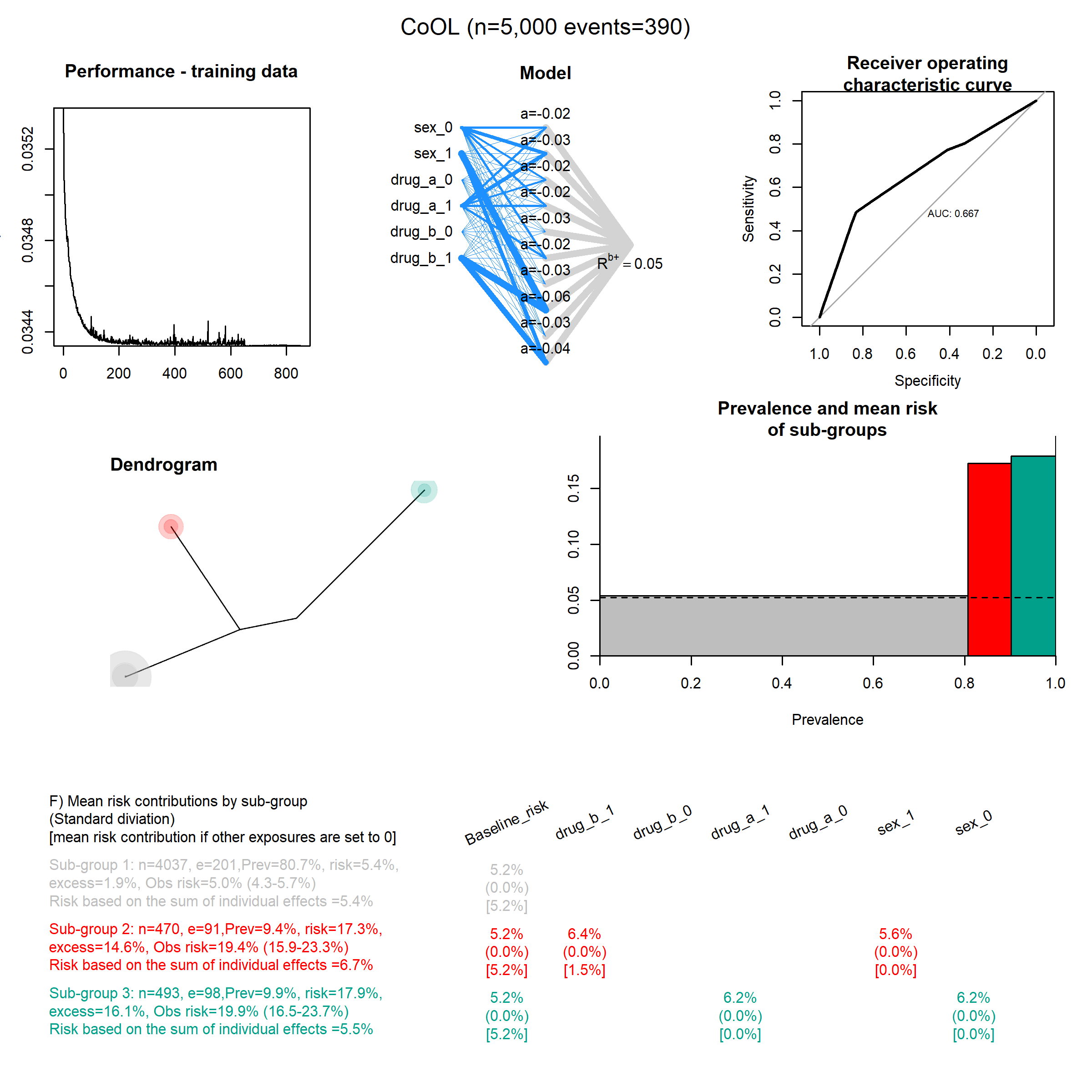

### Data_size_10000.png

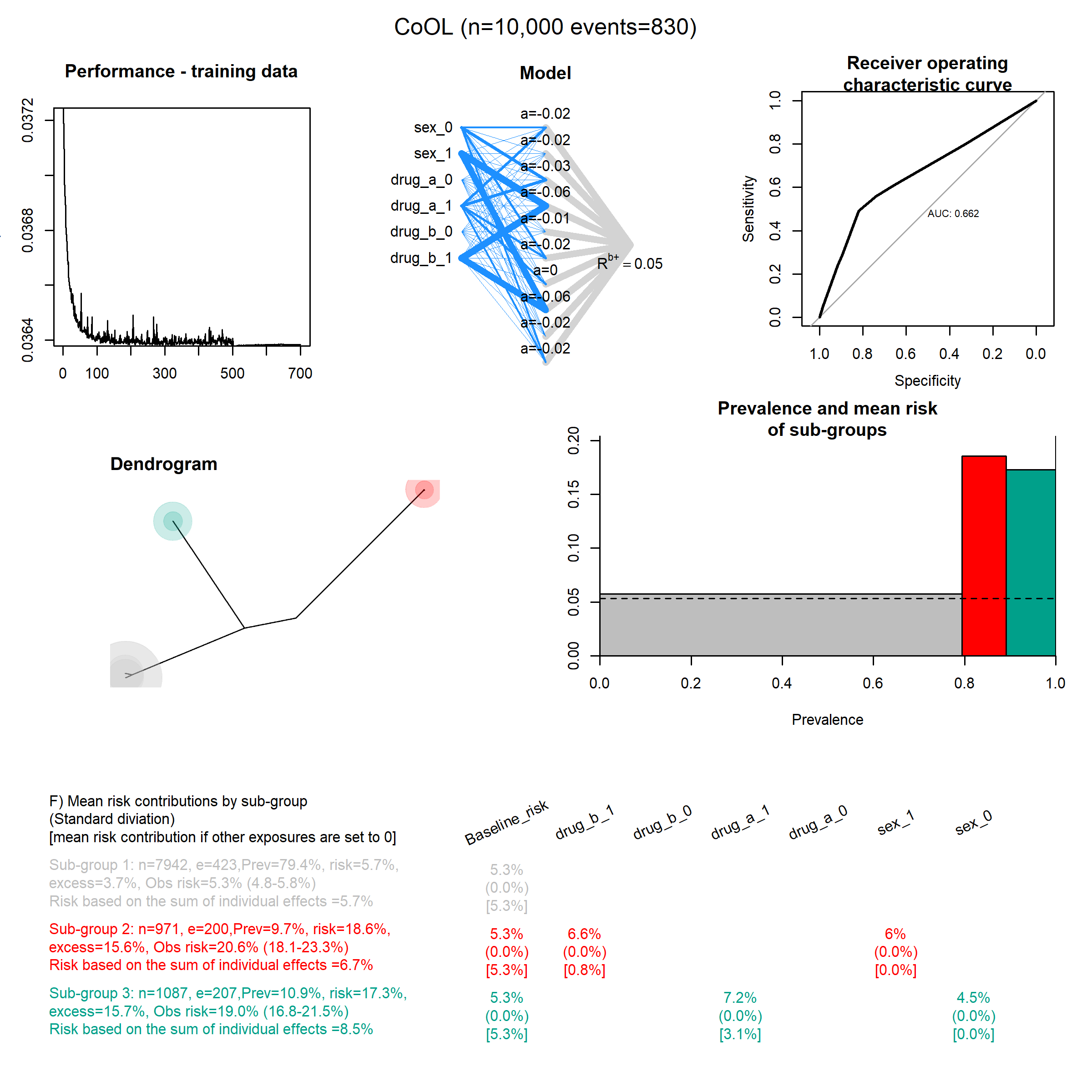

### Data_size_15000.png

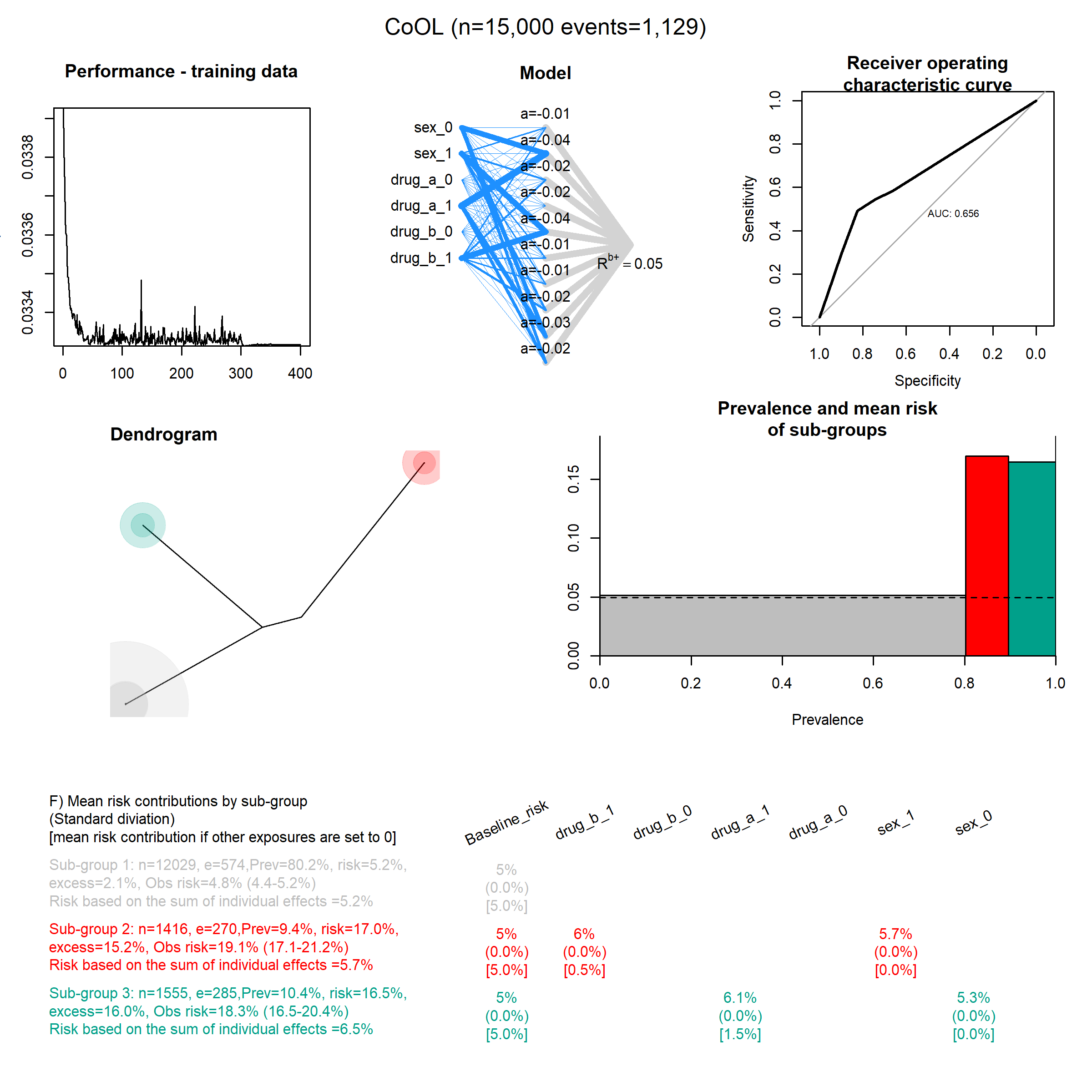

### Data_size_20000.png

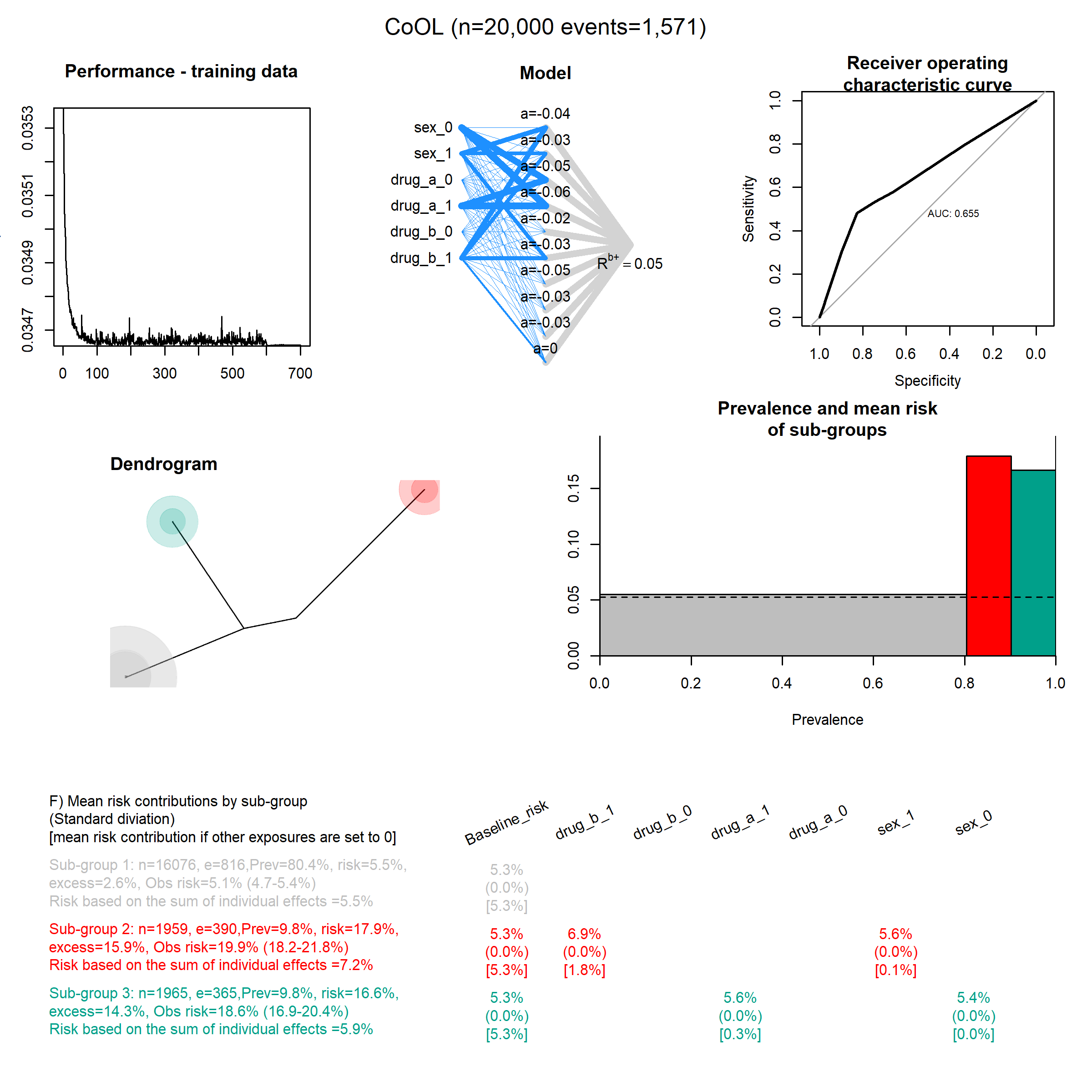

### Data_size_25000.png

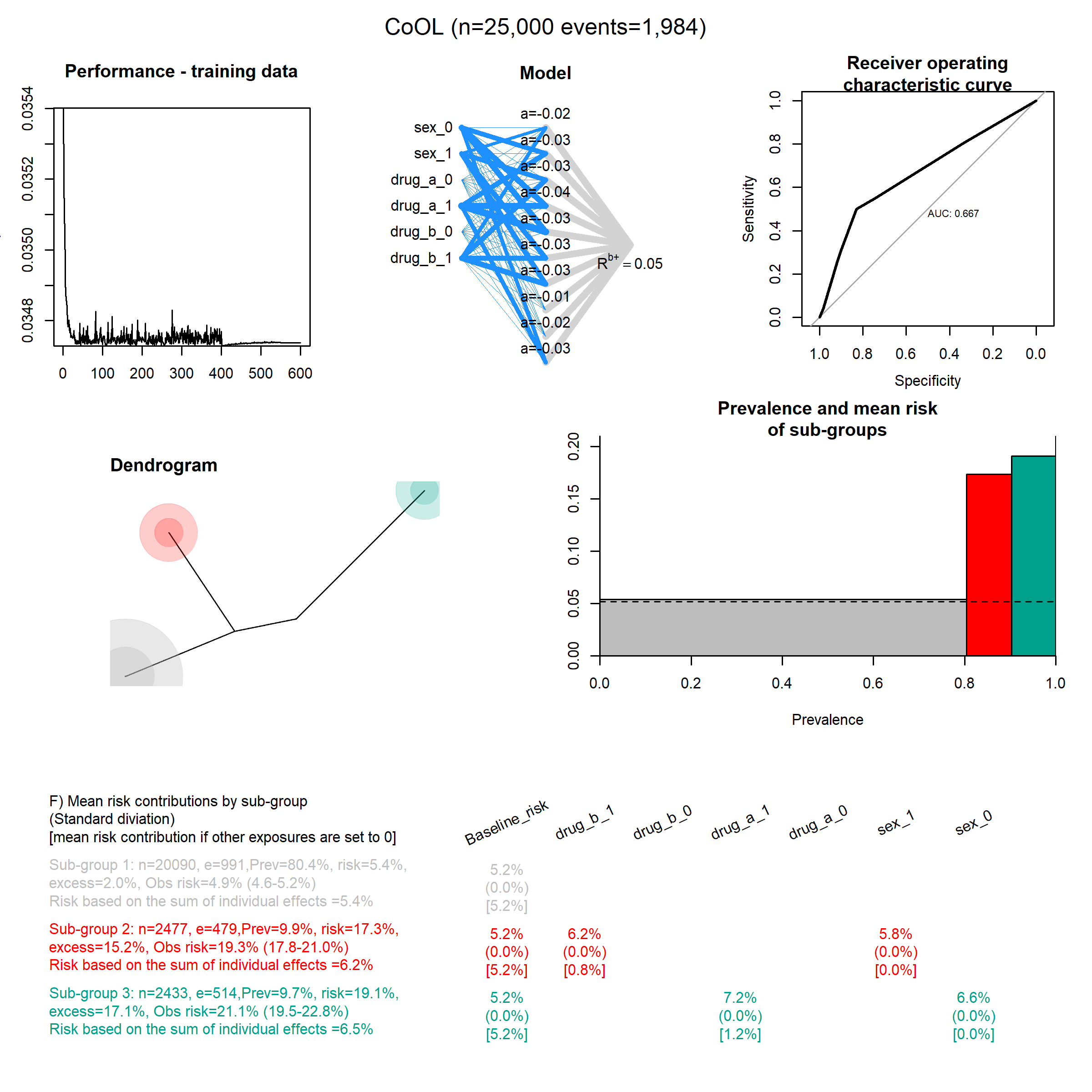

### Data_size_30000.png

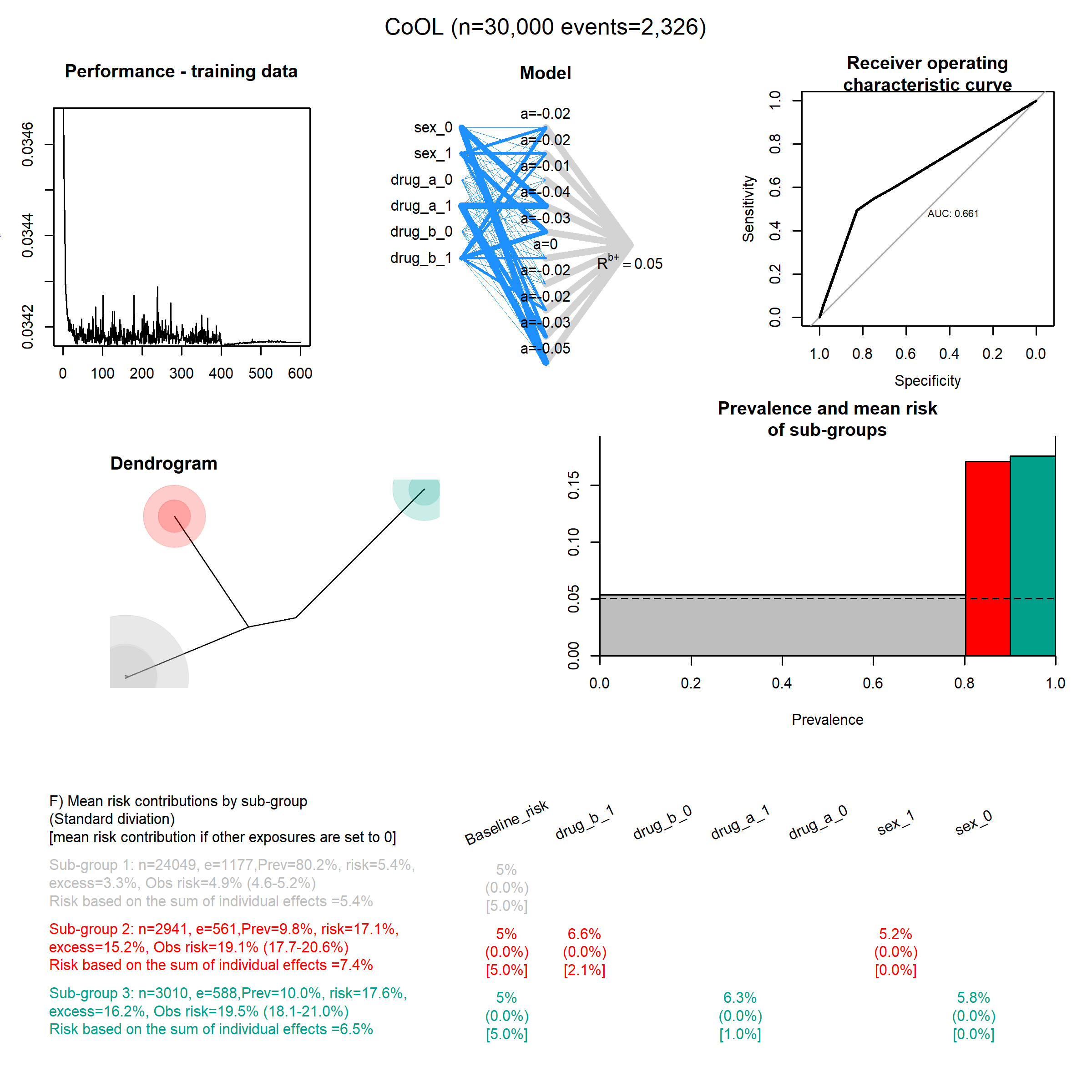

### Data_size_35000.png

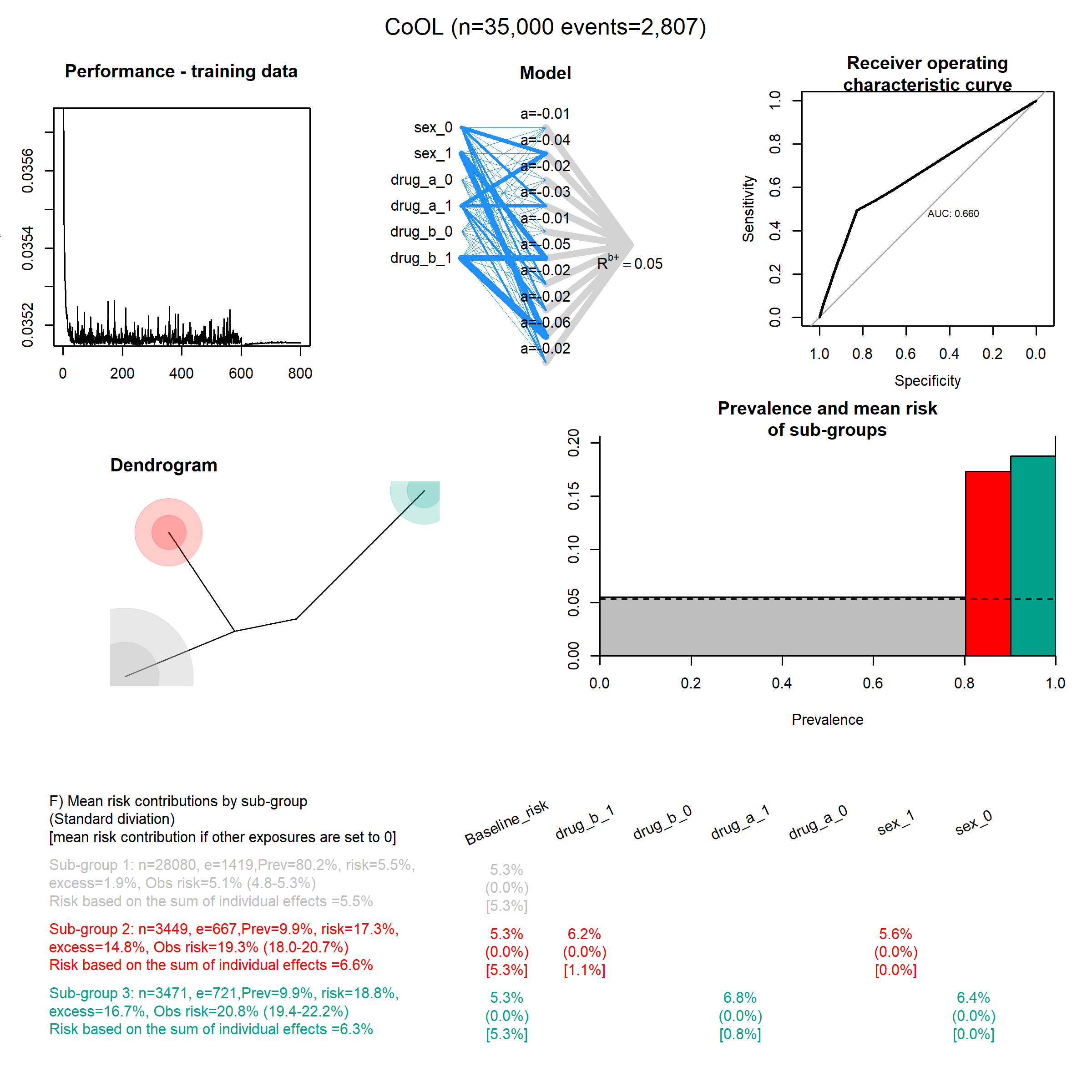

### Data_size_40000.png

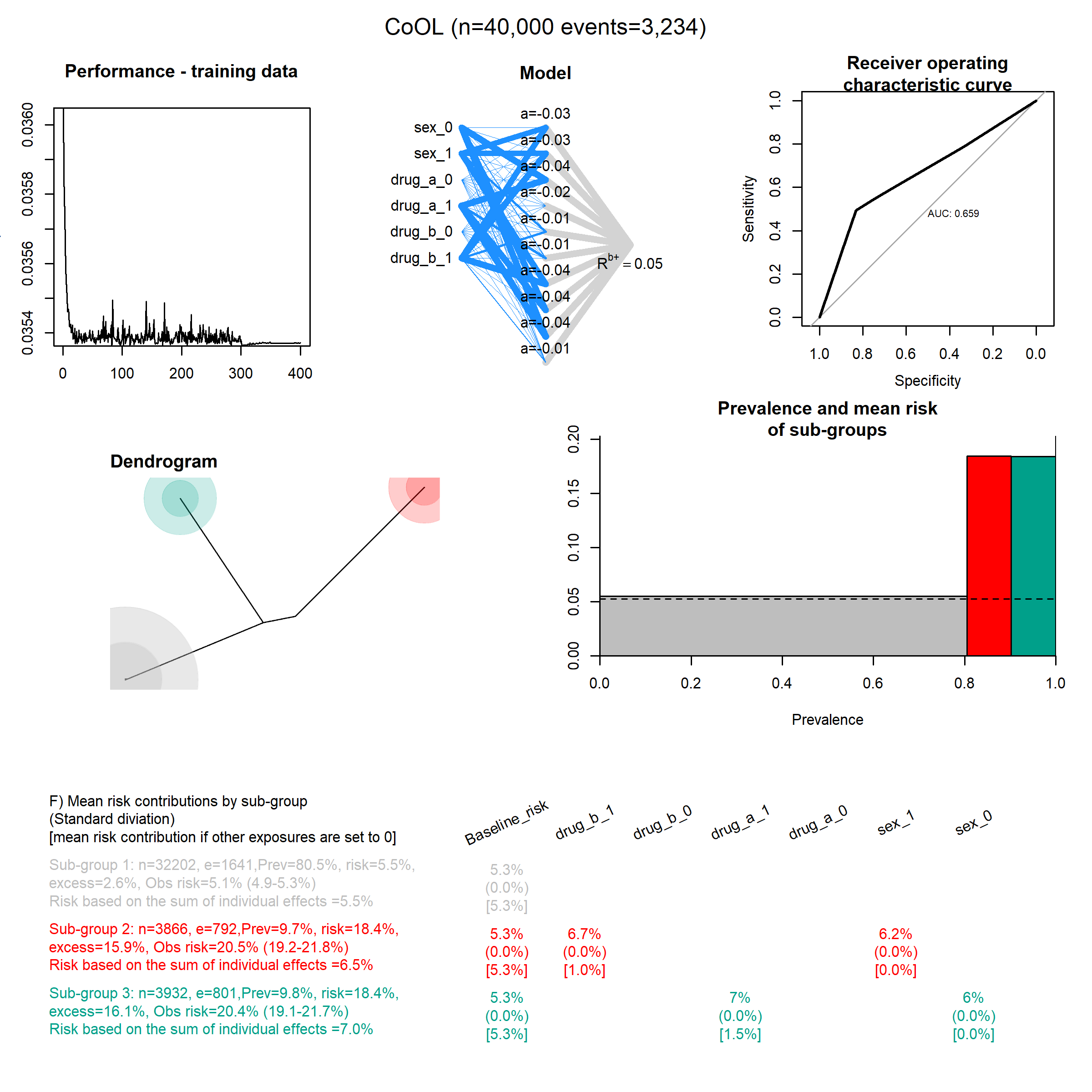

### Mediation_1.png

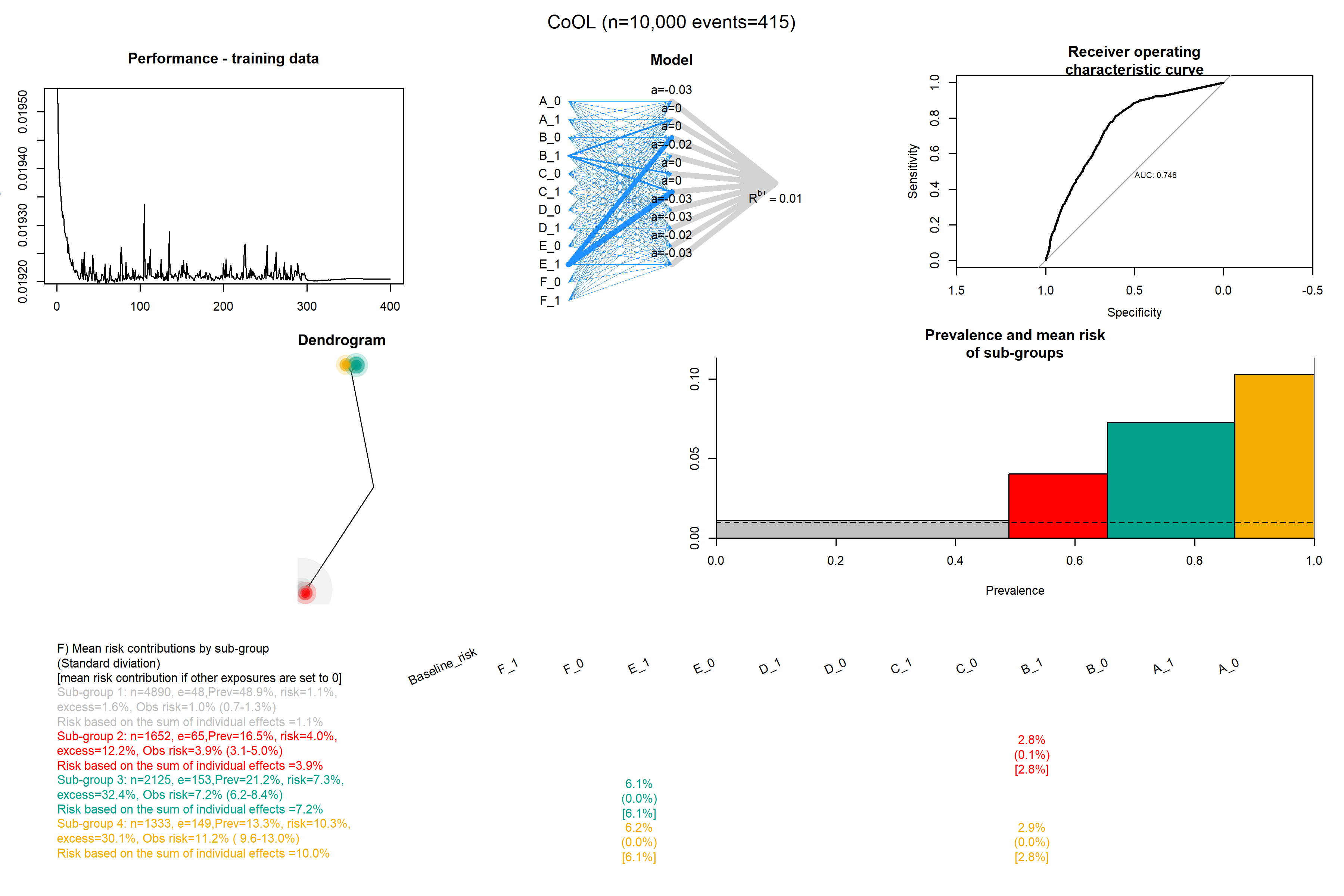

### Mediation_2.png

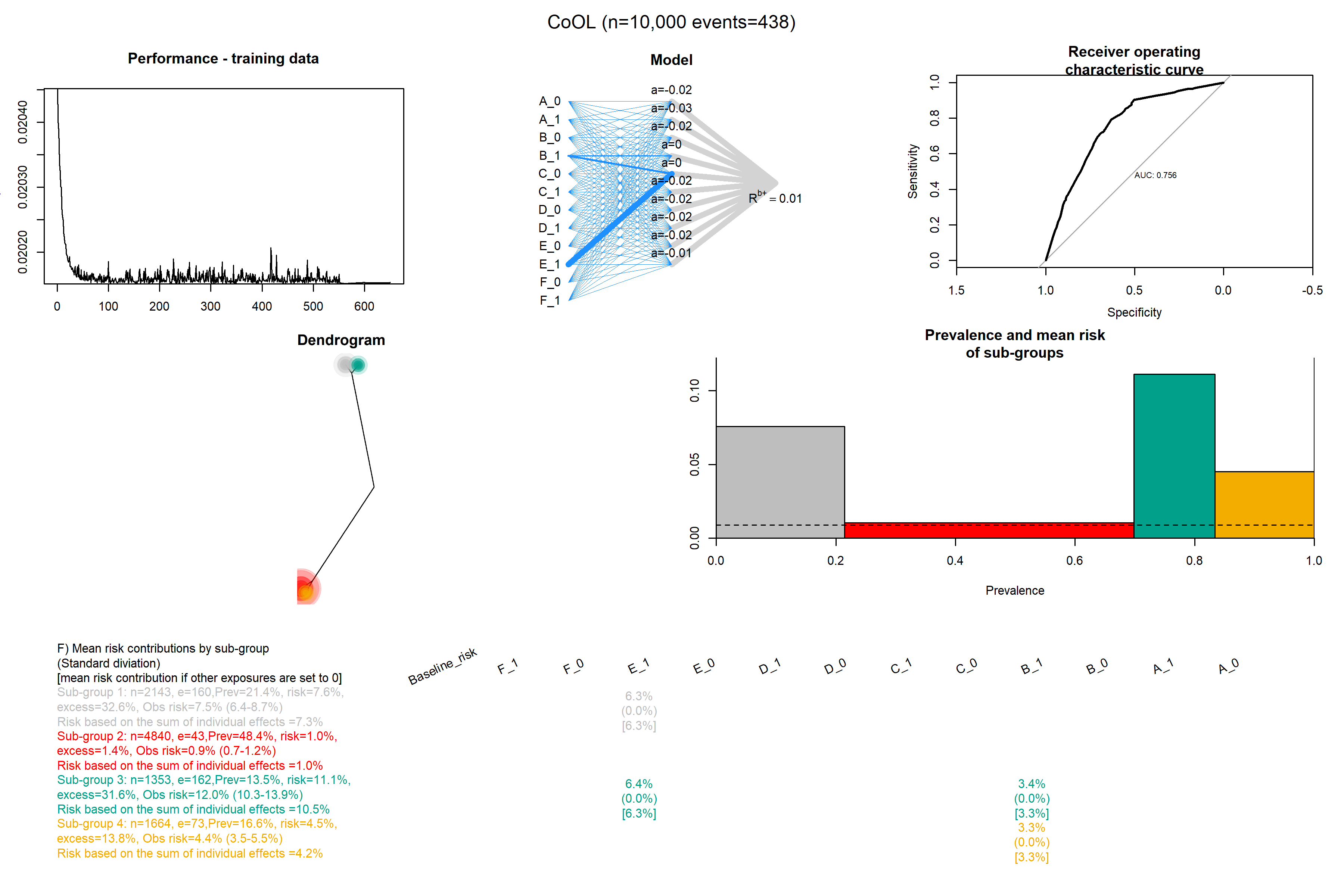

### Mediation_3.png

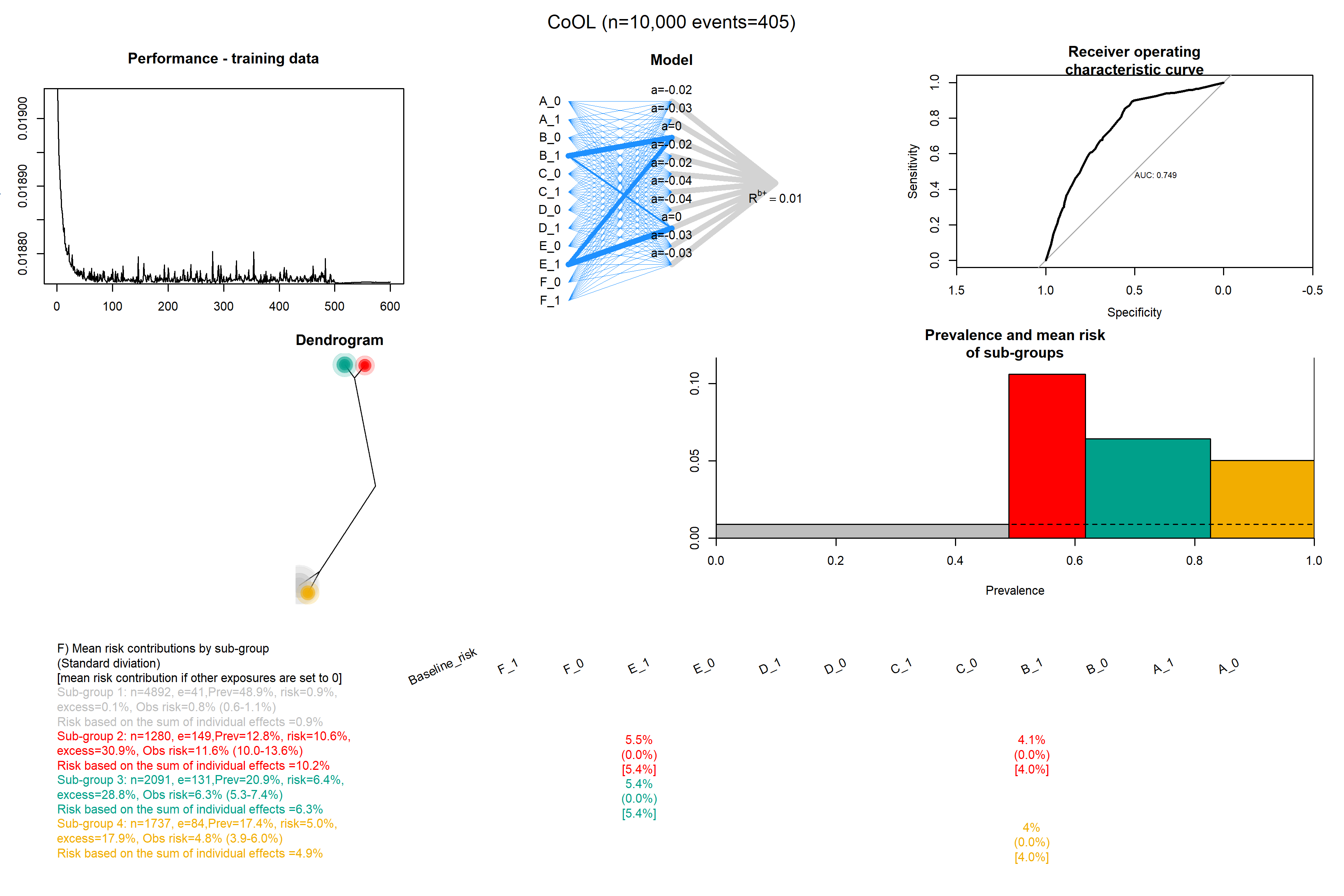

### Mediation_4.png

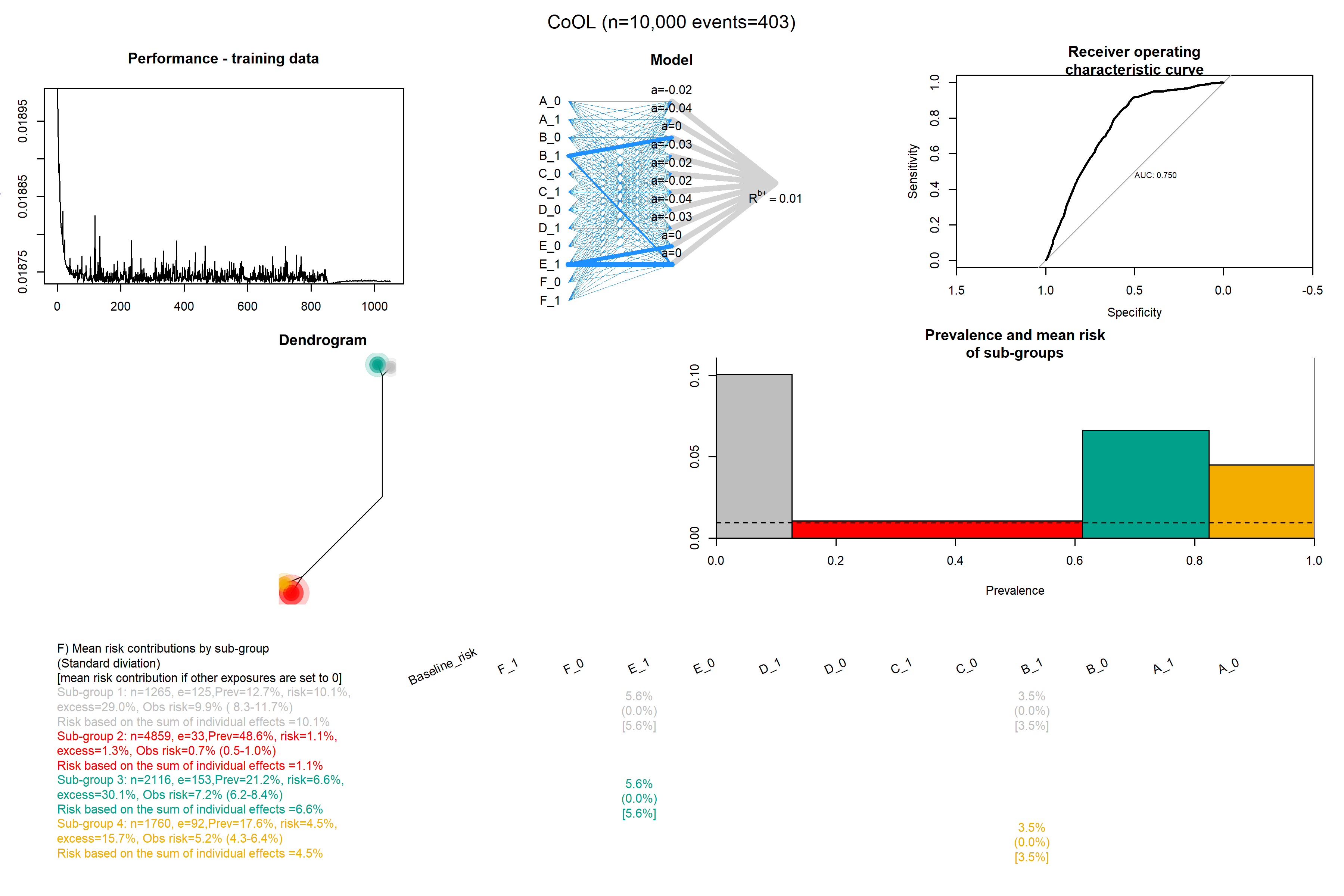

### Mediation_5.png

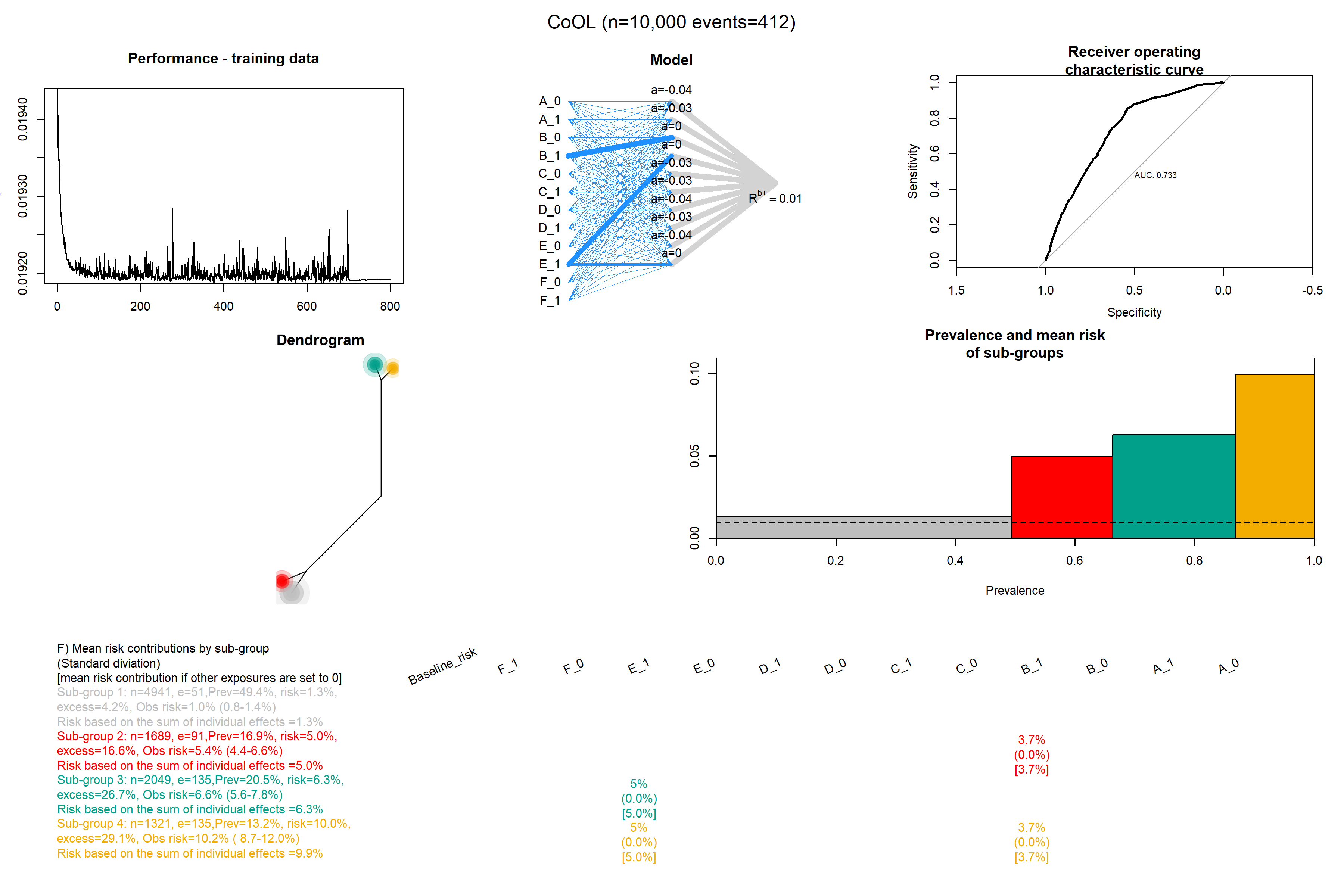

### Mediation_6.png

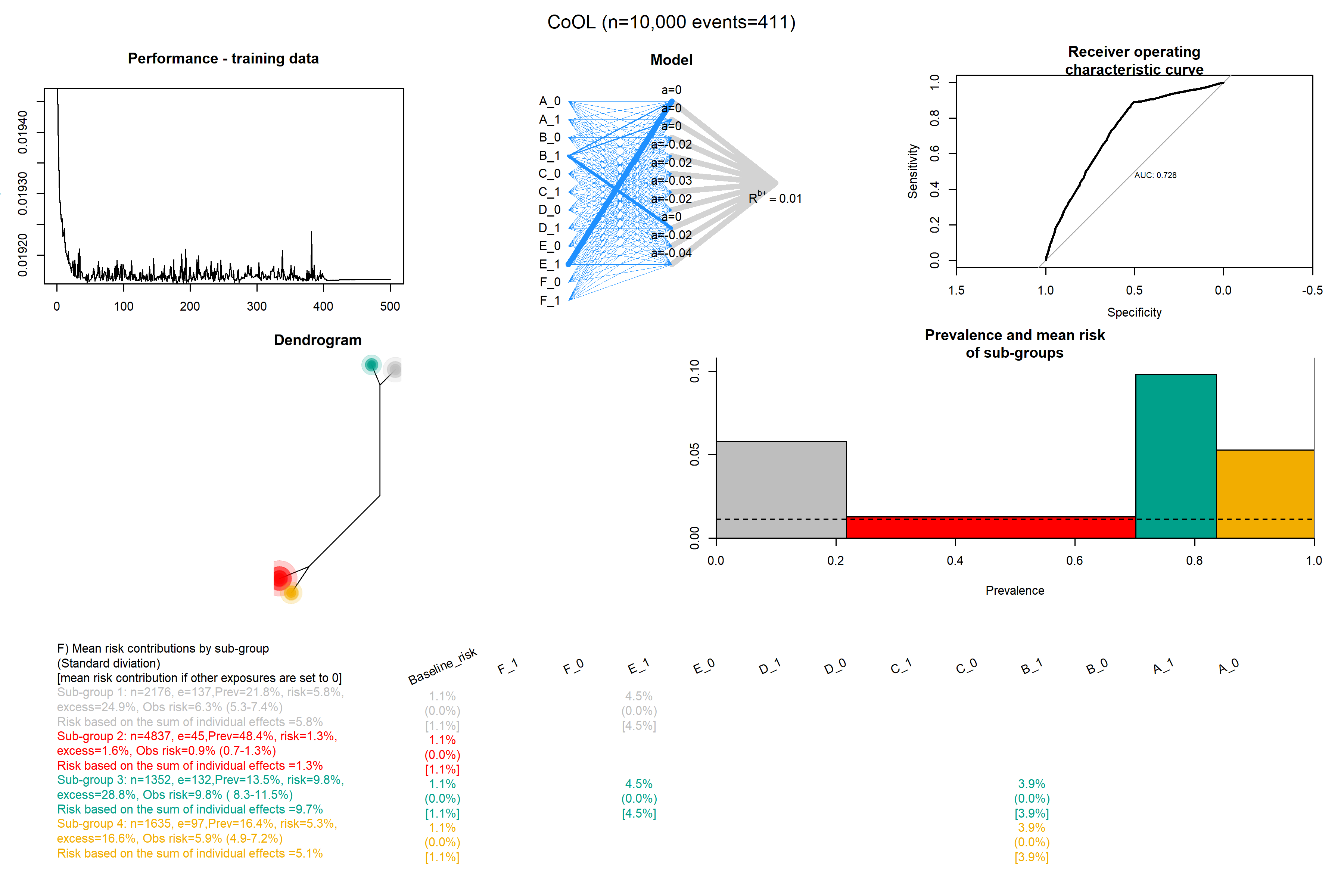

### Mediation_7.png

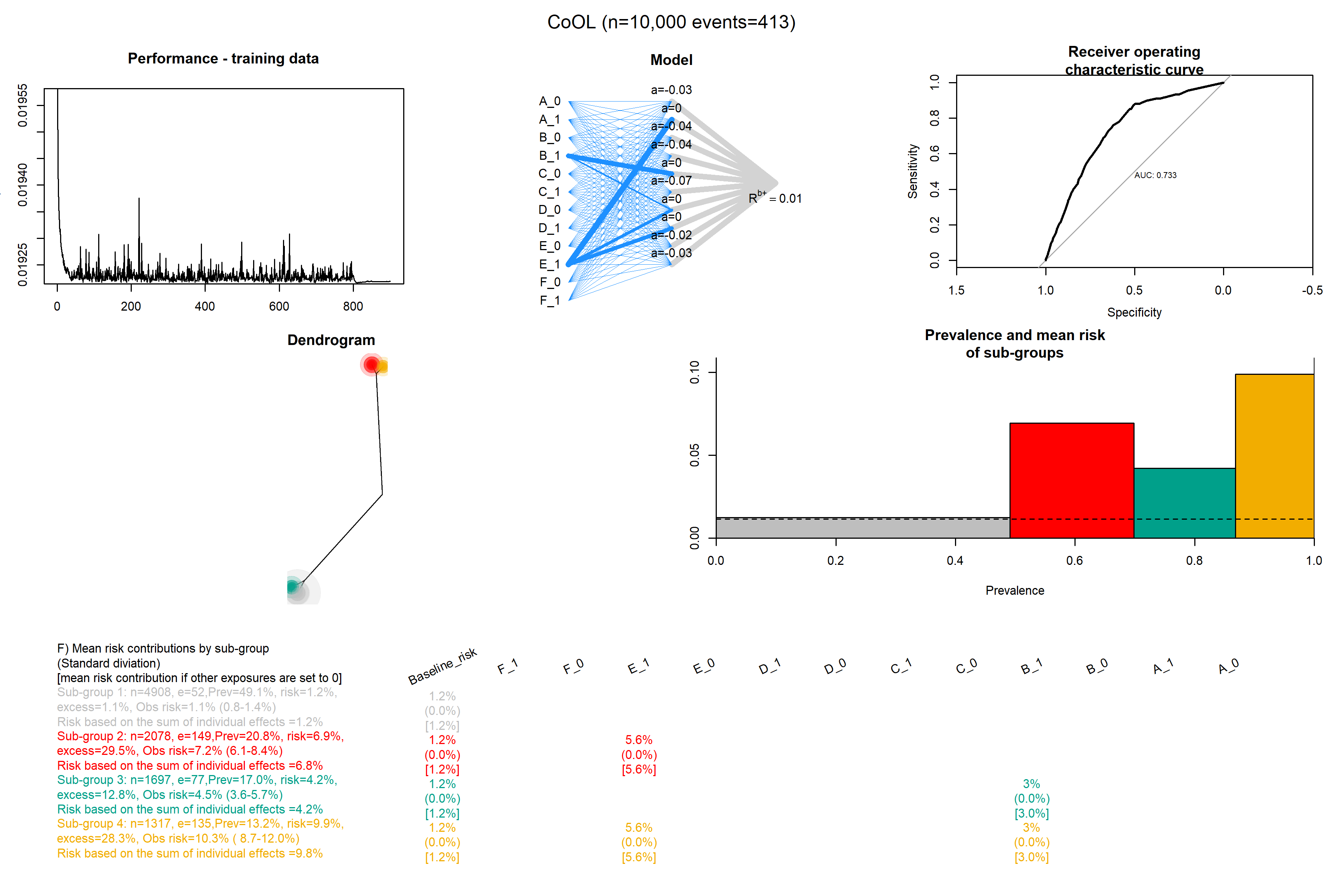

### Mediation_8.png

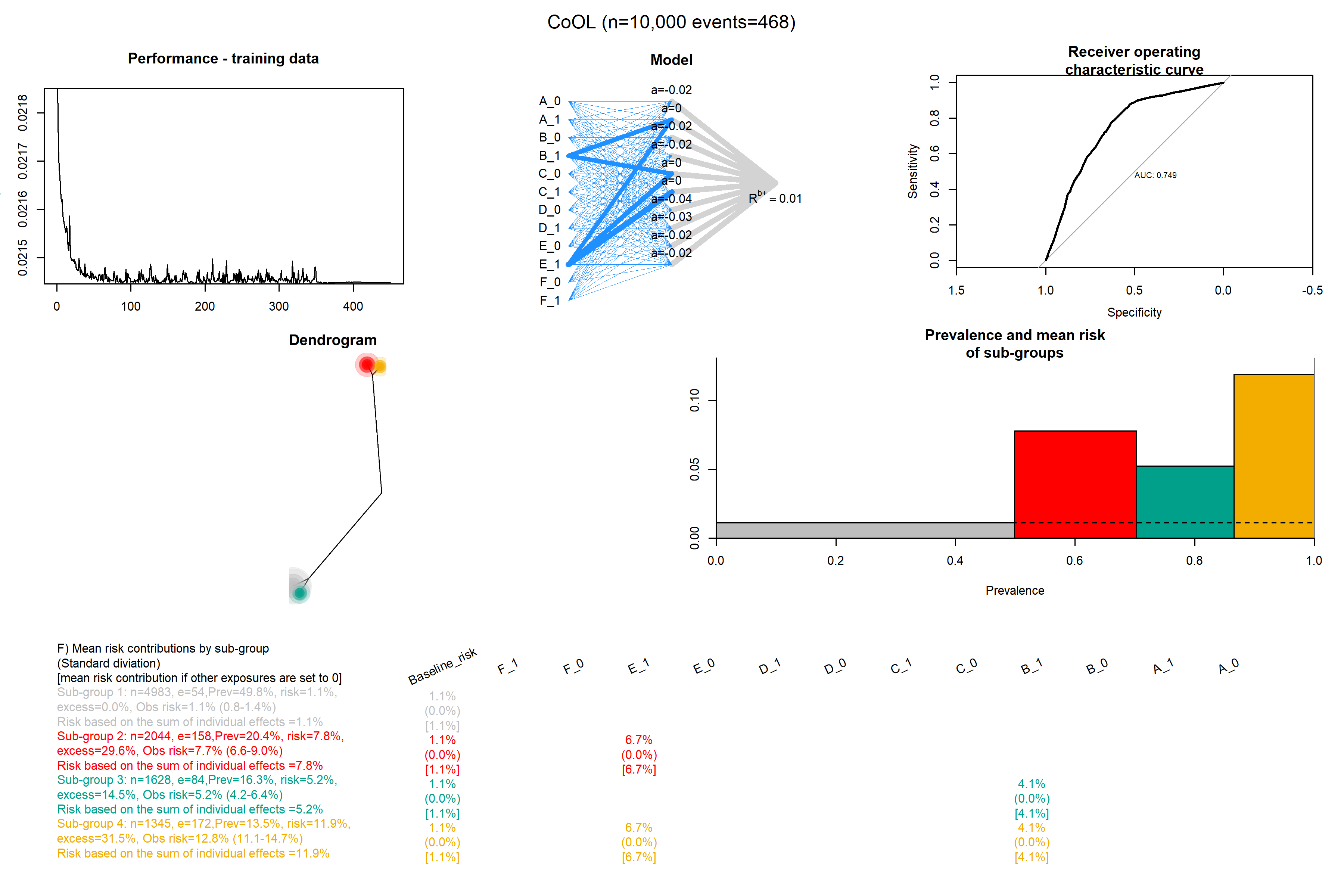

### Mediation_9.png

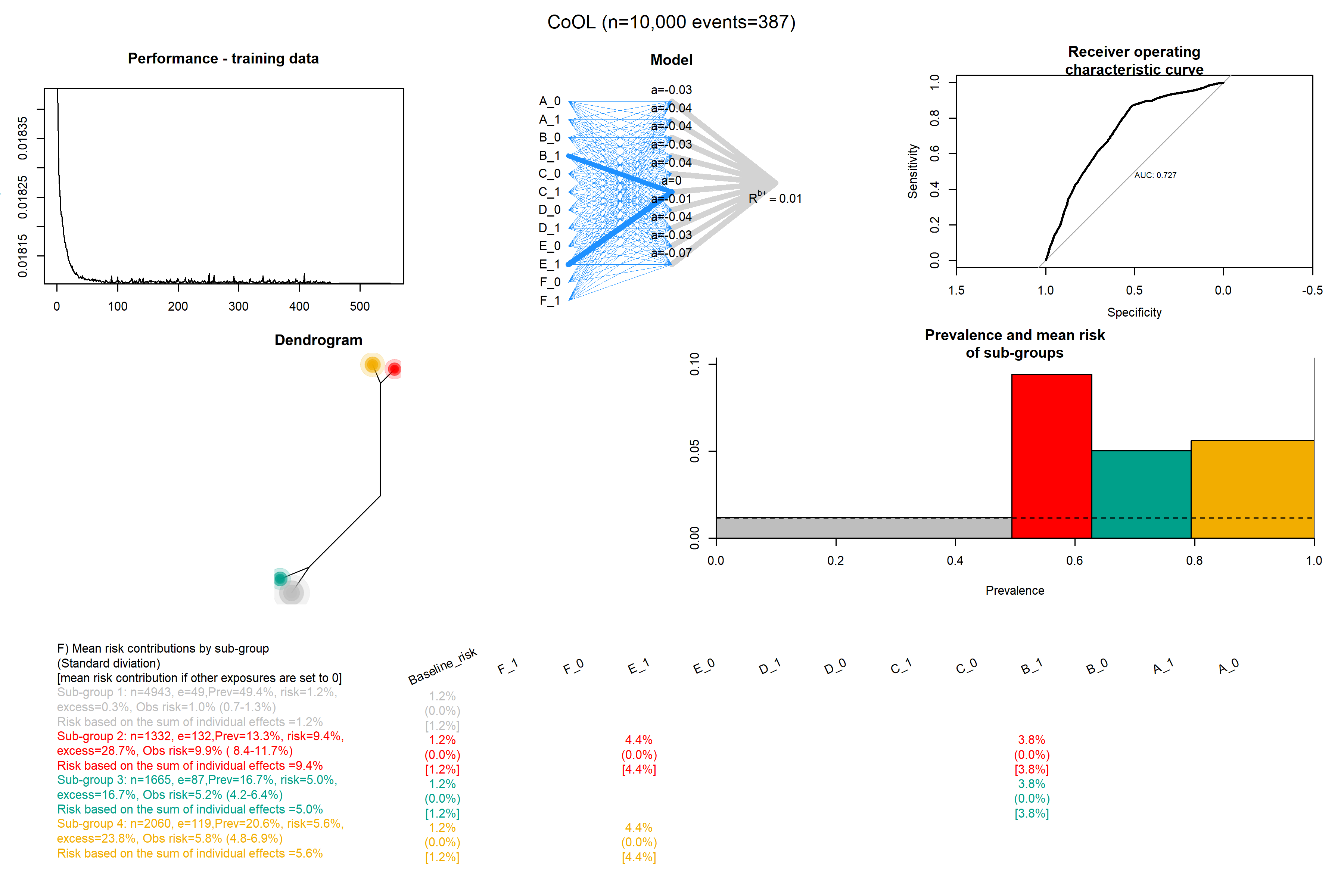

### Mediation_10.png

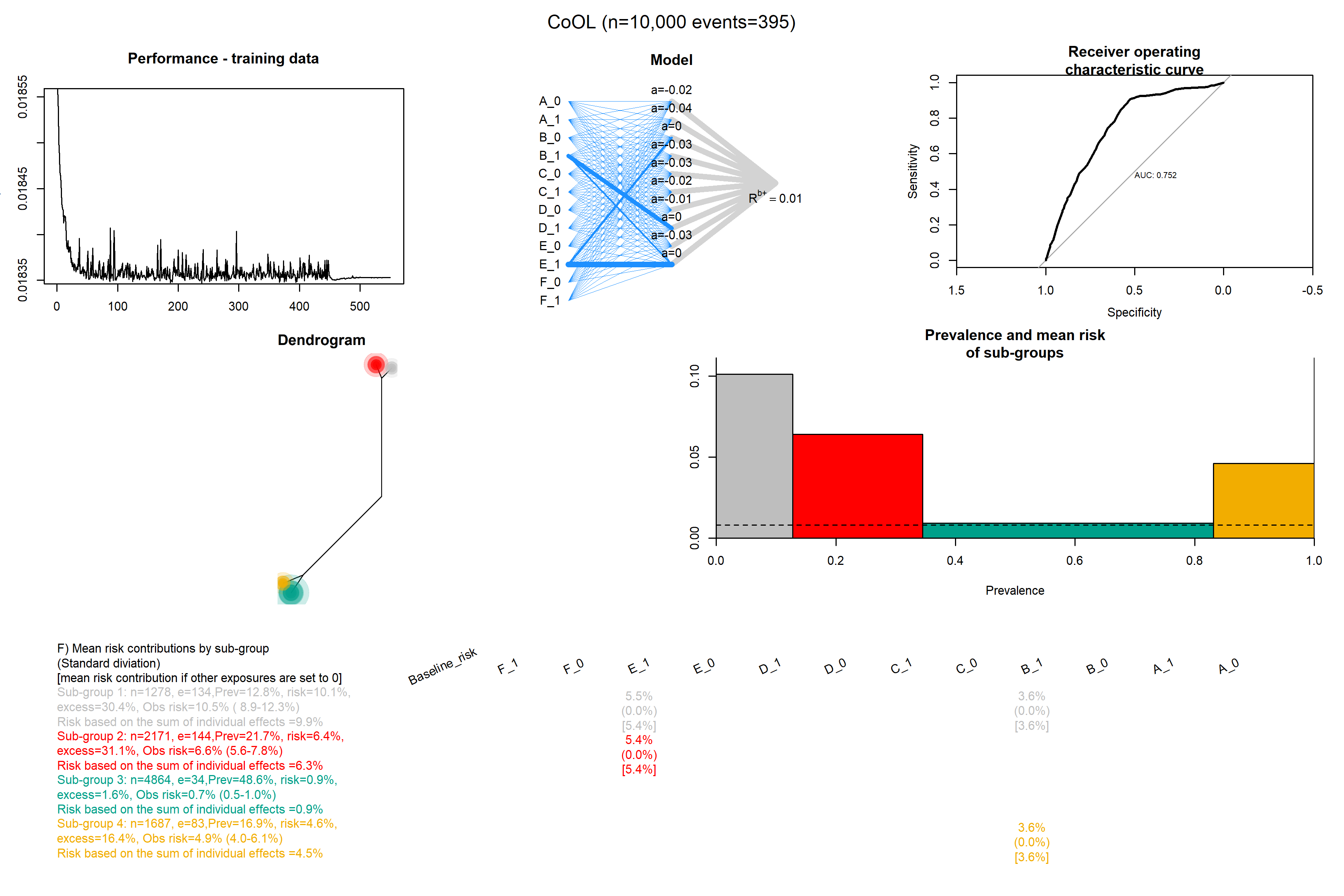

### Regularization_2.png

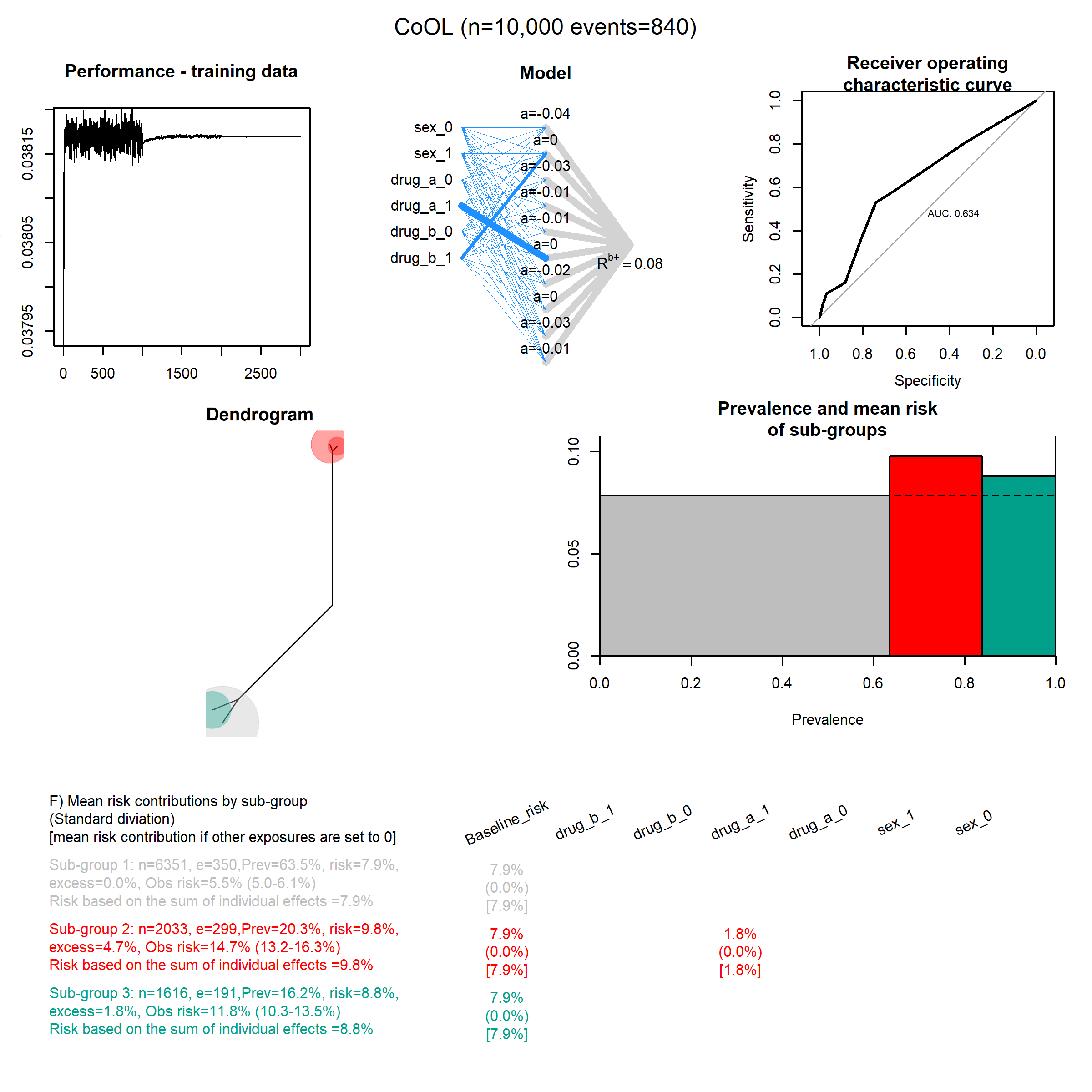

### Regularization_3.png

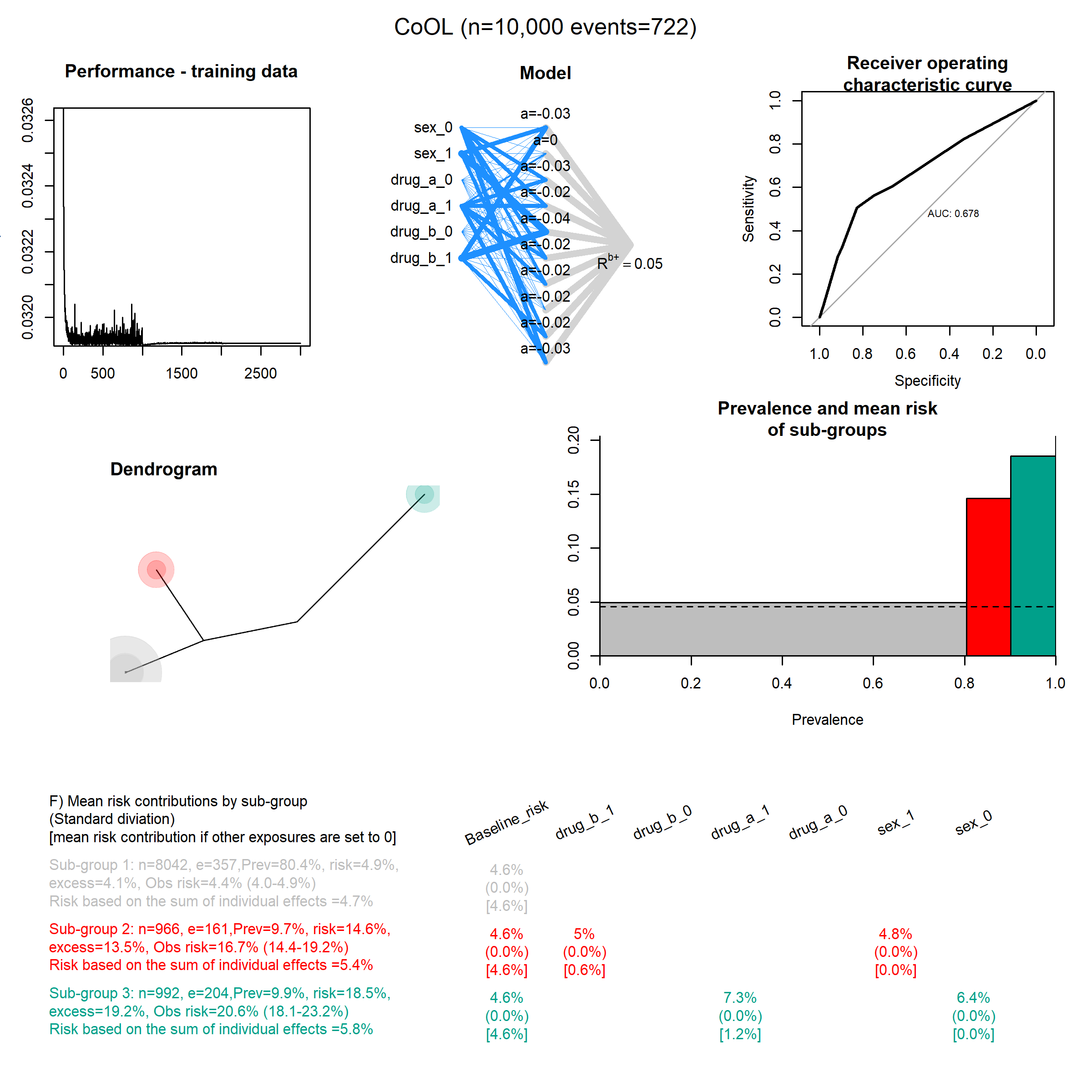

### Regularization_4.png

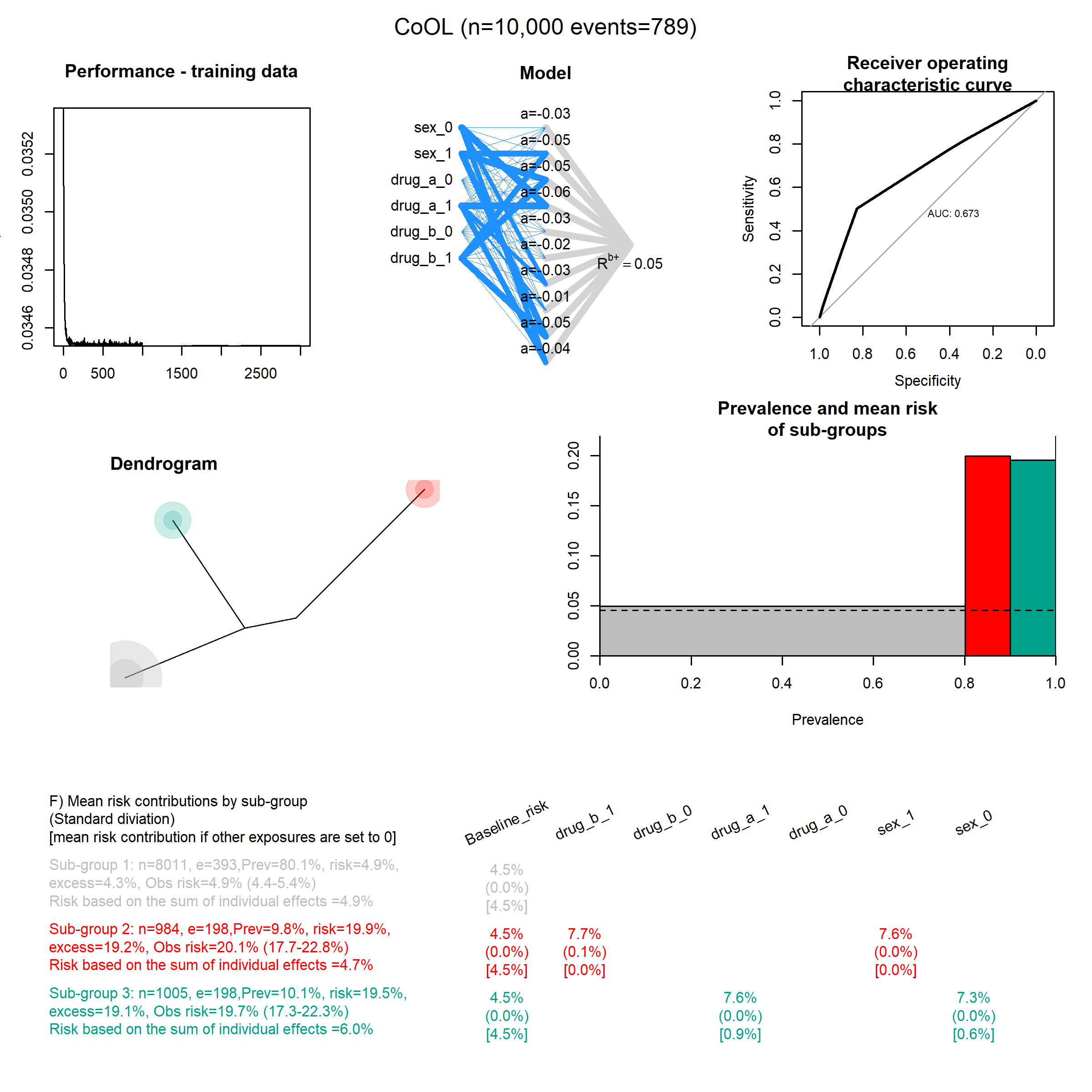

### Regularization_5.png

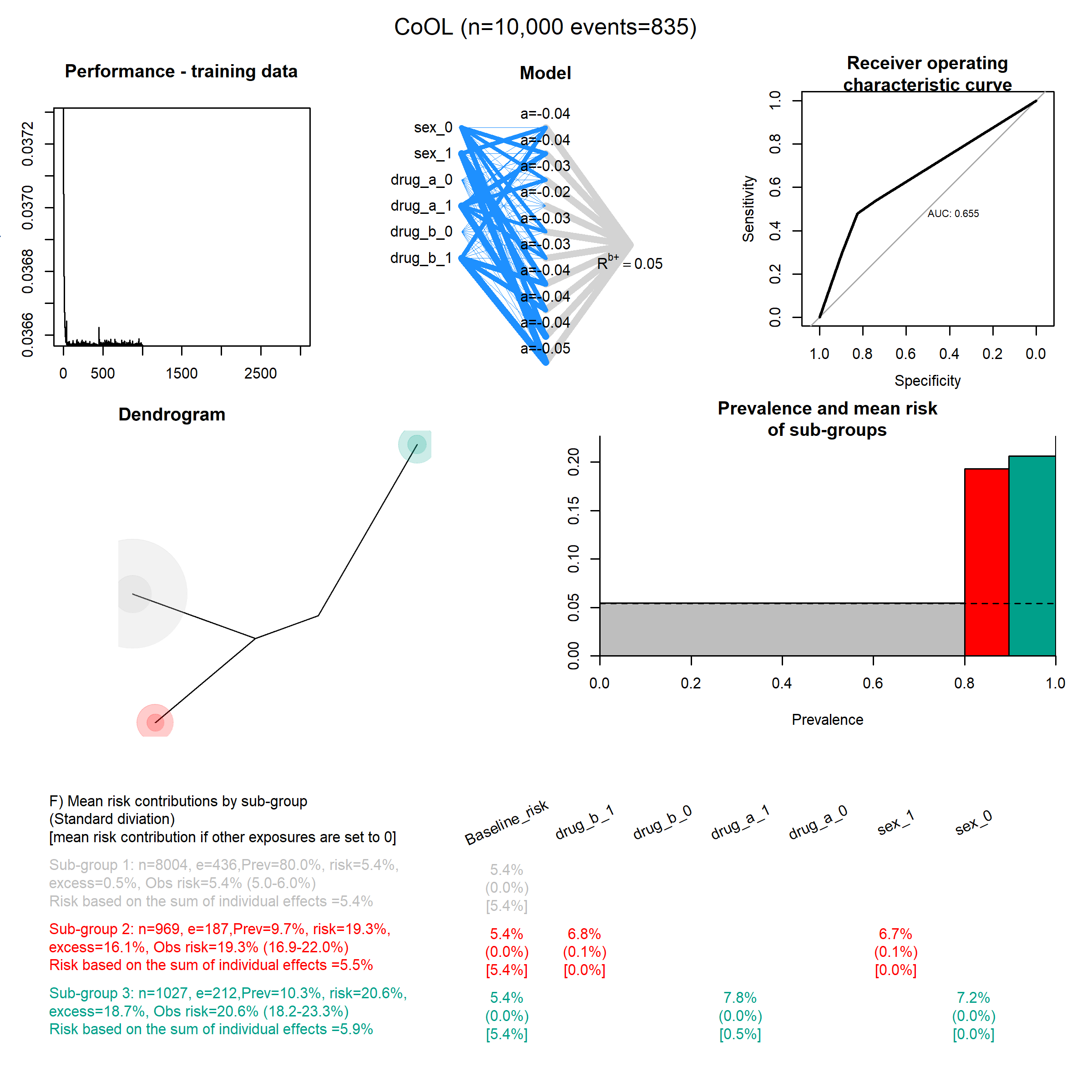

### Regularization_10.png

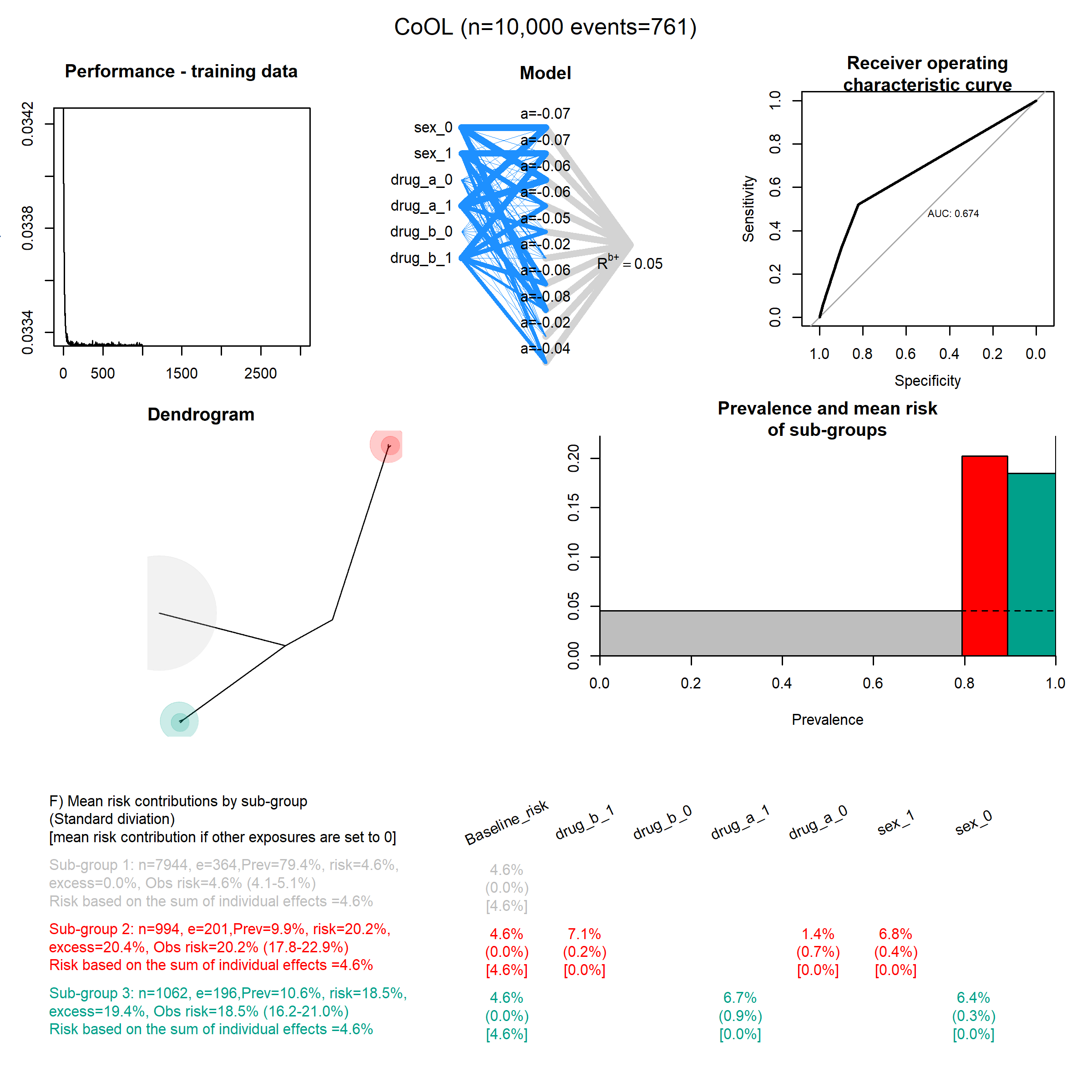

### Repeated_1.png

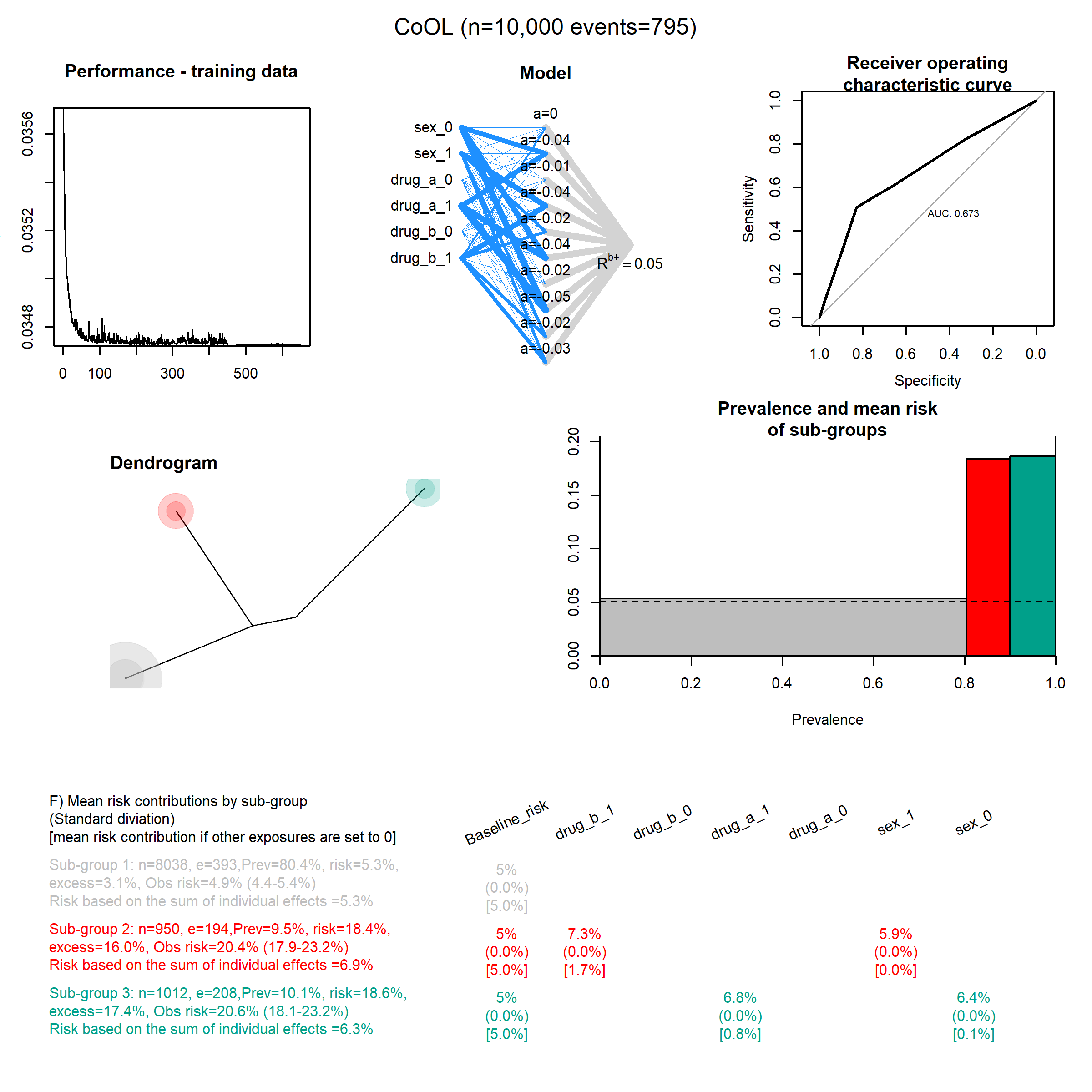

### Repeated_2.png

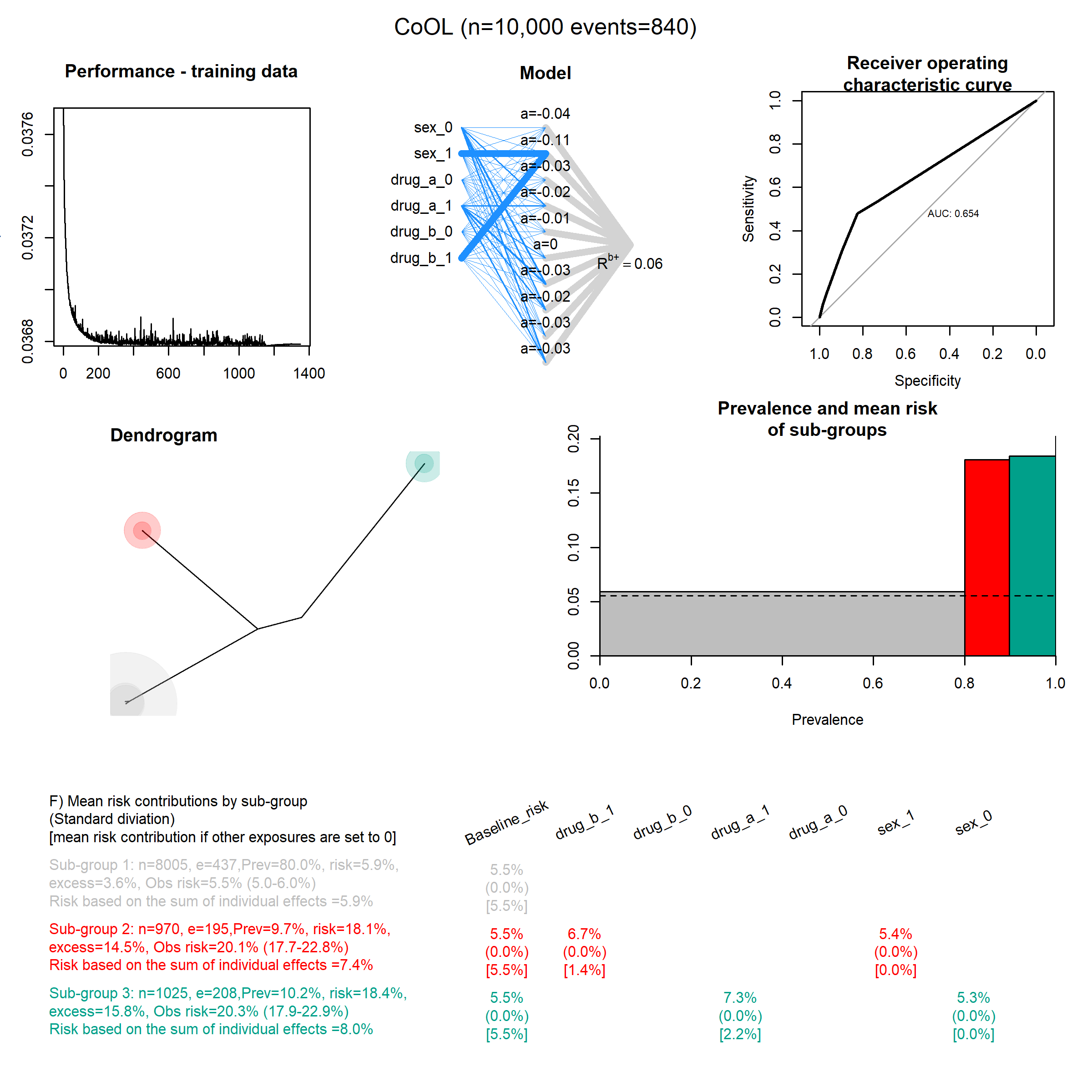

### Repeated_3.png

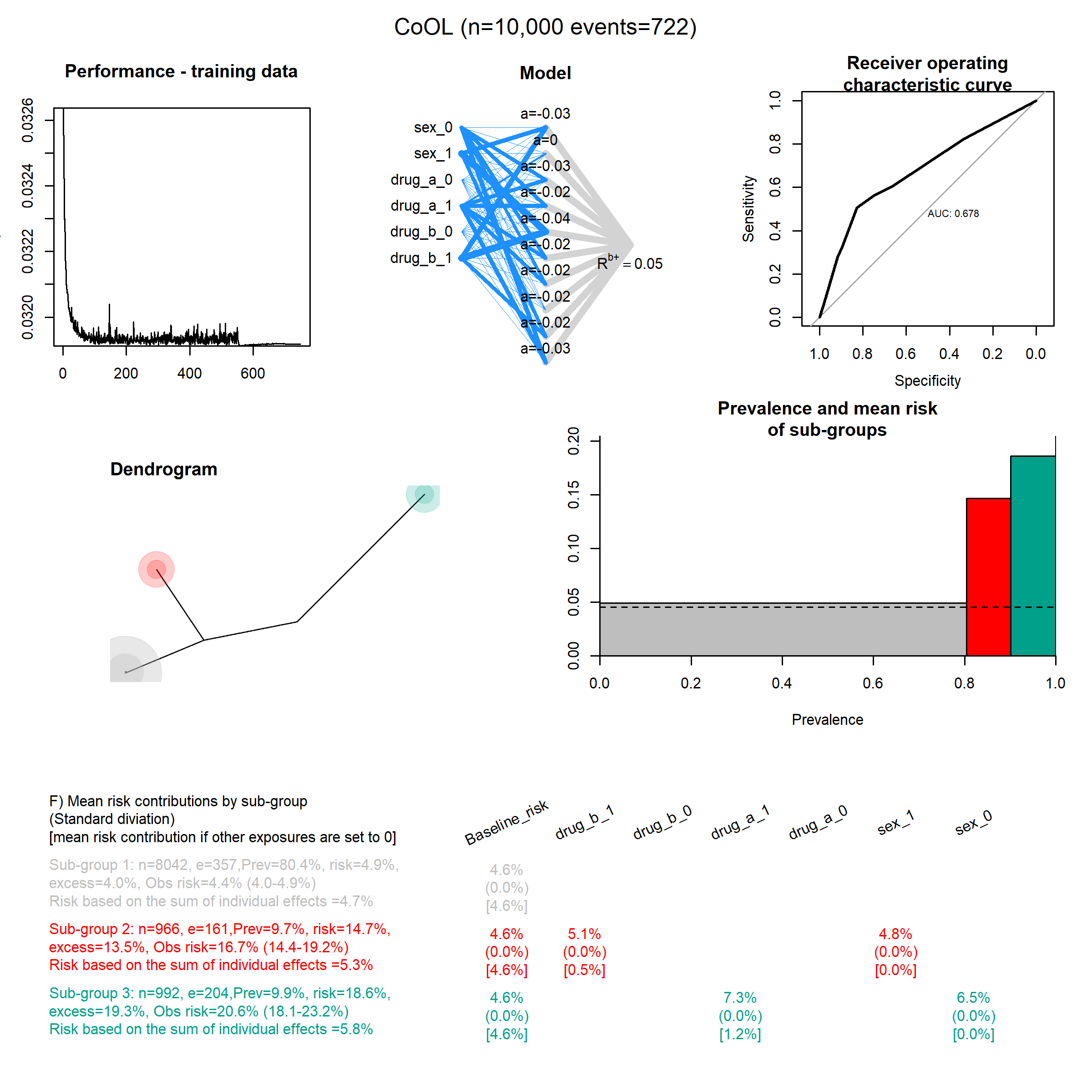

### Repeated_4.png

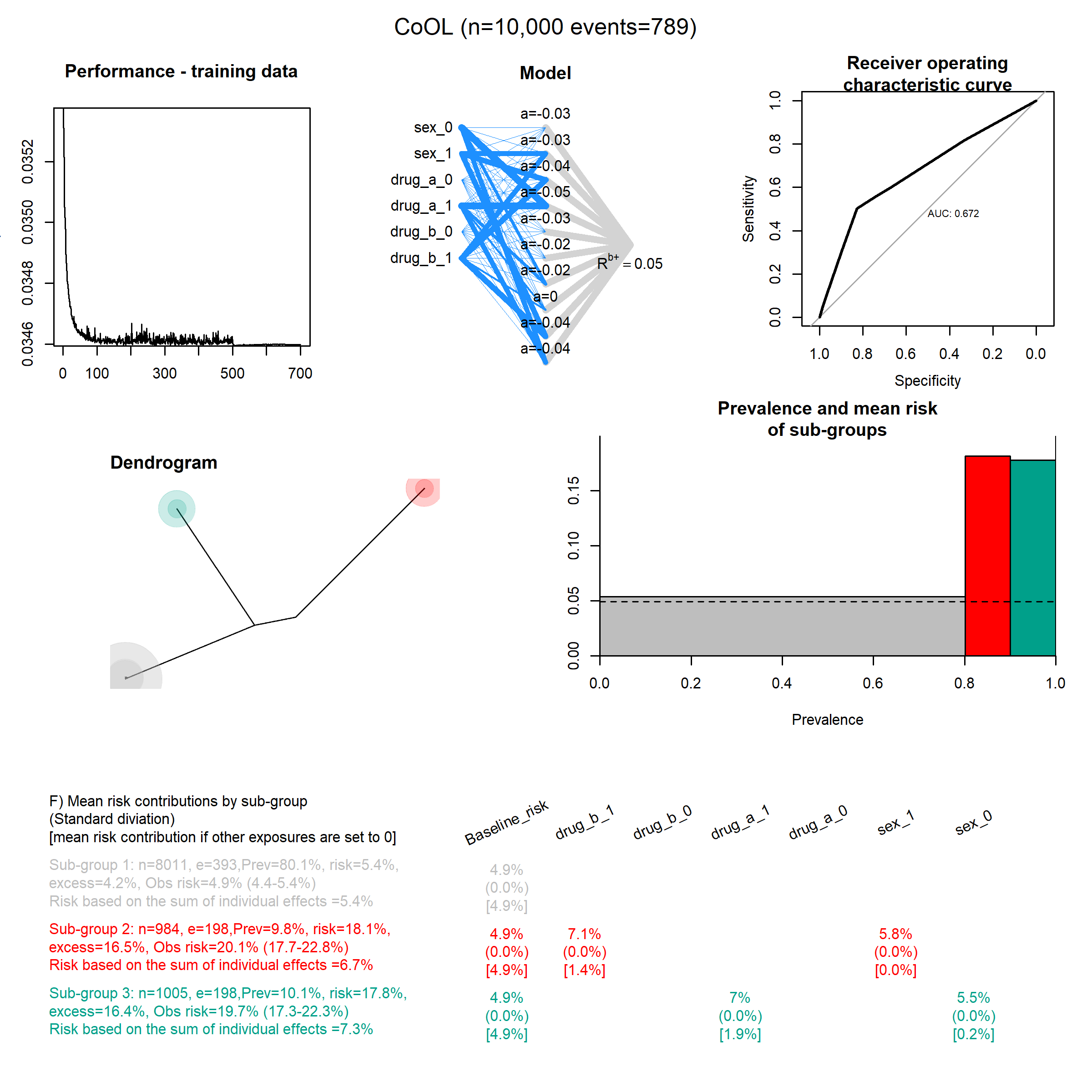

### Repeated_5.png

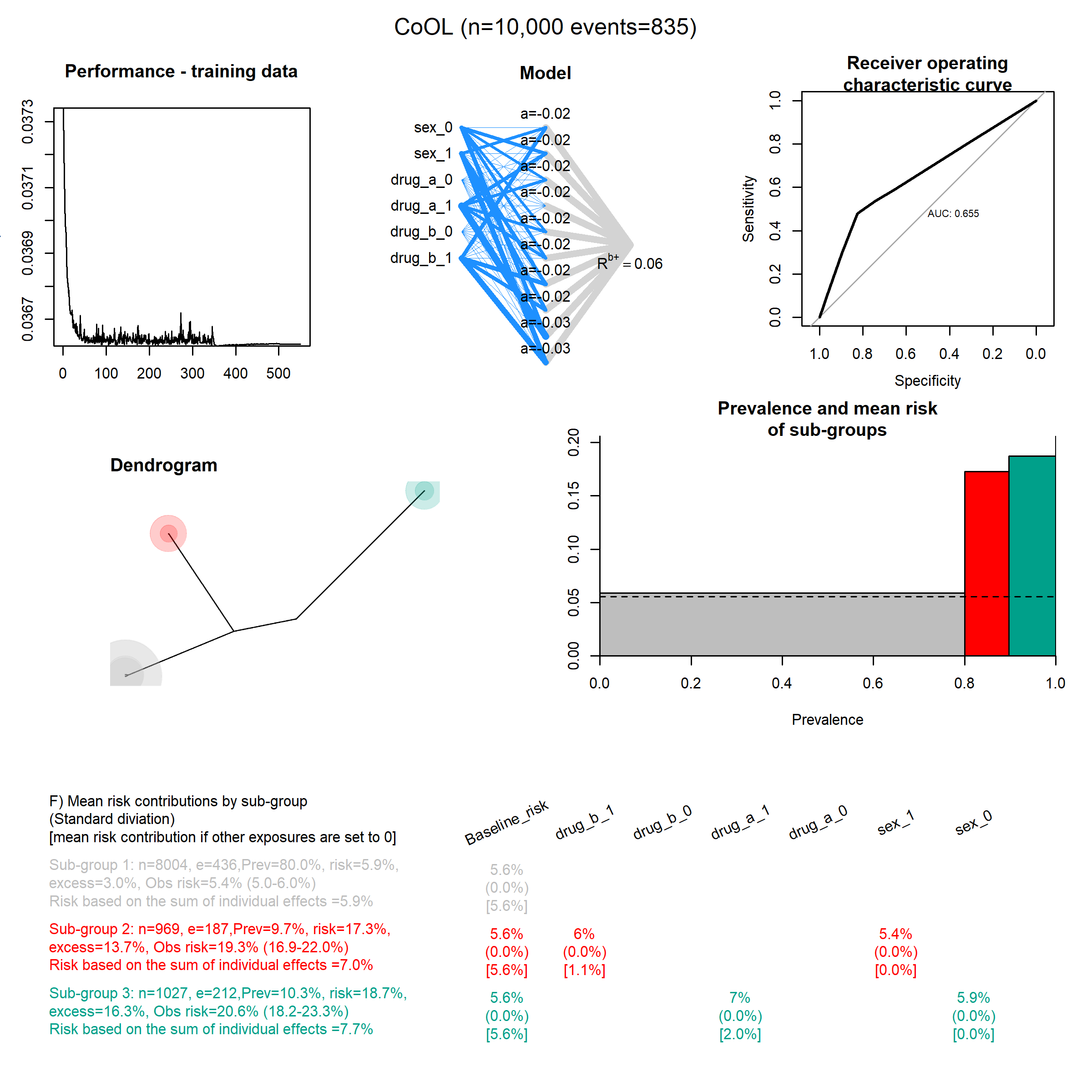

### Repeated_6.png

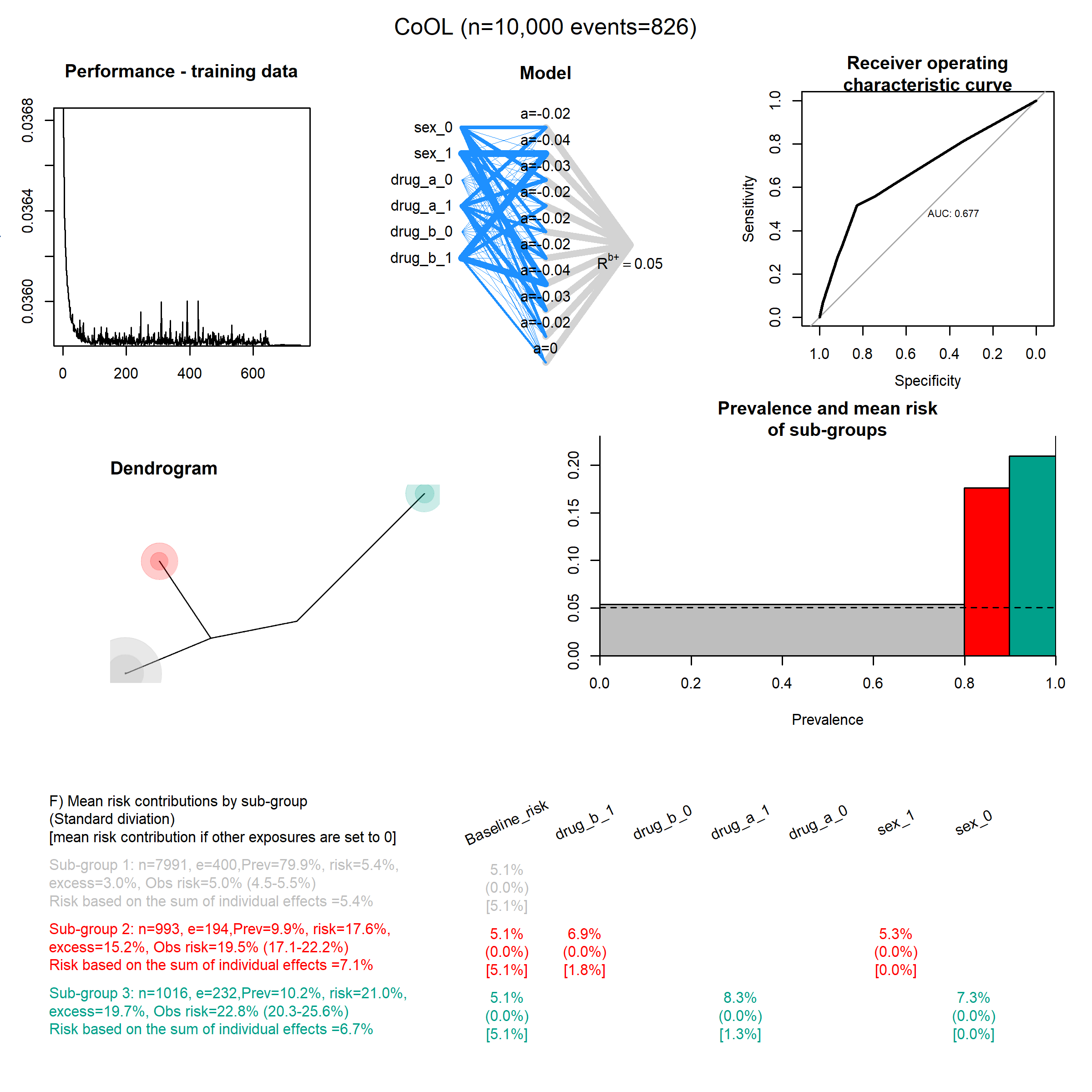

### Repeated_7.png

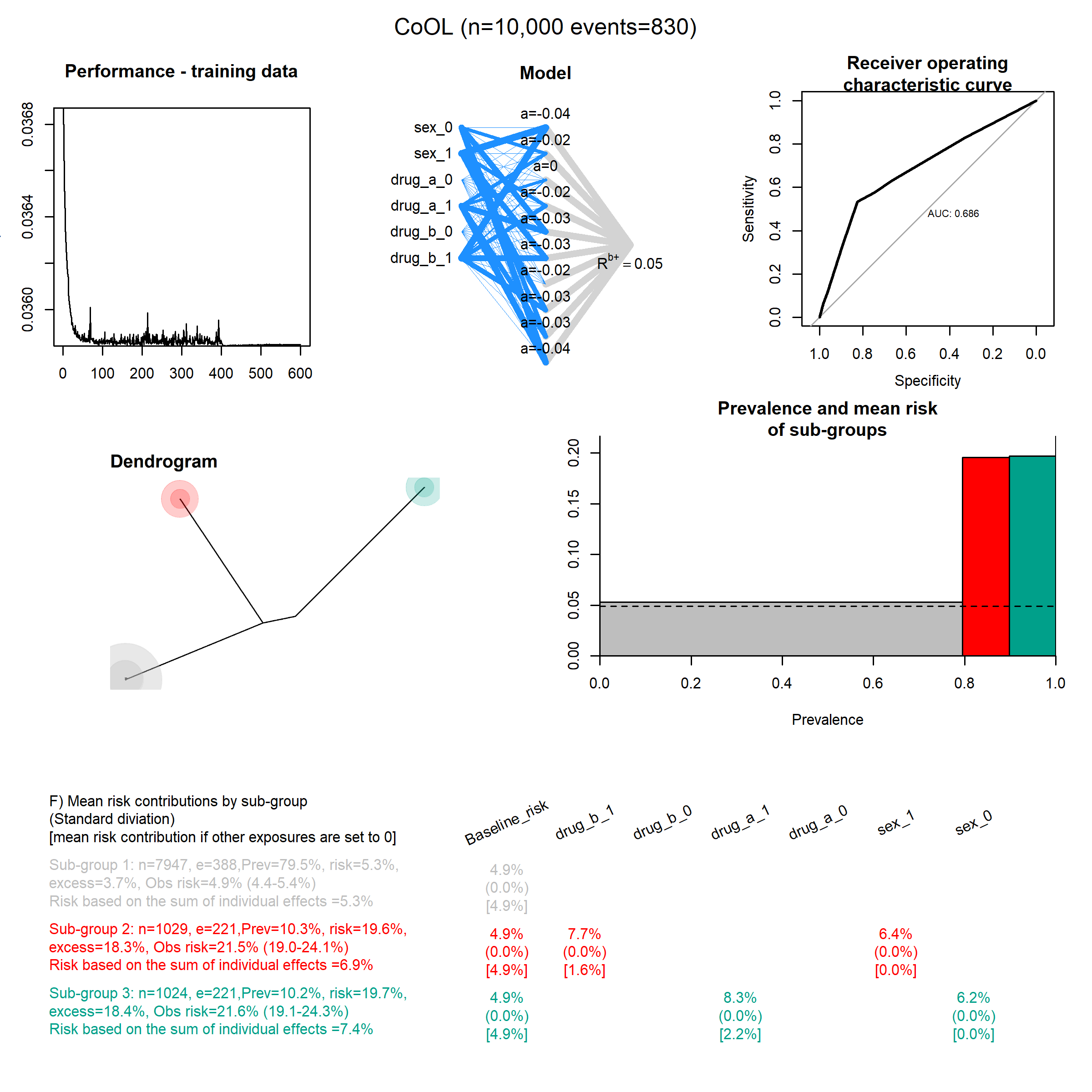

### Repeated_8.png

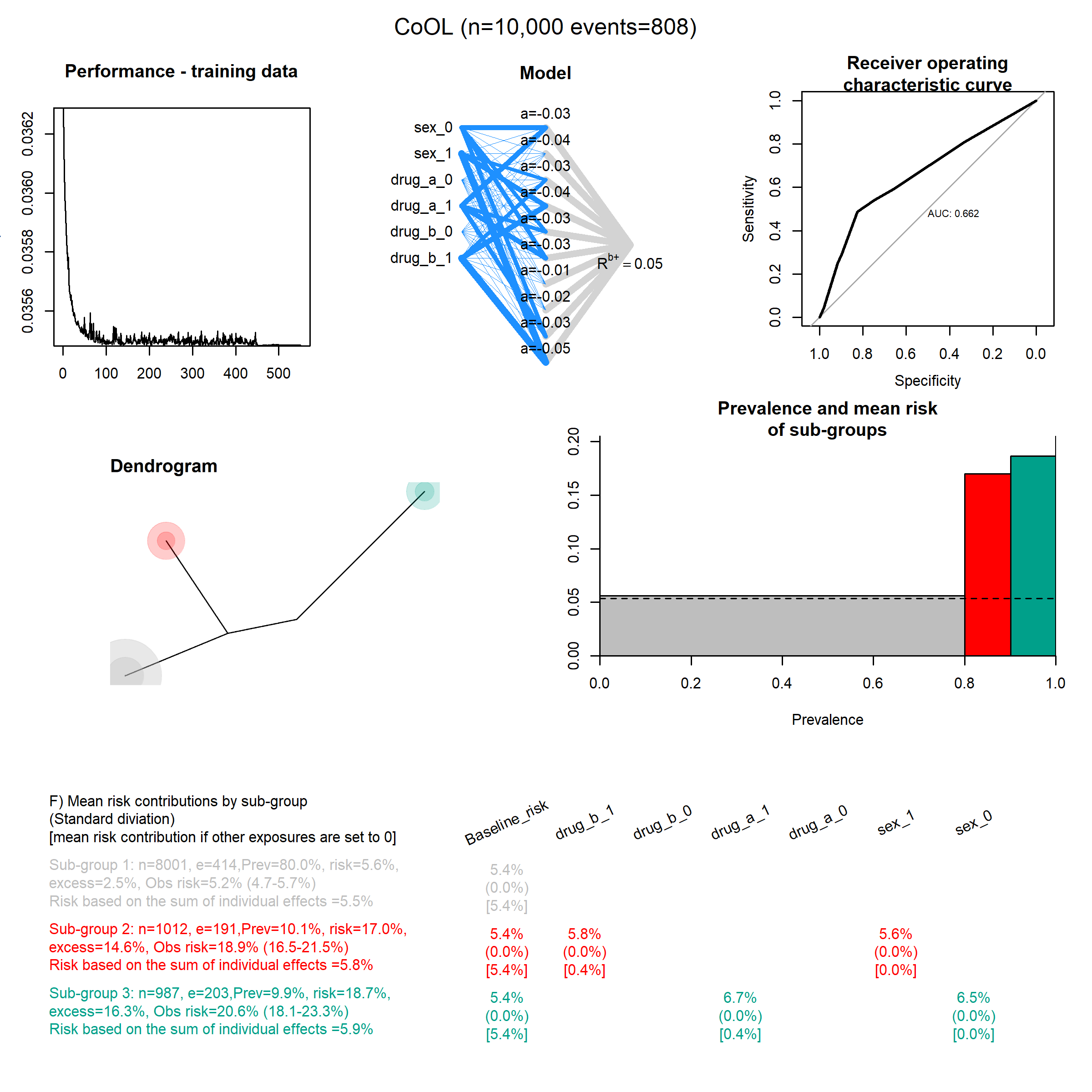

### Repeated_9.png

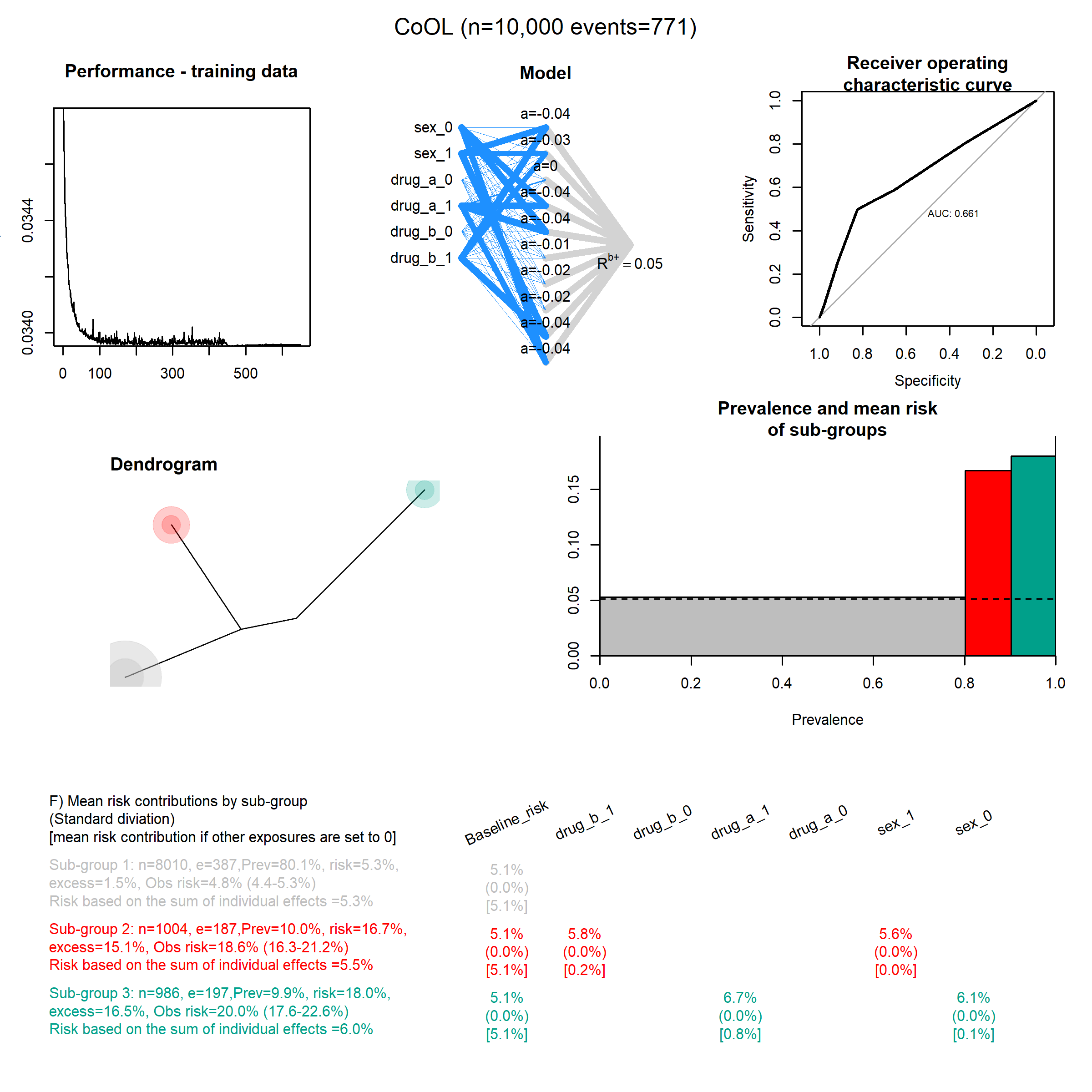

### Repeated_10.png

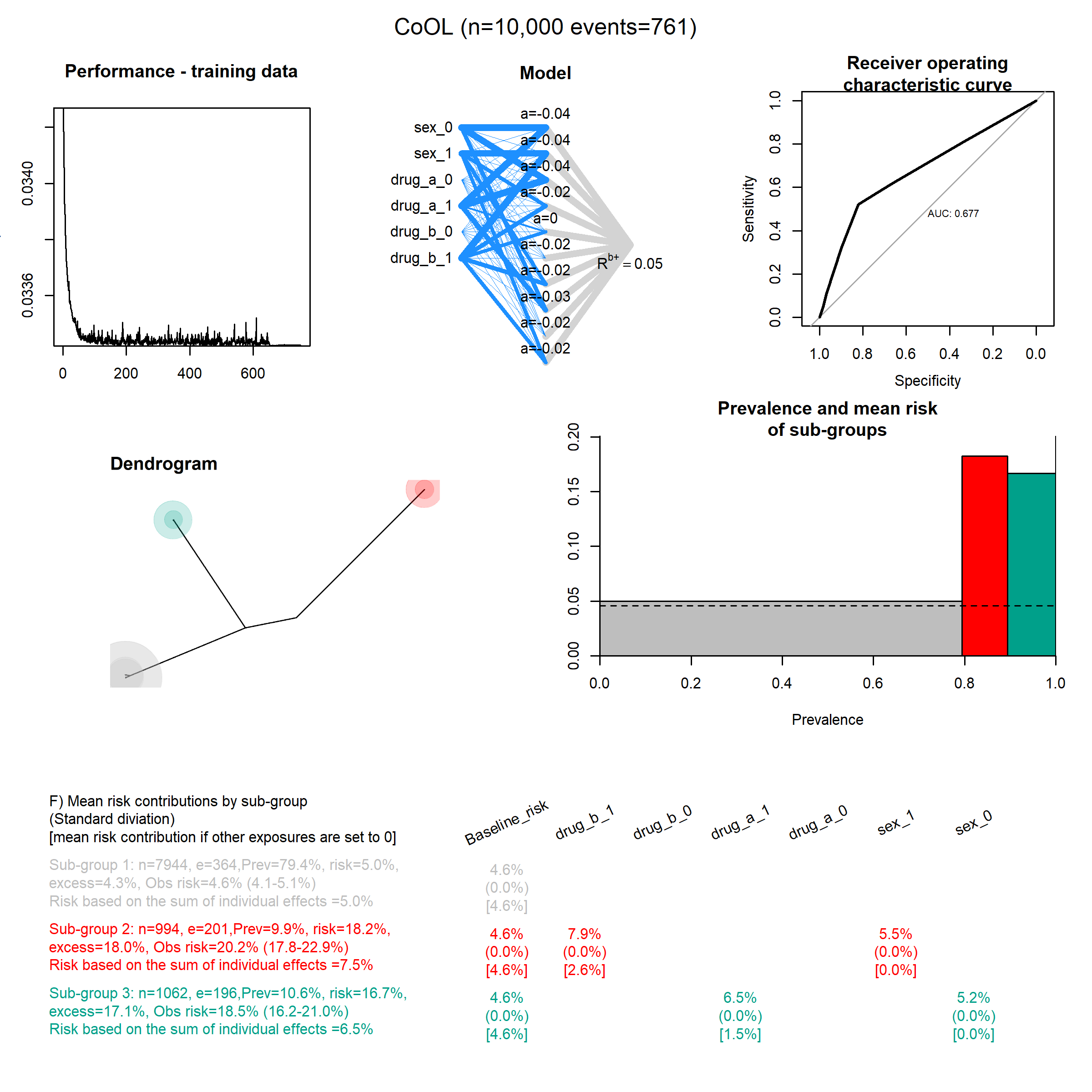

### Synergy_functions_1.png

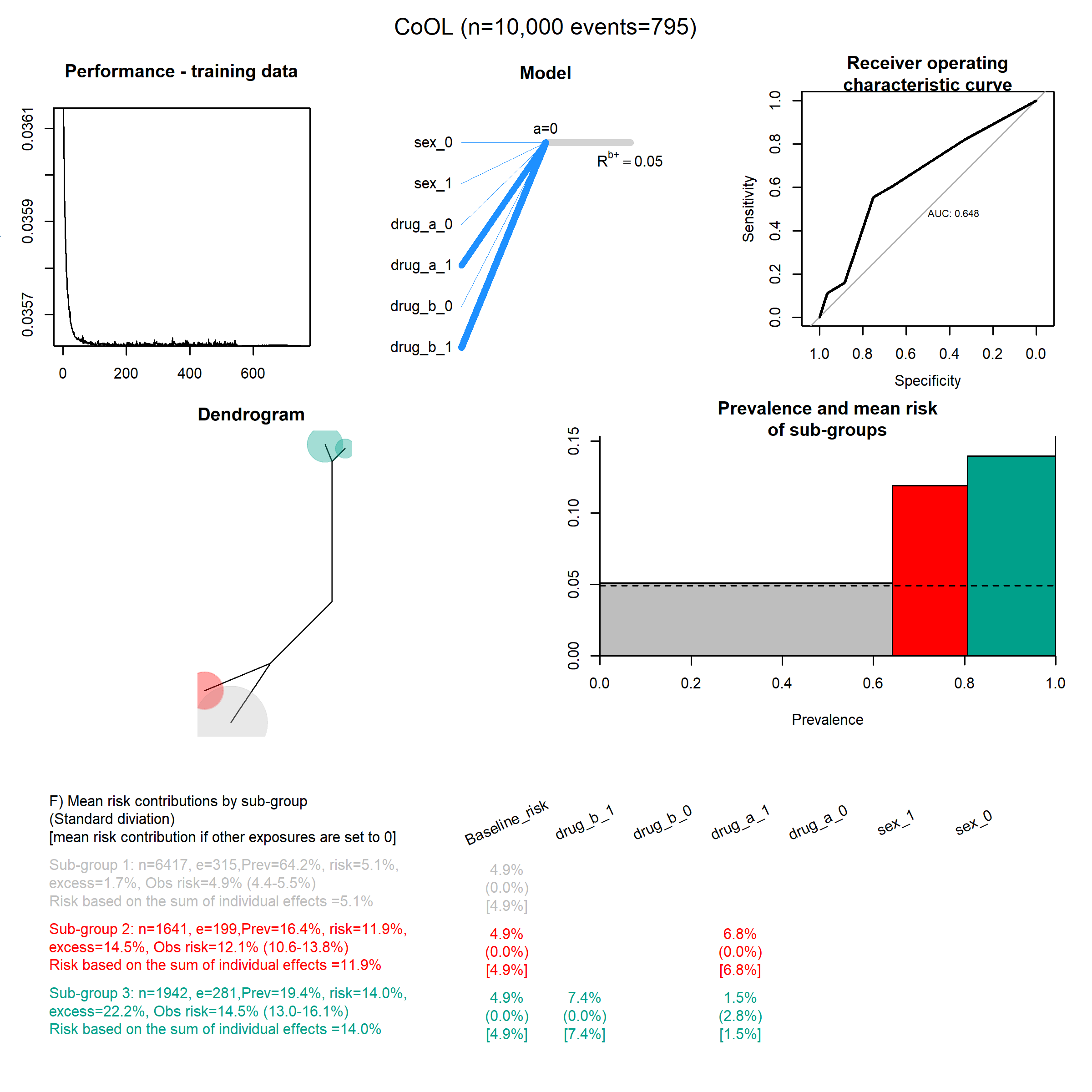

### Synergy_functions_2.png

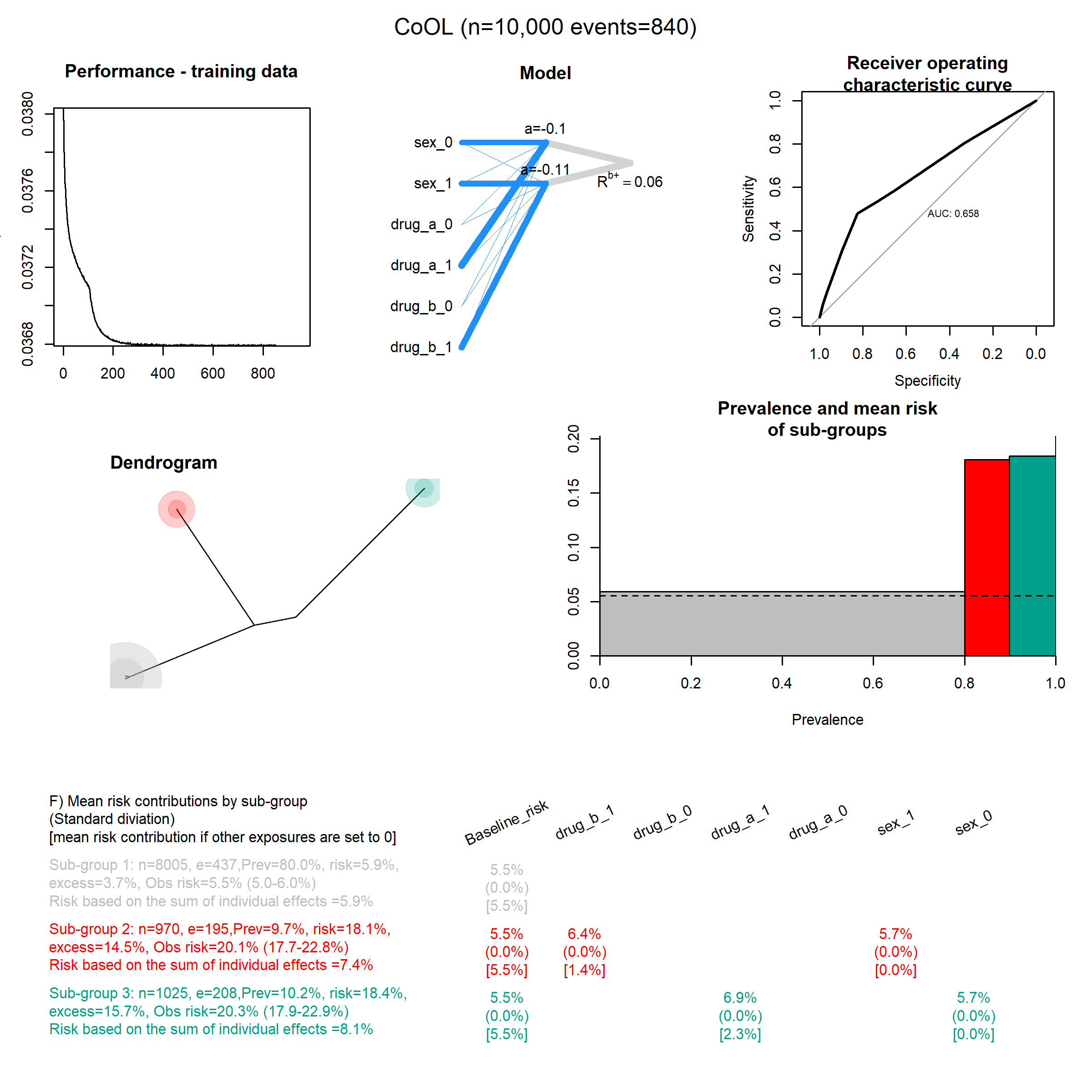

### Synergy_functions_3.png

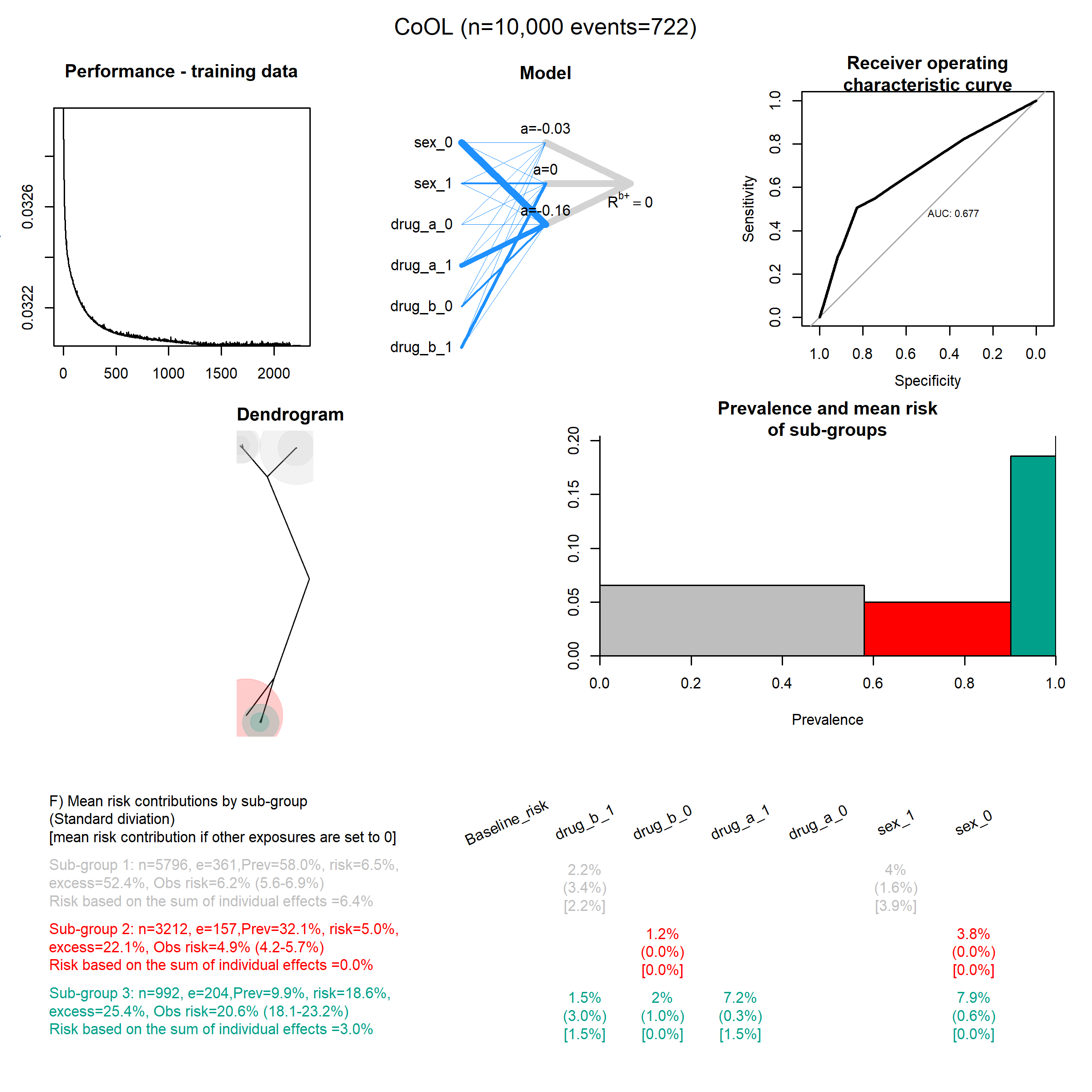

### Synergy_functions_4.png

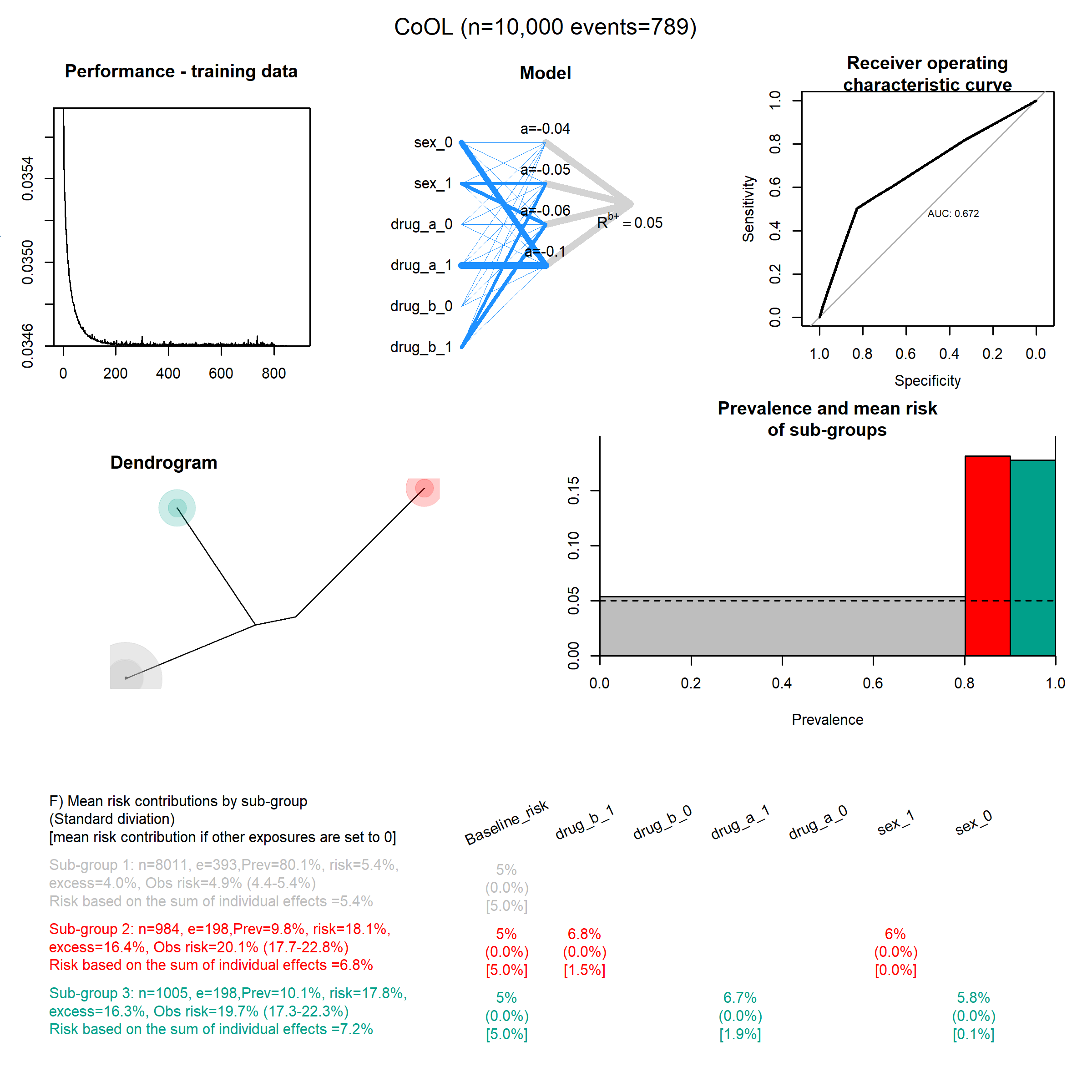

### Synergy_functions_5.png

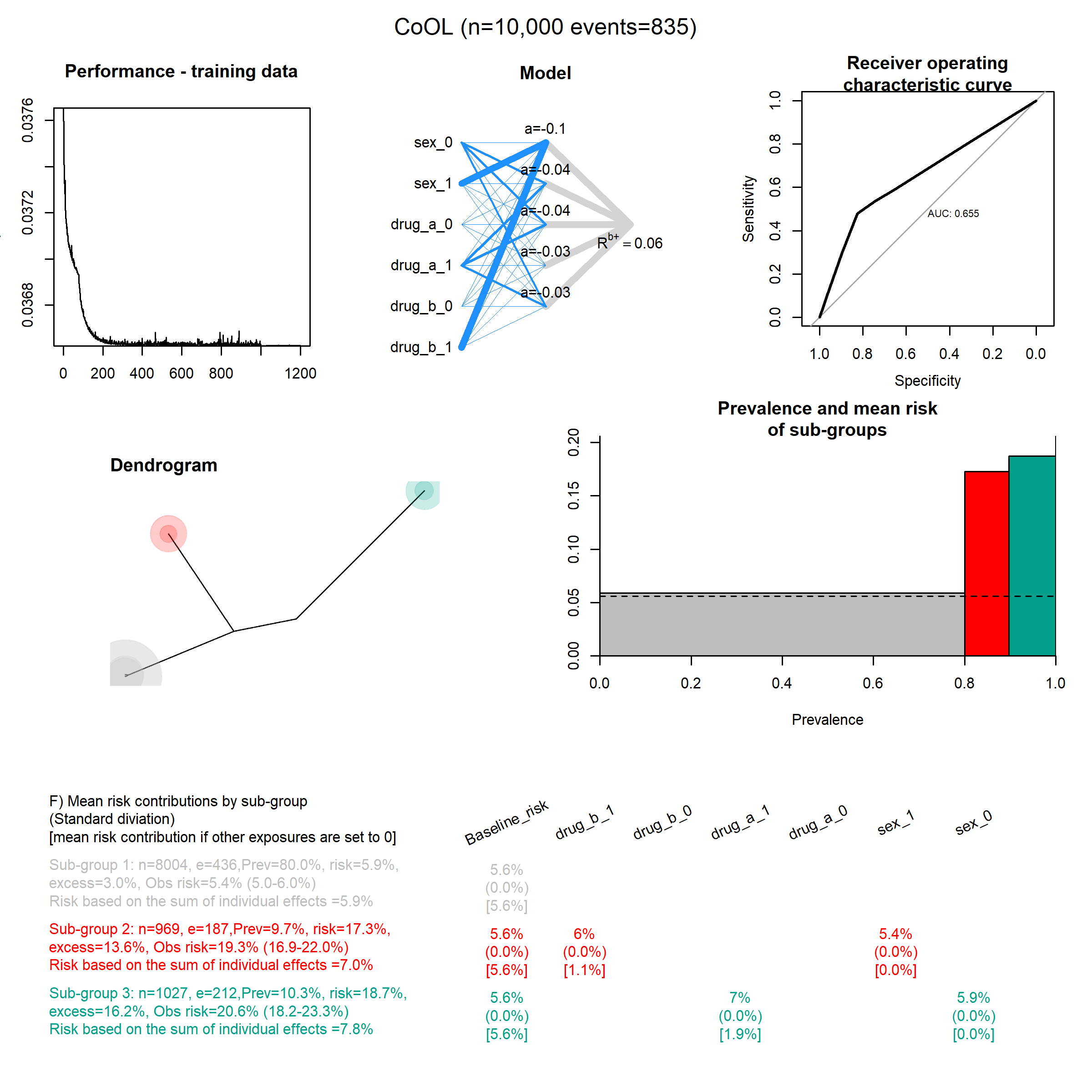

### Synergy_functions_15.png

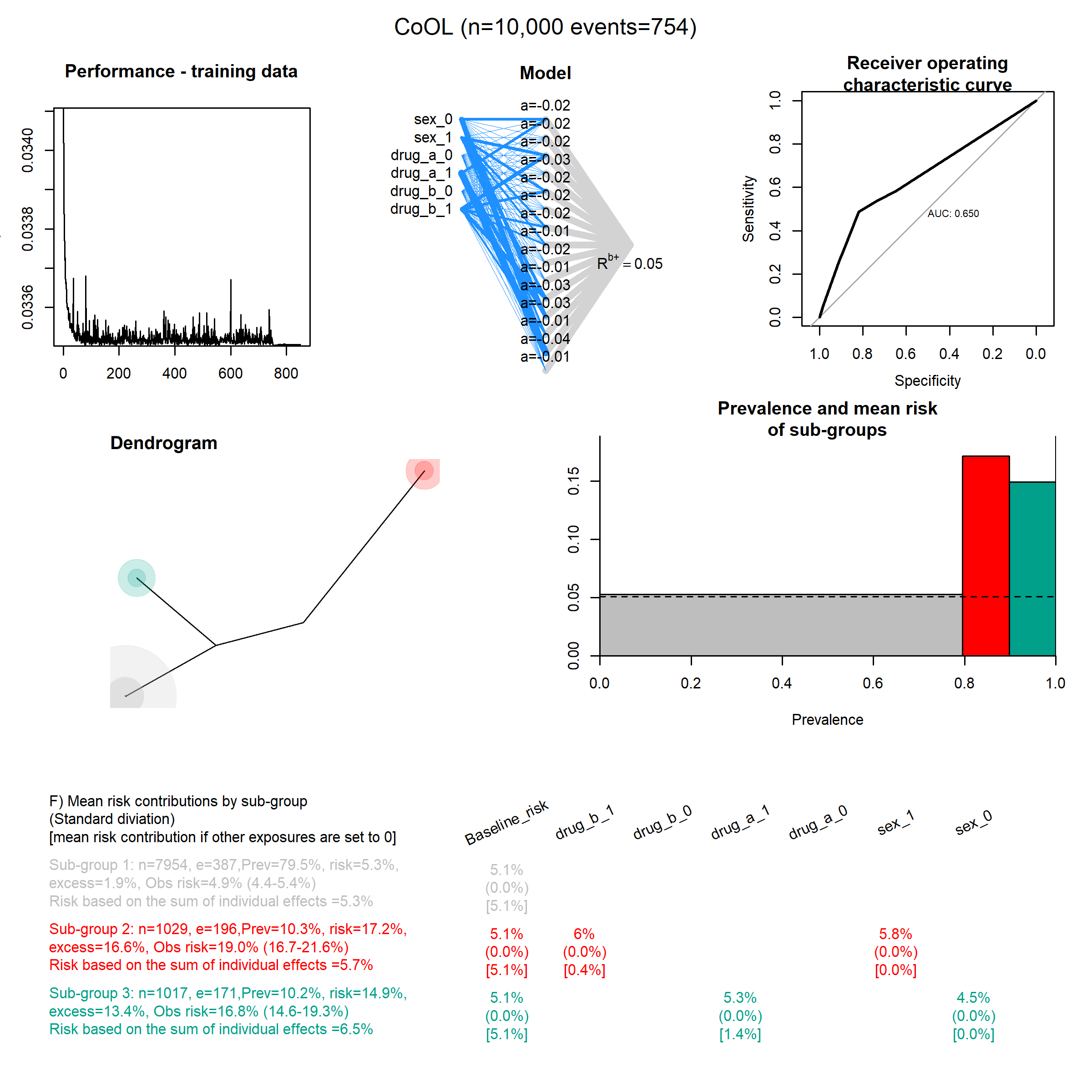

### Synergy_functions_20.png

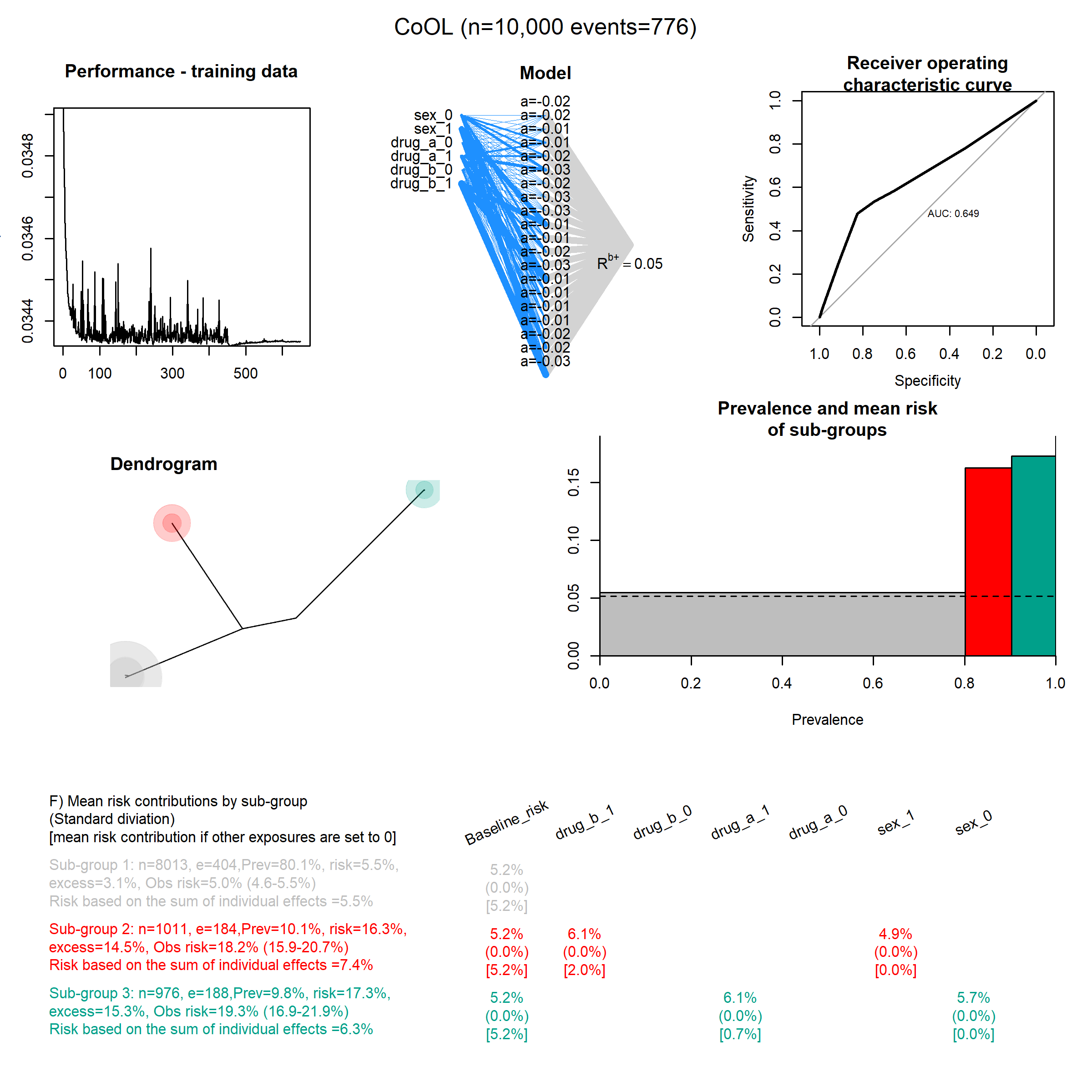
